# A machine learning-based phenotype for long COVID in children: an EHR-based study from the RECOVER program

**DOI:** 10.1101/2022.12.22.22283791

**Authors:** Vitaly Lorman, Hanieh Razzaghi, Xing Song, Keith Morse, Levon Utidjian, Andrea J. Allen, Suchitra Rao, Colin Rogerson, Tellen D. Bennett, Hiroki Morizono, Daniel Eckrich, Ravi Jhaveri, Yungui Huang, Daksha Ranade, Nathan Pajor, Grace M. Lee, Christopher B. Forrest, L. Charles Bailey

## Abstract

**Background:** As clinical understanding of pediatric Post-Acute Sequelae of SARS CoV-2 (PASC) develops, and hence the clinical definition evolves, it is desirable to have a method to reliably identify patients who are likely to have post-acute sequelae of SARS CoV-2 (PASC) in health systems data.

**Methods and Findings:** In this study, we developed and validated a machine learning algorithm to classify which patients have PASC (distinguishing between Multisystem Inflammatory Syndrome in Children (MIS-C) and non-MIS-C variants) from a cohort of patients with positive SARS-CoV-2 test results in pediatric health systems within the PEDSnet EHR network. Patient features included in the model were selected from conditions, procedures, performance of diagnostic testing, and medications using a tree-based scan statistic approach. We used an XGboost model, with hyperparameters selected through cross-validated grid search, and model performance was assessed using 5-fold cross-validation. Model predictions and feature importance were evaluated using Shapley Additive exPlanation (SHAP) values.

**Conclusions:** The model provides a tool for identifying patients with PASC and an approach to characterizing PASC using diagnosis, medication, laboratory, and procedure features in health systems data. Using appropriate threshold settings, the model can be used to identify PASC patients in health systems data at higher precision for inclusion in studies or at higher recall in screening for clinical trials, especially in settings where PASC diagnosis codes are used less frequently or less reliably. Analysis of how specific features contribute to the classification process may assist in gaining a better understanding of features that are associated with PASC diagnoses.

**Funding Source:** This research was funded by the National Institutes of Health (NIH) Agreement OT2HL161847-01 as part of the Researching COVID to Enhance Recovery (RECOVER) program of research.

**Disclaimer:** The content is solely the responsibility of the authors and does not necessarily represent the official views of the RECOVER Program, the NIH or other funders.

## Introduction

While long-term consequences of SARS-CoV2 infection have been studied from the perspectives of both clinical manifestations and underlying mechanisms^1,2^, formal definitions for Post-Acute Sequelae of SARS CoV-2 (PASC, or long COVID) are currently broad and necessarily nonspecific. Although advances have been made in describing features that characterize PASC, studies to date have suggested a heterogeneous presentation, particularly in children^3,4^. This heterogeneity has made it difficult to form a definition for predicting or classifying children with PASC in the absence of patient-specific expert review, resulting in challenges in obtaining “gold standard” labels for cases and controls, which consequently poses a challenge to conducting large-scale research towards improving patient outcomes.

To address this gap, researchers have developed machine learning algorithms intended to classify adult patients with PASC in large clinical databases^5^. In contrast with explicit rule-based definitions, machine learning algorithms have the advantage of being able to detect complex patterns involving thousands of covariates^6^.

Diagnosing PASC in the pediatric population is particularly challenging due to large differences in clinical manifestations across the age spectrum, compounded by a paucity of pediatric research to date. Multisystem Inflammatory Syndrome in Children (MIS-C), a clinically severe illness which follows SARS-COV-2 infection and satisfies the current time-based definition for PASC is considered a distinct entity in practice and does have a case definition which involves laboratory evidence of inflammation and involvement of at least two organ systems among SARS CoV-2-positive patients^7^. However, a fuller understanding of the subphenotypes of MIS-C, including the mechanisms by which they develop and their longer-term trajectories, is still emerging^8,9^. In the case of non-MIS-C PASC, the variety of presentations, variability of methods for diagnosis, and treatment modalities presents an even greater challenge in identifying such patients from EHR data^1–3,3,4,10,11^.

The goal of this study is to implement and validate a machine learning model to classify patients with a PASC diagnosis (including both MIS-C and non-MIS-C variants). Our model was trained to classify patients with a PASC diagnosis code in a cohort of SARS-CoV-2 positive patients, with the model’s utility found in its ability to detect patients who are likely to have had PASC based on a large collection of clinical features in settings where the diagnosis code was not reliably present. To define patient features for use in the model from large hierarchical vocabularies of clinical codes, we employed the tree-based scan statistic^12^, a data mining tool previously used for pharmacovigilance, vaccine safety surveillance, and occupational disease surveillance. In our context, the tree-based scan statistic was used to detect clusters of diagnosis, medication, procedure, and diagnostic test codes which occur disproportionately often among PASC-diagnosed patients; it detects the level of threshold hierarchical granularity at which to cluster the codes, allowing for feature selection without expert clinical input regarding which features to use and how to cluster them. To our knowledge, this is a novel application of the tree-based scan statistic to selecting features from a hierarchical structure for use in machine learning models and may be of independent methodological interest.

## Methods

### Study population

This retrospective cohort study is part of the NIH Researching COVID to Enhance Recovery (RECOVER) Initiative, which seeks to understand, treat, and prevent PASC. For more information on RECOVER, visit https://recovercovid.org/. The study population includes EHR data from the PEDSnet network and the following institutions: Children’s Hospital of Philadelphia, Cincinnati Children’s Hospital Medical Center, Children’s Hospital of Colorado, Ann & Robert H. Lurie Children’s Hospital of Chicago, Nationwide Children’s Hospital, Nemours Children’s Health System (in Delaware and Florida), Seattle Children’s Hospital, and Stanford Children’s Health. The Children’s Hospital of Philadelphia’s institutional review board designated this study as not human subjects research and the need for consent was waived. The PEDSnet RECOVER Database Version 202207 was used.

### Identifying PASC diagnoses

We identified patients with MIS-C by the presence of ICD-10-CM code M35.81, ICD 10 codes U10 or U10.9, OHDSI extension code OMOP5042964, or an (EHR) interface term containing the string ‘MIS-C’ or both the strings ‘multisystem’ and ‘inflam’. We identified patients with non-MIS-C subphenotypes of PASC using the ICD-10-CM diagnosis code U09.9 (available October 1, 2021^13^) or by an interface term in the EHR containing the string ‘long covid’ or ‘post-acute’ and ‘covid’ or ‘sars’. Based on evidence^14^ and early CDC guidance^15^ that the non-COVID-specific ICD-10-CM code B94.8 (“Sequelae of other specified infectious and parasitic diseases”) was used as a placeholder for long COVID prior to the introduction of U09.9, we elected to include among our cases patients who had an occurrence of this code or similar nonspecific post-viral infection codes in the SNOMED vocabulary that were not attributable to other conditions, when such codes occurred following a positive SARS CoV-2 test result.

Composite PASC diagnosis was defined as the presence of any of the above codes, including those for MIS-C. Non-MIS-C PASC diagnosed patients were defined as the subset of patients with a PASC diagnosis who did not have an MIS-C diagnosis.

### Cohort definition

Our cohort comprised patients less than 21 years old who had a positive SARS CoV-2 test (antigen, serology, or Reverse Transcriptase-Polymerase Chain Reaction (RT-PCR)), or PASC or MIS-C diagnosis at any point after January 1, 2021^7^. We used this cutoff date because it is when the MIS-C diagnosis code M35.81 first came into use. Because home viral testing became increasingly prevalent in 2022, and not all PASC-diagnosed patients in our cohort had a SARS CoV-2 test result, we did not require one. Further, for many PASC (including MIS-C)-diagnosed patients who did have a positive SARS CoV-2 test result, the test occurred on or near their earliest PASC diagnosis. Due to the resulting difficulties in capturing the true date of onset of SARS-CoV-2 infection in the EHR, we elected to impute the index date for all PASC-diagnosed patients by selecting a random date in the 28 to 90 days prior to their earliest PASC diagnosis. For non-PASC SARS-CoV-2 infected patients, we defined their index date as the date of first positive test result. We further required the presence of at least two visits following the index date for all patients in our cohort.

### Feature selection

Our feature set consisted of person-level demographics, conditions, performance of diagnostic testing, procedures, and medications. Demographic variables included site, index date month and year, age at index date, sex, and race/ethnicity. Diagnosis codes used to define the presence of PASC (U09.9, M35.81, B94.8) were excluded from the feature set.

The remaining variables were constructed from clusters selected from clinical vocabulary hierarchies using the tree-based scan statistic computed by the TreeScan software. In this study, we employed the unconditional analysis based on a Bernoulli probability model at each node of the tree. This approach identifies branches of the tree at which outcomes belonged most disproportionately to patients with PASC as compared to COVID-positive patients without PASC. We refer the reader to the TreeScan literature for more details^12,16^.

We used as input into the TreeScan software the SNOMED hierarchy for conditions, the RxNorm hierarchy for medications, the LOINC hierarchy for labs, and a union of the ICD-10-PCS, HCPCS, and CPT4 hierarchies for procedures. To ensure uniform follow-up time, the cohort we used for TreeScan feature selection was a subcohort of the full cohort of our study consisting of patients who had at least 3 months of follow-up. The code occurrences used as input to TreeScan were counted during the 28-180 days following the index date.

From the TreeScan output, we then selected the branches which were 1) significant at the p <0.05 threshold and 2) had at least 50 occurrences for conditions, diagnostic tests, and procedures. For medications, we required 1000 occurrences given the per-dose structure of inpatient medication records. We also omitted from consideration cuts above the 3^rd^ level of the respective vocabularies for conditions and above the 2^nd^ level for medications and diagnostic tests to avoid selecting clusters that were too general.

To create the feature space for our model, six binary variables were constructed for each branch selected by TreeScan to indicate whether the feature was present for each of the following time periods relative to index date: -1 to 0 months, 0 to 1 months, 1 to 2 months, 2-3 months, and 3-6 months.

### Model selection and evaluation

We used an XGBoost model with hyperparameters selected using cross-validated grid search. To distinguish between MIS-C and non-MIS-C PASC, the model was trained to classify patients as belonging to one of three classes (MIS-C, non-MIS-PASC, or no PASC) and model output for each patient consisted of one probability for each class such that the three probabilities sum to 1.

We used fivefold cross-validation to evaluate our model. The feature selection process was performed during each training step, and hence the dimension of the feature space varies slightly between the folds. Model performance was evaluated using a variety of metrics, including recall and precision at various probability thresholds, F1 scores, area under the Receiver Operating Characteristic (ROC) curve (AUROC), and area under the Precision-Recall (PR) curve (AUPR). Because both accuracy and AUROC can be misleading measures of model performance for imbalanced classification problems (in our case, cases form about 3% of our cohort), we favored AUPR as the primary metric to evaluate our models as it provides a concise summary of the recall-precision tradeoff across different levels of probability threshold^17^.

### Feature importance

To evaluate our model’s predictions and feature importance, we calculated SHapley Additive exPlanation (SHAP) values.^18,19^ SHAP values, a game-theoretic concept repurposed for machine learning, allow us to see how various features from our high dimensional feature space contribute to determining the model’s output for each patient. Feature importance ranking is based on the mean absolute value of SHAP values for each feature.

## Results

### Cohort

Our cohort comprised 87,398 children meeting eligibility criteria for SARS-CoV-2 infection and follow-up (Table 1). Of these, 1,049 had a diagnosis of MIS-C, and 1,215 were diagnosed with non-MIS-C PASC. Most patients entered the cohort between September 2021 and April 2022. Contributions from individual health systems varied from 1.7% to 26.8% of the cohort. Serology testing in the absence of viral test results was used to identify 2.3% of the controls. Consistent with code availability, a higher proportion of patients with MIS-C diagnoses entered the cohort in early 2021 than those with non-MIS-C PASC (Figure S1).

**Table 1:**
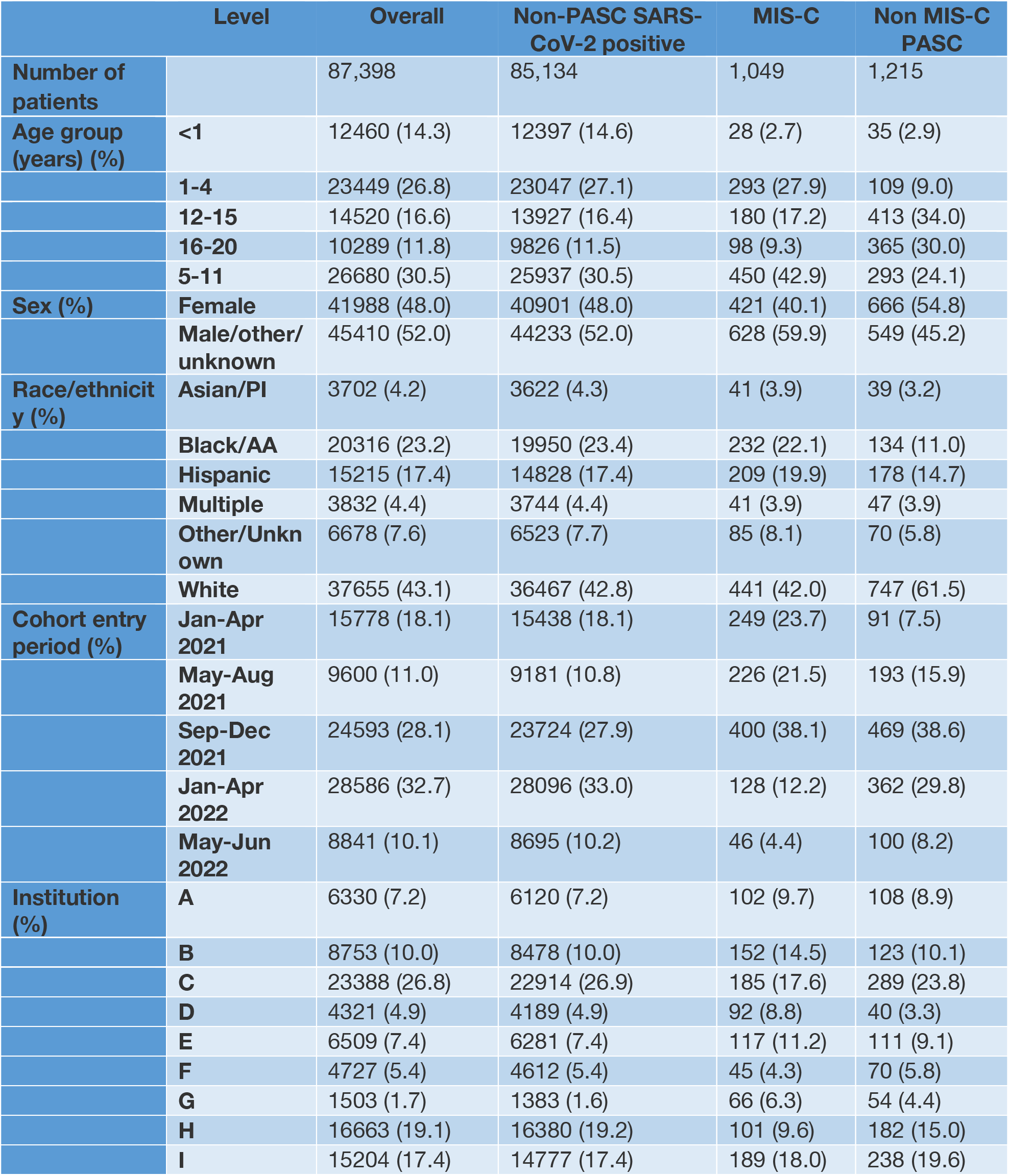
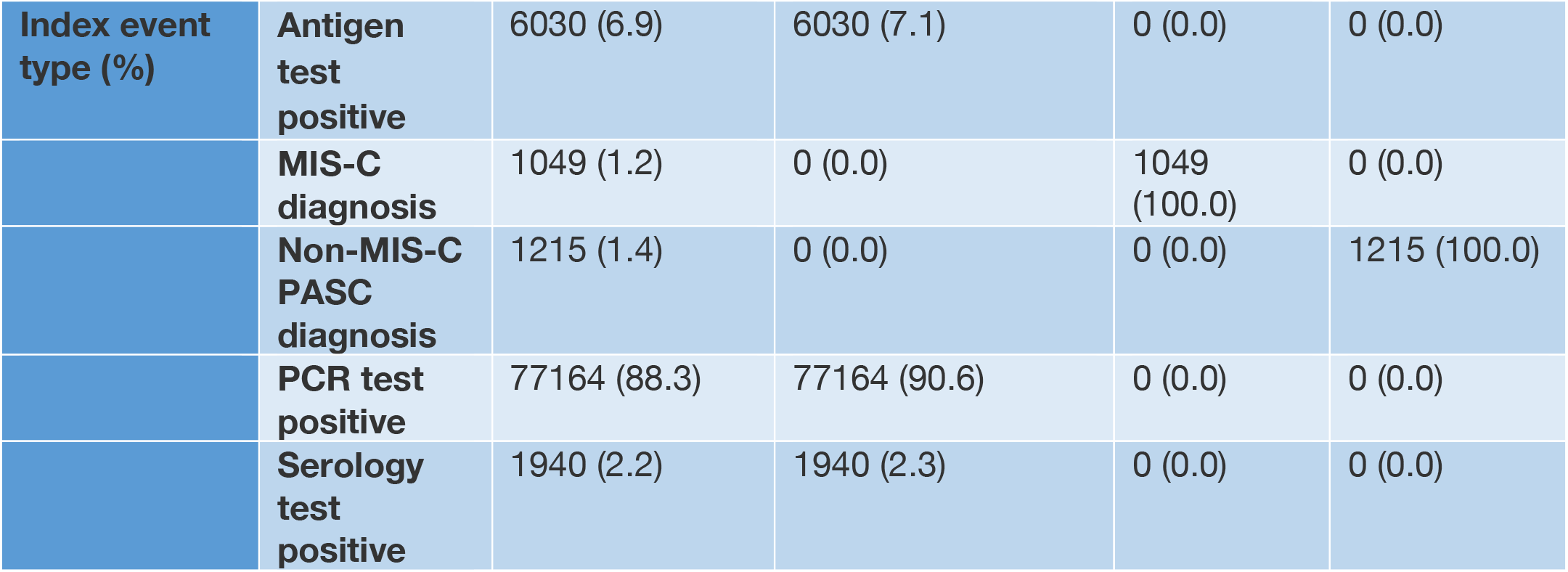
Cohort demographic and clinical characteristics

**Table 2:** Machine learning model performance in identifying patients with PASC (MIS-C or non-MIS-C variants) The table below displays several performance metrics for each of the three outcomes (PASC (any), non MIS-C PASC, and MIS-C). In addition to composite performance statistics, we report the area under the precision-recall curve (AUPR) and area under the receiver operating characteristic curve (AUROC). The curves themselves are shown and described in Figure 1.

|  | PASC (any) | MIS-C | Non MIS-C PASC |
| --- | --- | --- | --- |
| **Accuracy**<br>**(at p=0.5 threshold)** | 0.994 | 0.997 | 0.995 |
| **AUPR** | 0.917 | 0.939 | 0.810 |
| **AUROC** | 0.990 | 0.998 | 0.981 |
| **F1 score**<br>**(at p=0.5 threshold)** | 0.866 | 0.882 | 0.754 |
| **Precision**<br>**(at p=0.5 threshold)** | 0.945 | 0.954 | 0.864 |
| **Recall**<br>**(at p=0.5 threshold)** | 0.799 | 0.820 | 0.669 |

### TreeScan-selected features

TreeScan selection resulted in 645 condition features, 1,102 diagnostic test features, 197 procedure features, and 112 medication features. The terms from the SNOMED-CT ontology for which TreeScan found the greatest enrichment in patients diagnosed with PASC are shown in Table S1a. Some terms reflect specific pathologic findings (*e*.*g*. myocarditis), especially cardiovascular processes, and particularly those consistent with MIS-C. However, many of the terms with highest risk ratios are internal nodes in the SNOMED-CT hierarchy, and reflect the sum of contributions from multiple more specific terms. This pattern reflects the heterogeneity of presentation, particularly in patients without MIS-C, such that no single specific diagnosis best distinguishes between cases and non-cases. Results from medication data (Table S1c), diagnostic testing (Table S1d), and procedures (Table S1b) again reflect cardiovascular therapies, as well as immunomodulation, and supportive care for acute illness.

### Model performance

The final algorithm was an XGBoost^20^ model with a 10,339-dimensional feature space consisting of features described above evaluated at 5 different time windows around time of SARS-CoV-2 infection (−1 to 0 months, 0 to 1 months, 1 to 2 months, 2-3 months, and 3-6 months).

Using five-fold cross-validation we found that our model classified whether patients had PASC of any kind with 91.7% AUPR, 99.0% AUROC (Figure 1), with 79.9% recall and 94.5% precision at the p=0.5 threshold. In classifying whether patients had MIS-C, the model achieved 93.9% AUPR and 99.8% AUROC, with 82.0% recall and 95.4% precision at the p=0.5 threshold. In classifying non-MISC PASC patients, the model performed with 81.0% AUPR and 98.1% AUROC, with 66.9% recall and 86.4% precision at the p=0.5 threshold.

**Figure 1:**
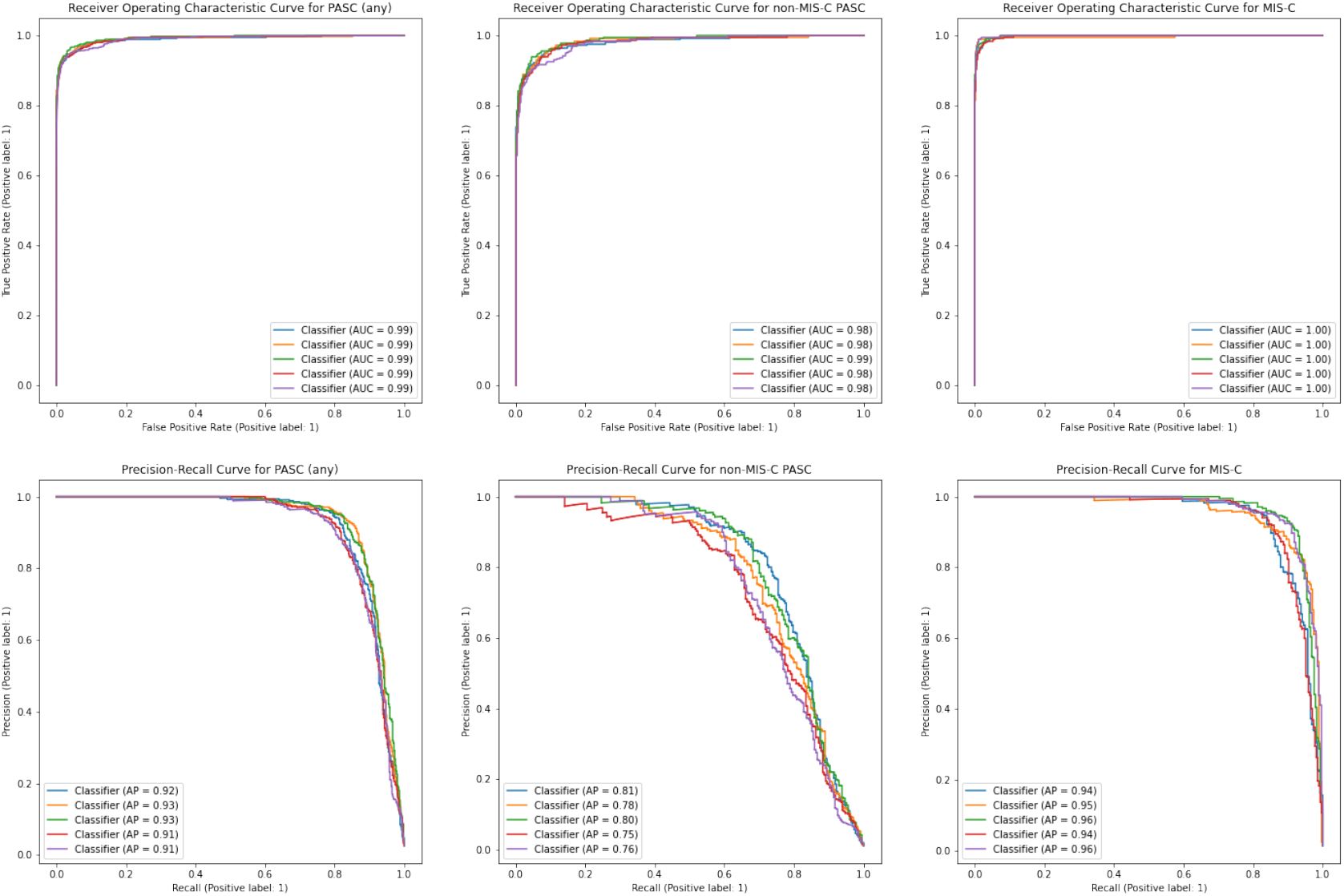
Receiver Operating Characteristic (ROC) and Precision-Recall (PR) curves describing model performance in identifying PASC with (MIS-C or non-MIS-C variants) For each of the three outcomes (PASC (any), non MIS-C PASC, and MIS-C) the Receiver Operating Characteristic (ROC) curves and Precision-Recall (PR) curves are estimated and plotted 5 times, once for each cross-validation fold.

### Feature importance

While SHAP summaries^18,19^ do not provide precise data about feature importance, they do provide approximations of features’ impact on model results. Results for the 3-outcome model are presented in Figure 2, aggregated by clinical domain. Because the model output is multinomial, the *x* axis is expressed in terms of log odds, rather than probabilities. Among demographic features, time of cohort entry was most impactful. Unsurprisingly, the clinical feature with the greatest positive result was the presence of an inflammatory diagnosis. Diagnosis features appeared more often as positive predictors, while laboratory testing appeared more often as negative predictors. Additional SHAP summaries for specific data domains are presented in Figure S2.

**Figure 2.**
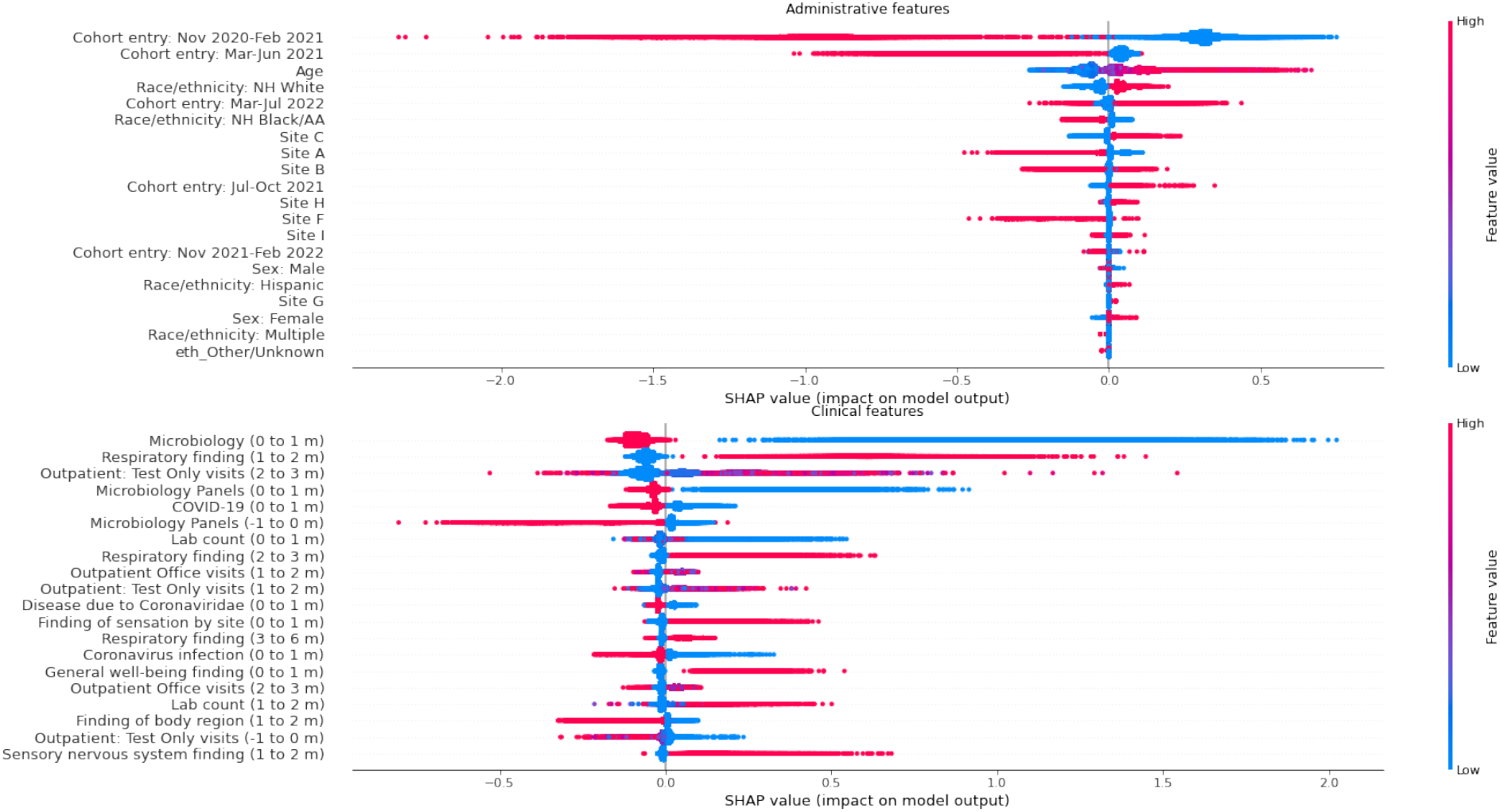
SHapley Additive exPlanation (SHAP) values for top administrative and clinical model features by class in predicting non-MIS-C PASC The plots show the most significant features as determined by the sum of SHAP value magnitudes over all samples. For each feature, SHAP values for each patient are plotted, with color representing the feature value (e.g. red if feature was present and blue if absent in case of a binary variable). For the SHAP values pictured, the x axis is interpreted as change in log odds (in particular, SHAP values are not confined to be between –1 and 1).

## Discussion

We have employed machine learning to identify potential patients with PASC based on EHR data collected during clinical care. As controls substantially outnumber cases in our cohort, accuracy and AUROC tend to be inflated, so we assessed our model’s performance primarily in terms of the Precision-Recall curve. At the p=0.5 threshold, our model was a high-precision classifier for PASC in general as well as for MIS-C and non-MIS-C PASC separately. Indeed, the model was able to classify patients with up to 70% recall with near perfect precision, with a drop-off in precision below 80% beginning only around 85% recall. At this threshold, our model’s high precision could be a valuable tool for identifying patients with PASC for study cohorts, particularly patients who did not receive a diagnosis due to unavailability of specific diagnosis codes, evolving clinical understanding of PASC, or less severe presentation. At lower probability thresholds, the model is potentially useful for identifying patients who may have features of PASC for further screening, such as might be done for clinical trial recruitment.

Review of feature contribution using SHAP value visualization reveals complex patterns, especially for non-MIS-C variants of PASC. In the more straightforward MIS-C case, the single diagnosis with consistent positive impact is ECG abnormalities; procedure codes show a similar pattern. Continuing this pattern, laboratory testing for inflammation and cardiac injury has a positive impact, while additional infection-related testing is more characteristic of unaffected patients. Aspirin, used only for a narrow set of indications in pediatrics, has the strongest positive influence among medications, with other immunomodulators associated with MIS-C (corticosteroid and immunoglobulin) appearing as well. SHAP data for the non-MIS-C group of patients reflects their greater heterogeneity. The skew toward later cohort entry despite masking of the PASC diagnosis code itself may reflect differences in PASC natural history, or in practice patterns for evaluation and treatment. Like children with MIS-C, lab utilization at the time of SARS-CoV-2 infection is not associated with caseness, but post-acute utilization is. However, no specific laboratory testing has a strong effect. Interestingly, several types of diagnostic imaging, but especially CT imaging during the acute phase of COVID19 may have positive predictive value. Among diagnoses, respiratory and neurologic codes in the early postacute period were observed to exert the greatest positive impact, consistent with respiratory medications, while aspirin and acetaminophen have negative impact, likely due to their role in MIS-C treatment. Coronavirus infection codes during the 1 to 2 month window suggests such codes are often still used for PASC-diagnosed patients during the post-acute period.

The model’s difficulties with the remaining 15% of PASC patients were driven by patients with non-MIS-C PASC. The lower recall for these patients could be a result of several factors. First, because the U09.9 diagnosis code was implemented in October 2021, there is a shorter overall duration for appropriately labelled cases to accrue in the training data. Though we attempted to mitigate this by including as cases patients who received more general post-viral disorder codes following a positive COVID test, it is possible that these more general codes were not used consistently. Additionally, non-MIS-C PASC patients had, on average, less follow-up time than patients with MIS-C. A second factor is that MIS-C is a more sharply defined condition with similarities to Kawasaki disease, an existing clinical phenotype, and thus there are features in the data related to diagnostic test utilization that align with consensus definitions of MIS-C (*cf*. Figure S2e). By contrast, we see more reliance on co-occurring symptoms and conditions to define non-MISC-PASC. These are more heterogeneous given the range of possible symptoms and presentations and likely include more characteristics that are poorly coded in discrete EHR data, such as fatigue or school difficulties.

We expect that our observed ceiling in recall at 85% may also reflect several limitations in the available training set. The primary challenge of using PASC diagnosis information in EHR data as a training standard, particularly for non-MIS-C PASC, is that the lack of a specific definition means the presence or absence of PASC-specific diagnosis codes does not constitute a gold standard for defining PASC. Moreover, patients who were seen with PASC and MIS-C may not have received the relevant diagnosis codes either due to the recent release of the codes or the early limited understanding of PASC, especially in the case of less severe sub-phenotypes. Thus, while diagnosis codes can serve as a starting point for assessing complications due to COVID-19, there is likely under-ascertainment of PASC which may result in false negative labels in our control population. The lack of a consensus definition may drive differences in clinical assignment of the code, and many patients may either be undiagnosed or never visit a clinician if the symptoms are not severe. On the other hand, false positives are also possible: many PASC features may reflect ongoing symptoms or conditions related to the acute COVID-19 infection, and therefore the lack of clear boundary between acute and post-acute symptoms and conditions may have led to patients in the acute phase of illness receiving a PASC diagnosis. Additionally, unlike adult care, there are relatively few institutional markers, such as dedicated long COVID-19 clinics, for patients with PASC, who receive their care in broader specialty settings such as cardiology or neurology. Lack of follow up for non-MIS-C PASC patients may also limit the model’s ability to identify distinguishing characteristics. The SHAP value plots indicate that many of the predictive features of PASC occur within the first three months from diagnosis, which may reflect this limitation.

When compared with adults, PASC in children may present with a milder clinical course^21^, leading to underdiagnosis in the health care setting. Some effects of PASC may have been seen in school performance or extracurricular participation, neither of which may have been captured in the HER. Pediatric PASC may less frequently involve focal organ dysfunction that prompts specific utilization, such as laboratory testing or diagnostic imaging which is well-captured in EHR data. Additionally, the majority of PASC manifestations in children possibly occur shortly following acute infection and resolve more rapidly than in adults^10^. These factors both require a distinct approach to classifying children with PASC and warrant further investigation as clinical guidelines and definitions are developed.

To our knowledge, this is the first study that has applied machine learning to an exclusively pediatric population to identify patients with PASC. A recent study^5^ developed a machine learning phenotype in the adult population trained on classifying whether patients were seen at a long COVID clinic. The model achieved 0.92 AUROC with 0.85 precision and 0.86 recall, the latter being slightly greater than observed here. Differences between our model and theirs include our pediatric population, and the use of PASC diagnoses as an outcome rather than specialty clinic attendance, due to limited use of dedicated PASC clinics at pediatric institutions.

Of note, we believe this study is also the first to use TreeScan for clinical feature selection in a machine learning model. The sets of possible features in each domain number in the hundreds of thousands, making reduction of dimensionality a necessary part of model development. Further, it is difficult to identify the correct level of granularity at which concepts should be clustered to define model features. This can be addressed through clinician expertise but is a time-consuming process which may incorporate bias stemming from individual clinicians’ practices or the coding practices at their institutions. However, vocabularies such as SNOMED-CT or RxNorm have rich hierarchies based on physiology or chemical structure, respectively. TreeScan therefore presents an appealing alternative: it takes the hierarchical structure of clinical vocabularies into account and does not require pre-specifying the level of granularity at which features should be selected.

Our study does have additional limitations, many in common with other studies which use EHR data to investigate PASC, a new condition with a broad definition. First, our model was trained to classify whether patients had a PASC diagnosis, which does not include all patients having PASC. Patients with milder manifestations may have been less likely to receive a diagnosis and thus would be less likely to be detected by our model. Second, our model was trained on data from tertiary and quaternary pediatric health systems, and so it may reflect biases in access to care as well as clinicians’ practices. Though a matched cohort design could be used to address some of these biases, it would run counter to our purpose of identifying patients likely to have PASC from routine EHR data. Third, while using data from a variety of sites may avoid overfitting the model to site-specific coding practices, there could be bias arising from coding practices across PEDSnet, as a network of pediatric health systems. A fourth potential limitation stems from our approach to imputing index dates for patients with PASC. Due to our difficulty in accurately capturing the initial SARS-CoV-2 infection date for such patients, we used a random date between 28 and 90 days prior to the first PASC diagnosis as the index date. However, this may not accurately reflect the date of the patient’s SARS-CoV-2 infection and as a result, relevant features may appear in a different time window relative to infection than when they actually occurred. Given the short follow-up for many patients, this could lead to a model that heavily weighs data close in time to infection. Finally, as noted above, the limited availability of PASC and MIS-C diagnosis codes may have not only reduced follow-up but led to models that focus on manifestation associated with more recent viral variants.

There is substantial potential for future work as understanding of PASC develops and more data become available. Training on data with more cases could help address the model’s potential limitation due to heterogeneity of PASC (particularly non-MIS-C PASC). Clinician review of false positives from the model could be used to determine the extent to which PASC-likely patients as determined by the model may have been undiagnosed, and review of false negatives could indicate which features the model is either misinterpreting or failing to detect. Further, our planned validation of the model on sites outside the PEDSnet network will help to evaluate the model’s generalizability. Supplementing the model with other sources of clinical data such as unstructured chart notes could further improve model performance.

Different features may be associated with PASC over different time periods relative to COVID infection. The tree temporal scan statistic has the potential to address this by picking out both features and time windows over which patients with PASC disproportionately have those features occur, and thereby construct a more refined feature space than the fixed windows we used.

## Conclusion

We have applied machine learning methods to available EHR data for children with SARS-CoV-2 infection and subsequent diagnosis of PASC. Using appropriate threshold settings, the model can be used to identify PASC patients in health systems data at higher precision for inclusion in studies or at higher recall in screening for clinical trials, especially in settings where PASC diagnosis codes are used less frequently or less reliably. Analysis of how specific features contribute to the classification process may assist in gaining a better understanding of features that are associated with PASC diagnoses. While additional work is required to improve identification of children with uncommon manifestations of PASC, the current classifier provides a valuable research tool, especially in cases where the scale or provenance of data make it infeasible to determine each patient’s PASC status by direct report. It may also find use in health care delivery, to identify undiagnosed patients who may benefit from further screening to identify and ameliorate effects of PASC.

## Data Availability

All data produced in the present study are available upon reasonable request to the authors.

## Notes

### Competing Interest Statement

Dr. Rao reports prior grant support from GSK and Biofire and is a consultant for Sequiris.
Dr. Jhaveri is a consultant for AstraZeneca, Seqirus, Dynavax, receives an editorial stipend from Elsevier and Pediatric Infectious Diseases Society and royalties from Up To Date/Wolters Kluwer.
Dr. Lee serves on the PASC Advisory Board for United Health Group.
Dr Bailey has received grants from Patient-Centered Outcomes Research Institute
All other authors have nothing to disclose.

### Author Declarations

BRANY IRB gave ethical approval for this work and waived documentation of informed consent.

