## Supplementary materials for "A machine learning-based phenotype for long COVID in children: an EHR-based study from the RECOVER program"

**Supplementary Figure 1: Histogram of cohort entry dates**

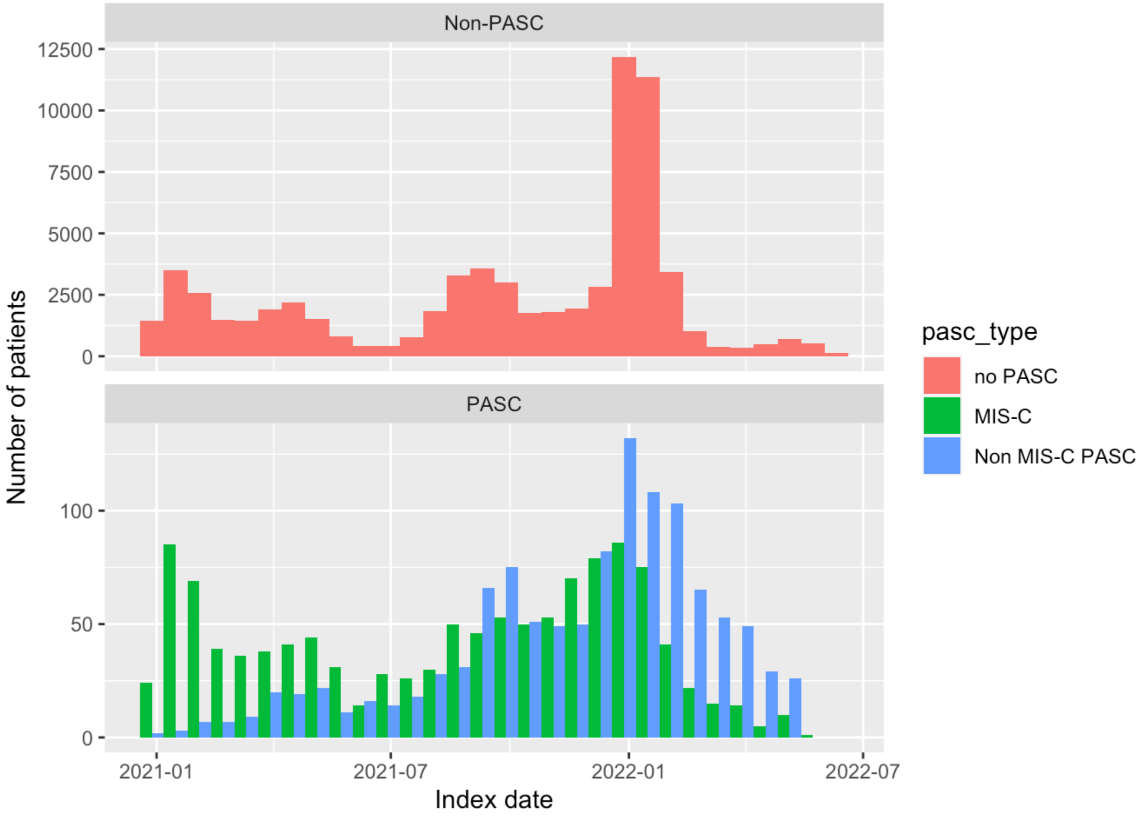

**Supplementary Figures 2a-g** SHapley Additive exPlanation (SHAP) values for model features by class (MIS-C/Non-MIS-C PASC)

Caption: The plots show the most significant features as determined by the sum of SHAP value magnitudes over all samples. For each feature, SHAP values for each patient are plotted, with color representing the feature value (e.g. red if feature was present and blue if absent in case of a binary variable). The SHAP values pictured are for the 3 class classification task and the x axis is interpreted as change in log odds for the corresponding outcome (e.g. MIS-C) as opposed to change in probability (in particular, SHAP values are not confined to be between –1 and 1).

**Figure 2a: Patient-level descriptives**

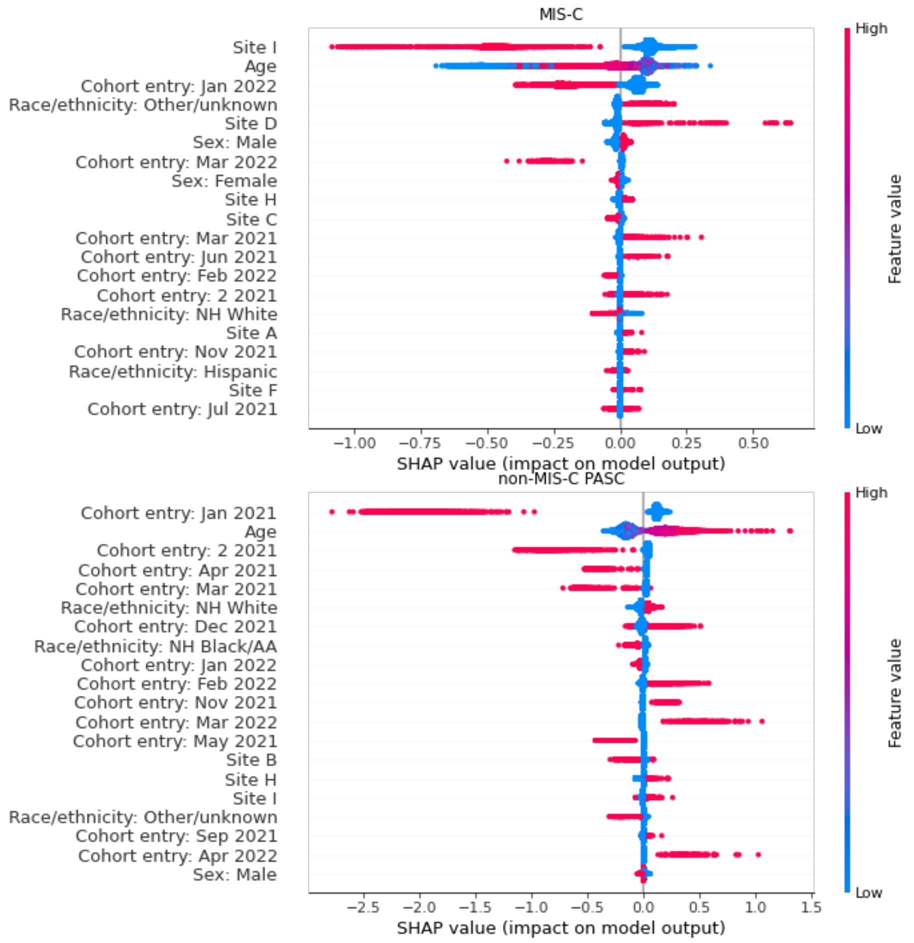

### Figure 2b: Overall utilization

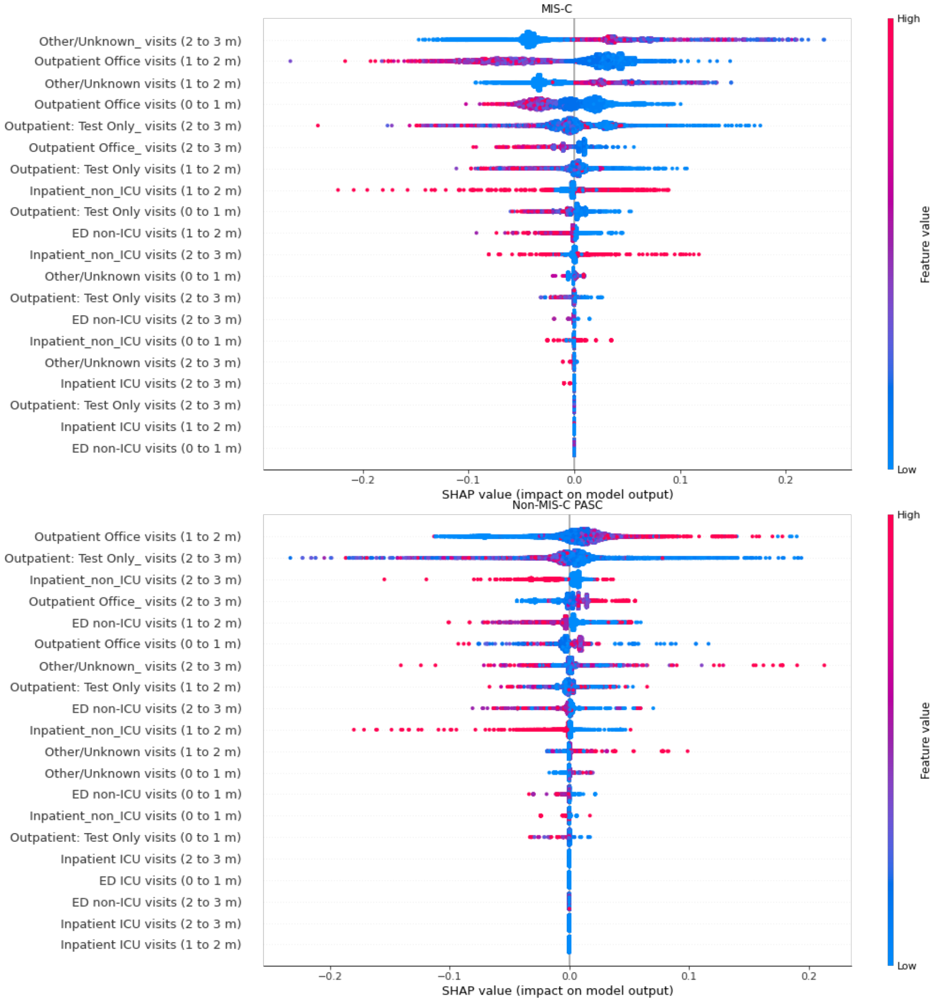

**Figure 2c: Lab counts:**

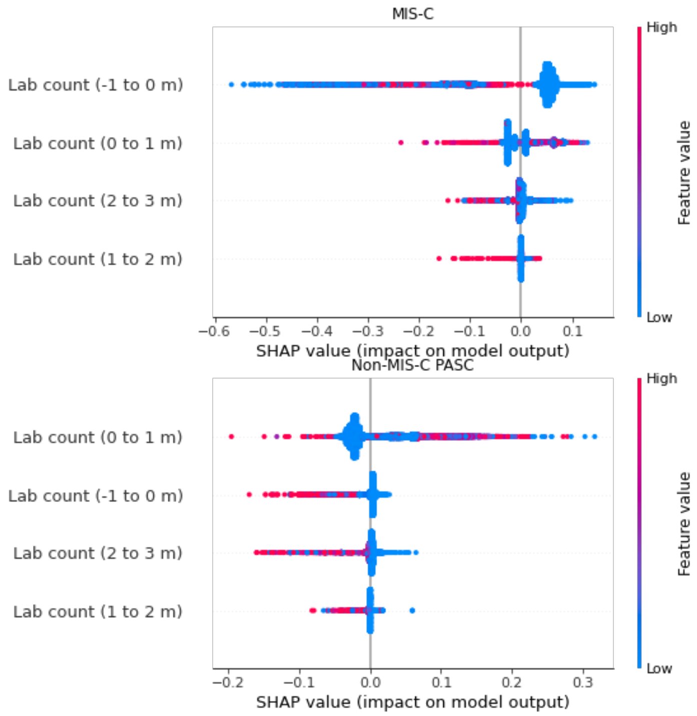

#### Figure 2d: Conditions

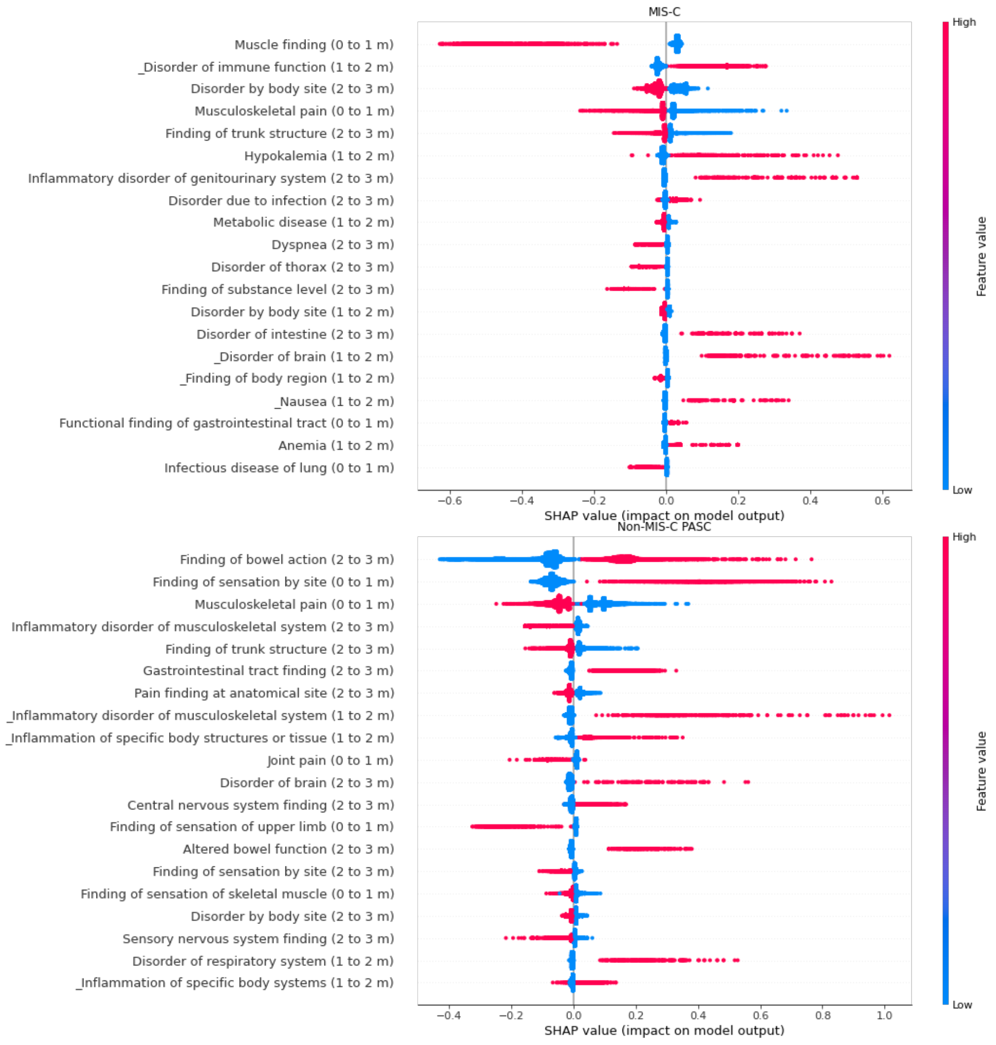

### Figure 2e: Labs

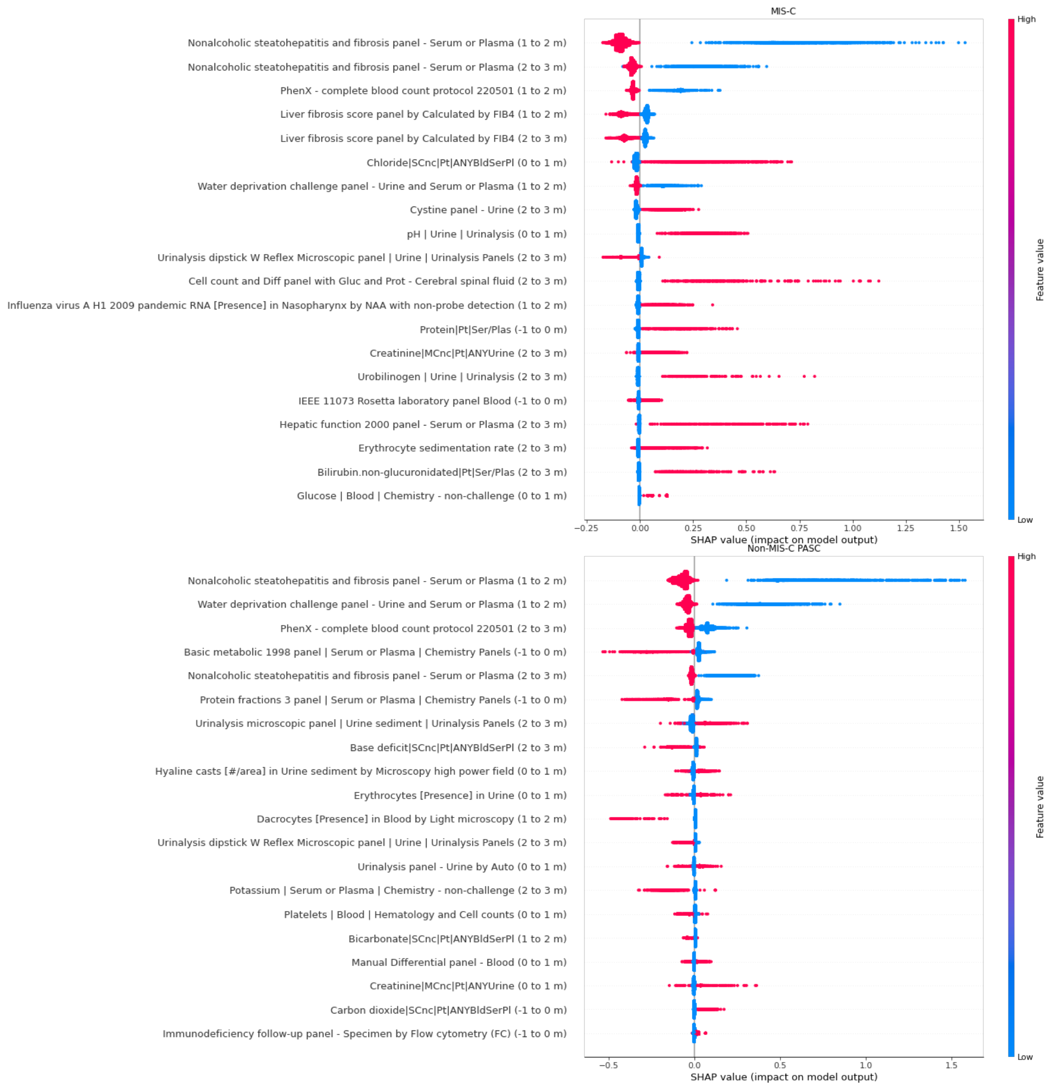

### Figure 2f: Medications

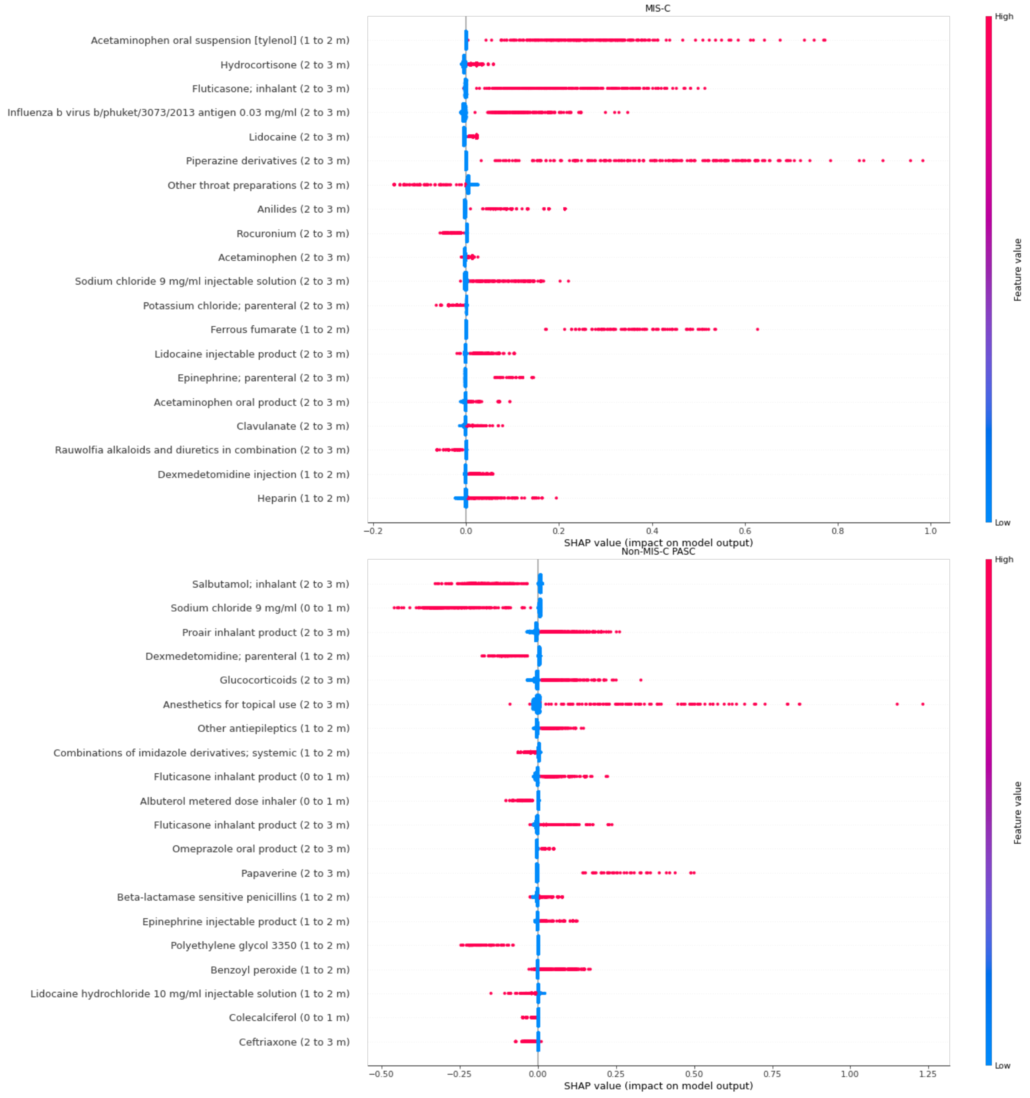

#### Figure 2g: Procedures

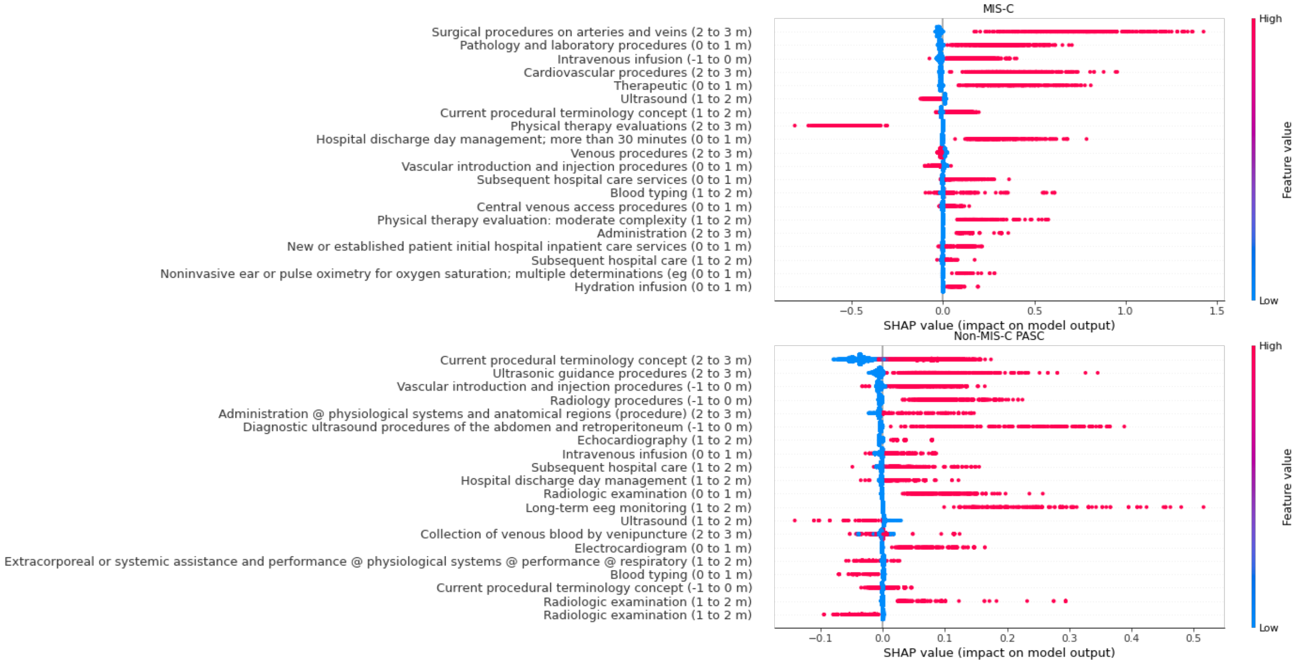

**Supplementary Tables 1a-d:** TreeScan-selected feature clusters

Caption: These tables show the TreeScan-selected cuts for conditions, labs, procedures, and medications. Each row describes the top node which characterizes the cluster. In other words, the node, together with all descendant codes, defines the feature cluster.

**Table 1a: TreeScan-selected condition features**

| Cut | Concept | Tree Level | Log Likelihood Ratio | SNOMED code |
|---|---|---|---|---|
| **1** | **Cardiovascular finding** | 3 | 3,725.46 | 106063007 |
| **2** | **Disorder of cardiovascular system** | 4 | 3,061.78 | 49601007 |
| **3** | **Finding of trunk structure** | 4 | 2,924.57 | 302292003 |
| **4** | **Finding of region of thorax** | 6 | 2,777.48 | 298705000 |
| **5** | **Finding of upper trunk** | 5 | 2,759.92 | 609623002 |
| **6** | **Mediastinal finding** | 7 | 2,734.20 | 301296002 |
| **7** | **Systemic disease** | 3 | 2,604.81 | 56019007 |
| **8** | **Cardiac finding** | 4 | 2,550.69 | 301095005 |
| **9** | **Measurement finding** | 3 | 2,452.99 | 118245000 |
| **10** | **Viscus structure finding** | 3 | 2,301.84 | 406123005 |
| **11** | **Disorder of mediastinum** | 5 | 2,090.49 | 49483002 |
| **12** | **Measurement finding outside reference range** | 4 | 1,919.94 | 442096005 |
| **13** | **Heart disease** | 5 | 1,901.47 | 56265001 |
| **14** | **Disorder of trunk** | 4 | 1,857.30 | 128121009 |
| **15** | **Disorder of thorax** | 6 | 1,827.75 | 118946009 |
| **16** | **Disorder of thoracic segment of trunk** | 5 | 1,810.61 | 609622007 |
| **17** | **Metabolic disease** | 3 | 1,616.32 | 75934005 |
| **18** | **Finding of body region** | 3 | 1,519.64 | 301857004 |
| **19** | **Disorder by body site** | 3 | 1,476.00 | 123946008 |
| **20** | **Disorder of fluid AND/OR electrolyte** | 4 | 1,431.94 | 76314005 |
| **21** | **Disorder of body system** | 4 | 1,356.31 | 362965005 |
| **22** | **Hematopoietic system finding** | 3 | 1,157.97 | 106200001 |
| **23** | **Abnormal blood cell count** | 5 | 1,147.99 | 762656009 |
| **24** | **Sensory nervous system finding** | 3 | 1,109.79 | 106147001 |
| **25** | **Pain / sensation finding** | 4 | 1,057.77 | 276435006 |
| **26** | **Finding of substance level** | 4 | 1,054.38 | 785671009 |
| **27** | **Finding of sensation by site** | 3 | 1,042.94 | 699697007 |
| **28** | **Pain** | 5 | 1,028.93 | 22253000 |
| **29** | **Disorder of electrolytes** | 5 | 1,008.45 | 237840007 |
| **30** | **Pain finding at anatomical site** | 4 | 976.00 | 279001004 |
| **31** | **Protein level - finding** | 5 | 924.66 | 365799007 |
| **32** | **Measurement finding below reference range** | 5 | 913.90 | 442686002 |
| **33** | **Shock** | 4 | 881.37 | 27942005 |
| **34** | **Structural disorder of heart** | 6 | 860.33 | 128599005 |
| **35** | **Hypo-osmolality and or hyponatremia** | 6 | 816.67 | 267447008 |
| **36** | **Inflammatory disorder of the cardiovascular system** | 5 | 807.78 | 128998007 |

| | | | | |
|---|---|---|---|---|
| **37** | **Measurement finding above reference range** | 5 | 781.51 | 442756004 |
| **38** | **Disorder of cardiac function** | 6 | 711.55 | 105981003 |
| **39** | **Disorder of cellular component of blood** | 3 | 703.94 | 414022008 |
| **40** | **Dyspnea** | 7 | 673.29 | 267036007 |
| **41** | **Finding of abdomen** | 6 | 670.89 | 609624008 |
| **42** | **Acute disease of cardiovascular system** | 4 | 670.64 | 128487001 |
| **43** | **Finding of abdominopelvic segment of trunk** | 5 | 642.54 | 822987005 |
| **44** | **General well-being finding** | 4 | 614.22 | 365275006 |
| **45** | **Myocardial finding** | 5 | 598.85 | 251052000 |
| **46** | **Myocardial disease** | 6 | 596.70 | 57809008 |
| **47** | **Lymphadenopathy** | 6 | 593.83 | 30746006 |
| **48** | **General body state finding** | 4 | 584.38 | 82832008 |
| **49** | **Pain of truncal structure** | 5 | 555.27 | 301366005 |
| **50** | **Cardiovascular measurement - finding** | 4 | 554.11 | 366157005 |
| **51** | **Ease of respiration - finding** | 5 | 553.57 | 366139009 |
| **52** | **Difficulty breathing** | 6 | 553.57 | 230145002 |
| **53** | **Disorder of lymph node** | 6 | 549.10 | 76616003 |
| **54** | **Blood vessel finding** | 4 | 547.52 | 21829004 |
| **55** | **Low blood pressure** | 5 | 521.61 | 45007003 |
| **56** | **Inflammatory disorder** | 3 | 518.97 | 128139000 |
| **57** | **Arterial finding** | 5 | 517.18 | 248718009 |
| **58** | **General finding of soft tissue** | 3 | 510.78 | 248402002 |
| **59** | **General finding of observation of patient** | 3 | 502.01 | 118222006 |
| **60** | **Finding of cellular component of blood** | 3 | 501.73 | 414253004 |
| **61** | **Finding of heart rate** | 4 | 496.76 | 301113001 |
| **62** | **Cardiac rhythm AND/OR rate finding** | 3 | 496.40 | 106066004 |
| **63** | **Finding of blood, lymphatics and immune system** | 3 | 496.13 | 299691001 |
| **64** | **Acute nephropathy** | 5 | 488.73 | 58574008 |
| **65** | **Vascular disorder** | 5 | 486.88 | 27550009 |
| **66** | **Carditis** | 6 | 482.78 | 399617002 |
| **67** | **Disorder of artery** | 6 | 465.24 | 359557001 |
| **68** | **Platelet count abnormal** | 5 | 458.41 | 165558001 |
| **69** | **Platelet count - finding** | 4 | 457.95 | 365632008 |
| **70** | **Platelet finding** | 4 | 457.04 | 250311007 |
| **71** | **Collagen disease** | 5 | 453.47 | 81573002 |
| **72** | **Acute genitourinary disorder** | 4 | 446.89 | 175176000 |
| **73** | **Hemostatic system finding** | 4 | 441.48 | 128153000 |
| **74** | **Myocarditis** | 7 | 435.02 | 50920009 |
| **75** | **Heart valve regurgitation** | 7 | 423.33 | 40445007 |
| **76** | **Fluid volume disorder** | 5 | 410.88 | 1860003 |
| **77** | **Platelet count below reference range** | 6 | 408.94 | 415116008 |
| **78** | **Non-rheumatic heart valve disorder** | 7 | 401.54 | 274097009 |
| **79** | **Disorder of abdomen** | 6 | 397.53 | 118948005 |
| **80** | **Cardiac arrhythmia** | 6 | 397.25 | 698247007 |
| **81** | **Disorder of retroperitoneum** | 7 | 380.47 | 734045002 |
| **82** | **Acute febrile mucocutaneous lymph node syndrome** | 4 | 375.96 | 75053002 |

| | | | | |
|---|---|---|---|---|
| **83** | **Vasculitis of medium sized vessel** | 7 | 373.14 | 724598001 |
| **84** | **Acute renal impairment** | 5 | 372.85 | 236424009 |
| **85** | **Acute renal failure syndrome** | 6 | 367.24 | 14669001 |
| **86** | **Primary systemic arteritis** | 5 | 366.38 | 407529009 |
| **87** | **Disorder of the urinary system** | 6 | 365.81 | 128606002 |
| **88** | **Heart valve finding** | 5 | 365.49 | 301098007 |
| **89** | **Abdominal organ finding** | 4 | 364.37 | 249561001 |
| **90** | **Disorder of soft tissue** | 4 | 364.09 | 19660004 |
| **91** | **Disorder of abdominopelvic segment of trunk** | 5 | 361.81 | 822988000 |
| **92** | **Disorder of lymphatic system** | 5 | 361.65 | 362971004 |
| **93** | **Heart valve disorder** | 6 | 358.47 | 368009 |
| **94** | **Kidney disease** | 6 | 356.80 | 90708001 |
| **95** | **White blood cell count abnormal** | 6 | 354.65 | 165509000 |
| **96** | **Disorder of lymphoid system** | 6 | 354.43 | 111590001 |
| **97** | **Head and neck lymphadenopathy** | 5 | 353.38 | 704281009 |
| **98** | **Cervical lymphadenopathy** | 5 | 351.53 | 127086001 |
| **99** | **Disorder of immune structure** | 5 | 351.30 | 414030009 |
| **100** | **Kidney finding** | 5 | 351.13 | 249578005 |
| **101** | **Disorder of kidney and/or ureter** | 7 | 350.14 | 443820000 |
| **102** | **Lymphadenopathy of head AND/OR neck** | 6 | 349.30 | 425061006 |
| **103** | **Tachycardia** | 5 | 347.46 | 3424008 |
| **104** | **RBC count abnormal** | 6 | 346.22 | 165427000 |
| **105** | **RBC count low** | 6 | 343.89 | 165423001 |
| **106** | **Systemic vasculitis** | 4 | 343.63 | 46956008 |
| **107** | **Renal failure syndrome** | 5 | 343.45 | 42399005 |
| **108** | **Urinary system finding** | 7 | 342.79 | 106098005 |
| **109** | **Arteritis** | 7 | 342.60 | 52089001 |
| **110** | **Hemoglobin low** | 5 | 338.25 | 165397008 |
| **111** | **Hemoglobin level outside reference range** | 4 | 338.25 | 441793007 |
| **112** | **Disorder of coronary artery** | 6 | 328.63 | 414024009 |
| **113** | **Coronary artery finding** | 5 | 326.00 | 251015000 |
| **114** | **Vasculitis** | 6 | 318.23 | 31996006 |
| **115** | **Cytopenia** | 4 | 315.76 | 50820005 |
| **116** | **Erythropenia** | 5 | 314.19 | 62574001 |
| **117** | **Anemia** | 4 | 314.19 | 271737000 |
| **118** | **Disorder of hemostatic system** | 3 | 305.85 | 362970003 |
| **119** | **Distention of artery** | 6 | 302.10 | 248494009 |
| **120** | **Disorder of hematopoietic structure** | 5 | 298.87 | 414027002 |
| **121** | **Chest pain** | 6 | 298.50 | 29857009 |
| **122** | **Energy and stamina finding** | 4 | 296.44 | 359752005 |
| **123** | **Metabolic finding** | 3 | 295.75 | 106089007 |
| **124** | **Acute disease** | 3 | 293.73 | 2704003 |
| **125** | **White blood cell disorder** | 4 | 291.92 | 54097007 |
| **126** | **Heart failure** | 7 | 288.16 | 84114007 |
| **127** | **Distention of blood vessel** | 5 | 287.70 | 248493003 |
| **128** | **Aneurysm of coronary vessels** | 6 | 286.00 | 50570003 |

| | | | | |
|---|---|---|---|---|
| **129** | **Abdominal pain** | 5 | 284.42 | 21522001 |
| **130** | **Finding of sensation of abdomen** | 4 | 280.61 | 247330004 |
| **131** | **Finding of respiration** | 4 | 278.53 | 301282008 |
| **132** | **Aneurysm of artery of trunk** | 5 | 273.48 | 301433005 |
| **133** | **Cardiogenic shock** | 5 | 271.18 | 89138009 |
| **134** | **Disease affecting entire cardiovascular system** | 5 | 271.18 | 105980002 |
| **135** | **Aneurysm** | 5 | 267.05 | 432119003 |
| **136** | **Fatigue** | 5 | 265.46 | 84229001 |
| **137** | **Non-malignant white cell disorder** | 5 | 264.61 | 234414004 |
| **138** | **Dehydration** | 6 | 263.91 | 34095006 |
| **139** | **Lymphocytoid disorder** | 6 | 262.90 | 234428005 |
| **140** | **Lymphocytopenia** | 6 | 262.90 | 48813009 |
| **141** | **Disorder of immune function** | 3 | 262.75 | 414029004 |
| **142** | **Arterial aneurysm** | 6 | 261.06 | 233981004 |
| **143** | **Enzyme level - finding** | 6 | 260.82 | 365767001 |
| **144** | **C-reactive protein abnormal** | 5 | 260.05 | 166584001 |
| **145** | **Pleural effusion** | 7 | 258.32 | 60046008 |
| **146** | **Renal impairment** | 4 | 257.96 | 236423003 |
| **147** | **Disorder of the genitourinary system** | 5 | 257.15 | 42030000 |
| **148** | **Lymphocyte disorder** | 5 | 253.24 | 3239007 |
| **149** | **General problem AND/OR complaint** | 4 | 252.16 | 105721009 |
| **150** | **Gastrointestinal tract finding** | 4 | 250.15 | 386618008 |
| **151** | **Lymphocyte count abnormal** | 7 | 248.22 | 165534000 |
| **152** | **Conduction disorder of the heart** | 7 | 246.77 | 44808001 |
| **153** | **Disorder of pleura and pleural cavity** | 6 | 241.69 | 233635001 |
| **154** | **Urogenital finding** | 6 | 236.71 | 118238000 |
| **155** | **Mitral valve disorder** | 7 | 233.26 | 11851006 |
| **156** | **Mitral valve finding** | 6 | 233.26 | 301101006 |
| **157** | **Systemic arterial finding** | 6 | 233.23 | 301139003 |
| **158** | **Electrocardiogram finding** | 3 | 232.45 | 301120008 |
| **159** | **Electrocardiogram abnormal** | 4 | 231.64 | 102594003 |
| **160** | **Mitral valve regurgitation** | 8 | 231.11 | 48724000 |
| **161** | **Organ dysfunction syndrome** | 4 | 231.06 | 238147009 |
| **162** | **Elevated C-reactive protein** | 6 | 228.29 | 119971000119104 |
| **163** | **Increased globulin** | 6 | 226.39 | 124019005 |
| **164** | **Non-rheumatic mitral regurgitation** | 9 | 225.55 | 194978002 |
| **165** | **Sepsis** | 4 | 224.01 | 91302008 |
| **166** | **Disorder of protein metabolism** | 4 | 223.70 | 363090004 |
| **167** | **Disorder of plasma protein metabolism** | 5 | 223.70 | 147211000119101 |
| **168** | **Disorder of connective tissue** | 4 | 220.73 | 105969002 |
| **169** | **Non-rheumatic mitral valve disease** | 8 | 220.67 | 708121009 |
| **170** | **Disorder characterized by fever** | 3 | 220.66 | 416113008 |
| **171** | **Globulin level - finding** | 6 | 213.42 | 365805001 |
| **172** | **Dizziness** | 5 | 208.81 | 404640003 |

| | | | | |
|---|---|---|---|---|
| **173** | **Erythrocyte sedimentation rate - finding** | 4 | 206.94 | 365649001 |
| **174** | **ESR abnormal** | 5 | 206.94 | 165465007 |
| **175** | **ESR raised** | 6 | 206.94 | 165468009 |
| **176** | **Acute heart disease** | 5 | 206.89 | 127337006 |
| **177** | **Coag./bleeding tests abnormal** | 4 | 206.21 | 165563002 |
| **178** | **Digestive system finding** | 3 | 204.26 | 386617003 |
| **179** | **Functional finding of gastrointestinal tract** | 5 | 201.79 | 300358007 |
| **180** | **Pericardial finding** | 4 | 199.33 | 301123005 |
| **181** | **Disorder of pericardium** | 5 | 199.33 | 55855009 |
| **182** | **Vital signs finding** | 5 | 193.87 | 118227000 |
| **183** | **Leukopenia** | 5 | 192.59 | 84828003 |
| **184** | **Disorder of lower respiratory system** | 5 | 192.14 | 128272009 |
| **185** | **Hypervolemia** | 6 | 189.46 | 21639008 |
| **186** | **Abnormal body temperature** | 6 | 179.67 | 123979008 |
| **187** | **Body temperature finding** | 5 | 179.27 | 105723007 |
| **188** | **Blood coagulation disorder** | 4 | 179.22 | 64779008 |
| **189** | **Lung finding** | 4 | 178.32 | 301230006 |
| **190** | **Body temperature above reference range** | 7 | 177.46 | 50177009 |
| **191** | **Fever** | 8 | 177.33 | 386661006 |
| **192** | **Temperature-associated finding** | 4 | 176.71 | 301343009 |
| **193** | **Hypokalemia** | 7 | 170.80 | 43339004 |
| **194** | **Platelet disorder** | 4 | 169.79 | 22716005 |
| **195** | **Pericardial effusion** | 6 | 168.99 | 373945007 |
| **196** | **Respiratory failure** | 6 | 168.71 | 409622000 |
| **197** | **Disorder characterized by pain** | 3 | 168.02 | 373673007 |
| **198** | **Disorder of cardiac ventricle** | 6 | 167.54 | 415991003 |
| **199** | **Chronic pain** | 6 | 166.94 | 82423001 |
| **200** | **Potassium disorder** | 6 | 166.59 | 24529006 |
| **201** | **Respiratory insufficiency** | 5 | 165.70 | 409623005 |
| **202** | **Musculoskeletal pain** | 4 | 164.91 | 279069000 |
| **203** | **Injury of kidney** | 6 | 164.48 | 40095003 |
| **204** | **Injury of urinary organ** | 7 | 163.61 | 733386005 |
| **205** | **Increased blood leukocyte number** | 6 | 162.95 | 414478003 |
| **206** | **Dyspnea on exertion** | 8 | 162.93 | 60845006 |
| **207** | **Headache disorder** | 4 | 162.87 | 230461009 |
| **208** | **D-dimer above reference range** | 5 | 155.68 | 449830004 |
| **209** | **Urogenital injury** | 6 | 155.63 | 283879004 |
| **210** | **Heart block** | 8 | 153.92 | 233916004 |
| **211** | **Septic shock** | 5 | 153.75 | 76571007 |
| **212** | **Internal injury of abdominal organ** | 5 | 153.42 | 49011004 |
| **213** | **Pericardial effusion - noninflammatory** | 7 | 150.78 | 194970009 |
| **214** | **Bradycardia** | 5 | 150.40 | 48867003 |
| **215** | **Injury of internal organ** | 4 | 148.52 | 105612003 |
| **216** | **Conjunctival finding** | 4 | 147.43 | 246875002 |
| **217** | **Edema of trunk** | 4 | 146.62 | 301867009 |
| **218** | **Ocular surface finding** | 6 | 146.17 | 246869006 |

| | | | | |
|---|---|---|---|---|
| **219** | **Anterior segment finding** | 5 | 145.62 | 418727003 |
| **220** | **Pulmonary edema** | 5 | 145.46 | 19242006 |
| **221** | **Blood chemistry abnormal** | 5 | 145.28 | 166318006 |
| **222** | **Abnormal finding on evaluation procedure** | 3 | 144.93 | 442618008 |
| **223** | **Lymphadenitis** | 6 | 144.75 | 19471005 |
| **224** | **Infectious disease of heart** | 6 | 143.15 | 128403000 |
| **225** | **Acute respiratory failure** | 5 | 143.14 | 65710008 |
| **226** | **Respiratory finding** | 3 | 141.73 | 106048009 |
| **227** | **Elevated liver enzymes level** | 6 | 141.63 | 707724006 |
| **228** | **Liver enzyme levels - finding** | 7 | 140.97 | 365769003 |
| **229** | **Liver enzymes abnormal** | 5 | 140.97 | 166643006 |
| **230** | **Orbit finding** | 4 | 139.71 | 246912006 |
| **231** | **Injury of abdomen** | 6 | 137.57 | 128069005 |
| **232** | **Inflammatory disorder of the eye** | 6 | 133.92 | 128295000 |
| **233** | **Tricuspid valve disorder** | 7 | 133.58 | 20721001 |
| **234** | **Tricuspid valve finding** | 6 | 133.29 | 301108000 |
| **235** | **Inflammatory disorder of immune system** | 6 | 133.12 | 363177008 |
| **236** | **Tricuspid valve disorder, non-rheumatic** | 8 | 132.95 | 194989009 |
| **237** | **Disorder of conjunctiva** | 5 | 132.43 | 59698003 |
| **238** | **Disorder of anterior segment of eye** | 6 | 132.01 | 128535002 |
| **239** | **Infectious disease of cardiovascular system** | 5 | 130.84 | 128402005 |
| **240** | **Inflammation of specific body systems** | 5 | 130.34 | 363171009 |
| **241** | **Finding related to awareness of heart beat** | 5 | 129.71 | 366182005 |
| **242** | **Palpitations** | 6 | 129.71 | 80313002 |
| **243** | **Inflammation of specific body structures or tissue** | 4 | 129.56 | 363170005 |
| **244** | **Disorder of lung** | 5 | 129.47 | 19829001 |
| **245** | **Tricuspid valve regurgitation** | 8 | 128.92 | 111287006 |
| **246** | **Tricuspid incompetence, non-rheumatic** | 9 | 128.09 | 194990000 |
| **247** | **Disorder of gastrointestinal tract** | 5 | 127.75 | 119292006 |
| **248** | **Lower respiratory tract finding** | 4 | 127.74 | 301226008 |
| **249** | **Kidney lesion** | 6 | 127.08 | 79131000119100 |
| **250** | **Thrombocytopenic disorder** | 4 | 126.92 | 302215000 |
| **251** | **Disorder of orbit proper** | 5 | 125.37 | 371436007 |
| **252** | **Conjunctivitis** | 6 | 120.63 | 9826008 |
| **253** | **Inflammation of orbit** | 6 | 120.03 | 95770005 |
| **254** | **Chronic headache disorder** | 4 | 119.59 | 431237007 |
| **255** | **Mental state finding** | 4 | 118.93 | 36456004 |
| **256** | **Tricuspid valve lesion** | 7 | 118.41 | 301109008 |
| **257** | **Finding of left ventricle** | 5 | 117.15 | 301096006 |
| **258** | **Disorder of acid-base balance** | 4 | 115.79 | 26436007 |
| **259** | **Disorder of digestive system** | 4 | 115.12 | 53619000 |
| **260** | **Atrioventricular conduction disorder** | 8 | 114.36 | 418341009 |
| **261** | **Atrioventricular block** | 7 | 114.36 | 233917008 |
| **262** | **Acute heart failure** | 6 | 112.84 | 56675007 |
| **263** | **Pain in limb** | 5 | 112.14 | 90834002 |

| | | | | |
|---|---|---|---|---|
| **264** | **Inflammation of specific body organs** | 5 | 111.45 | 363169009 |
| **265** | **Giddiness** | 5 | 111.40 | 404641004 |
| **266** | **Dizziness and giddiness** | 6 | 111.40 | 271789005 |
| **267** | **Eruption** | 7 | 111.21 | 271807003 |
| **268** | **Soft tissue lesion** | 5 | 110.40 | 239953001 |
| **269** | **Finding of defecation** | 6 | 108.76 | 300373008 |
| **270** | **Myocardial dysfunction** | 7 | 108.65 | 233928007 |
| **271** | **Anomaly of eye** | 6 | 108.47 | 11131000119108 |
| **272** | **Finding of bowel action** | 7 | 106.38 | 366256008 |
| **273** | **Altered bowel function** | 8 | 106.32 | 88111009 |
| **274** | **Skin AND/OR mucosa finding** | 3 | 106.26 | 415531008 |
| **275** | **Headache** | 5 | 103.14 | 25064002 |
| **276** | **Mental state, behavior and/or psychosocial function finding** | 3 | 102.76 | 384821006 |
| **277** | **Imaging finding** | 3 | 102.40 | 365853002 |
| **278** | **Disorder of right atrium** | 7 | 102.36 | 472762000 |
| **279** | **Disorder of left cardiac ventricle** | 6 | 100.37 | 415993000 |
| **280** | **Pain in lower limb** | 5 | 98.00 | 10601006 |
| **281** | **Finding of sensation of lower limb** | 4 | 95.82 | 713314006 |
| **282** | **Hyponatremia** | 7 | 93.96 | 89627008 |
| **283** | **Atrial cardiopathy** | 6 | 93.31 | 870575001 |
| **284** | **Pneumonia** | 7 | 92.50 | 233604007 |
| **285** | **Disorder of phosphate, calcium and vitamin D metabolism** | 5 | 92.47 | 237879001 |
| **286** | **Lung consolidation** | 6 | 92.38 | 95436008 |
| **287** | **Immune system finding** | 4 | 90.48 | 106182000 |
| **288** | **Cardiovascular function finding** | 4 | 89.62 | 301458000 |
| **289** | **Globe finding** | 4 | 89.29 | 246915008 |
| **290** | **Joint pain** | 5 | 88.54 | 57676002 |
| **291** | **Mucosal finding** | 4 | 86.46 | 128145008 |
| **292** | **Radiologic finding** | 4 | 85.82 | 118247008 |
| **293** | **Imaging result abnormal** | 4 | 85.75 | 408574004 |
| **294** | **Localized enlarged lymph nodes** | 7 | 85.30 | 274744005 |
| **295** | **Finding of sensation of joint** | 4 | 84.50 | 298249004 |
| **296** | **Acute injury of kidney** | 6 | 83.52 | 14350001000004108 |
| **297** | **Disorder of eye** | 5 | 83.12 | 371405004 |
| **298** | **Injury of trunk** | 5 | 82.93 | 48125009 |
| **299** | **Vomiting** | 6 | 82.23 | 422400008 |
| **300** | **Sodium disorder** | 6 | 81.79 | 123807007 |
| **301** | **Disorder of ocular adnexa** | 6 | 81.21 | 118941004 |
| **302** | **Finding of vomiting** | 6 | 80.42 | 300359004 |
| **303** | **Disorder of mineral metabolism** | 4 | 79.57 | 45744005 |
| **304** | **Finding of neck region** | 5 | 77.83 | 298378000 |
| **305** | **Disorder of mucous membrane** | 5 | 77.43 | 95351003 |

| | | | | |
|---|---|---|---|---|
| **306** | **Infective pneumonia** | 7 | 77.11 | 312342009 |
| **307** | **Finding of lung field** | 5 | 76.60 | 301231005 |
| **308** | **Lung field abnormal** | 6 | 76.60 | 274710003 |
| **309** | **Hypertensive disorder** | 5 | 76.50 | 38341003 |
| **310** | **Disorder of blood gas** | 4 | 75.12 | 238157005 |
| **311** | **Leukocytosis** | 5 | 75.04 | 111583006 |
| **312** | **Diarrhea** | 9 | 74.95 | 62315008 |
| **313** | **Cardiomegaly** | 7 | 74.93 | 8186001 |
| **314** | **Disorder characterized by edema** | 3 | 74.26 | 118654009 |
| **315** | **Nausea** | 5 | 71.74 | 422587007 |
| **316** | **Vascular headache** | 5 | 71.62 | 128187005 |
| **317** | **Pain of cardiovascular structure** | 4 | 71.52 | 301358001 |
| **318** | **Disorder of digestive tract** | 5 | 71.51 | 84410009 |
| **319** | **Disorder of soft tissue of head** | 5 | 69.56 | 280131007 |
| **320** | **Disease of liver** | 5 | 69.52 | 235856003 |
| **321** | **Migraine** | 6 | 69.44 | 37796009 |
| **322** | **Malaise** | 5 | 69.30 | 367391008 |
| **323** | **Inflammatory disease of mucous membrane** | 6 | 66.91 | 95361005 |
| **324** | **Skin or mucosa lesion** | 4 | 65.90 | 247440002 |
| **325** | **Joint finding** | 4 | 65.76 | 118952005 |
| **326** | **Liver finding** | 4 | 65.27 | 249565005 |
| **327** | **Disease due to Gram-positive bacteria** | 5 | 64.20 | 371582002 |
| **328** | **Radiology result abnormal** | 5 | 63.94 | 168501001 |
| **329** | **Infectious disease of lung** | 6 | 63.63 | 128601007 |
| **330** | **Generalized abdominal pain** | 6 | 63.54 | 102614006 |
| **331** | **Acidosis** | 5 | 62.12 | 51387008 |
| **332** | **Chronic cough** | 6 | 61.61 | 68154008 |
| **333** | **Bacterial infectious disease** | 4 | 60.79 | 87628006 |
| **334** | **Lesion of skin and/or skin-associated mucous membrane** | 5 | 60.11 | 714974000 |
| **335** | **Hypoxia** | 5 | 59.30 | 389086002 |
| **336** | **Hypoxemia** | 5 | 59.23 | 389087006 |
| **337** | **Disorder of upper gastrointestinal tract** | 6 | 58.95 | 119291004 |
| **338** | **Skin lesion** | 6 | 58.02 | 95324001 |
| **339** | **Disturbance of consciousness** | 8 | 57.58 | 3006004 |
| **340** | **Level of consciousness - finding** | 7 | 57.49 | 365931003 |
| **341** | **Consciousness related finding** | 6 | 57.41 | 106167005 |
| **342** | **Disorder due to infection** | 3 | 57.33 | 40733004 |
| **343** | **Disease due to Gram-positive coccus** | 6 | 56.96 | 408637006 |
| **344** | **Abdominal organomegaly** | 5 | 56.23 | 714254003 |
| **345** | **Disorder of respiratory system** | 4 | 56.22 | 50043002 |
| **347** | **Atelectasis** | 6 | 55.39 | 46621007 |
| **348** | **Upper abdominal pain** | 6 | 55.16 | 83132003 |
| **349** | **Disorder of neck** | 4 | 54.69 | 118939000 |
| **350** | **Obesity** | 3 | 54.20 | 414916001 |
| **351** | **Disorder of eye region** | 5 | 53.66 | 371409005 |

| | | | | |
|---|---|---|---|---|
| **352** | **Inflammatory disorder of musculoskeletal system** | 5 | 52.68 | 363179006 |
| **353** | **Muscle pain** | 5 | 51.33 | 68962001 |
| **354** | **Finding of sensation of skeletal muscle** | 4 | 51.22 | 298287007 |
| **355** | **Thrombosis** | 5 | 51.02 | 439127006 |
| **356** | **Musculoskeletal finding** | 3 | 48.51 | 106028002 |
| **357** | **Disease due to Gram-negative bacteria** | 5 | 48.30 | 371583007 |
| **358** | **Disease due to Gram-negative bacillus** | 6 | 47.70 | 408638001 |
| **359** | **Connective tissue disorder by body site** | 5 | 46.66 | 363044007 |
| **360** | **Disorder of digestive organ** | 5 | 46.56 | 76712006 |
| **361** | **Disorder of phosphorus metabolism** | 6 | 46.42 | 87049008 |
| **362** | **Nausea and vomiting** | 6 | 44.41 | 16932000 |
| **363** | **Emotional state finding** | 5 | 43.30 | 106126000 |
| **364** | **Pain in left lower limb** | 6 | 43.17 | 287047008 |
| **365** | **Urinary tract infectious disease** | 6 | 42.84 | 68566005 |
| **366** | **Central abdominal pain** | 6 | 42.76 | 162046002 |
| **367** | **Migraine without aura** | 6 | 42.00 | 56097005 |
| **368** | **Pain in right lower limb** | 6 | 41.90 | 287048003 |
| **369** | **Pulmonary valve finding** | 6 | 40.93 | 301104003 |
| **370** | **Anxiety** | 6 | 39.49 | 48694002 |
| **371** | **Observation of sensation** | 5 | 39.40 | 271712005 |
| **372** | **Pneumonitis** | 6 | 39.29 | 205237003 |
| **373** | **Infection due to Enterobacteriaceae** | 7 | 39.22 | 128945009 |
| **374** | **Disorder of nervous system** | 5 | 39.21 | 118940003 |
| **375** | **Muscle finding** | 3 | 39.15 | 106030000 |
| **376** | **Constipation** | 7 | 38.73 | 14760008 |
| **377** | **Decreased level of consciousness** | 9 | 38.56 | 443371007 |
| **378** | **Loss of consciousness** | 10 | 38.56 | 419045004 |
| **379** | **Consciousness and/or awareness finding** | 5 | 38.24 | 365929007 |
| **380** | **Pulmonary valve lesion** | 7 | 37.99 | 301105002 |
| **381** | **Pulmonary valve disorder** | 7 | 37.58 | 76267008 |
| **382** | **Backache** | 5 | 37.38 | 161891005 |
| **383** | **Epigastric pain** | 4 | 37.23 | 79922009 |
| **384** | **Viral pneumonia** | 7 | 37.21 | 75570004 |
| **385** | **Cardiac auscultation finding** | 4 | 37.14 | 106070007 |
| **386** | **Finding of heart sounds** | 5 | 37.14 | 301126002 |
| **387** | **Essential hypertension** | 6 | 37.04 | 59621000 |
| **388** | **Thrombosis of blood vessel** | 6 | 37.00 | 439129009 |
| **389** | **Abnormality of pulmonary valve** | 7 | 36.73 | 448643005 |
| **390** | **Finding by auscultation** | 3 | 36.53 | 118241009 |
| **391** | **Streptococcal infectious disease** | 7 | 36.28 | 85769006 |
| **392** | **Heart murmur** | 5 | 34.97 | 88610006 |
| **393** | **Murmur** | 4 | 34.97 | 414786004 |
| **394** | **Skin finding** | 4 | 34.28 | 106076001 |
| **395** | **Bleeding from nose** | 3 | 34.24 | 249366005 |
| **396** | **Chronic disease** | 3 | 34.03 | 27624003 |
| **397** | **Disorder of vein** | 6 | 33.60 | 90507008 |

| | | | | |
|---|---|---|---|---|
| **398** | **Venous thrombosis** | 7 | 33.48 | 111293003 |
| **399** | **Infectious disease of genitourinary system** | 5 | 33.25 | 189176002 |
| **400** | **Finding related to ability to move** | 6 | 33.06 | 364833005 |
| **401** | **Syncope** | 5 | 33.02 | 271594007 |
| **402** | **Venous finding** | 5 | 32.80 | 248727005 |
| **403** | **Eye / vision finding** | 3 | 32.51 | 118235002 |
| **404** | **Autoimmune disease** | 4 | 31.94 | 85828009 |
| **405** | **Functional finding** | 3 | 31.67 | 118228005 |
| **406** | **Disorder of blood vessels of thorax** | 6 | 31.62 | 373434004 |
| **407** | **Bowel finding** | 5 | 31.31 | 249562008 |
| **408** | **Finding of urine substance level** | 5 | 31.12 | 785672002 |
| **409** | **Lesion of lung** | 5 | 30.27 | 301232003 |
| **410** | **Disorder of intestine** | 6 | 30.24 | 85919009 |
| **411** | **Disorder of lower gastrointestinal tract** | 6 | 30.17 | 79787007 |
| **412** | **Herpesvirus infection** | 5 | 29.24 | 23513009 |
| **413** | **Pneumonia caused by Human coronavirus** | 7 | 28.59 | 713084008 |
| **414** | **Visual system disorder** | 4 | 28.51 | 128127008 |
| **415** | **Integumentary system finding** | 3 | 28.40 | 106077005 |
| **416** | **Disorder of stomach** | 6 | 27.99 | 29384001 |
| **417** | **Stomach finding** | 5 | 27.69 | 118434000 |
| **418** | **Coronavirus infection** | 6 | 27.39 | 186747009 |
| **419** | **Disease due to Coronaviridae** | 5 | 27.35 | 27619001 |
| **420** | **Viral disease** | 4 | 27.33 | 34014006 |
| **421** | **Lower abdominal pain** | 6 | 26.77 | 54586004 |
| **422** | **Disorder of aorta** | 5 | 26.54 | 47040006 |
| **423** | **Finding of aorta** | 5 | 26.22 | 473451005 |
| **424** | **Disease due to Enterovirus** | 6 | 26.15 | 53648006 |
| **425** | **Disease due to Picornaviridae** | 5 | 25.94 | 105633007 |
| **426** | **At risk of disease** | 4 | 25.85 | 313424005 |
| **427** | **Disease of non-coronary systemic artery** | 7 | 25.63 | 473449006 |
| **428** | **Disorder of endocrine system** | 5 | 25.46 | 362969004 |
| **429** | **Viral lower respiratory infection** | 6 | 25.30 | 312134000 |
| **430** | **Pneumonia caused by SARS-CoV-2** | 8 | 23.90 | 882784691000119100 |
| **431** | **Finding of functional performance and activity** | 4 | 23.69 | 248536006 |
| **432** | **Nutritional deficiency associated condition** | 3 | 23.52 | 363246002 |
| **433** | **Nutritional anemia** | 5 | 23.32 | 66612000 |
| **434** | **Iron deficiency anemia** | 4 | 23.05 | 87522002 |
| **435** | **Arthritis** | 6 | 22.99 | 3723001 |
| **436** | **Inflamed joint** | 5 | 22.88 | 298160000 |
| **437** | **Swelling of body region** | 4 | 21.73 | 300888008 |
| **438** | **Disorder of upper digestive tract** | 6 | 21.23 | 50410009 |
| **439** | **Lower respiratory infection caused by SARS-CoV-2** | 7 | 21.22 | 880529761000119102 |
| **440** | **Swelling of body structure** | 3 | 21.10 | 300872008 |

| | | | | |
|---|---|---|---|---|
| **441** | **Finding related to ability to perform gross motor function** | 5 | 20.78 | 364832000 |
| **442** | **Disorder of skin** | 5 | 20.69 | 95320005 |
| **443** | **Hyperglycemia** | 6 | 20.68 | 80394007 |
| **444** | **Arthralgia of the pelvic region and thigh** | 6 | 20.43 | 267952008 |
| **445** | **Lower respiratory tract infection** | 5 | 20.31 | 50417007 |
| **446** | **Anxiety disorder** | 4 | 20.14 | 197480006 |
| **447** | **Finding related to sleep** | 4 | 20.08 | 106168000 |
| **448** | **Asthma** | 5 | 19.77 | 195967001 |
| **449** | **Abnormality of atrial septum** | 7 | 19.53 | 253363004 |
| **450** | **Impaired mobility** | 8 | 19.35 | 82971005 |
| **451** | **Uncomplicated asthma** | 6 | 19.32 | 707444001 |
| **452** | **Undernutrition** | 5 | 19.28 | 65404009 |
| **453** | **Disorder of adrenal gland** | 5 | 19.10 | 30171000 |
| **454** | **Dyssomnia** | 4 | 19.00 | 44186003 |
| **455** | **Nutritional deficiency disorder** | 4 | 18.94 | 70241007 |
| **456** | **Finding related to ability to mobilize** | 7 | 18.63 | 365092005 |
| **457** | **Finding of back** | 4 | 18.52 | 414252009 |
| **458** | **Bacterial respiratory infection** | 6 | 18.52 | 312117008 |
| **459** | **Hereditary disease** | 4 | 18.47 | 32895009 |
| **460** | **Sleep disorder** | 3 | 18.39 | 39898005 |
| **461** | **Cardiac septal defects** | 7 | 18.16 | 253273004 |
| **462** | **Inflammatory disorder of lower respiratory tract** | 5 | 17.95 | 128997002 |
| **463** | **Disorder of skin and/or subcutaneous tissue** | 4 | 17.93 | 80659006 |
| **464** | **At risk - finding** | 3 | 17.93 | 281694009 |
| **465** | **Chronic disease of respiratory system** | 4 | 17.55 | 17097001 |
| **466** | **Genetic disease** | 3 | 17.44 | 782964007 |
| **467** | **COVID-19** | 7 | 16.90 | 840539006 |
| **468** | **Disorder of brain** | 5 | 16.81 | 81308009 |
| **469** | **Disorder of the central nervous system** | 4 | 16.70 | 23853001 |
| **470** | **Congenital cardiovascular disorder** | 4 | 16.66 | 762228008 |
| **471** | **Disorder of integument** | 4 | 16.65 | 128598002 |
| **472** | **Neutrophil count abnormal** | 7 | 16.63 | 165519006 |
| **473** | **Disorder of glucose metabolism** | 5 | 16.54 | 126877002 |
| **474** | **Finding of sensation of upper limb** | 4 | 16.45 | 298750003 |
| **475** | **Inflammatory disorder of genitourinary system** | 6 | 16.31 | 373406006 |
| **476** | **Congenital anomaly of cardiovascular structure of trunk** | 6 | 16.13 | 363028003 |
| **477** | **Congenital anomaly of cardiovascular system** | 5 | 15.61 | 9904008 |
| **478** | **Low back pain** | 6 | 15.53 | 279039007 |
| **479** | **Mental disorder** | 3 | 15.39 | 74732009 |
| **480** | **Asthenia** | 6 | 15.11 | 13791008 |
| **481** | **Primary immune deficiency disorder** | 5 | 15.06 | 58606001 |
| **482** | **Infectious disease of abdomen** | 5 | 15.05 | 128070006 |
| **483** | **Disorder of carbohydrate metabolism** | 4 | 14.37 | 20957000 |
| **484** | **Drug-related disorder** | 3 | 14.30 | 87858002 |
| **485** | **Finding of lower limb** | 5 | 14.13 | 116312005 |

| | | | | |
|---|---|---|---|---|
| **486** | **Gastritis** | 6 | 13.95 | 4556007 |
| **487** | **Finding of brain** | 4 | 13.90 | 299718000 |
| **488** | **Pain in upper limb** | 5 | 13.80 | 102556003 |
| **489** | **Inflammatory disorder of digestive system** | 5 | 13.75 | 373407002 |
| **490** | **Disorder of muscle** | 4 | 13.08 | 129565002 |
| **491** | **Central nervous system finding** | 3 | 12.98 | 246556002 |
| **492** | **Immunodeficiency disorder** | 4 | 12.44 | 234532001 |
| **493** | **Deficiency of micronutrients** | 6 | 12.04 | 238111008 |
| **494** | **Respiratory rate AND/OR rhythm finding** | 5 | 11.83 | 106049001 |
| **495** | **Vitamin deficiency** | 5 | 11.42 | 85670002 |
| **496** | **Congenital anomaly of thorax** | 7 | 11.31 | 363035006 |
| **497** | **Disorder of hematopoietic morphology** | 3 | 11.31 | 414026006 |
| **498** | **Congenital anomaly of upper trunk** | 6 | 11.29 | 363037003 |
| **499** | **Vitamin D deficiency** | 6 | 11.23 | 34713006 |
| **500** | **Disorder of vitamin D** | 5 | 11.23 | 386068004 |
| **501** | **Vitamin disease** | 4 | 11.21 | 7521001 |
| **502** | **Congenital heart disease** | 7 | 10.14 | 13213009 |
| **503** | **Finding of limb structure** | 4 | 10.12 | 302293008 |
| **504** | **Hereditary disorder by system** | 5 | 9.91 | 363137000 |
| **505** | **Gastrointestinal infection** | 6 | 9.89 | 715852004 |
| **506** | **Arthropathy** | 5 | 9.73 | 399269003 |
| **507** | **Acute pain** | 6 | 9.49 | 274663001 |
| **508** | **Health-related behavior finding** | 5 | 9.34 | 365949003 |
| **509** | **Neck pain** | 5 | 9.20 | 81680005 |
| **510** | **Finding of joint movement** | 3 | 9.19 | 298179002 |
| **511** | **Nutritional disorder** | 3 | 9.13 | 2492009 |
| **512** | **Red blood cell disorder** | 4 | 8.70 | 38292009 |
| **513** | **Deficiency of macronutrients** | 6 | 8.63 | 238107002 |
| **514** | **Disorder of body wall** | 4 | 8.62 | 399986003 |
| **515** | **Intestinal infectious disease** | 6 | 8.22 | 266071000 |
| **516** | **Dependence on enabling machine or device** | 5 | 8.07 | 105501005 |
| **517** | **Abnormal breathing** | 5 | 7.89 | 386813002 |
| **518** | **Infectious enteritis** | 6 | 7.66 | 55184003 |
| **519** | **Complication** | 3 | 7.48 | 116223007 |
| **520** | **Childhood obesity** | 5 | 7.04 | 444862003 |

**Table 1b: TreeScan-selected lab features**

| Cut | Concept | Tree Level | Log Likelihood Ratio | LOINC code |
|---|---|---|---|---|
| **1** | **Chemistry - non-challenge** | 3 | 38,720.65 | LP7786-9 |
| **2** | **Chemistry and Chemistry - challenge** | 2 | 38,497.68 | LP343631-0 |
| **3** | **Urinalysis** | 2 | 32,431.89 | LP7851-1 |
| **4** | **Chemistry - challenge** | 3 | 30,558.90 | LP7784-4 |
| **5** | **Hematology and Cell counts** | 2 | 28,384.49 | LP7803-2 |

| | | | | |
|---|---|---|---|---|
| **6** | **Chemistry Panels** | 4 | 27,479.15 | LP7834-7 |
| **7** | **Chemistry order set** | 2 | 27,098.39 | PANEL.CHEM |
| **8** | **Hematology & blood count order set** | 2 | 19,283.53 | PANEL.HEM/BC |
| **9** | **Hematology and Cell Count Panels** | 3 | 19,273.88 | LP7833-9 |
| **10** | **Female fertility** | 3 | 17,674.67 | LP32818-4 |
| **11** | **Fertility testing** | 2 | 17,593.83 | LP7798-4 |
| **12** | **Ophthalmology and Optometry** | 2 | 16,433.59 | LP7797-6 |
| **13** | **NEI eyeGENE slit lamp biomicroscopy** | 3 | 16,433.59 | LP203653-3 |
| **14** | **Cell types** | 4 | 16,267.06 | LP32763-2 |
| **15** | **Male fertility** | 3 | 16,267.06 | LP32817-6 |
| **16** | **CBC W Reflex Manual Differential panel \| Blood \| Hematology and Cell Count Panels** | 4 | 16,034.57 | LP393882-8 |
| **17** | **CBC W Reflex Manual Differential panel - Blood** | 3 | 16,034.57 | 57022-6 |
| **18** | **Cell markers** | 2 | 14,338.54 | LP7783-6 |
| **19** | **Analytes** | 3 | 13,710.14 | LP40317-7 |
| **20** | **Blood bank** | 2 | 13,543.69 | LP7776-0 |
| **21** | **CBC W Ordered Manual Differential panel - Blood** | 3 | 13,493.44 | 57782-5 |
| **22** | **Other markers** | 3 | 13,385.20 | LP32943-0 |
| **23** | **Comprehensive metabolic 2000 panel - Serum or Plasma** | 3 | 12,998.33 | 24323-8 |
| **24** | **Comprehensive metabolic 2000 panel \| Serum or Plasma \| Chemistry Panels** | 5 | 12,998.33 | LP386863-7 |
| **25** | **Hemogram without Platelets and with Manual Differential panel - Blood** | 3 | 10,942.93 | 24359-2 |
| **26** | **Hemogram without Platelets and with Manual Differential panel \| Blood \| Hematology and Cell Count Panels** | 4 | 10,942.93 | LP393926-3 |
| **27** | **Metabolic panel.large animal \| Serum or Plasma \| Chemistry Panels** | 5 | 10,362.23 | LP386983-3 |
| **28** | **Metabolic panel.large animal - Serum or Plasma** | 3 | 10,362.23 | 54232-4 |
| **29** | **Comprehensive metabolic 1998 panel - Serum or Plasma** | 3 | 10,333.18 | 24322-0 |
| **30** | **Comprehensive metabolic 1998 panel \| Serum or Plasma \| Chemistry Panels** | 5 | 10,333.18 | LP386862-9 |
| **31** | **Metabolic panel.small animal \| Serum or Plasma \| Chemistry Panels** | 5 | 10,269.06 | LP386984-1 |
| **32** | **Metabolic panel.small animal - Serum or Plasma** | 3 | 10,269.06 | 54233-2 |
| **33** | **Leukogram panel - Blood** | 3 | 9,060.36 | 48808-0 |
| **34** | **Leukogram panel \| Blood \| Hematology and Cell Count Panels** | 4 | 9,060.36 | LP393932-1 |
| **35** | **Manual Differential panel - Blood** | 3 | 9,060.36 | 24318-8 |
| **36** | **Manual Differential panel \| Blood \| Hematology and Cell Count Panels** | 4 | 9,060.36 | LP393933-9 |
| **37** | **CBC W Differential panel, method unspecified \| Blood \| Hematology and Cell Count Panels** | 4 | 9,032.67 | LP393880-2 |
| **38** | **CBC W Differential panel, method unspecified - Blood** | 3 | 9,032.67 | 69742-5 |

| | | | | |
|---|---|---|---|---|
| **39** | **Microbiology and Antimicrobial susceptibility** | 2 | 8,971.90 | LP343406-7 |
| **40** | **Microbiology** | 3 | 8,954.28 | LP7819-8 |
| **41** | **Renal function 2000 panel - Serum or Plasma** | 3 | 8,215.07 | 24362-6 |
| **42** | **Renal function 2000 panel \| Serum or Plasma \| Chemistry Panels** | 5 | 8,215.07 | LP387047-6 |
| **43** | **PhenX Panel** | 2 | 8,075.58 | PANEL.PHENX |
| **44** | **Basic metabolic and albumin panel - Serum or Plasma** | 3 | 8,067.29 | 89044-2 |
| **45** | **Basic metabolic and albumin panel \| Serum or Plasma \| Chemistry Panels** | 5 | 8,067.29 | LP386816-5 |
| **46** | **PhenX - complete blood count protocol 220501** | 3 | 7,807.08 | 93340-8 |
| **47** | **Obstetrics order set** | 2 | 7,759.90 | PANEL.OBS |
| **48** | **Obstetric 1996 panel - Serum and Blood** | 3 | 7,759.90 | 24364-2 |
| **49** | **CBC W Auto Differential panel - Blood** | 3 | 7,731.22 | 57021-8 |
| **50** | **CBC W Auto Differential panel \| Blood \| Hematology and Cell Count Panels** | 4 | 7,731.22 | LP393878-6 |
| **51** | **Urinalysis order set** | 2 | 7,363.21 | PANEL.UA |
| **52** | **Urinalysis Panels** | 3 | 7,363.21 | LP7838-8 |
| **53** | **Basic metabolic 2000 panel - Serum or Plasma** | 3 | 7,200.82 | 24321-2 |
| **54** | **Basic metabolic 2000 panel \| Serum or Plasma \| Chemistry Panels** | 5 | 7,200.82 | LP386818-1 |
| **55** | **Metabolic panel.dialysis patient \| Serum or Plasma \| Chemistry Panels** | 5 | 6,999.02 | LP386982-5 |
| **56** | **Metabolic panel.dialysis patient - Serum or Plasma** | 3 | 6,999.02 | 88843-8 |
| **57** | **Coagulation** | 2 | 6,637.96 | LP7788-5 |
| **58** | **Short blood count panel - Blood** | 3 | 6,545.52 | 55429-5 |
| **59** | **Short blood count panel \| Blood \| Hematology and Cell Count Panels** | 4 | 6,545.52 | LP393938-8 |
| **60** | **Differential panel, method unspecified - Blood** | 3 | 6,515.25 | 69738-3 |
| **61** | **Differential panel, method unspecified \| Blood \| Hematology and Cell Count Panels** | 4 | 6,515.25 | LP393917-2 |
| **62** | **Routine** | 3 | 6,460.25 | LP31624-7 |
| **63** | **Urinalysis complete W Reflex Culture panel - Urine** | 3 | 6,373.43 | 58077-9 |
| **64** | **Urinalysis complete W Reflex Culture panel \| Urine \| Urinalysis Panels** | 4 | 6,373.43 | LP402547-6 |
| **65** | **Basic metabolic 2008 panel with ionized calcium - Serum or Plasma** | 3 | 6,349.29 | 70219-1 |
| **66** | **Basic metabolic 2008 panel with ionized calcium \| Serum or Plasma \| Chemistry Panels** | 5 | 6,349.29 | LP386819-9 |
| **67** | **Basic metabolic 1998 panel \| Serum or Plasma \| Chemistry Panels** | 5 | 6,290.32 | LP386817-3 |
| **68** | **Basic metabolic 1998 panel - Serum or Plasma** | 3 | 6,290.32 | 24320-4 |
| **69** | **Urinalysis complete panel \| Urine \| Urinalysis Panels** | 4 | 6,226.33 | LP402546-8 |
| **70** | **Urinalysis dipstick W Reflex Microscopic panel \| Urine \| Urinalysis Panels** | 4 | 6,226.33 | LP402550-0 |
| **71** | **Urinalysis dipstick W Reflex Microscopic panel - Urine** | 3 | 6,226.33 | 57020-0 |
| **72** | **Urinalysis complete panel - Urine** | 3 | 6,226.33 | 24356-8 |

| | | | | |
|---|---|---|---|---|
| **73** | **Coagulation Panels** | 3 | 5,958.81 | LP31893-8 |
| **74** | **Coagulation order set** | 2 | 5,958.81 | PANEL.COAG |
| **75** | **Intravascular coagulation and fibrinolysis panel \| Patient \| Coagulation Panels** | 4 | 5,894.75 | LP428844-7 |
| **76** | **Intravascular coagulation and fibrinolysis panel** | 3 | 5,894.75 | 98125-8 |
| **77** | **Hepatic function 2000 panel - Serum or Plasma** | 3 | 5,517.73 | 24325-3 |
| **78** | **Hepatic function 2000 panel \| Serum or Plasma \| Chemistry Panels** | 5 | 5,517.73 | LP386948-6 |
| **79** | **Microbiology Panels** | 4 | 5,285.46 | LP7835-4 |
| **80** | **Microbiology order set** | 2 | 5,284.27 | PANEL.MICRO |
| **81** | **Gases and acid/Base** | 4 | 5,128.12 | LP31400-2 |
| **82** | **Challenge Bank Panels** | 4 | 5,053.25 | LP31895-3 |
| **83** | **Challenge order set** | 2 | 5,053.25 | PANEL.CHAL |
| **84** | **CBC panel - Blood by Automated count** | 3 | 5,033.15 | 58410-2 |
| **85** | **CBC panel \| Blood \| Hematology and Cell Count Panels** | 4 | 5,033.15 | LP393905-7 |
| **86** | **Cytokines panel - Serum or Plasma** | 3 | 4,981.18 | 82335-1 |
| **87** | **Cytokines panel \| Serum or Plasma \| Chemistry Panels** | 5 | 4,981.18 | LP386881-9 |
| **88** | **IEEE 11073 Rosetta laboratory panel Blood** | 2 | 4,745.09 | 75967-0 |
| **89** | **PT and aPTT and Fibrinogen panel - Platelet poor plasma by Coagulation assay** | 3 | 4,659.20 | 49045-8 |
| **90** | **PT and aPTT and Fibrinogen panel \| Platelet poor plasma \| Coagulation Panels** | 4 | 4,659.20 | LP394456-0 |
| **91** | **Hepatic function 1996 panel - Serum or Plasma** | 3 | 4,606.83 | 24324-6 |
| **92** | **Hepatic function 1996 panel \| Serum or Plasma \| Chemistry Panels** | 5 | 4,606.83 | LP386947-8 |
| **93** | **Urinalysis dipstick W Reflex Culture panel \| Urine \| Urinalysis Panels** | 4 | 4,582.46 | LP402549-2 |
| **94** | **Urinalysis dipstick W Reflex Culture panel - Urine** | 3 | 4,582.46 | 57019-2 |
| **95** | **Vasopressin challenge post water deprivation panel \| Urine and Serum or Plasma \| Challenge Bank Panels** | 5 | 4,544.62 | LP417168-4 |
| **96** | **Water deprivation challenge panel \| Urine and Serum or Plasma \| Challenge Bank Panels** | 5 | 4,544.62 | LP417169-2 |
| **97** | **Vasopressin challenge post water deprivation panel - Urine and Serum or Plasma** | 3 | 4,544.62 | 94128-6 |
| **98** | **Water deprivation challenge panel - Urine and Serum or Plasma** | 3 | 4,544.62 | 94129-4 |
| **99** | **Urinalysis macro (dipstick) panel - Urine** | 3 | 4,432.78 | 24357-6 |
| **100** | **Urinalysis macro (dipstick) panel \| Urine \| Urinalysis Panels** | 4 | 4,432.78 | LP402551-8 |
| **101** | **Interleukins** | 4 | 4,081.27 | LP30867-3 |
| **102** | **Electrolytes 1998 and Venous pH panel \| Serum or Plasma + Blood venous \| Chemistry Panels** | 5 | 3,934.67 | LP386892-6 |
| **103** | **Electrolytes 1998 and Venous pH panel - Serum or Plasma + Blood venous** | 3 | 3,934.67 | 34554-6 |
| **104** | **Respiratory measures and Ventilator management** | 2 | 3,788.05 | LP7840-4 |

| | | | | |
|---|---|---|---|---|
| **105** | **Electrolytes 1998 panel \| Serum or Plasma \| Chemistry Panels** | 5 | 3,653.46 | LP386893-4 |
| **106** | **Electrolytes 1998 panel - Serum or Plasma** | 3 | 3,653.46 | 24326-1 |
| **107** | **Liver fibrosis score panel \| Serum or Plasma \| Chemistry Panels** | 5 | 3,392.36 | LP386977-5 |
| **108** | **Blood Indices** | 3 | 3,291.82 | LP30866-5 |
| **109** | **PT and aPTT panel - Platelet poor plasma by Coagulation assay** | 3 | 3,258.15 | 34529-8 |
| **110** | **PT and aPTT panel \| Platelet poor plasma \| Coagulation Panels** | 4 | 3,258.15 | LP394457-8 |
| **111** | **Red cell indices** | 4 | 3,174.73 | LP31669-2 |
| **112** | **Smear morphology panel \| Blood \| Hematology and Cell Count Panels** | 4 | 2,706.31 | LP393939-6 |
| **113** | **Smear morphology panel - Blood** | 3 | 2,706.31 | 34994-4 |
| **114** | **Auto Differential panel \| Blood \| Hematology and Cell Count Panels** | 4 | 2,701.53 | LP393877-8 |
| **115** | **Auto Differential panel - Blood** | 3 | 2,701.53 | 57023-4 |
| **116** | **Liver fibrosis score panel - Serum or Plasma Calculated by FibroMeter** | 3 | 2,527.37 | 78699-6 |
| **117** | **PT panel - Platelet poor plasma by Coagulation assay** | 3 | 2,464.44 | 34528-0 |
| **118** | **PT panel \| Platelet poor plasma \| Coagulation Panels** | 4 | 2,464.44 | LP394458-6 |
| **119** | **Warfarin tracking panel - Platelet poor plasma** | 3 | 2,464.44 | 55401-4 |
| **120** | **Warfarin tracking panel \| Platelet poor plasma \| Coagulation Panels** | 4 | 2,464.44 | LP394464-4 |
| **121** | **Cardiology** | 2 | 2,442.26 | LP29708-2 |
| **122** | **Hemogram without Platelets panel - Blood** | 3 | 2,429.01 | 24358-4 |
| **123** | **Hemogram without Platelets panel \| Blood \| Hematology and Cell Count Panels** | 4 | 2,429.01 | LP393927-1 |
| **124** | **Serology - non-micro** | 2 | 2,363.43 | LP7844-6 |
| **125** | **Respiratory pathogens DNA and RNA panel - Nasopharynx by NAA with non-probe detection** | 3 | 2,350.28 | 82159-5 |
| **126** | **Sugars/Sugar metabolism** | 4 | 2,298.21 | LP31399-6 |
| **127** | **Clinical Risk Panel** | 2 | 2,292.57 | LP267622-1 |
| **128** | **Sequential Organ Failure Assessment \| Patient \| Clinical Risk Panel** | 3 | 2,249.71 | LP428913-0 |
| **129** | **Sequential Organ Failure Assessment SOFA** | 4 | 2,249.71 | 96789-3 |
| **130** | **Gas panel - Venous blood** | 3 | 2,192.50 | 24339-4 |
| **131** | **Gas panel \| Blood venous \| Chemistry Panels** | 5 | 2,192.50 | LP386940-3 |
| **132** | **Gas and Carbon monoxide panel - Venous blood** | 3 | 2,153.10 | 24344-4 |
| **133** | **Gas and Carbon monoxide panel \| Blood venous \| Chemistry Panels** | 5 | 2,153.10 | LP386932-0 |
| **134** | **Urinalysis microscopic panel \| Urine sediment \| Urinalysis Panels** | 4 | 2,129.37 | LP402554-2 |
| **135** | **Drug doses** | 2 | 2,065.10 | LP7791-9 |
| **136** | **Mineral, bone, joint, connective tissue** | 4 | 2,061.07 | LP31413-5 |

| | | | | |
|---|---|---|---|---|
| **137** | **Nonalcoholic steatohepatitis and fibrosis panel \| Serum or Plasma \| Chemistry Panels** | 5 | 2,013.86 | LP417161-9 |
| **138** | **Nonalcoholic steatohepatitis and fibrosis panel - Serum or Plasma** | 3 | 2,013.86 | 93691-4 |
| **139** | **Menorrhagia coagulation panel - Platelet poor plasma** | 3 | 1,995.73 | 44791-2 |
| **140** | **Menorrhagia coagulation panel \| Platelet poor plasma \| Coagulation Panels** | 4 | 1,995.73 | LP394448-7 |
| **141** | **Erythrocyte morphology panel \| Blood \| Hematology and Cell Count Panels** | 4 | 1,926.69 | LP393918-0 |
| **142** | **Erythrocyte morphology panel - Blood** | 3 | 1,926.69 | 58408-6 |
| **143** | **Electrocardiogram measures** | 2 | 1,909.36 | EKG.MEAS |
| **144** | **EKG measurements** | 3 | 1,909.36 | LP7795-0 |
| **145** | **Respiratory pathogens DNA and RNA panel \| Nasopharynx \| Microbiology Panels** | 5 | 1,903.45 | LP380083-8 |
| **146** | **Liver fibrosis score panel \| Patient \| Chemistry Panels** | 5 | 1,901.28 | LP428840-5 |
| **147** | **Liver fibrosis score panel by Calculated by FIB4** | 3 | 1,901.28 | 98491-4 |
| **148** | **Urinalysis microscopic panel - Urine sediment** | 3 | 1,884.46 | 24365-9 |
| **149** | **Protein fractions 3 panel \| Serum or Plasma \| Chemistry Panels** | 5 | 1,861.12 | LP387030-2 |
| **150** | **Protein fractions 3 panel - Serum or Plasma** | 3 | 1,861.12 | 48811-4 |
| **151** | **Rheumatoid arthritis disease activity panel - Serum or Plasma by VectraDA** | 3 | 1,784.58 | 75635-3 |
| **152** | **Rheumatoid arthritis disease activity panel \| Serum or Plasma \| Chemistry Panels** | 5 | 1,784.58 | LP387049-2 |
| **153** | **Human coronavirus** | 2 | 1,779.97 | LG32771-4 |
| **154** | **Respiratory pathogens DNA and RNA 12b panel \| XXX \| Microbiology Panels** | 5 | 1,775.08 | LP380080-4 |
| **155** | **Respiratory pathogens DNA and RNA 12b panel - Specimen by NAA with probe detection** | 3 | 1,775.08 | 60566-7 |
| **156** | **Gas panel - Blood** | 3 | 1,748.43 | 24338-6 |
| **157** | **Gas panel \| Blood \| Chemistry Panels** | 5 | 1,748.43 | LP386934-6 |
| **158** | **Chemistry - routine challenge** | 3 | 1,712.87 | LP234174-3 |
| **159** | **Gas and Carbon monoxide panel \| Blood \| Chemistry Panels** | 5 | 1,704.99 | LP386929-6 |
| **160** | **Gas and Carbon monoxide panel - Blood** | 3 | 1,704.99 | 24343-6 |
| **161** | **Respiratory pathogens DNA and RNA 12a panel - Specimen by NAA with probe detection** | 3 | 1,691.74 | 50219-5 |
| **162** | **Respiratory pathogens DNA and RNA 12a panel \| XXX \| Microbiology Panels** | 5 | 1,691.74 | LP380079-6 |
| **163** | **Protein electrophoresis and Immunoglobulins panel - Serum** | 3 | 1,616.81 | 55295-0 |
| **164** | **Protein electrophoresis and Immunoglobulins panel \| Serum \| Chemistry Panels** | 5 | 1,616.81 | LP387029-4 |
| **165** | **Troponin I.cardiac \| Serum or Plasma \| Chemistry - non-challenge** | 4 | 1,610.19 | LP385942-0 |
| **166** | **Troponin I.cardiac\|MCnc\|Pt\|ANYBldSerPl** | 2 | 1,606.00 | LG433-9 |
| **167** | **Troponin I.cardiac [Mass/volume] in Serum or Plasma** | 2 | 1,606.00 | 10839-9 |

| | | | | |
|---|---|---|---|---|
| **168** | **Sodium and Potassium panel [Moles/volume] - Serum or Plasma** | 3 | 1,605.85 | 34548-8 |
| **169** | **Sodium and Potassium panel \| Serum or Plasma \| Chemistry Panels** | 5 | 1,605.85 | LP387058-3 |
| **170** | **Cell marker order sets** | 2 | 1,565.12 | PANEL.CELLMARK |
| **171** | **Cellmarker Panels** | 3 | 1,565.12 | LP36900-6 |
| **172** | **Protein electrophoresis panel \| Serum or Plasma \| Chemistry Panels** | 5 | 1,505.57 | LP387033-6 |
| **173** | **Protein electrophoresis panel - Serum or Plasma** | 3 | 1,505.57 | 24351-9 |
| **174** | **Immunoelectrophoresis panel \| Serum \| Chemistry Panels** | 5 | 1,505.57 | LP386957-7 |
| **175** | **Immunoelectrophoresis panel - Serum** | 3 | 1,505.57 | 29586-5 |
| **176** | **Erythrocyte sedimentation rate \| Blood \| Hematology and Cell counts** | 3 | 1,447.29 | LP392450-5 |
| **177** | **Fibrinogen \| Platelet poor plasma \| Coagulation** | 4 | 1,442.97 | LP394019-6 |
| **178** | **Fibrinogen [Mass/volume] in Platelet poor plasma by Coagulation assay** | 2 | 1,442.97 | 3255-7 |
| **179** | **Iron panel \| Serum or Plasma \| Chemistry Panels** | 5 | 1,404.08 | LP386968-4 |
| **180** | **Ferritin\|MCnc\|Pt\|ANYBldSerPl** | 2 | 1,404.08 | LG6299-4 |
| **181** | **Ferritin \| Serum or Plasma \| Chemistry - non-challenge** | 4 | 1,404.08 | LP385083-3 |
| **182** | **Ferritin [Mass/volume] in Serum or Plasma** | 2 | 1,404.08 | 2276-4 |
| **183** | **Iron panel - Serum or Plasma** | 3 | 1,404.08 | 75689-0 |
| **184** | **Ferritin\|Pt\|Ser/Plas** | 2 | 1,404.08 | LG43688-7 |
| **185** | **Erythrogram panel - Blood** | 3 | 1,395.21 | 48809-8 |
| **186** | **Erythrogram panel \| Blood \| Hematology and Cell Count Panels** | 4 | 1,395.21 | LP393919-8 |
| **187** | **C reactive protein \| Serum or Plasma \| Chemistry - non-challenge** | 4 | 1,365.96 | LP384546-0 |
| **188** | **C reactive protein\|MCnc\|Pt\|ANYBldSerPl** | 2 | 1,354.03 | LG6212-7 |
| **189** | **C reactive protein [Mass/volume] in Serum or Plasma** | 2 | 1,354.03 | 1988-5 |
| **190** | **C reactive protein\|Pt\|Ser/Plas** | 2 | 1,354.03 | LG48228-7 |
| **191** | **Hepatitis C virus** | 2 | 1,303.00 | LG32754-0 |
| **192** | **Human coronavirus RNA panel \| XXX \| Microbiology Panels** | 5 | 1,290.38 | LP380015-0 |
| **193** | **Human coronavirus RNA panel - Specimen by NAA with probe detection** | 3 | 1,290.38 | 63430-3 |
| **194** | **Hepatitis C virus FibroSURE panel - Serum or Plasma** | 3 | 1,289.03 | 48796-7 |
| **195** | **Hepatitis C virus FibroSURE panel \| Serum or Plasma \| Chemistry Panels** | 5 | 1,289.03 | LP386949-4 |
| **196** | **Cardiac studies order set** | 2 | 1,277.14 | PANEL.CARDIAC |
| **197** | **Cardiac Panels** | 3 | 1,277.14 | LP31905-0 |
| **198** | **Lupus anticoagulant three screening tests W Reflex panel \| Platelet poor plasma \| Coagulation Panels** | 4 | 1,271.02 | LP394446-1 |
| **199** | **Lupus anticoagulant aPTT, dRVVT and PT screening panel W Reflex** | 3 | 1,271.02 | 75881-3 |

| | | | | |
|---|---|---|---|---|
| **200** | **Erythrocyte sedimentation rate** | 2 | 1,270.36 | 30341-2 |
| **201** | **PT mixing study panel \| Platelet poor plasma \| Coagulation Panels** | 4 | 1,239.92 | LP411451-0 |
| **202** | **PT mixing study panel - Platelet poor plasma by Coagulation assay** | 3 | 1,239.92 | 93321-8 |
| **203** | **Urinalysis panel \| Urine \| Urinalysis Panels** | 4 | 1,235.12 | LP402556-7 |
| **204** | **Urinalysis panel - Urine by Auto** | 3 | 1,235.12 | 50564-4 |
| **205** | **Prothrombin time (PT) \| Platelet poor plasma \| Coagulation** | 4 | 1,232.50 | LP394044-4 |
| **206** | **INR in Platelet poor plasma by Coagulation assay** | 2 | 1,231.65 | 6301-6 |
| **207** | **INR \| Platelet poor plasma \| Coagulation** | 4 | 1,231.65 | LP394061-8 |
| **208** | **Prothrombin time (PT)** | 2 | 1,231.20 | 5902-2 |
| **209** | **Hemoglobin and Hematocrit panel \| Blood \| Hematology and Cell Count Panels** | 4 | 1,228.24 | LP393920-6 |
| **210** | **Hemoglobin and Hematocrit panel - Blood** | 3 | 1,228.24 | 24360-0 |
| **211** | **Short Fibrin D-dimer FEU and DDU panel - Platelet poor plasma** | 3 | 1,202.75 | 55398-2 |
| **212** | **Short Fibrin D-dimer FEU and DDU panel \| Platelet poor plasma \| Coagulation Panels** | 4 | 1,202.75 | LP394459-4 |
| **213** | **Fibrin D-dimer FEU \| Platelet poor plasma \| Coagulation** | 4 | 1,202.75 | LP394015-4 |
| **214** | **Fibrin D-dimer FEU [Mass/volume] in Platelet poor plasma** | 2 | 1,202.75 | 48065-7 |
| **215** | **Liver fibrosis score panel - Serum or Plasma Calculated by HepaScore** | 3 | 1,190.13 | 78698-8 |
| **216** | **PhenX domain - Respiratory** | 3 | 1,172.21 | 62611-9 |
| **217** | **T-cell subsets CD4 and CD8 panel - Blood** | 3 | 1,172.13 | 65759-3 |
| **218** | **T-cell subsets CD4 and CD8 panel \| Blood \| Cellmarker Panels** | 4 | 1,172.13 | LP400872-0 |
| **219** | **Antibiotic susceptibilities** | 3 | 1,136.27 | LP7755-4 |
| **220** | **Bilirubin direct and total panel [Mass/volume] - Serum or Plasma** | 3 | 1,134.44 | 34543-9 |
| **221** | **Bilirubin direct and total panel \| Serum or Plasma \| Chemistry Panels** | 5 | 1,134.44 | LP386833-0 |
| **222** | **Sodium\|SCnc\|Pt\|ANYBldSerPl** | 2 | 1,134.26 | LG11363-5 |
| **223** | **Potassium\|SCnc\|Pt\|ANYBldSerPl** | 2 | 1,128.27 | LG10990-6 |
| **224** | **Miscellaneous Hematology** | 3 | 1,070.32 | LP30865-7 |
| **225** | **Natriuretic peptide.B\|Pt\|Ser/Plas** | 2 | 1,062.42 | LG44906-2 |
| **226** | **Natriuretic peptide B \| Serum or Plasma \| Chemistry - non-challenge** | 4 | 1,062.42 | LP385935-4 |
| **227** | **Natriuretic peptide B [Mass/volume] in Serum or Plasma** | 2 | 1,062.42 | 30934-4 |
| **228** | **Natriuretic peptide.B\|MCnc\|Pt\|ANYBldSerPl** | 2 | 1,062.42 | LG12080-4 |
| **229** | **Calcium-phosphorus product panel - Serum or Plasma** | 3 | 1,050.89 | 50676-6 |
| **230** | **Calcium-phosphorus product panel \| Serum or Plasma \| Chemistry Panels** | 5 | 1,050.89 | LP386841-3 |
| **231** | **Glucose\|MCnc\|Pt\|ANYBldSerPl** | 2 | 1,037.13 | LG7967-5 |

| | | | | |
|---|---|---|---|---|
| **232** | **Basic metabolic panel - Blood** | 3 | 1,037.07 | 51990-0 |
| **233** | **Basic metabolic panel \| Blood \| Chemistry Panels** | 5 | 1,037.07 | LP386820-7 |
| **234** | **Immunodeficiency follow-up panel \| XXX \| Cellmarker Panels** | 4 | 1,032.06 | LP400855-5 |
| **235** | **Immunodeficiency follow-up panel - Specimen by Flow cytometry (FC)** | 3 | 1,032.06 | 49120-9 |
| **236** | **12 lead EKG panel** | 2 | 1,031.59 | 34534-8 |
| **237** | **12 lead EKG panel \| Heart \| Cardiac Panels** | 4 | 1,031.59 | LP408301-2 |
| **238** | **Amylase and Creatinine clearance panel \| Urine and Serum or Plasma \| Chemistry Panels** | 5 | 996.71 | LP386810-8 |
| **239** | **Amylase and Creatinine clearance panel - Urine and Serum or Plasma** | 3 | 996.71 | 44789-6 |
| **240** | **Creatinine renal clearance adjusted for body surface area panel \| Urine and Serum or Plasma \| Chemistry Panels** | 5 | 984.34 | LP386873-6 |
| **241** | **Creatinine 24H renal clearance adjusted for body surface area panel** | 3 | 984.34 | 58446-6 |
| **242** | **Genetic Antimicrobial Resistance** | 4 | 968.15 | LP64213-9 |
| **243** | **Platelets panel \| Blood \| Hematology and Cell Count Panels** | 4 | 961.35 | LP393936-2 |
| **244** | **Platelets panel - Blood by Automated count** | 3 | 961.35 | 53800-9 |
| **245** | **Lymphocytes \| Blood \| Hematology and Cell counts** | 3 | 961.23 | LP392919-9 |
| **246** | **Lymphocytes\|NCnc\|Pt\|Bld** | 2 | 961.23 | LG32863-9 |
| **247** | **Creatinine renal clearance panel \| Urine and Serum or Plasma \| Chemistry Panels** | 5 | 949.80 | LP386874-4 |
| **248** | **Creatinine 24H renal clearance panel** | 3 | 949.80 | 34555-3 |
| **249** | **Alkaline phosphatase isoenz panel - Serum or Plasma** | 3 | 942.20 | 24332-9 |
| **250** | **Alkaline phosphatase isoenz panel \| Serum or Plasma \| Chemistry Panels** | 5 | 942.20 | LP386802-5 |
| **251** | **Albumin\|MCnc\|Pt\|ANYBldSerPl** | 2 | 941.19 | LG5465-2 |
| **252** | **Albumin \| Serum or Plasma \| Chemistry - non-challenge** | 3 | 941.19 | LP384485-1 |
| **253** | **Alkaline phosphatase \| Serum or Plasma \| Chemistry - non-challenge** | 4 | 940.53 | LP382722-9 |
| **254** | **Alkaline phosphatase\|CCnc\|Pt\|ANYBldSerPl** | 2 | 940.53 | LG5665-7 |
| **255** | **Alkaline phosphatase [Enzymatic activity/volume] in Serum or Plasma** | 2 | 940.53 | 6768-6 |
| **256** | **Cells panel \| Urine sediment \| Urinalysis Panels** | 4 | 931.04 | LP402542-7 |
| **257** | **Cells panel - Urine sediment** | 3 | 931.04 | 58434-2 |
| **258** | **Multiple sclerosis panel - Serum and CSF** | 3 | 922.95 | 55121-8 |
| **259** | **Multiple sclerosis panel \| Serum and CSF \| Chemistry Panels** | 5 | 922.95 | LP386996-5 |
| **260** | **Alanine aminotransferase\|CCnc\|Pt\|ANYBldSerPl** | 2 | 921.27 | LG5272-2 |
| **261** | **Alanine aminotransferase \| Serum or Plasma \| Chemistry - non-challenge** | 4 | 921.27 | LP382703-9 |
| **262** | **Chloride\|SCnc\|Pt\|ANYBldSerPl** | 2 | 919.68 | LG6373-7 |

| | | | | |
|---|---|---|---|---|
| **263** | **Protein electrophoresis and M protein isotype panel - Serum or Plasma** | 3 | 917.78 | 90991-1 |
| **264** | **Monoclonal gammopathy panel \| Serum or Plasma \| Chemistry Panels** | 5 | 917.78 | LP386994-0 |
| **265** | **Protein electrophoresis and M protein isotype panel \| Serum or Plasma \| Chemistry Panels** | 5 | 917.78 | LP387028-6 |
| **266** | **Monoclonal gammopathy panel - Serum or Plasma** | 3 | 917.78 | 90992-9 |
| **267** | **Protein \| Serum or Plasma \| Chemistry - non-challenge** | 4 | 910.90 | LP384468-7 |
| **268** | **Protein [Mass/volume] in Serum or Plasma** | 2 | 910.90 | 2885-2 |
| **269** | **Protein\|Pt\|Ser/Plas** | 2 | 910.90 | LG49422-5 |
| **270** | **Protein\|MCnc\|Pt\|ANYBldSerPl** | 2 | 910.90 | LG1777-4 |
| **271** | **Carbon dioxide\|SCnc\|Pt\|ANYBldSerPl** | 2 | 909.02 | LG4454-7 |
| **272** | **Monocytes\|NCnc\|Pt\|Bld** | 2 | 902.17 | LG32885-2 |
| **273** | **Monocytes \| Blood \| Hematology and Cell counts** | 3 | 902.17 | LP393028-8 |
| **274** | **Aspartate aminotransferase \| Serum or Plasma \| Chemistry - non-challenge** | 4 | 899.19 | LP382836-7 |
| **275** | **Aspartate aminotransferase\|CCnc\|Pt\|ANYBldSerPl** | 2 | 899.19 | LG6033-7 |
| **276** | **Creatinine and Glomerular filtration rate.predicted panel \| Serum, Plasma or Blood \| Chemistry Panels** | 5 | 893.99 | LP386871-0 |
| **277** | **Creatinine and Glomerular filtration rate.predicted panel - Serum, Plasma or Blood** | 3 | 893.99 | 45066-8 |
| **278** | **Urea nitrogen\|MCnc\|Pt\|ANYBldSerPl** | 2 | 876.85 | LG1314-6 |
| **279** | **Creatinine\|MCnc\|Pt\|ANYBldSerPl** | 2 | 874.25 | LG6657-3 |
| **280** | **Carbon dioxide \| Serum or Plasma \| Chemistry - non-challenge** | 3 | 874.01 | LP383334-2 |
| **281** | **Albumin [Mass/volume] in Serum or Plasma** | 2 | 873.98 | 1751-7 |
| **282** | **Albumin\|Pt\|Ser/Plas\|2754.105 g/mole** | 2 | 873.98 | LG49829-1 |
| **283** | **Parathyrin.intact and Calcium panel \| Serum or Plasma \| Chemistry Panels** | 5 | 873.89 | LP387010-4 |
| **284** | **Parathyrin.intact and Calcium panel - Serum or Plasma** | 3 | 873.89 | 24346-9 |
| **285** | **Calcium\|Pt\|Ser/Plas\|40.078 g/mole** | 2 | 867.66 | LG49864-8 |
| **286** | **Parathyrin.mid molecule and Calcium panel - Serum or Plasma** | 3 | 867.66 | 24347-7 |
| **287** | **Parathyrin.mid molecule and Calcium panel \| Serum or Plasma \| Chemistry Panels** | 5 | 867.66 | LP387012-0 |
| **288** | **Calcium [Mass/volume] in Serum or Plasma** | 2 | 867.66 | 17861-6 |
| **289** | **Calcium\|MCnc\|Pt\|ANYBldSerPl** | 2 | 867.66 | LG7247-2 |
| **290** | **Calcium \| Serum or Plasma \| Chemistry - non-challenge** | 5 | 867.66 | LP385966-9 |
| **291** | **Bilirubin \| Serum or Plasma \| Chemistry - non-challenge** | 4 | 867.23 | LP385283-9 |
| **292** | **Bilirubin\|MCnc\|Pt\|ANYBldSerPl** | 2 | 867.23 | LG6199-6 |
| **293** | **Bilirubin.total [Mass/volume] in Serum or Plasma** | 2 | 867.23 | 1975-2 |
| **294** | **Bilirubin\|Pt\|Ser/Plas\|584.673 g/mole** | 2 | 867.23 | LG49848-1 |

| | | | | |
|---|---|---|---|---|
| **295** | **PhenX - respiratory - arterial blood gas - ABG protocol 090201** | 3 | 854.40 | 62613-5 |
| **296** | **Erythrocytes\|Urine/Urine sed** | 2 | 853.99 | LG40868-8 |
| **297** | **Natriuretic peptide.B prohormone N-Terminal [Mass/volume] in Serum or Plasma** | 2 | 851.78 | 33762-6 |
| **298** | **Natriuretic peptide.B prohormone N-Terminal\|MCnc\|Pt\|ANYBldSerPl** | 2 | 851.78 | LG13322-9 |
| **299** | **Natriuretic peptide.B prohormone N-Terminal\|Pt\|Ser/Plas** | 2 | 851.78 | LG47358-3 |
| **300** | **Natriuretic peptide.B prohormone N-Terminal \| Serum or Plasma \| Chemistry - non-challenge** | 4 | 851.78 | LP385939-6 |
| **301** | **Serology order set** | 2 | 848.69 | PANEL.SERO |
| **302** | **Urea nitrogen [Mass/volume] in Serum or Plasma** | 2 | 847.01 | 3094-0 |
| **303** | **Urea nitrogen\|Pt\|Ser/Plas** | 2 | 847.01 | LG49763-2 |
| **304** | **Urea nitrogen \| Serum or Plasma \| Chemistry - non-challenge** | 4 | 846.81 | LP385464-5 |
| **305** | **Chloride \| Serum or Plasma \| Chemistry - non-challenge** | 4 | 846.80 | LP386588-0 |
| **306** | **Chloride [Moles/volume] in Serum or Plasma** | 2 | 846.80 | 2075-0 |
| **307** | **Potassium\|Pt\|Ser/Plas\|39.098 g/mole** | 2 | 846.70 | LG49936-4 |
| **308** | **Potassium \| Serum or Plasma \| Chemistry - non-challenge** | 4 | 846.70 | LP386618-5 |
| **309** | **Potassium [Moles/volume] in Serum or Plasma** | 2 | 846.70 | 2823-3 |
| **310** | **Carbon dioxide, total [Moles/volume] in Serum or Plasma** | 2 | 845.73 | 2028-9 |
| **311** | **Creatinine [Mass/volume] in Serum or Plasma** | 2 | 844.50 | 2160-0 |
| **312** | **Creatinine \| Serum or Plasma \| Chemistry - non-challenge** | 4 | 844.50 | LP385359-7 |
| **313** | **Creatinine\|Pt\|Ser/Plas\|113.12 g/mole** | 2 | 844.50 | LG50024-5 |
| **314** | **Gastrointestinal pathogens DNA and RNA panel - Stool by NAA with non-probe detection** | 3 | 843.31 | 82195-9 |
| **315** | **Gastrointestinal pathogens DNA and RNA panel \| Stool \| Microbiology Panels** | 5 | 843.31 | LP379970-9 |
| **316** | **Alanine aminotransferase [Enzymatic activity/volume] in Serum or Plasma** | 2 | 840.09 | 1742-6 |
| **317** | **Neutrophils\|NCnc\|Pt\|Bld** | 2 | 838.66 | LG32886-0 |
| **318** | **Neutrophils \| Blood \| Hematology and Cell counts** | 3 | 838.66 | LP392641-9 |
| **319** | **Basophils\|NCnc\|Pt\|Bld** | 2 | 838.25 | LG32848-0 |
| **320** | **Basophils \| Blood \| Hematology and Cell counts** | 3 | 838.25 | LP392736-7 |
| **321** | **Eosinophils/100 leukocytes \| Blood \| Hematology and Cell counts** | 3 | 834.97 | LP392798-7 |
| **322** | **Basophils/100 leukocytes \| Blood \| Hematology and Cell counts** | 3 | 831.03 | LP392761-5 |
| **323** | **Eosinophils\|NCnc\|Pt\|Bld** | 2 | 825.17 | LG32849-8 |
| **324** | **Eosinophils \| Blood \| Hematology and Cell counts** | 3 | 825.17 | LP392778-9 |
| **325** | **Lymphocyte T-cell and B-cell and Natural killer subsets panel \| Blood \| Cellmarker Panels** | 4 | 823.64 | LP400859-7 |

| | | | | |
|---|---|---|---|---|
| **326** | **Lymphocyte T-cell and B-cell and Natural killer subsets panel - Blood** | 3 | 823.64 | 80721-4 |
| **327** | **SARSCoV2 antibody detection** | 2 | 822.57 | LG51018-6 |
| **328** | **Aspartate aminotransferase [Enzymatic activity/volume] in Serum or Plasma** | 2 | 819.54 | 1920-8 |
| **329** | **Platelets\|NCnc\|Pt\|Bld** | 2 | 809.07 | LG32892-8 |
| **330** | **Platelets \| Blood \| Hematology and Cell counts** | 3 | 808.55 | LP393218-5 |
| **331** | **Urinalysis dipstick panel \| Urine \| Urinalysis Panels** | 4 | 804.69 | LP402548-4 |
| **332** | **Urinalysis dipstick panel - Urine by Automated test strip** | 3 | 804.69 | 50556-0 |
| **333** | **Electrolytes panel \| Blood \| Chemistry Panels** | 5 | 803.78 | LP386898-3 |
| **334** | **Electrolytes panel - Blood** | 3 | 803.78 | 55231-5 |
| **335** | **aPTT \| Platelet poor plasma \| Coagulation** | 4 | 801.33 | LP393946-1 |
| **336** | **Hematocrit \| Blood \| Hematology and Cell counts** | 3 | 797.03 | LP392479-4 |
| **337** | **Glucose\|Pt\|Ser/Plas\|180.156 g/mole** | 2 | 795.68 | LG49881-2 |
| **338** | **Glucose [Mass/volume] in Serum or Plasma** | 2 | 795.68 | 2345-7 |
| **339** | **Glucose \| Serum or Plasma \| Chemistry - non-challenge** | 3 | 795.68 | LP385540-2 |
| **340** | **Glucose tolerance 2 hours gestational panel - Urine and Serum or Plasma** | 3 | 795.48 | 24353-5 |
| **341** | **Glucose tolerance 2 hours gestational panel \| Urine and Serum or Plasma \| Challenge Bank Panels** | 5 | 795.48 | LP387239-9 |
| **342** | **aPTT in Platelet poor plasma by Coagulation assay** | 2 | 794.58 | 14979-9 |
| **343** | **aPTT panel - Platelet poor plasma** | 3 | 794.58 | 50197-3 |
| **344** | **aPTT panel \| Platelet poor plasma \| Coagulation Panels** | 4 | 794.58 | LP394433-9 |
| **345** | **Activated protein C resistance panel - Platelet poor plasma** | 3 | 794.04 | 48596-1 |
| **346** | **Activated protein C resistance panel \| Platelet poor plasma \| Coagulation Panels** | 4 | 794.04 | LP394432-1 |
| **347** | **Hemodynamics** | 3 | 793.51 | LP29712-4 |
| **348** | **Specific hemodynamics** | 4 | 793.51 | LP7805-7 |
| **349** | **aPTT mixing study panel \| Platelet poor plasma \| Coagulation Panels** | 4 | 793.50 | LP427541-0 |
| **350** | **aPTT mixing study panel - Platelet poor plasma** | 3 | 793.50 | 97024-4 |
| **351** | **Vital signs** | 2 | 792.72 | LP30605-7 |
| **352** | **Glucose post fasting and meal stimulation panel \| Serum or Plasma \| Challenge Bank Panels** | 5 | 791.93 | LP419313-4 |
| **353** | **Glucose post fasting and meal stimulation panel - Serum or Plasma** | 3 | 791.93 | 95102-0 |
| **354** | **Erythrocyte mean corpuscular volume \| Red Blood Cells \| Hematology and Cell counts** | 5 | 790.32 | LP393361-3 |
| **355** | **Monocytes/100 leukocytes \| Blood \| Hematology and Cell counts** | 3 | 769.80 | LP393095-7 |
| **356** | **Leukocytes \| Blood \| Hematology and Cell counts** | 3 | 765.68 | LP392599-9 |
| **357** | **Leukocytes\|NCnc\|Pt\|Bld** | 2 | 765.68 | LG32857-1 |

| | | | | |
|---|---|---|---|---|
| **358** | **Erythrocyte mean corpuscular hemoglobin \| Red Blood Cells \| Hematology and Cell counts** | 5 | 765.05 | LP393353-0 |
| **359** | **MCH [Entitic mass] by Automated count** | 2 | 765.05 | 785-6 |
| **360** | **MCHC [Mass/volume] by Automated count** | 2 | 764.97 | 786-4 |
| **361** | **Erythrocyte mean corpuscular hemoglobin concentration \| Red Blood Cells \| Hematology and Cell counts** | 5 | 764.97 | LP393357-1 |
| **362** | **Serology Panels** | 3 | 759.92 | LP7837-0 |
| **363** | **Sodium [Moles/volume] in Serum or Plasma** | 2 | 759.70 | 2951-2 |
| **364** | **Sodium \| Serum or Plasma \| Chemistry - non-challenge** | 4 | 759.70 | LP386648-2 |
| **365** | **Procalcitonin \| Serum or Plasma \| Chemistry - non-challenge** | 3 | 756.60 | LP381196-7 |
| **366** | **Procalcitonin\|MCnc\|Pt\|ANYBldSerPl** | 2 | 756.60 | LG15749-1 |
| **367** | **Cardiovascular order set** | 2 | 756.36 | PANEL.CV |
| **368** | **Cardiovascular physiologic and EKG assessment panel** | 3 | 756.36 | 45033-8 |
| **369** | **Lipid and glucose panel \| Serum or Plasma \| Chemistry Panels** | 5 | 745.81 | LP419298-7 |
| **370** | **Lipid and glucose panel - Serum or Plasma** | 3 | 745.81 | 95126-9 |
| **371** | **Lymphocytes/100 leukocytes \| Blood \| Hematology and Cell counts** | 3 | 743.24 | LP392998-3 |
| **372** | **Hemoglobin \| Blood \| Hematology and Cell counts** | 3 | 734.76 | LP392452-1 |
| **373** | **Multiple markers** | 3 | 719.29 | LP32772-3 |
| **374** | **Leukocytes\|Urine/Urine sed** | 2 | 702.92 | LG40867-0 |
| **375** | **Protein\|Urine/Urine sed** | 2 | 700.46 | LG40870-4 |
| **376** | **Protein \| Urine \| Urinalysis** | 4 | 700.46 | LP402534-4 |
| **377** | **Platelet mean volume [Entitic volume] in Blood by Automated count** | 2 | 685.18 | 32623-1 |
| **378** | **Platelet indices** | 3 | 685.05 | LP31668-4 |
| **379** | **Platelet mean volume \| Blood \| Hematology and Cell counts** | 4 | 685.05 | LP393244-1 |
| **380** | **Hemoglobin [Mass/volume] in Blood** | 2 | 680.59 | 718-7 |
| **381** | **Hemoglobin\|Pt\|Bld** | 2 | 680.59 | LG44868-4 |
| **382** | **Lactate\|SCnc\|Pt\|ANYBldSerPl** | 2 | 643.30 | LG6039-4 |
| **383** | **Erythrocytes\|NCnc\|Pt\|Bld** | 2 | 630.14 | LG32850-6 |
| **384** | **Erythrocytes \| Blood \| Hematology and Cell counts** | 3 | 630.14 | LP392503-1 |
| **385** | **Clinical panels** | 2 | 627.98 | LP94654-8 |
| **386** | **Clinical NEC (not elsewhere classified) set** | 2 | 627.66 | PANEL.CLIN |
| **387** | **Lactate dehydrogenase 1 \| Serum or Plasma \| Chemistry - non-challenge** | 4 | 619.72 | LP383089-2 |
| **388** | **Lactate dehydrogenase 1 [Enzymatic activity/volume] in Serum or Plasma** | 2 | 619.72 | 2537-9 |
| **389** | **Lactate dehydrogenase 1\|CCnc\|Pt\|ANYBldSerPl** | 2 | 619.72 | LG9048-2 |
| **390** | **Erythrocyte distribution width \| Red Blood Cells \| Hematology and Cell counts** | 5 | 615.94 | LP393348-0 |
| **391** | **Granulocytes.immature\|NCnc\|Pt\|Bld** | 2 | 608.03 | LG33011-4 |

| | | | | |
|---|---|---|---|---|
| **392** | **Immature granulocytes \| Blood \| Hematology and Cell counts** | 3 | 608.03 | LP392634-4 |
| **393** | **Specific gravity\|Rden\|Urine** | 2 | 605.59 | LG35800-8 |
| **394** | **Lymphocytes [#/volume] in Blood** | 2 | 600.78 | 26474-7 |
| **395** | **Procalcitonin [Mass/volume] in Serum or Plasma** | 2 | 600.54 | 33959-8 |
| **396** | **Procalcitonin\|Pt\|Ser/Plas** | 2 | 600.54 | LG51318-0 |
| **397** | **pH\|LsCnc\|Pt\|ANYBldSerPl** | 2 | 597.13 | LG345-5 |
| **398** | **Magnesium and phosphate and lactate panel - Serum or Plasma** | 3 | 595.97 | 97759-5 |
| **399** | **Magnesium and phosphate and lactate panel \| Serum or Plasma \| Chemistry Panels** | 5 | 595.97 | LP427534-5 |
| **400** | **Hemodynamics molecular** | 2 | 593.61 | HEMODYN.MOLEC |
| **401** | **Interleukin 6 \| Serum or Plasma \| Chemistry - non-challenge** | 5 | 593.46 | LP385040-3 |
| **402** | **Interleukin 6 [Mass/volume] in Serum or Plasma** | 2 | 593.46 | 26881-3 |
| **403** | **Bilirubin \| Urine \| Urinalysis** | 4 | 592.45 | LP402524-5 |
| **404** | **Bilirubin\|Urine/Urine sed** | 2 | 592.45 | LG40907-4 |
| **405** | **Sodium and Potassium panel [Moles/volume] - Blood** | 3 | 588.12 | 74353-4 |
| **406** | **Sodium and Potassium panel \| Blood \| Chemistry Panels** | 5 | 588.12 | LP387057-5 |
| **407** | **pH\|Urine/Urine sed** | 2 | 585.96 | LG40956-1 |
| **408** | **pH \| Urine \| Urinalysis** | 4 | 585.96 | LP402533-6 |
| **409** | **Ketones\|Urine/Urine sed** | 2 | 585.69 | LG40908-2 |
| **410** | **Ketones \| Urine \| Urinalysis** | 4 | 585.69 | LP402528-6 |
| **411** | **Small molecules** | 4 | 583.30 | LP31415-0 |
| **412** | **Protein\|MCnc\|Urine** | 2 | 580.19 | LG34512-0 |
| **413** | **pH\|LsCnc\|Urine** | 2 | 579.17 | LG35753-9 |
| **414** | **Basophils/100 leukocytes in Blood** | 2 | 578.23 | 30180-4 |
| **415** | **Ketones\|MCnc\|Urine** | 2 | 577.92 | LG36553-2 |
| **416** | **Neonatal bilirubin panel [Mass/volume] - Serum or Plasma** | 3 | 573.99 | 50189-0 |
| **417** | **Neonatal bilirubin panel \| Serum or Plasma \| Chemistry Panels** | 5 | 573.99 | LP386997-3 |
| **418** | **Color\|Type\|Urine** | 2 | 571.15 | LG36556-5 |
| **419** | **Lactate dehydrogenase \| Serum or Plasma \| Chemistry - non-challenge** | 4 | 567.34 | LP383084-3 |
| **420** | **Lactate dehydrogenase panel - Serum or Plasma** | 3 | 565.03 | 42929-0 |
| **421** | **Lactate dehydrogenase panel \| Serum or Plasma \| Chemistry Panels** | 5 | 565.03 | LP386971-8 |
| **422** | **Hematocrit [Volume Fraction] of Blood** | 2 | 556.55 | 20570-8 |
| **423** | **Platelet and HLA glycoprotein antibody panel \| Blood \| Serology Panels** | 4 | 554.74 | LP419399-3 |
| **424** | **Platelet and HLA glycoprotein antibody panel - Blood** | 3 | 554.74 | 95270-5 |
| **425** | **Nitrite\|Urine/Urine sed** | 2 | 553.97 | LG40866-2 |
| **426** | **Nitrite \| Urine \| Urinalysis** | 4 | 553.97 | LP402532-8 |
| **427** | **Leukocytes \| Urine sediment \| Urinalysis** | 3 | 548.04 | LP402498-2 |

| | | | | |
|---|---|---|---|---|
| **428** | **Platelets [#/volume] in Blood** | 2 | 543.40 | 26515-7 |
| **429** | **Monocytes [#/volume] in Blood** | 2 | 542.43 | 26484-6 |
| **430** | **Bilirubin\|PrThr\|Urine** | 2 | 542.13 | LG35153-2 |
| **431** | **Granulocytes.immature/100 leukocytes \| Blood \| Hematology and Cell counts** | 3 | 538.33 | LP392635-1 |
| **432** | **Neutrophils.segmented/100 leukocytes \| Blood \| Hematology and Cell counts** | 3 | 537.83 | LP392668-2 |
| **433** | **Segmented neutrophils/100 leukocytes in Blood** | 2 | 537.83 | 26505-8 |
| **434** | **Carbon dioxide \| Blood venous \| Chemistry - non-challenge** | 3 | 535.71 | LP383328-4 |
| **435** | **Leukocytes [#/area] in Urine sediment by Microscopy high power field** | 2 | 532.58 | 5821-4 |
| **436** | **Ketones [Mass/volume] in Urine by Test strip** | 2 | 528.93 | 5797-6 |
| **437** | **Ketones\|Pt\|Urine\|Test strip\|270.413 g/mole** | 2 | 528.93 | LG48911-8 |
| **438** | **pH of Urine by Test strip** | 2 | 528.56 | 5803-2 |
| **439** | **Neutrophils.band form/100 leukocytes \| Blood \| Hematology and Cell counts** | 3 | 528.05 | LP392682-3 |
| **440** | **Tumor necrosis factor.alpha\|MCnc\|Pt\|ANYBldSerPl** | 2 | 526.49 | LG1977-0 |
| **441** | **Tumor necrosis factor.alpha \| Serum or Plasma \| Chemistry - non-challenge** | 4 | 526.49 | LP385059-3 |
| **442** | **Tumor necrosis factor.alpha [Mass/volume] in Serum or Plasma** | 2 | 526.49 | 3074-2 |
| **443** | **Tumor necrosis factor.alpha\|Pt\|Ser/Plas** | 2 | 526.49 | LG48007-5 |
| **444** | **Bacteria identified \| Blood \| Microbiology** | 4 | 525.88 | LP373700-6 |
| **445** | **Leukocyte morphology panel - Blood** | 3 | 523.68 | 58407-8 |
| **446** | **Leukocyte morphology panel \| Blood \| Hematology and Cell Count Panels** | 4 | 523.68 | LP393931-3 |
| **447** | **Bicarbonate\|SCnc\|Pt\|ANYBldSerPl** | 2 | 523.33 | LG2807-8 |
| **448** | **Protein [Mass/volume] in Urine by Test strip** | 2 | 518.80 | 5804-0 |
| **449** | **Oxygen saturation\|MFr\|Pt\|Chal:None** | 2 | 506.64 | LG33051-0 |
| **450** | **Nursing physiologic assessment panel** | 3 | 504.42 | 80346-0 |
| **451** | **Nursing physiologic assessment panel \| Patient \| Clinical panels** | 3 | 504.42 | LP428885-0 |
| **452** | **Gas and electrolytes panel \| Blood arterial \| Chemistry Panels** | 5 | 501.04 | LP431676-8 |
| **453** | **Gas and electrolytes panel - Arterial blood** | 3 | 501.04 | 99548-0 |
| **454** | **Carbon dioxide\|PPres\|Pt\|ANYBldSerPl** | 2 | 499.34 | LG344-8 |
| **455** | **Allergy** | 2 | 497.96 | LP7756-2 |
| **456** | **Microbiology CNAMTS panel - Urine** | 3 | 494.83 | 88848-7 |
| **457** | **Microbiology CNAMTS panel \| Urine \| Microbiology Panels** | 5 | 494.83 | LP380052-3 |
| **458** | **Neutrophils [#/volume] in Blood** | 2 | 494.74 | 26499-4 |
| **459** | **Erythrocyte staining** | 3 | 492.93 | LP30927-5 |
| **460** | **Nitrite\|PrThr\|Urine** | 2 | 492.49 | LG35986-5 |
| **461** | **Immature granulocytes/100 leukocytes in Blood by Automated count** | 2 | 490.25 | 71695-1 |

| | | | | |
|---|---|---|---|---|
| **462** | **aPTT in Blood by Coagulation assay** | 2 | 486.04 | 3173-2 |
| **463** | **Adenovirus DNA \| XXX \| Microbiology** | 4 | 485.86 | LP377083-3 |
| **464** | **aPTT \| Blood \| Coagulation** | 4 | 485.58 | LP393943-8 |
| **465** | **Gas and Carbon monoxide and Electrolytes panel - Arterial blood** | 3 | 485.26 | 93685-6 |
| **466** | **Gas and Carbon monoxide and Electrolytes panel \| Blood arterial \| Chemistry Panels** | 5 | 485.26 | LP417156-9 |
| **467** | **Bilirubin.total [Presence] in Urine by Test strip** | 2 | 484.06 | 5770-3 |
| **468** | **Lymphocytes/100 leukocytes in Blood** | 2 | 478.08 | 26478-8 |
| **469** | **Erythrocytes \| Urine \| Urinalysis** | 3 | 477.20 | LP402476-8 |
| **470** | **Basophils [#/volume] in Blood** | 2 | 474.82 | 26444-0 |
| **471** | **Monocytes/100 leukocytes in Blood** | 2 | 472.17 | 26485-3 |
| **472** | **Nitrite [Presence] in Urine by Test strip** | 2 | 467.84 | 5802-4 |
| **473** | **Erythrocytes [#/volume] in Blood by Automated count** | 2 | 462.96 | 789-8 |
| **474** | **Genitourinary assessment panel** | 4 | 460.60 | 80330-4 |
| **475** | **Glucose\|MCnc\|Urine** | 2 | 459.41 | LG35166-4 |
| **476** | **MCV [Entitic volume] by Automated count** | 2 | 458.20 | 787-2 |
| **477** | **Leukocytes [#/volume] in Blood by Automated count** | 2 | 457.70 | 6690-2 |
| **478** | **Eosinophils/100 leukocytes in Blood** | 2 | 457.66 | 26450-7 |
| **479** | **Eosinophils [#/volume] in Blood by Automated count** | 2 | 454.34 | 711-2 |
| **480** | **T-cell helper (CD4) subset panel \| Blood \| Cellmarker Panels** | 4 | 453.27 | LP400869-6 |
| **481** | **T-cell helper (CD4) subset panel - Blood** | 3 | 453.27 | 65758-5 |
| **482** | **SARS-CoV-2 (COVID-19) N protein IgG Ab [Presence] in Serum or Plasma by Immunoassay** | 2 | 451.27 | 99596-9 |
| **483** | **Interleukin 13 \| Serum or Plasma \| Chemistry - non-challenge** | 5 | 449.79 | LP385054-4 |
| **484** | **Interleukin 13 [Mass/volume] in Serum or Plasma** | 2 | 449.79 | 33822-8 |
| **485** | **Interleukin 1 beta [Mass/volume] in Serum or Plasma** | 2 | 448.13 | 13629-1 |
| **486** | **Interleukin 12 [Mass/volume] in Serum or Plasma** | 2 | 448.13 | 41760-0 |
| **487** | **Interleukin 1 beta \| Serum or Plasma \| Chemistry - non-challenge** | 5 | 448.13 | LP385020-5 |
| **488** | **Interleukin 2 \| Serum or Plasma \| Chemistry - non-challenge** | 5 | 448.13 | LP385023-9 |
| **489** | **Interleukin 8 \| Serum or Plasma \| Chemistry - non-challenge** | 5 | 448.13 | LP385046-0 |
| **490** | **Interleukin 10 \| Serum or Plasma \| Chemistry - non-challenge** | 5 | 448.13 | LP385049-4 |
| **491** | **Interleukin 10 [Mass/volume] in Serum or Plasma** | 2 | 448.13 | 26848-2 |
| **492** | **Interleukin 12 \| Serum or Plasma \| Chemistry - non-challenge** | 5 | 448.13 | LP385051-0 |
| **493** | **Interleukin 2 [Mass/volume] in Serum or Plasma** | 2 | 448.13 | 33939-0 |
| **494** | **Interleukin 8 [Mass/volume] in Serum or Plasma** | 2 | 448.13 | 33211-4 |
| **495** | **Mucus\|Urine/Urine sed** | 2 | 445.88 | LG40937-1 |
| **496** | **Drug toxicology** | 2 | 444.44 | LP7790-1 |

| | | | | |
|---|---|---|---|---|
| **497** | **Specific gravity\|Urine/Urine sed** | 2 | 436.52 | LG40958-7 |
| **498** | **Specific gravity \| Urine \| Urinalysis** | 4 | 436.52 | LP402537-7 |
| **499** | **History** | 2 | 430.67 | LP7800-8 |
| **500** | **Gas panel \| Blood arterial \| Chemistry Panels** | 5 | 426.10 | LP386935-3 |
| **501** | **Gas panel - Arterial blood** | 3 | 426.10 | 24336-0 |
| **502** | **Urobilinogen\|MCnc\|Urine** | 2 | 424.03 | LG35156-5 |
| **503** | **Single allergens** | 4 | 417.80 | LP30727-9 |
| **504** | **Molds and Other Microorganisms** | 3 | 417.80 | LP30672-7 |
| **505** | **Metamyelocytes/100 leukocytes \| Blood \| Hematology and Cell counts** | 3 | 413.32 | LP392831-6 |
| **506** | **Gas and Carbon monoxide panel - Arterial blood** | 3 | 410.12 | 24341-0 |
| **507** | **Gas and Carbon monoxide panel \| Blood arterial \| Chemistry Panels** | 5 | 410.12 | LP386930-4 |
| **508** | **Calcium.ionized\|SCnc\|Pt\|ANYBldSerPl** | 2 | 409.01 | LG655-7 |
| **509** | **Cell count and Diff panel with Gluc and Prot - Cerebral spinal fluid** | 3 | 405.70 | 54230-8 |
| **510** | **Cell count and Diff panel with Gluc and Prot \| Cerebral spinal fluid \| Hematology and Cell Count Panels** | 4 | 405.70 | LP393894-3 |
| **511** | **Immature granulocytes [#/volume] in Blood by Automated count** | 2 | 404.84 | 53115-2 |
| **512** | **pH \| Blood venous \| Chemistry - non-challenge** | 4 | 402.79 | LP383438-1 |
| **513** | **Bordetella pertussis and parapertussis and holmesii DNA panel - Specimen by NAA with probe detection** | 3 | 402.55 | 85809-2 |
| **514** | **Bordetella sp DNA panel \| XXX \| Microbiology Panels** | 5 | 402.55 | LP379897-4 |
| **515** | **Bordetella pertussis and Bordetella parapertussis DNA panel \| XXX \| Microbiology Panels** | 5 | 402.55 | LP379891-7 |
| **516** | **Bordetella pertussis and parapertussis and holmesii DNA panel \| XXX \| Microbiology Panels** | 5 | 402.55 | LP379893-3 |
| **517** | **Bordetella pertussis and Bordetella parapertussis DNA panel - Specimen by NAA with probe detection** | 3 | 402.55 | 41875-6 |
| **518** | **Bordetella sp DNA panel - Specimen by NAA with probe detection** | 3 | 402.55 | 62426-2 |
| **519** | **Acylcarnitine panel - Serum or Plasma** | 3 | 400.84 | 43433-2 |
| **520** | **Acylcarnitine panel \| Serum or Plasma \| Chemistry Panels** | 5 | 400.84 | LP386797-7 |
| **521** | **Mucus \| Urine sediment \| Urinalysis** | 3 | 395.88 | LP402520-3 |
| **522** | **Other elements \| Urine sediment \| Urinalysis Panels** | 3 | 393.98 | LP402545-0 |
| **523** | **Other elements in Urine sediment** | 3 | 393.98 | 58441-7 |
| **524** | **Leukocyte inclusions** | 3 | 390.44 | LP30934-1 |
| **525** | **Glucose \| Urine \| Urinalysis** | 3 | 390.43 | LP402526-0 |
| **526** | **Glucose\|Urine/Urine sed** | 2 | 390.43 | LG40865-4 |
| **527** | **Bacteria identified in Blood by Culture** | 2 | 388.65 | 600-7 |
| **528** | **Interferon gamma [Mass/volume] in Serum or Plasma** | 2 | 386.47 | 27415-9 |
| **529** | **Interferon gamma \| Serum or Plasma \| Hematology and Cell counts** | 3 | 386.47 | LP393788-7 |

| | | | | |
|---|---|---|---|---|
| **530** | **Leukocyte esterase \| Urine \| Urinalysis** | 4 | 384.67 | LP402530-2 |
| **531** | **Leukocyte esterase\|Urine/Urine sed** | 2 | 384.67 | LG40953-8 |
| **532** | **Leukocyte esterase\|PrThr\|Urine** | 2 | 384.67 | LG36980-7 |
| **533** | **Differential cell count method \| Blood \| Hematology and Cell counts** | 4 | 383.79 | LP393809-1 |
| **534** | **Differential cell count method - Blood** | 2 | 383.79 | 49024-3 |
| **535** | **SARS-CoV-2 (COVID-19) Ab panel - Serum or Plasma by Immunoassay** | 3 | 382.42 | 94504-8 |
| **536** | **SARS-CoV-2 (COVID-19) Ab panel \| Serum or Plasma \| Microbiology Panels** | 5 | 382.42 | LP419286-2 |
| **537** | **Urobilinogen \| Urine \| Chemistry - non-challenge** | 4 | 381.95 | LP385341-5 |
| **538** | **Erythrocytes \| Urine sediment \| Urinalysis** | 3 | 376.98 | LP402477-6 |
| **539** | **Albumin/Globulin \| Serum or Plasma \| Chemistry - non-challenge** | 3 | 376.36 | LP384513-0 |
| **540** | **Albumin/Globulin [Mass Ratio] in Serum or Plasma** | 2 | 376.36 | 1759-0 |
| **541** | **Albumin/Globulin\|MRto\|Pt\|ANYBldSerPl** | 2 | 376.36 | LG5164-1 |
| **542** | **Carbon dioxide \| Blood \| Chemistry - non-challenge** | 3 | 375.21 | LP383321-9 |
| **543** | **Interleukin 4 [Mass/volume] in Serum or Plasma** | 2 | 375.06 | 27161-9 |
| **544** | **Interleukin 4 \| Serum or Plasma \| Chemistry - non-challenge** | 5 | 375.06 | LP385034-6 |
| **545** | **Oxygen \| Blood venous \| Chemistry - non-challenge** | 3 | 374.74 | LP383367-2 |
| **546** | **Myelocytes/100 leukocytes \| Blood \| Hematology and Cell counts** | 3 | 372.32 | LP392851-4 |
| **547** | **Calcium.ionized \| Blood \| Chemistry - non-challenge** | 5 | 370.78 | LP385973-5 |
| **548** | **Polychromasia \| Blood \| Hematology and Cell counts** | 4 | 367.50 | LP393447-0 |
| **549** | **Polychromasia [Presence] in Blood by Light microscopy** | 2 | 367.50 | 10378-8 |
| **550** | **MCV [Entitic volume]** | 2 | 367.43 | 30428-7 |
| **551** | **Anion gap\|SCnc\|Pt\|ANYBldSerPl** | 2 | 365.84 | LG13614-9 |
| **552** | **IEEE 11073 Rosetta cardiovascular panel Chorionic villus sample** | 2 | 364.14 | 75962-1 |
| **553** | **Erythrocytes\|PrThr\|Urine** | 2 | 362.39 | LG36247-1 |
| **554** | **Respiratory pathogens RNA 8 panel - Specimen by NAA with probe detection** | 3 | 362.37 | 76771-5 |
| **555** | **Respiratory pathogens RNA 8 panel \| XXX \| Microbiology Panels** | 5 | 362.37 | LP380090-3 |
| **556** | **Leukocyte esterase [Presence] in Urine by Test strip** | 2 | 359.99 | 5799-2 |
| **557** | **Bilirubin.indirect [Mass/volume] in Serum or Plasma** | 2 | 358.74 | 1971-1 |
| **558** | **Bilirubin.non-glucuronidated\|MCnc\|Pt\|ANYBldSerPl** | 2 | 358.74 | LG3235-1 |
| **559** | **Bilirubin.non-glucuronidated \| Serum or Plasma \| Chemistry - non-challenge** | 4 | 358.74 | LP385316-7 |
| **560** | **Bilirubin.non-glucuronidated\|Pt\|Ser/Plas** | 2 | 358.74 | LG43430-4 |
| **561** | **Drug allergens** | 3 | 357.60 | LP30682-6 |
| **562** | **Sodium [Moles/volume] in Blood** | 2 | 355.61 | 2947-0 |
| **563** | **Sodium \| Blood \| Chemistry - non-challenge** | 4 | 355.61 | LP386631-8 |
| **564** | **Band form neutrophils/100 leukocytes in Blood** | 2 | 353.26 | 26508-2 |

| | | | | |
|---|---|---|---|---|
| **565** | **Metamyelocytes/100 leukocytes in Blood** | 2 | 352.88 | 28541-1 |
| **566** | **Parainfluenza virus 1 RNA \| XXX \| Microbiology** | 4 | 351.39 | LP378758-9 |
| **567** | **Influenza virus types A and B and subtypes RNA panel - Specimen by NAA with probe detection** | 3 | 351.04 | 97733-0 |
| **568** | **Influenza virus types A and B and subtypes RNA panel \| XXX \| Microbiology Panels** | 5 | 351.04 | LP427045-2 |
| **569** | **Human metapneumovirus RNA \| XXX \| Microbiology** | 4 | 348.97 | LP378241-6 |
| **570** | **Human metapneumovirus RNA [Presence] in Specimen by NAA with probe detection** | 2 | 348.81 | 38917-1 |
| **571** | **Parainfluenza virus 1 RNA [Presence] in Specimen by NAA with probe detection** | 2 | 348.23 | 29908-1 |
| **572** | **Erythrocytes [Presence] in Urine** | 2 | 347.78 | 33051-4 |
| **573** | **Urobilinogen\|Pt\|Urine\|592.737 g/mole** | 2 | 344.79 | LG47119-9 |
| **574** | **Urobilinogen [Mass/volume] in Urine** | 2 | 344.79 | 3107-0 |
| **575** | **Clusters of differentiation** | 3 | 343.69 | LP32944-8 |
| **576** | **Gamma glutamyl transferase [Enzymatic activity/volume] in Serum or Plasma** | 2 | 342.41 | 2324-2 |
| **577** | **Gamma glutamyl transferase \| Serum or Plasma \| Chemistry - non-challenge** | 4 | 342.41 | LP383003-3 |
| **578** | **Leukocytes [#/volume] in Blood** | 2 | 338.88 | 26464-8 |
| **579** | **Cell count and Differential panel \| Cerebral spinal fluid \| Hematology and Cell Count Panels** | 4 | 337.48 | LP393885-1 |
| **580** | **Cell count and Differential panel - Cerebral spinal fluid** | 3 | 337.48 | 34564-5 |
| **581** | **Lactate dehydrogenase [Enzymatic activity/volume] in Serum or Plasma** | 2 | 337.45 | 2532-0 |
| **582** | **Escherichia sp** | 4 | 335.10 | LP30565-3 |
| **583** | **Magnesium [Mass/volume] in Serum or Plasma** | 2 | 334.75 | 19123-9 |
| **584** | **Magnesium\|MCnc\|Pt\|ANYBldSerPl** | 2 | 334.75 | LG5903-2 |
| **585** | **Magnesium\|Pt\|Ser/Plas\|24.305 g/mole** | 2 | 334.64 | LG50041-9 |
| **586** | **Magnesium \| Serum or Plasma \| Chemistry - non-challenge** | 5 | 334.64 | LP386028-7 |
| **587** | **Human coronavirus NL63 RNA [Presence] in Specimen by NAA with probe detection** | 2 | 334.21 | 41005-0 |
| **588** | **Human coronavirus NL63 RNA \| XXX \| Microbiology** | 4 | 334.21 | LP377181-5 |
| **589** | **Platelet morphology panel \| Blood \| Hematology and Cell Count Panels** | 4 | 333.53 | LP393935-4 |
| **590** | **Platelet morphology panel - Blood** | 3 | 333.53 | 58406-0 |
| **591** | **Human coronavirus 229E RNA \| XXX \| Microbiology** | 4 | 332.77 | LP377166-6 |
| **592** | **Human coronavirus 229E RNA [Presence] in Specimen by NAA with probe detection** | 2 | 332.77 | 41003-5 |
| **593** | **Adenovirus DNA [Presence] in Specimen by NAA with probe detection** | 2 | 330.84 | 39528-5 |
| **594** | **Human coronavirus OC43 RNA \| XXX \| Microbiology** | 4 | 330.28 | LP377189-8 |
| **595** | **Human coronavirus OC43 RNA [Presence] in Specimen by NAA with probe detection** | 2 | 330.28 | 41009-2 |
| **596** | **Oxygen [Partial pressure] in Venous blood** | 2 | 327.92 | 2705-2 |

| | | | | |
|---|---|---|---|---|
| **597** | **pH of Venous blood** | 2 | 327.92 | 2746-6 |
| **598** | **Meningitis+Encephalitis pathogens panel \| Cerebral spinal fluid \| Microbiology Panels** | 5 | 327.73 | LP380040-8 |
| **599** | **Color of Urine by Auto** | 2 | 322.61 | 50553-7 |
| **600** | **Eosinophils/100 leukocytes in Blood by Automated count** | 2 | 318.85 | 713-8 |
| **601** | **Cardiac ultrasound** | 3 | 318.56 | LP29672-0 |
| **602** | **Lactate [Moles/volume] in Venous blood** | 2 | 317.88 | 2519-7 |
| **603** | **Lactate \| Blood venous \| Chemistry - non-challenge** | 5 | 317.88 | LP385566-7 |
| **604** | **Lactate\|Pt\|BldV** | 2 | 317.88 | LG48937-3 |
| **605** | **Bordetella pertussis** | 2 | 316.24 | LG41615-2 |
| **606** | **Parainfluenza virus 3 RNA \| XXX \| Microbiology** | 4 | 314.13 | LP378798-5 |
| **607** | **Parainfluenza virus 3 RNA [Presence] in Specimen by NAA with probe detection** | 2 | 310.93 | 29910-7 |
| **608** | **Epstein Barr virus nuclear IgG \| Serum \| Microbiology** | 4 | 304.46 | LP377555-0 |
| **609** | **Creatine kinase \| Serum or Plasma \| Chemistry - non-challenge** | 4 | 303.48 | LP382927-4 |
| **610** | **Creatine kinase [Enzymatic activity/volume] in Serum or Plasma** | 2 | 303.48 | 2157-6 |
| **611** | **Bicarbonate [Moles/volume] in Venous blood** | 2 | 303.37 | 14627-4 |
| **612** | **Bicarbonate \| Blood venous \| Chemistry - non-challenge** | 4 | 303.37 | LP386559-1 |
| **613** | **Creatine kinase panel \| Serum or Plasma \| Chemistry Panels** | 5 | 302.94 | LP386869-4 |
| **614** | **Creatine kinase panel - Serum or Plasma** | 3 | 302.94 | 24335-2 |
| **615** | **Carbon dioxide [Partial pressure] in Venous blood** | 2 | 296.52 | 2021-4 |
| **616** | **Platelets [#/volume] in Blood by Automated count** | 2 | 295.81 | 777-3 |
| **617** | **Neutrophils/100 leukocytes \| Blood \| Hematology and Cell counts** | 3 | 293.40 | LP392722-7 |
| **618** | **Human coronavirus HKU1 RNA [Presence] in Specimen by NAA with probe detection** | 2 | 293.37 | 62423-9 |
| **619** | **Human coronavirus HKU1 RNA \| XXX \| Microbiology** | 4 | 293.37 | LP377174-0 |
| **620** | **Basophils [#/volume] in Blood by Automated count** | 2 | 292.55 | 704-7 |
| **621** | **Globulin \| Serum \| Chemistry - non-challenge** | 4 | 289.26 | LP384596-5 |
| **622** | **Glucose [Mass/volume] in Urine by Test strip** | 2 | 288.15 | 5792-7 |
| **623** | **Glucose\|Pt\|Urine\|Test strip\|180.156 g/mole** | 2 | 288.15 | LG33250-8 |
| **624** | **Eosinophils [#/volume] in Blood** | 2 | 286.93 | 26449-9 |
| **625** | **Nucleated erythrocytes \| Blood \| Hematology and Cell counts** | 3 | 286.62 | LP392558-5 |
| **626** | **Erythrocytes.nucleated\|NCnc\|Pt\|Bld** | 2 | 286.62 | LG32999-1 |
| **627** | **PhenX - respiratory - spirometry protocol 091601** | 3 | 286.60 | 62639-0 |
| **628** | **Differential panel \| Cerebral spinal fluid \| Hematology and Cell Count Panels** | 4 | 286.43 | LP393910-7 |
| **629** | **Differential panel - Cerebral spinal fluid** | 3 | 286.43 | 29584-0 |
| **630** | **Calcium.ionized\|Pt\|Bld** | 2 | 284.31 | LG42928-8 |

| | | | | |
|---|---|---|---|---|
| **631** | **Mycoplasma pneumoniae DNA [Presence] in Specimen by NAA with probe detection** | 2 | 283.90 | 29257-3 |
| **632** | **Mycoplasma pneumoniae DNA \| XXX \| Microbiology** | 4 | 283.90 | LP375335-9 |
| **633** | **Cell count and Diff panel with Gluc and Prot \| Body fluid \| Hematology and Cell Count Panels** | 4 | 280.59 | LP393893-5 |
| **634** | **Cell count and Diff panel with Gluc and Prot - Body fluid** | 3 | 280.59 | 54234-0 |
| **635** | **Nucleated erythrocytes [#/volume] in Blood** | 2 | 278.15 | 30392-5 |
| **636** | **Cell count and Differential panel - Body fluid** | 3 | 277.60 | 34557-9 |
| **637** | **Cell count and Differential panel \| Body fluid \| Hematology and Cell Count Panels** | 4 | 277.60 | LP393884-4 |
| **638** | **Lipoprotein lipase [Enzymatic activity/volume] in Serum or Plasma** | 2 | 277.44 | 2572-6 |
| **639** | **Lipoprotein lipase \| Serum or Plasma \| Chemistry - non-challenge** | 4 | 277.44 | LP383130-4 |
| **640** | **Lymphocytes [#/volume] in Blood by Automated count** | 2 | 274.88 | 731-0 |
| **641** | **Neutrophils [#/volume] in Blood by Automated count** | 2 | 273.88 | 751-8 |
| **642** | **Neutrophil.PMA stimulated DHR panel by Flow cytometry (FC)** | 3 | 273.88 | 98124-1 |
| **643** | **Neutrophil.fMLP and PMA stimulated DHR panel by Flow cytometry (FC)** | 3 | 273.88 | 98122-5 |
| **644** | **Neutrophil.fMLP stimulated DHR panel \| Patient \| Hematology and Cell Count Panels** | 4 | 273.88 | LP428842-1 |
| **645** | **Neutrophil.fMLP and PMA stimulated DHR panel \| Patient \| Hematology and Cell Count Panels** | 4 | 273.88 | LP428841-3 |
| **646** | **Neutrophil.PMA stimulated DHR panel \| Patient \| Hematology and Cell Count Panels** | 4 | 273.88 | LP428843-9 |
| **647** | **Neutrophil.fMLP stimulated DHR panel by Flow cytometry (FC)** | 3 | 273.88 | 98123-3 |
| **648** | **Monocytes [#/volume] in Blood by Automated count** | 2 | 273.73 | 742-7 |
| **649** | **Microbiology CNAMTS panel \| XXX \| Microbiology Panels** | 5 | 272.69 | LP380054-9 |
| **650** | **Microbiology CNAMTS panel - Specimen** | 3 | 272.69 | 88845-3 |
| **651** | **Bilirubin.direct [Mass/volume] in Serum or Plasma** | 2 | 267.29 | 1968-7 |
| **652** | **Bilirubin.glucuronidated+Bilirubin.albumin bound\|Pt\|Ser/Plas** | 2 | 267.29 | LG43215-9 |
| **653** | **Bilirubin.glucuronidated+Bilirubin.albumin bound \| Serum or Plasma \| Chemistry - non-challenge** | 4 | 267.29 | LP385310-0 |
| **654** | **Bilirubin.glucuronidated+Bilirubin.albumin bound\|MCnc\|Pt\|ANYBldSerPl** | 2 | 267.29 | LG6165-7 |
| **655** | **Natural killer cell panel \| Blood \| Cellmarker Panels** | 4 | 263.66 | LP400865-4 |
| **656** | **Natural killer cell panel - Blood by Flow cytometry (FC)** | 3 | 263.66 | 53759-7 |
| **657** | **Natural killer and natural killer T cell subsets panel - Blood** | 3 | 263.31 | 98073-0 |
| **658** | **Natural killer and natural killer T cell subsets panel \| Blood \| Cellmarker Panels** | 4 | 263.31 | LP428204-4 |

| | | | | |
|---|---|---|---|---|
| **659** | **Calcium.ionized [Moles/volume] in Blood** | 2 | 262.65 | 1994-3 |
| **660** | **Chlamydophila pneumoniae DNA \| XXX \| Microbiology** | 4 | 262.16 | LP374549-6 |
| **661** | **Chlamydophila pneumoniae DNA [Presence] in Specimen by NAA with probe detection** | 2 | 262.16 | 34645-2 |
| **662** | **Output assessment panel** | 4 | 262.03 | 81677-7 |
| **663** | **Platelet Function** | 3 | 260.60 | LP31621-3 |
| **664** | **Erythrocytes.nucleated/100 leukocytes \| Blood \| Hematology and Cell counts** | 3 | 260.04 | LP392590-8 |
| **665** | **Anion gap in Serum or Plasma** | 2 | 255.68 | 33037-3 |
| **666** | **Anion gap \| Serum or Plasma \| Chemistry - non-challenge** | 4 | 255.68 | LP386539-3 |
| **667** | **Phosphate [Moles/volume] in Specimen** | 2 | 254.50 | 22733-0 |
| **668** | **Phosphate \| XXX \| Chemistry - non-challenge** | 5 | 254.50 | LP386062-6 |
| **669** | **Phosphate\|Pt\|XXX\|94.97 g/mole** | 2 | 254.50 | LG49946-3 |
| **670** | **CBC W Differential panel \| Blood cord \| Hematology and Cell Count Panels** | 4 | 253.17 | LP393879-4 |
| **671** | **CBC WO Differential panel \| Blood cord \| Hematology and Cell Count Panels** | 4 | 253.17 | LP393883-6 |
| **672** | **Erythrocyte distribution width \| Blood cord \| Hematology and Cell counts** | 5 | 253.17 | LP393347-2 |
| **673** | **CBC W Differential panel - Cord blood** | 3 | 253.17 | 74412-8 |
| **674** | **CBC WO Differential panel - Cord blood** | 3 | 253.17 | 47288-6 |
| **675** | **Erythrocyte distribution width [Ratio] in Cord blood** | 2 | 253.17 | 47277-9 |
| **676** | **Q-T interval \| {Electrocardiograph lead} \| EKG measurements** | 4 | 252.89 | LP407041-5 |
| **677** | **Q-T interval {Electrocardiograph lead}** | 3 | 252.89 | 44975-1 |
| **678** | **QRS duration {Electrocardiograph lead}** | 3 | 252.39 | 44973-6 |
| **679** | **QRS duration \| {Electrocardiograph lead} \| EKG measurements** | 4 | 252.39 | LP407001-9 |
| **680** | **P-R Interval \| {Electrocardiograph lead} \| EKG measurements** | 4 | 251.08 | LP406955-7 |
| **681** | **P-R Interval {Electrocardiograph lead}** | 3 | 251.08 | 44976-9 |
| **682** | **Lymphocytes.variant/100 leukocytes \| Blood \| Hematology and Cell counts** | 3 | 250.56 | LP392973-6 |
| **683** | **Variant lymphocytes/100 leukocytes in Blood by Manual count** | 2 | 250.56 | 735-1 |
| **684** | **Base deficit\|SCnc\|Pt\|ANYBldSerPl** | 2 | 250.35 | LG6037-8 |
| **685** | **Erythrocyte distribution width [Ratio]** | 2 | 249.69 | 30385-9 |
| **686** | **Hematocrit [Volume Fraction] of Blood by Automated count** | 2 | 249.52 | 4544-3 |
| **687** | **Color of Urine** | 2 | 249.44 | 5778-6 |
| **688** | **Erythrocyte distribution width [Ratio] by Automated count** | 2 | 248.98 | 788-0 |
| **689** | **FEV1 \| Respiratory system \| Respiratory measures and Ventilator management** | 3 | 247.99 | LP404751-2 |

| | | | | |
|---|---|---|---|---|
| **690** | **Erythrocyte shape \| Blood \| Hematology and Cell counts** | 3 | 246.86 | LP393396-9 |
| **691** | **Erythrocyte shape [Morphology] in Blood** | 2 | 246.86 | 18225-3 |
| **692** | **Urinalysis microscopic panel [#/area] - Urine sediment by Automated count** | 3 | 246.43 | 53293-7 |
| **693** | **Cardiac 2D echo panel** | 3 | 245.90 | 34552-0 |
| **694** | **Cardiac 2D echo panel \| Heart \| Cardiac Panels** | 4 | 245.90 | LP431680-0 |
| **695** | **Forced vital capacity \| Respiratory system \| Respiratory measures and Ventilator management** | 3 | 240.17 | LP408945-6 |
| **696** | **Zimnitsky urine concentration panel \| Urine \| Urinalysis Panels** | 4 | 237.90 | LP428714-2 |
| **697** | **Zimnitsky urine concentration panel - Urine** | 3 | 237.90 | 98126-6 |
| **698** | **Drug and Toxicology Panels** | 3 | 236.76 | LP29683-7 |
| **699** | **Burr cells [Presence] in Blood by Light microscopy** | 2 | 236.19 | 7790-9 |
| **700** | **Burr cells \| Blood \| Hematology and Cell counts** | 3 | 236.19 | LP393403-3 |
| **701** | **Anti-infection drugs** | 3 | 234.75 | LP31450-7 |
| **702** | **Parainfluenza virus 2 RNA \| XXX \| Microbiology** | 4 | 234.55 | LP378778-7 |
| **703** | **Cell count panel - Body fluid** | 3 | 234.46 | 34556-1 |
| **704** | **Cell count panel \| Body fluid \| Hematology and Cell Count Panels** | 4 | 234.46 | LP393895-0 |
| **705** | **Potassium \| Blood \| Chemistry - non-challenge** | 4 | 234.25 | LP386601-1 |
| **706** | **Potassium [Moles/volume] in Blood** | 2 | 234.25 | 6298-4 |
| **707** | **Potassium\|Pt\|Bld\|39.098 g/mole** | 2 | 234.25 | LG49939-8 |
| **708** | **Drug level & Toxicology order set** | 2 | 234.18 | PANEL.DRUG/TOX |
| **709** | **pH \| Blood \| Chemistry - non-challenge** | 4 | 232.71 | LP383431-6 |
| **710** | **LMW Heparin \| Platelet poor plasma \| Coagulation** | 4 | 232.25 | LP394265-5 |
| **711** | **Rhinovirus+Enterovirus RNA \| XXX \| Microbiology** | 4 | 231.64 | LP377510-5 |
| **712** | **Rhinovirus+Enterovirus RNA [Presence] in Specimen by NAA with probe detection** | 2 | 231.64 | 40991-2 |
| **713** | **Parainfluenza virus 2 RNA [Presence] in Specimen by NAA with probe detection** | 2 | 231.49 | 29909-9 |
| **714** | **Mycobacterium tuberculosis stimulated gamma interferon panel - Blood** | 3 | 231.34 | 71775-1 |
| **715** | **Mycobacterium tuberculosis stimulated gamma interferon panel \| Blood \| Microbiology Panels** | 5 | 231.34 | LP380067-1 |
| **716** | **Immunodeficiency panel \| Blood \| Cellmarker Panels** | 4 | 230.75 | LP400856-3 |
| **717** | **Immunodeficiency panel - Blood by Flow cytometry (FC)** | 3 | 230.75 | 45268-0 |
| **718** | **Coagulation factor VIII Activity.Xa activator [Units/volume] in Platelet poor plasma by Chromogenic method** | 2 | 229.74 | 3212-8 |
| **719** | **Coagulation factor VIII Activity.Xa activator \| Platelet poor plasma \| Coagulation** | 4 | 229.74 | LP394183-0 |
| **720** | **Epstein Barr virus nuclear IgG Ab [Units/volume] in Serum by Immunoassay** | 2 | 227.59 | 30083-0 |
| **721** | **Parainfluenza virus 4 RNA \| XXX \| Microbiology** | 4 | 225.84 | LP378809-0 |

| | | | | |
|---|---|---|---|---|
| **722** | **Parainfluenza virus 4 RNA [Presence] in Specimen by NAA with probe detection** | 2 | 225.84 | 41010-0 |
| **723** | **pH of Blood** | 2 | 225.44 | 11558-4 |
| **724** | **Clarity\|Type\|Urine** | 2 | 224.34 | LG35818-0 |
| **725** | **Clarity of Urine** | 2 | 224.34 | 32167-9 |
| **726** | **Bacteria\|Urine/Urine sed** | 2 | 223.96 | LG40875-3 |
| **727** | **Bacteria \| Urine sediment \| Urinalysis** | 3 | 223.96 | LP402462-8 |
| **728** | **Myelocytes/100 leukocytes in Blood** | 2 | 223.71 | 26498-6 |
| **729** | **Bordetella pertussis DNA [Presence] in Specimen by NAA with probe detection** | 2 | 222.80 | 23826-1 |
| **730** | **Bordetella pertussis DNA \| XXX \| Microbiology** | 4 | 222.80 | LP373986-1 |
| **731** | **Amylase and triacylglycerol lipase panel - Serum or Plasma** | 3 | 221.52 | 72272-8 |
| **732** | **Amylase and triacylglycerol lipase panel \| Serum or Plasma \| Chemistry Panels** | 5 | 221.52 | LP386811-6 |
| **733** | **Cytomegalovirus IgG \| Serum or Plasma \| Microbiology** | 4 | 221.24 | LP377253-2 |
| **734** | **Epithelial cells.squamous\|Urine/Urine sed** | 2 | 219.53 | LG40856-3 |
| **735** | **Respiratory syncytial virus RNA [Presence] in Isolate by NAA with probe detection** | 2 | 218.48 | 60271-4 |
| **736** | **Respiratory syncytial virus RNA \| Isolate \| Microbiology** | 4 | 218.48 | LP378955-1 |
| **737** | **Immature granulocytes [#/volume] in Blood** | 2 | 218.41 | 51584-1 |
| **738** | **SARS-CoV-2 (COVID-19) IgG \| Serum or Plasma \| Microbiology** | 4 | 218.39 | LP418688-0 |
| **739** | **Anisocytosis [Presence] in Blood by Light microscopy** | 2 | 215.79 | 702-1 |
| **740** | **Anisocytosis \| Blood \| Hematology and Cell counts** | 3 | 215.79 | LP393399-3 |
| **741** | **Globulin [Mass/volume] in Serum by calculation** | 2 | 215.31 | 10834-0 |
| **742** | **Appearance of Urine** | 2 | 213.24 | 5767-9 |
| **743** | **Lymphocytes/100 leukocytes in Blood by Automated count** | 2 | 210.61 | 736-9 |
| **744** | **Urobilinogen\|Urine/Urine sed** | 2 | 210.40 | LG40864-7 |
| **745** | **Urobilinogen \| Urine \| Urinalysis** | 4 | 210.40 | LP402538-5 |
| **746** | **Neutrophils/100 leukocytes in Blood by Automated count** | 2 | 210.03 | 770-8 |
| **747** | **Adulterants panel \| Urine \| Chemistry Panels** | 5 | 209.27 | LP386799-3 |
| **748** | **Adulterants panel - Urine** | 3 | 209.27 | 58715-4 |
| **749** | **Monocytes/100 leukocytes in Blood by Automated count** | 2 | 209.02 | 5905-5 |
| **750** | **Hemoglobin [Presence] in Urine by Test strip** | 2 | 207.99 | 5794-3 |
| **751** | **Hemoglobin\|PrThr\|Urine** | 2 | 207.99 | LG36977-3 |
| **752** | **Vancomycin \| Serum or Plasma \| Drug toxicology** | 4 | 207.62 | LP389062-3 |
| **753** | **Neutrophils.vacuolated \| Blood \| Hematology and Cell counts** | 3 | 205.60 | LP392707-8 |
| **754** | **Neutrophils.vacuolated [Presence] in Blood by Light microscopy** | 2 | 205.60 | 18319-4 |

| | | | | |
|---|---|---|---|---|
| **755** | **Specific gravity of Urine by Test strip** | 2 | 205.47 | 5811-5 |
| **756** | **Hyaline casts\|Urine/Urine sed** | 2 | 204.61 | LG40890-2 |
| **757** | **Base excess in Blood by calculation** | 2 | 201.76 | 11555-0 |
| **758** | **Base excess \| Blood \| Chemistry - non-challenge** | 5 | 201.76 | LP383304-5 |
| **759** | **Cytomegalovirus IgM \| Serum or Plasma \| Microbiology** | 4 | 200.09 | LP377260-7 |
| **760** | **Complement profile panel [Mass/volume] - Serum or Plasma** | 3 | 199.75 | 34547-0 |
| **761** | **Complement C3 and C4 panel \| Serum or Plasma \| Hematology and Cell Count Panels** | 4 | 199.75 | LP393903-2 |
| **762** | **Complement profile panel \| Serum or Plasma \| Hematology and Cell Count Panels** | 4 | 199.75 | LP393904-0 |
| **763** | **Complement C3 and C4 panel [Mass/volume] - Serum or Plasma** | 3 | 199.75 | 34544-7 |
| **764** | **Base excess \| Blood venous \| Chemistry - non-challenge** | 5 | 199.29 | LP383311-0 |
| **765** | **Lactate \| Serum or Plasma \| Chemistry - non-challenge** | 5 | 198.97 | LP385575-8 |
| **766** | **Lactate\|Pt\|Ser/Plas** | 2 | 198.97 | LG48941-5 |
| **767** | **Meningitis+Encephalitis pathogens panel - Cerebral spinal fluid by NAA with probe detection** | 3 | 198.75 | 99087-9 |
| **768** | **Monocytes/100 leukocytes \| Cerebral spinal fluid \| Hematology and Cell counts** | 3 | 196.39 | LP393099-9 |
| **769** | **Hemoglobin \| Urine \| Urinalysis** | 3 | 195.52 | LP402527-8 |
| **770** | **Hemoglobin\|Urine/Urine sed** | 2 | 195.52 | LG40952-0 |
| **771** | **Fibrin D-dimer [Presence] in Platelet poor plasma** | 2 | 195.29 | 15179-5 |
| **772** | **Fibrin D-dimer \| Platelet poor plasma \| Coagulation** | 4 | 195.29 | LP394010-5 |
| **773** | **Giant platelets \| Blood \| Hematology and Cell counts** | 3 | 194.22 | LP393233-4 |
| **774** | **Giant platelets [Presence] in Blood by Light microscopy** | 2 | 194.22 | 5908-9 |
| **775** | **Epithelial cells.squamous \| Urine sediment \| Urinalysis** | 3 | 193.85 | LP402472-7 |
| **776** | **Monocytes/100 leukocytes in Cerebral spinal fluid** | 2 | 193.82 | 26486-1 |
| **777** | **Glucose \| Urine \| Chemistry - non-challenge** | 3 | 193.56 | LP385545-1 |
| **778** | **Base excess in Venous blood by calculation** | 2 | 192.10 | 1927-3 |
| **779** | **National EMS Information System - version 3 set** | 3 | 191.60 | 84428-2 |
| **780** | **National EMS Information System data set** | 2 | 191.60 | PANEL.NEMSIS |
| **781** | **Cytology** | 2 | 191.10 | LP7789-3 |
| **782** | **Toxic granules [Presence] in Blood by Light microscopy** | 2 | 191.01 | 803-7 |
| **783** | **Toxic granules \| Blood \| Hematology and Cell counts** | 4 | 191.01 | LP393455-3 |
| **784** | **Oxygen \| Blood \| Chemistry - non-challenge** | 3 | 190.92 | LP383360-7 |
| **785** | **Mucus [Presence] in Urine sediment by Light microscopy** | 2 | 190.86 | 8247-9 |
| **786** | **Leukocytes\|NCnc\|Urine** | 2 | 190.64 | LG35159-9 |
| **787** | **Leukocytes \| Urine \| Urinalysis** | 3 | 190.64 | LP402497-4 |

| | | | | |
|---|---|---|---|---|
| **788** | **Carbon dioxide, total [Moles/volume] in Blood by calculation** | 2 | 190.43 | 34728-6 |
| **789** | **Urinalysis microscopic panel \| Urine \| Urinalysis Panels** | 4 | 190.12 | LP402553-4 |
| **790** | **Bilirubin.conjugated [Mass/volume] in Serum or Plasma** | 2 | 190.11 | 15152-2 |
| **791** | **Bilirubin.glucuronidated\|Pt\|Ser/Plas** | 2 | 190.11 | LG42870-2 |
| **792** | **Bilirubin.glucuronidated \| Serum or Plasma \| Chemistry - non-challenge** | 4 | 190.11 | LP385304-3 |
| **793** | **Lactate dehydrogenase [Enzymatic activity/volume] in Serum or Plasma by Pyruvate to lactate reaction** | 2 | 189.44 | 14805-6 |
| **794** | **Basophils/100 leukocytes in Blood by Automated count** | 2 | 187.01 | 706-2 |
| **795** | **Heterophile Ab \| Serum \| Microbiology** | 4 | 185.32 | LP377561-8 |
| **796** | **Phosphate \| Serum or Plasma \| Chemistry - non-challenge** | 5 | 185.22 | LP386057-6 |
| **797** | **Phosphate [Mass/volume] in Serum or Plasma** | 2 | 185.22 | 2777-1 |
| **798** | **Phosphate\|Pt\|Ser/Plas\|94.97 g/mole** | 2 | 185.22 | LG49949-7 |
| **799** | **Phosphate\|MCnc\|Pt\|ANYBldSerPl** | 2 | 185.22 | LG6426-3 |
| **800** | **Virtual Medical Record for Clinical Decision Support panel \| Patient \| Clinical panels** | 3 | 185.08 | LP428898-3 |
| **801** | **Virtual Medical Record for Clinical Decision Support panel HL7.VMR-CDS** | 3 | 185.08 | 74028-2 |
| **802** | **PhenX - personal medical history of allergies, infectious diseases, and immunizations - child protocol 161601** | 3 | 185.08 | 62894-1 |
| **803** | **PhenX domain - Infectious diseases and immunity** | 3 | 185.08 | 62863-6 |
| **804** | **Enterovirus RNA \| XXX \| Microbiology** | 4 | 184.96 | LP377499-1 |
| **805** | **Enterovirus RNA [Presence] in Specimen by NAA with probe detection** | 2 | 184.96 | 29591-5 |
| **806** | **Oxygen [Partial pressure] in Blood** | 2 | 183.12 | 11556-8 |
| **807** | **Band form neutrophils/100 leukocytes in Blood by Manual count** | 2 | 182.82 | 764-1 |
| **808** | **P-R Interval** | 3 | 182.75 | 8625-6 |
| **809** | **P-R Interval \| Heart \| EKG measurements** | 4 | 182.75 | LP406956-5 |
| **810** | **Epstein Barr virus capsid IgM Ab [Units/volume] in Serum by Immunoassay** | 2 | 182.72 | 5159-9 |
| **811** | **Influenza virus A H1 2009 pandemic RNA [Presence] in Specimen by NAA with probe detection** | 2 | 182.25 | 55465-9 |
| **812** | **Influenza virus A H1 2009 pandemic RNA \| XXX \| Microbiology** | 4 | 182.25 | LP378451-1 |
| **813** | **SaO2 \| Blood venous \| Specific hemodynamics** | 3 | 182.13 | LP416069-5 |
| **814** | **Oxygen saturation in Venous blood** | 3 | 182.13 | 2711-0 |
| **815** | **Q-T interval corrected \| Heart \| EKG measurements** | 4 | 182.07 | LP407055-5 |
| **816** | **Q-T interval** | 3 | 182.07 | 8634-8 |
| **817** | **QRS duration \| Heart \| EKG measurements** | 4 | 182.07 | LP407002-7 |
| **818** | **Q-T interval \| Heart \| EKG measurements** | 4 | 182.07 | LP407042-3 |

| | | | | |
|---|---|---|---|---|
| **819** | **QRS duration** | 3 | 182.07 | 8633-0 |
| **820** | **Epstein Barr virus capsid IgG Ab [Units/volume] in Serum by Immunoassay** | 2 | 180.63 | 5157-3 |
| **821** | **Influenza virus A H1 RNA [Presence] in Specimen by NAA with probe detection** | 2 | 180.55 | 49521-8 |
| **822** | **Influenza virus A H1 RNA \| XXX \| Microbiology** | 4 | 180.55 | LP378410-7 |
| **823** | **Lactate [Moles/volume] in Blood** | 2 | 180.13 | 32693-4 |
| **824** | **Microorganisms panel - Urine sediment** | 3 | 179.90 | 58435-9 |
| **825** | **Microorganisms panel \| Urine sediment \| Urinalysis Panels** | 4 | 179.90 | LP402544-3 |
| **826** | **Erythrocytes [#/area] in Urine sediment by Microscopy high power field** | 2 | 179.89 | 13945-1 |
| **827** | **Lactate\|Pt\|Bld** | 2 | 179.78 | LG48935-7 |
| **828** | **Lactate \| Blood \| Chemistry - non-challenge** | 5 | 179.78 | LP385561-8 |
| **829** | **Bordetella parapertussis DNA [Presence] in Specimen by NAA with probe detection** | 2 | 179.76 | 29723-4 |
| **830** | **Bordetella parapertussis DNA \| XXX \| Microbiology** | 4 | 179.76 | LP373969-7 |
| **831** | **Erythrocyte sedimentation rate by Westergren method** | 2 | 179.21 | 4537-7 |
| **832** | **Hyaline casts \| Urine sediment \| Urinalysis** | 3 | 179.08 | LP402442-0 |
| **833** | **FEV1** | 2 | 178.68 | 20150-9 |
| **834** | **Clotting factors** | 3 | 178.13 | LP31623-9 |
| **835** | **Carbon dioxide [Partial pressure] in Blood** | 2 | 177.93 | 11557-6 |
| **836** | **Meningitis+Encephalitis pathogens panel - Cerebral spinal fluid by NAA with non-probe detection** | 3 | 176.83 | 82180-1 |
| **837** | **Globulin\|MCnc\|Pt\|ANYBldSerPl** | 2 | 174.57 | LG1776-6 |
| **838** | **Cells Counted Total [#] in Blood** | 2 | 171.81 | 11282-1 |
| **839** | **Cells Counted Total \| Blood \| Hematology and Cell counts** | 3 | 171.81 | LP393545-1 |
| **840** | **Urinalysis microscopic panel [#/volume] - Urine by Automated count** | 3 | 171.38 | 50554-5 |
| **841** | **Atypical pneumonia pathogens panel - Respiratory specimen by NAA with probe detection** | 3 | 171.35 | 90439-1 |
| **842** | **Atypical pneumonia pathogens panel \| Respiratory specimen \| Microbiology Panels** | 5 | 171.35 | LP379913-9 |
| **843** | **Specimen volume\|Vol\|Urine** | 2 | 171.06 | LG34516-1 |
| **844** | **Glucose\|Pt\|Bld\|180.156 g/mole** | 2 | 170.59 | LG49883-8 |
| **845** | **Specific gravity \| Urine \| Chemistry - non-challenge** | 4 | 170.17 | LP385426-4 |
| **846** | **Specific gravity of Urine** | 2 | 170.17 | 2965-2 |
| **847** | **Mucus [#/area] in Urine sediment by Microscopy low power field** | 2 | 168.53 | 28545-2 |
| **848** | **Erythrocytes [#/volume] in Blood** | 2 | 167.71 | 26453-1 |
| **849** | **Microorganism identified in Specimen by Culture** | 2 | 167.10 | 11475-1 |
| **850** | **Cytomegalovirus IgG Ab [Units/volume] in Serum or Plasma by Immunoassay** | 2 | 166.87 | 5124-3 |
| **851** | **Erythrocytes [#/volume] in Body fluid** | 2 | 165.97 | 26455-6 |

| | | | | |
|---|---|---|---|---|
| **852** | **Erythrocytes\|NCnc\|Pt\|Body fld** | 2 | 165.52 | LG32852-2 |
| **853** | **Erythrocytes \| Body fluid \| Hematology and Cell counts** | 3 | 165.52 | LP392505-6 |
| **854** | **Specific gravity of Urine by Refractometry** | 2 | 164.35 | 5810-7 |
| **855** | **Coxsackievirus B Ab panel [Titer] - Serum by Complement fixation** | 3 | 163.24 | 41485-4 |
| **856** | **Coxsackievirus B Ab panel \| Serum \| Microbiology Panels** | 5 | 163.24 | LP379945-1 |
| **857** | **Erythrocytes [#/area] in Urine sediment by Automated count** | 2 | 163.19 | 46419-8 |
| **858** | **Forced vital capacity [Volume] Respiratory system** | 2 | 163.19 | 19870-5 |
| **859** | **SARS-CoV-2 (COVID-19) IgG Ab [Presence] in Serum or Plasma by Immunoassay** | 2 | 162.25 | 94563-4 |
| **860** | **Procalcitonin [Mass/volume] in Serum or Plasma by Immunoassay** | 2 | 162.02 | 75241-0 |
| **861** | **Glucose [Mass/volume] in Blood** | 2 | 161.80 | 2339-0 |
| **862** | **Glucose \| Blood \| Chemistry - non-challenge** | 3 | 161.49 | LP385526-1 |
| **863** | **Interleukin 2 Receptor Soluble \| Serum or Plasma \| Chemistry - non-challenge** | 5 | 161.19 | LP385027-0 |
| **864** | **Epstein Barr virus capsid IgM Ab [Presence] in Serum by Immunoassay** | 2 | 160.99 | 24115-8 |
| **865** | **Epstein Barr virus capsid IgG Ab [Presence] in Serum by Immunoassay** | 2 | 160.99 | 24114-1 |
| **866** | **FEF 25-75% \| Respiratory system \| Respiratory measures and Ventilator management** | 3 | 160.18 | LP408958-9 |
| **867** | **Left ventricular End-diastolic volume by Imaging** | 3 | 159.28 | 8821-1 |
| **868** | **Cardiopulmonary** | 2 | 159.28 | LP172861-9 |
| **869** | **Left ventricular Ejection fraction** | 2 | 159.28 | 10230-1 |
| **870** | **Ejection fraction \| Heart ventricle - left \| Specific hemodynamics** | 4 | 159.28 | LP416061-2 |
| **871** | **End-diastolic volume \| Heart ventricle - left \| Specific hemodynamics** | 3 | 159.28 | LP416057-0 |
| **872** | **Hyaline casts [#/area] in Urine sediment by Microscopy high power field** | 2 | 157.90 | 46135-0 |
| **873** | **Base deficit \| Blood venous \| Chemistry - non-challenge** | 5 | 157.00 | LP383303-7 |
| **874** | **Base deficit in Venous blood** | 2 | 157.00 | 1924-0 |
| **875** | **Urea nitrogen/Creatinine [Mass Ratio] in Serum or Plasma** | 2 | 156.97 | 3097-3 |
| **876** | **Urea nitrogen/Creatinine \| Serum or Plasma \| Chemistry - non-challenge** | 4 | 156.97 | LP385472-8 |
| **877** | **Urea nitrogen/Creatinine\|MRto\|Pt\|ANYBldSerPl** | 2 | 156.97 | LG12083-8 |
| **878** | **Bordetella parapertussis IS1001 DNA \| Nasopharynx \| Microbiology** | 4 | 156.89 | LP373970-5 |
| **879** | **Bordetella parapertussis IS1001 DNA [Presence] in Nasopharynx by NAA with non-probe detection** | 2 | 156.89 | 87621-9 |

| | | | | |
|---|---|---|---|---|
| **880** | **Bordetella parapertussis IS1001 DNA\|PrThr\|Sys:ANYResp** | 2 | 156.89 | LG50885-9 |
| **881** | **Borrelia** | 2 | 155.77 | LG41633-5 |
| **882** | **Human coronavirus HKU1 RNA\|PrThr\|Sys:ANYResp** | 2 | 154.61 | LG34135-0 |
| **883** | **Human coronavirus HKU1 RNA [Presence] in Nasopharynx by NAA with non-probe detection** | 2 | 154.61 | 82161-1 |
| **884** | **Human coronavirus HKU1 RNA \| Nasopharynx \| Microbiology** | 4 | 154.61 | LP377171-6 |
| **885** | **Streptococcal DNAse B and ASO antibodies panel - Serum** | 3 | 154.35 | 58713-9 |
| **886** | **DNAse B Ab.Streptococcal and Streptolysin O antibodies panel \| Serum \| Microbiology Panels** | 5 | 154.35 | LP379951-9 |
| **887** | **LMW Heparin [Units/volume] in Platelet poor plasma by Chromogenic method** | 2 | 153.52 | 3271-4 |
| **888** | **Erythrocyte distribution width [Entitic volume]** | 2 | 151.76 | 30384-2 |
| **889** | **Anion gap in Blood** | 2 | 151.19 | 41276-7 |
| **890** | **Anion gap \| Blood \| Chemistry - non-challenge** | 4 | 151.19 | LP386537-7 |
| **891** | **Bacteria identified in Urine by Culture** | 2 | 150.96 | 630-4 |
| **892** | **Bacteria identified\|Prid\|Urine** | 2 | 150.96 | LG34664-9 |
| **893** | **Bacteria identified \| Urine \| Microbiology** | 4 | 150.96 | LP373754-3 |
| **894** | **Specific antigens** | 3 | 150.65 | LP57347-4 |
| **895** | **Lactate dehydrogenase [Enzymatic activity/volume] in Specimen** | 2 | 150.40 | 32324-6 |
| **896** | **Lactate dehydrogenase \| XXX \| Chemistry - non-challenge** | 4 | 150.40 | LP383087-6 |
| **897** | **Myelocytes/100 leukocytes in Blood by Manual count** | 2 | 150.00 | 749-2 |
| **898** | **Interleukin 2 Receptor Soluble [Mass/volume] in Serum or Plasma** | 2 | 149.92 | 76039-7 |
| **899** | **Interleukin 2 receptor.soluble\|Pt\|Ser/Plas** | 2 | 149.92 | LG47065-4 |
| **900** | **Poikilocytosis \| Blood \| Hematology and Cell counts** | 3 | 149.14 | LP393417-3 |
| **901** | **Poikilocytosis [Presence] in Blood by Light microscopy** | 2 | 149.14 | 779-9 |
| **902** | **Cytomegalovirus T-cell immunodeficiency panel - Blood by Flow cytometry (FC)** | 3 | 148.75 | 95184-8 |
| **903** | **Cytomegalovirus T-cell immunodeficiency panel \| Blood \| Cellmarker Panels** | 4 | 148.75 | LP419322-5 |
| **904** | **Coxsackievirus A Ab panel [Titer] - Serum by Complement fixation** | 3 | 146.80 | 41484-7 |
| **905** | **Coxsackievirus A Ab panel \| Serum \| Microbiology Panels** | 5 | 146.80 | LP379944-4 |
| **906** | **SARS-CoV-2 (COVID-19) Ab \| Serum or Plasma \| Microbiology** | 4 | 146.46 | LP418684-9 |
| **907** | **Microcytes \| Blood \| Hematology and Cell counts** | 3 | 146.41 | LP393436-3 |
| **908** | **Bartonella henselae and Bartonella quintana IgG and IgM panel \| Serum \| Microbiology Panels** | 5 | 146.35 | LP379884-2 |
| **909** | **Extractable nuclear Ab panel - Serum** | 3 | 145.94 | 43119-7 |
| **910** | **Extractable nuclear Ab panel \| Serum \| Serology Panels** | 4 | 145.94 | LP404104-4 |

| | | | | |
|---|---|---|---|---|
| **911** | **Cytomegalovirus IgM Ab [Units/volume] in Serum or Plasma by Immunoassay** | 2 | 145.63 | 5126-8 |
| **912** | **Vanillylmandelate and Creatinine panel \| Urine \| Chemistry Panels** | 5 | 144.96 | LP387084-9 |
| **913** | **Vanillylmandelate and Creatinine panel - 24 hour Urine** | 3 | 144.96 | 43099-1 |
| **914** | **Antiphospholipid syndrome** | 3 | 144.70 | LP40278-1 |
| **915** | **SARS-CoV-2 (COVID-19) RNA [Presence] in Nasopharynx by NAA with non-probe detection** | 2 | 144.12 | 94565-9 |
| **916** | **Microbiology CNAMTS panel - Blood** | 3 | 143.31 | 88850-3 |
| **917** | **Microbiology CNAMTS panel \| Blood \| Microbiology Panels** | 5 | 143.31 | LP380041-6 |
| **918** | **Retinol binding protein panel\|-\|Urine** | 2 | 142.93 | LG51055-8 |
| **919** | **Epithelial cells.squamous [Presence] in Urine sediment by Light microscopy** | 2 | 142.84 | 12258-0 |
| **920** | **Cystine panel \| Urine \| Chemistry Panels** | 5 | 142.04 | LP386879-3 |
| **921** | **Cystine panel - Urine** | 3 | 142.04 | 43124-7 |
| **922** | **Chloride panel - 24 hour Urine** | 3 | 141.30 | 43128-8 |
| **923** | **Chloride panel \| Urine \| Chemistry Panels** | 5 | 141.30 | LP386858-7 |
| **924** | **SARS-CoV-2 (COVID-19) Ab [Presence] in Serum or Plasma by Immunoassay** | 2 | 141.06 | 94762-2 |
| **925** | **Mycobacterium tuberculosis stimulated gamma interferon and spot count panel \| Blood \| Microbiology Panels** | 5 | 140.64 | LP380066-3 |
| **926** | **Mycobacterium tuberculosis stimulated gamma interferon and spot count panel - Blood** | 3 | 140.64 | 74281-7 |
| **927** | **Mycobacterium tuberculosis stimulated gamma interferon [Interpretation] in Blood Qualitative** | 2 | 140.64 | 71773-6 |
| **928** | **Mycobacterium tuberculosis stimulated gamma interferon \| Blood \| Microbiology** | 3 | 140.64 | LP379748-9 |
| **929** | **Parainfluenza virus 3 RNA [Presence] in Nasopharynx by NAA with non-probe detection** | 2 | 140.61 | 82173-6 |
| **930** | **Nucleated erythrocytes/100 leukocytes [Ratio] in Blood** | 2 | 140.02 | 19048-8 |
| **931** | **Parainfluenza virus 3 RNA\|PrThr\|Sys:ANYResp** | 2 | 139.83 | LG34069-1 |
| **932** | **Parainfluenza virus 3 RNA \| Nasopharynx \| Microbiology** | 4 | 139.83 | LP378794-4 |
| **933** | **Interleukin 5 \| Serum or Plasma \| Chemistry - non-challenge** | 5 | 138.33 | LP385037-9 |
| **934** | **Interleukin 5 [Mass/volume] in Serum or Plasma** | 2 | 138.33 | 33938-2 |
| **935** | **Specimen source [Identifier] of Body fluid** | 2 | 138.03 | 47938-6 |
| **936** | **Bartonella henselae and Bartonella quintana IgG and IgM panel [Titer] - Serum** | 3 | 137.75 | 90251-0 |
| **937** | **Monocytes/100 leukocytes in Blood by Manual count** | 2 | 136.93 | 744-3 |
| **938** | **Heterophile Ab [Presence] in Serum by Latex agglutination** | 2 | 134.43 | 5213-4 |
| **939** | **Dohle body \| Blood \| Hematology and Cell counts** | 4 | 133.32 | LP393453-8 |

| | | | | |
|---|---|---|---|---|
| **940** | **Dohle body [Presence] in Blood by Light microscopy** | 2 | 133.32 | 7792-5 |
| **941** | **Protein electrophoresis panel \| Urine \| Chemistry Panels** | 5 | 132.94 | LP387034-4 |
| **942** | **Protein fractions panel\|-\|Urine** | 2 | 132.94 | LG36112-7 |
| **943** | **Protein electrophoresis panel - Urine** | 3 | 132.94 | 34539-7 |
| **944** | **Immunoelectrophoresis panel\|-\|Urine** | 2 | 132.94 | LG35785-1 |
| **945** | **Immunoelectrophoresis panel \| Urine \| Chemistry Panels** | 5 | 132.94 | LP386958-5 |
| **946** | **Immunoelectrophoresis panel - Urine** | 3 | 132.94 | 29585-7 |
| **947** | **Carbon dioxide, total [Moles/volume] in Venous blood by calculation** | 2 | 132.33 | 48391-7 |
| **948** | **Immunoglobulin panel [Mass/volume] - Serum** | 3 | 131.98 | 34550-4 |
| **949** | **Immunoglobulin panel \| Serum \| Chemistry Panels** | 5 | 131.98 | LP386963-5 |
| **950** | **Bacteria identified in Blood by Aerobe culture** | 2 | 131.54 | 17928-3 |
| **951** | **Neutrophils.segmented\|NCnc\|Pt\|Bld** | 2 | 130.96 | LG32792-0 |
| **952** | **Segmented neutrophils [#/volume] in Blood** | 2 | 130.96 | 30451-9 |
| **953** | **Segmented neutrophils \| Blood \| Hematology and Cell counts** | 3 | 130.96 | LP392658-3 |
| **954** | **Erythrocyte morphology finding [Identifier] in Blood** | 2 | 129.96 | 6742-1 |
| **955** | **Erythrocyte morphology finding \| Blood \| Hematology and Cell counts** | 3 | 129.96 | LP393394-4 |
| **956** | **Nucleated erythrocytes/100 leukocytes [Ratio] in Blood by Automated count** | 2 | 128.13 | 58413-6 |
| **957** | **Rhinovirus RNA \| XXX \| Microbiology** | 4 | 128.09 | LP378971-8 |
| **958** | **Rhinovirus RNA [Presence] in Specimen by NAA with probe detection** | 2 | 128.09 | 7993-9 |
| **959** | **Epstein Barr virus early IgG \| Serum \| Microbiology** | 4 | 126.96 | LP377545-1 |
| **960** | **SARS-CoV-2 (COVID-19) RNA \| Nasopharynx \| Microbiology** | 4 | 126.84 | LP418694-8 |
| **961** | **Microbiology CNAMTS panel - Cerebral spinal fluid** | 3 | 126.82 | 88842-0 |
| **962** | **Microbiology CNAMTS panel \| Cerebral spinal fluid \| Microbiology Panels** | 5 | 126.82 | LP380044-0 |
| **963** | **Hypochromia [Presence] in Blood by Light microscopy** | 2 | 126.63 | 728-6 |
| **964** | **Hypochromia \| Blood \| Hematology and Cell counts** | 4 | 126.63 | LP393445-4 |
| **965** | **SaO2 \| Blood venous \| Chemistry - non-challenge** | 3 | 126.52 | LP383406-8 |
| **966** | **Single allergens** | 4 | 126.00 | LP30729-5 |
| **967** | **Occupational allergens** | 3 | 126.00 | LP30689-1 |
| **968** | **Sapovirus genogroups I+II+IV+V RNA [Presence] in Stool by NAA with non-probe detection** | 2 | 124.67 | 82213-0 |
| **969** | **Sapovirus genogroups I+II+IV+V RNA \| Stool \| Microbiology** | 4 | 124.67 | LP379037-7 |
| **970** | **Oxygen saturation in Blood** | 3 | 124.14 | 20564-1 |
| **971** | **SaO2 \| Blood \| Specific hemodynamics** | 3 | 124.14 | LP416065-3 |
| **972** | **Pathology** | 2 | 123.03 | LP7839-6 |
| **973** | **P wave axis** | 3 | 122.74 | 8626-4 |
| **974** | **P wave axis \| Heart \| EKG measurements** | 4 | 122.74 | LP406934-2 |

| | | | | |
|---|---|---|---|---|
| **975** | **Glucose\|Pt\|Urine\|180.156 g/mole** | 2 | 122.57 | LG49894-5 |
| **976** | **Glucose [Mass/volume] in Urine** | 2 | 122.57 | 2350-7 |
| **977** | **T wave axis** | 3 | 120.97 | 8638-9 |
| **978** | **Q-T interval corrected** | 3 | 120.97 | 8636-3 |
| **979** | **T wave axis \| Heart \| EKG measurements** | 4 | 120.97 | LP407295-7 |
| **980** | **QRS axis \| Heart \| EKG measurements** | 4 | 120.97 | LP406998-7 |
| **981** | **QRS axis** | 3 | 120.97 | 8632-2 |
| **982** | **Ovalocytes \| Blood \| Hematology and Cell counts** | 3 | 120.95 | LP393409-0 |
| **983** | **Ovalocytes [Presence] in Blood by Light microscopy** | 2 | 120.95 | 774-0 |
| **984** | **Epstein Barr virus early IgG Ab [Units/volume] in Serum** | 2 | 120.79 | 24007-7 |
| **985** | **Glucose\|PrThr\|Urine** | 2 | 120.24 | LG35248-0 |
| **986** | **Nuclear Ab \| Serum \| Serology - non-micro** | 3 | 120.01 | LP403792-7 |
| **987** | **Amylase \| Serum or Plasma \| Chemistry - non-challenge** | 4 | 119.87 | LP382784-9 |
| **988** | **Amylase\|CCnc\|Pt\|ANYBldSerPl** | 2 | 119.87 | LG5805-9 |
| **989** | **Amylase isoenzyme 7 panel \| Serum \| Chemistry Panels** | 5 | 119.87 | LP386813-2 |
| **990** | **Amylase [Enzymatic activity/volume] in Serum or Plasma** | 2 | 119.87 | 1798-8 |
| **991** | **Amylase isoenzyme 7 panel - Serum** | 3 | 119.87 | 24334-5 |
| **992** | **Amylase isoenzyme 3 panel - Serum or Plasma** | 3 | 119.49 | 24333-7 |
| **993** | **Amylase isoenzyme 3 panel \| Serum or Plasma \| Chemistry Panels** | 5 | 119.49 | LP386812-4 |
| **994** | **Triglyceride\|Pt\|Ser/Plas** | 2 | 117.59 | LG47044-9 |
| **995** | **Triglyceride \| Serum or Plasma \| Chemistry - non-challenge** | 3 | 117.59 | LP382509-0 |
| **996** | **Vancomycin^trough\|Pt\|Ser/Plas** | 2 | 116.81 | LG50477-5 |
| **997** | **Vancomycin [Mass/volume] in Serum or Plasma – trough** | 2 | 116.81 | 4092-3 |
| **998** | **Erythrocytes\|NCnc\|Urine** | 2 | 115.75 | LG35160-7 |
| **999** | **Iron and Iron binding capacity panel - Serum or Plasma** | 3 | 115.58 | 50190-8 |
| **1,000** | **Iron and Iron binding capacity panel \| Serum or Plasma \| Chemistry Panels** | 5 | 115.58 | LP386967-6 |
| **1,001** | **Bicarbonate \| Serum or Plasma \| Chemistry - non-challenge** | 4 | 115.35 | LP386569-0 |
| **1,002** | **Bicarbonate [Moles/volume] in Serum or Plasma** | 2 | 115.35 | 1963-8 |
| **1,003** | **Human coronavirus NL63 RNA\|PrThr\|Sys:ANYResp** | 2 | 113.07 | LG34136-8 |
| **1,004** | **Human coronavirus 229E RNA\|PrThr\|Sys:ANYResp** | 2 | 113.07 | LG34137-6 |
| **1,005** | **Human coronavirus OC43 RNA \| Nasopharynx \| Microbiology** | 4 | 113.07 | LP377186-4 |
| **1,006** | **Human coronavirus 229E RNA \| Nasopharynx \| Microbiology** | 4 | 113.07 | LP377163-3 |
| **1,007** | **Human coronavirus NL63 RNA [Presence] in Nasopharynx by NAA with non-probe detection** | 2 | 113.07 | 82162-9 |

| | | | | |
|---|---|---|---|---|
| **1,008** | **Human coronavirus NL63 RNA \| Nasopharynx \| Microbiology** | 4 | 113.07 | LP377178-1 |
| **1,009** | **Human coronavirus OC43 RNA\|PrThr\|Sys:ANYResp** | 2 | 113.07 | LG34138-4 |
| **1,010** | **Human coronavirus 229E RNA [Presence] in Nasopharynx by NAA with non-probe detection** | 2 | 113.07 | 82163-7 |
| **1,011** | **Human coronavirus OC43 RNA [Presence] in Nasopharynx by NAA with non-probe detection** | 2 | 113.07 | 82164-5 |
| **1,012** | **Influenza virus A H1 2009 pandemic RNA \| Nasopharynx \| Microbiology** | 4 | 112.62 | LP378449-5 |
| **1,013** | **Influenza virus A H1 2009 pandemic RNA [Presence] in Nasopharynx by NAA with non-probe detection** | 2 | 112.62 | 82168-6 |
| **1,014** | **Influenza virus A H1 2009 pandemic RNA\|PrThr\|Sys:ANYResp** | 2 | 112.62 | LG34094-9 |
| **1,015** | **PhenX - urinary microalbumin assay for kidney function protocol 141501** | 3 | 112.50 | 62809-9 |
| **1,016** | **PhenX - fasting C-peptide assay for residual beta cell function protocol 141201** | 3 | 112.50 | 62803-2 |
| **1,017** | **PhenX - urinary creatinine assay for kidney function protocol 141601** | 3 | 112.50 | 62811-5 |
| **1,018** | **PhenX - serum creatinine assay for kidney function protocol 141401** | 3 | 112.50 | 62807-3 |
| **1,019** | **PhenX - assay for syphilis protocol 160701** | 3 | 112.50 | 62877-6 |
| **1,020** | **PhenX - liver function assay protocol 190801** | 3 | 112.50 | 62967-5 |
| **1,021** | **PhenX - colorectal procedures and outcomes protocol 190401** | 3 | 112.50 | 62959-2 |
| **1,022** | **PhenX domain - Gastrointestinal** | 3 | 112.50 | 62949-3 |
| **1,023** | **PhenX - personal and family history of psoriasis protocol 170501** | 3 | 112.50 | 62906-3 |
| **1,024** | **PhenX - assay for human leukocyte antigen (HLA) genotyping protocol 160601** | 3 | 112.50 | 62875-0 |
| **1,025** | **PhenX - assay herpes simplex virus types 1 - 2 protocol 160501** | 3 | 112.50 | 62873-5 |
| **1,026** | **PhenX - human immunodeficiency virus (HIV) protocol 160901** | 3 | 112.50 | 62882-6 |
| **1,027** | **PhenX - assay for hepatitis B protocol 160401** | 3 | 112.50 | 62871-9 |
| **1,028** | **Process variables for assays PhenX** | 3 | 112.50 | 69863-9 |
| **1,029** | **PhenX - assay for cytokine panel 12 protocol 160201** | 3 | 112.50 | 62867-7 |
| **1,030** | **PhenX - physical functioning - objective protocol 150501** | 3 | 112.50 | 62825-5 |
| **1,031** | **Service comment** | 2 | 112.50 | 8251-1 |
| **1,032** | **PhenX - assay for hepatitis C protocol 160301** | 3 | 112.50 | 62869-3 |
| **1,033** | **PhenX - celiac sprue assay protocol 190301** | 3 | 112.50 | 62957-6 |
| **1,034** | **PhenX - assay for chlamydia - gonorrhea protocol 160101** | 3 | 112.50 | 62865-1 |
| **1,035** | **PhenX - oral glucose tolerance test protocol 141001** | 3 | 112.50 | 62856-0 |
| **1,036** | **PhenX - fasting serum insulin protocol 141301** | 3 | 112.50 | 62805-7 |

| | | | | |
|---|---|---|---|---|
| **1,037** | **PhenX - respiratory - respiratory rate - child protocol 091402** | 3 | 112.50 | 62633-3 |
| **1,038** | **Process variables for assays - human leukocyte antigen - HLA PhenX** | 3 | 112.50 | 69866-2 |
| **1,039** | **PhenX domain - Diabetes** | 3 | 112.50 | 62787-7 |
| **1,040** | **Process variables for assays - gonorrhea-chlamydia PhenX** | 3 | 112.50 | 69865-4 |
| **1,041** | **PhenX - alkaline phosphatase protocol 170101** | 3 | 112.50 | 62898-2 |
| **1,042** | **PhenX - signs of essential tremor protocol 131301** | 3 | 112.50 | 62784-4 |
| **1,043** | **PhenX - fracture history protocol 170901** | 3 | 112.50 | 64390-8 |
| **1,044** | **PhenX - fasting plasma glucose for diabetes screening - blood draw protocol 140801** | 3 | 112.50 | 62851-1 |
| **1,045** | **PhenX domain - Neurology** | 3 | 112.50 | 62755-4 |
| **1,046** | **PhenX - personal medical history of allergies, infectious diseases, and immunizations - adult protocol 161602** | 3 | 112.50 | 62895-8 |
| **1,047** | **PhenX - glycosylated hemoglobin assay reflecting long-term glucose concentration protocol 140901** | 3 | 112.50 | 62854-5 |
| **1,048** | **Influenza virus A H1 RNA [Presence] in Nasopharynx by NAA with non-probe detection** | 2 | 112.46 | 82167-8 |
| **1,049** | **Influenza virus A H1 RNA\|PrThr\|Sys:ANYResp** | 2 | 112.16 | LG34092-3 |
| **1,050** | **Influenza virus A H1 RNA \| Nasopharynx \| Microbiology** | 4 | 112.16 | LP378408-1 |
| **1,051** | **Influenza virus A H3 RNA [Presence] in Nasopharynx by NAA with non-probe detection** | 2 | 112.01 | 82169-4 |
| **1,052** | **Influenza virus A H1 2009 pandemic RNA panel \| XXX \| Microbiology Panels** | 5 | 111.81 | LP380029-1 |
| **1,053** | **Influenza virus A H1 2009 pandemic RNA panel - Specimen by NAA with probe detection** | 3 | 111.81 | 55466-7 |
| **1,054** | **PhenX - respiratory - exercise capacity - 6 minute walk test protocol 090601** | 3 | 111.38 | 62619-2 |
| **1,055** | **PhenX domain - Physical activity and physical fitness** | 3 | 111.38 | 62812-3 |
| **1,056** | **PhenX - respiratory - sleep apnea - child protocol 091502** | 3 | 111.38 | 62637-4 |
| **1,057** | **Parainfluenza virus 2 RNA [Presence] in Nasopharynx by NAA with non-probe detection** | 2 | 111.26 | 82172-8 |
| **1,058** | **Influenza virus A H3 RNA\|PrThr\|Sys:ANYResp** | 2 | 110.99 | LG34093-1 |
| **1,059** | **Influenza virus A H3 RNA \| Nasopharynx \| Microbiology** | 4 | 110.99 | LP378418-0 |
| **1,060** | **Bacteria [#/area] in Urine sediment by Microscopy high power field** | 2 | 110.69 | 5769-5 |
| **1,061** | **Exercise/Activity** | 2 | 110.27 | LG41761-4 |
| **1,062** | **Parainfluenza virus 2 RNA \| Nasopharynx \| Microbiology** | 4 | 110.22 | LP378774-6 |
| **1,063** | **Parainfluenza virus 2 RNA\|PrThr\|Sys:ANYResp** | 2 | 110.22 | LG34068-3 |
| **1,064** | **Lymphocytes [#/volume] in Blood by Manual count** | 2 | 110.18 | 732-8 |

| | | | | |
|---|---|---|---|---|
| **1,065** | **Glycerol and glycerol-corrected triglyceride panel \| Serum or Plasma \| Chemistry Panels** | 5 | 109.51 | LP419297-9 |
| **1,066** | **Triglyceride\|MCnc\|Pt\|ANYBldSerPl** | 2 | 109.51 | LG9600-0 |
| **1,067** | **Triglyceride [Mass/volume] in Serum or Plasma** | 2 | 109.51 | 2571-8 |
| **1,068** | **Glycerol and glycerol-corrected triglyceride panel - Serum or Plasma** | 3 | 109.51 | 94859-6 |
| **1,069** | **Reticulocytes panel - Blood** | 3 | 108.98 | 50262-5 |
| **1,070** | **Reticulocytes panel \| Blood \| Hematology and Cell Count Panels** | 4 | 108.98 | LP393937-0 |
| **1,071** | **Microcytes [Presence] in Blood by Light microscopy** | 2 | 108.38 | 741-9 |
| **1,072** | **Epstein Barr virus capsid IgG and IgM panel - Serum** | 3 | 108.10 | 24316-2 |
| **1,073** | **Complement C3 \| Serum or Plasma \| Hematology and Cell counts** | 3 | 107.88 | LP393725-9 |
| **1,074** | **Complement C3 [Mass/volume] in Serum or Plasma** | 2 | 107.88 | 4485-9 |
| **1,075** | **Controlled substances and drugs of abuse** | 3 | 107.29 | LP31448-1 |
| **1,076** | **Free T4 and TSH panel - Serum or Plasma** | 3 | 107.02 | 24348-5 |
| **1,077** | **Free T4 and TSH panel \| Serum or Plasma \| Chemistry Panels** | 5 | 107.02 | LP386925-4 |
| **1,078** | **Monocytes [#/volume] in Blood by Manual count** | 2 | 106.48 | 743-5 |
| **1,079** | **Thyroid Hormones-** | 4 | 106.40 | LP31666-8 |
| **1,080** | **Microscopic observation [Identifier] in Specimen by Gram stain** | 2 | 105.90 | 664-3 |
| **1,081** | **Dyshemoglobin** | 5 | 105.81 | LP31772-4 |
| **1,082** | **PhenX domain - Skin, bone, muscle and joint** | 3 | 105.14 | 62896-6 |
| **1,083** | **Rhinovirus+Enterovirus RNA \| Nasopharynx \| Microbiology** | 4 | 105.01 | LP377507-1 |
| **1,084** | **Rhinovirus+Enterovirus RNA\|PrThr\|Sys:ANYResp** | 2 | 104.65 | LG34139-2 |
| **1,085** | **pH^^adjusted to patient's actual temperature\|LsCnc\|Pt\|ANYBldSerPl** | 2 | 104.28 | LG13263-5 |
| **1,086** | **Influenza virus B RNA \| XXX \| Microbiology** | 4 | 103.97 | LP378559-1 |
| **1,087** | **Lipase [Enzymatic activity/volume] in Serum or Plasma** | 2 | 103.83 | 3040-3 |
| **1,088** | **Triacylglycerol lipase \| Serum or Plasma \| Chemistry - non-challenge** | 4 | 103.83 | LP383262-5 |
| **1,089** | **Differential panel \| Body fluid \| Hematology and Cell Count Panels** | 4 | 103.20 | LP393907-3 |
| **1,090** | **Differential panel - Body fluid** | 3 | 103.20 | 29580-8 |
| **1,091** | **Oxygen saturation Calculated from oxygen partial pressure in Venous blood** | 2 | 102.43 | 51731-8 |
| **1,092** | **Influenza virus B RNA [Presence] in Specimen by NAA with probe detection** | 2 | 102.14 | 40982-1 |
| **1,093** | **Eosinophils [#/volume] in Blood by Manual count** | 2 | 101.70 | 712-0 |
| **1,094** | **Lactate [Mass/volume] in Serum or Plasma** | 2 | 101.64 | 14118-4 |
| **1,095** | **17-Ketosteroids and 17-Ketogenic steroids panel \| Urine \| Chemistry Panels** | 5 | 101.34 | LP386790-2 |
| **1,096** | **Adrenal cancer risk assessment and urine steroid fractions panel** | 3 | 101.34 | 95556-7 |

| | | | | |
|---|---|---|---|---|
| **1,097** | **Specimen set** | 2 | 101.34 | PANEL.SPEC |
| **1,098** | **Urine collection associated observations panel \| Urine \| Specimen panels** | 2 | 101.34 | LP404336-2 |
| **1,099** | **Urine collection associated observations panel - 24 hour Urine** | 3 | 101.34 | 81323-8 |
| **1,100** | **Microalbumin panel - 24 hour Urine** | 3 | 101.34 | 58431-8 |
| **1,101** | **Microalbumin panel \| Urine \| Chemistry Panels** | 5 | 101.34 | LP386990-8 |
| **1,102** | **17-Ketosteroids and 17-Ketogenic steroids panel - 24 hour Urine** | 3 | 101.34 | 43135-3 |
| **1,103** | **Lactate\|MCnc\|Pt\|ANYBldSerPl** | 2 | 101.22 | LG2275-8 |
| **1,104** | **Microscopic observation \| XXX \| Microbiology** | 3 | 101.22 | LP379843-8 |
| **1,105** | **Retinol binding protein panel - 24 hour Urine** | 3 | 101.04 | 96399-1 |
| **1,106** | **Uranium.depleted panel - 24 hour Urine** | 3 | 101.04 | 49670-3 |
| **1,107** | **Uranium.depleted panel \| Urine \| Drug and Toxicology Panels** | 4 | 101.04 | LP392132-9 |
| **1,108** | **Neutrophil cytoplasmic Ab panel \| Serum \| Serology Panels** | 4 | 100.82 | LP404117-6 |
| **1,109** | **Neutrophil cytoplasmic Ab panel - Serum by Immunofluorescence** | 3 | 100.82 | 87427-1 |
| **1,110** | **Parainfluenza virus 1 RNA\|PrThr\|Sys:ANYResp** | 2 | 100.37 | LG34067-5 |
| **1,111** | **Parainfluenza virus 1 RNA \| Nasopharynx \| Microbiology** | 4 | 100.37 | LP378754-8 |
| **1,112** | **Microalbumin/Creatinine ratio panel \| Urine \| Chemistry Panels** | 5 | 99.46 | LP386991-6 |
| **1,113** | **Porphyrin fractions panel \| Urine \| Chemistry Panels** | 5 | 99.46 | LP387021-1 |
| **1,114** | **Porphyrin fractions panel - 24 hour Urine** | 3 | 99.46 | 43116-3 |
| **1,115** | **Microalbumin/Creatinine ratio panel\|-\|Urine** | 2 | 99.46 | LG36111-9 |
| **1,116** | **Microalbumin/Creatinine panel in 24H Urine** | 3 | 99.46 | 58447-4 |
| **1,117** | **Porphyrin fractions panel\|-\|Urine** | 2 | 99.46 | LG50937-8 |
| **1,118** | **Cortisol.free panel - 24 hour Urine** | 3 | 99.31 | 43126-2 |
| **1,119** | **Cortisol.free panel \| Urine \| Chemistry Panels** | 5 | 99.31 | LP386866-0 |
| **1,120** | **Bordetella pertussis.pertussis toxin promoter region [Presence] in Nasopharynx by NAA with non-probe detection** | 2 | 99.20 | 82179-3 |
| **1,121** | **Mycoplasma pneumoniae DNA\|PrThr\|Sys:ANYResp** | 2 | 99.20 | LG34140-0 |
| **1,122** | **Mycoplasma pneumoniae DNA [Presence] in Nasopharynx by NAA with non-probe detection** | 2 | 99.20 | 82177-7 |
| **1,123** | **Chlamydophila pneumoniae DNA [Presence] in Nasopharynx by NAA with non-probe detection** | 2 | 99.20 | 82178-5 |
| **1,124** | **Chlamydophila pneumoniae DNA \| Nasopharynx \| Microbiology** | 4 | 99.20 | LP374544-7 |
| **1,125** | **Mycoplasma pneumoniae DNA \| Nasopharynx \| Microbiology** | 4 | 99.20 | LP375331-8 |
| **1,126** | **Influenza virus A RNA [Presence] in Nasopharynx by NAA with non-probe detection** | 2 | 99.20 | 82166-0 |
| **1,127** | **Influenza virus B RNA [Presence] in Nasopharynx by NAA with non-probe detection** | 2 | 99.08 | 82170-2 |

| | | | | |
|---|---|---|---|---|
| **1,128** | **Rhinovirus+Enterovirus RNA [Presence] in Nasopharynx by NAA with non-probe detection** | 2 | 99.08 | 82175-1 |
| **1,129** | **Parainfluenza virus 4 RNA [Presence] in Nasopharynx by NAA with non-probe detection** | 2 | 99.08 | 82174-4 |
| **1,130** | **Parainfluenza virus 1 RNA [Presence] in Nasopharynx by NAA with non-probe detection** | 2 | 99.08 | 82171-0 |
| **1,131** | **Human metapneumovirus RNA [Presence] in Nasopharynx by NAA with non-probe detection** | 2 | 98.95 | 82165-2 |
| **1,132** | **Parainfluenza virus 4 RNA\|PrThr\|Sys:ANYResp** | 2 | 98.82 | LG34070-9 |
| **1,133** | **Parainfluenza virus 4 RNA \| Nasopharynx \| Microbiology** | 4 | 98.82 | LP378805-8 |
| **1,134** | **Bordetella pertussis.pertussis toxin promoter region\|PrThr\|Sys:ANYResp** | 2 | 98.82 | LG34124-4 |
| **1,135** | **Bordetella pertussis toxin** | 4 | 98.82 | LP14047-2 |
| **1,136** | **Bordetella pertussis.pertussis toxin promoter region \| Nasopharynx \| Microbiology** | 5 | 98.82 | LP374002-6 |
| **1,137** | **Human metapneumovirus RNA \| Nasopharynx \| Microbiology** | 4 | 98.70 | LP378237-4 |
| **1,138** | **Human metapneumovirus RNA\|PrThr\|Sys:ANYResp** | 2 | 98.70 | LG34090-7 |
| **1,139** | **Chlamydophila pneumoniae DNA\|PrThr\|Sys:ANYResp** | 2 | 98.70 | LG34076-6 |
| **1,140** | **Glucose [Presence] in Urine** | 2 | 98.55 | 2349-9 |
| **1,141** | **Globulin [Mass/volume] in Plasma** | 2 | 98.19 | 13536-8 |
| **1,142** | **Globulin \| Plasma \| Chemistry - non-challenge** | 4 | 98.19 | LP384593-2 |
| **1,143** | **Protein [Units/volume] in Urine** | 2 | 98.12 | 27298-9 |
| **1,144** | **Epithelial cells.renal [Presence] in Urine sediment by Light microscopy** | 2 | 97.92 | 12248-1 |
| **1,145** | **Lactate [Moles/volume] in Serum or Plasma** | 2 | 97.44 | 2524-7 |
| **1,146** | **Respiratory syncytial virus RNA [Presence] in Nasopharynx by NAA with non-probe detection** | 2 | 96.45 | 82176-9 |
| **1,147** | **Epithelial cells.renal\|Urine/Urine sed** | 2 | 96.33 | LG40860-5 |
| **1,148** | **Epithelial cells.renal \| Urine sediment \| Urinalysis** | 3 | 96.33 | LP402470-1 |
| **1,149** | **Immunoglobulin light chains panel \| Urine \| Chemistry Panels** | 5 | 96.03 | LP386961-9 |
| **1,150** | **Immunoglobulin light chains panel - 24 hour Urine** | 3 | 96.03 | 43110-6 |
| **1,151** | **Immunoglobulin light chains panel\|-\|Urine** | 2 | 96.03 | LG36305-7 |
| **1,152** | **Volume of 24 hour Urine** | 2 | 96.03 | 3167-4 |
| **1,153** | **Clostridium sp** | 4 | 96.00 | LP30497-9 |
| **1,154** | **Polymorphonuclear cells/100 leukocytes \| Blood \| Hematology and Cell counts** | 3 | 95.47 | LP393172-4 |
| **1,155** | **Borrelia burgdorferi IgG+IgM with reflex to immune blot panel - Serum** | 3 | 95.47 | 62341-3 |
| **1,156** | **Borrelia burgdorferi IgG+IgM with reflex to immune blot panel \| Serum \| Microbiology Panels** | 5 | 95.47 | LP379906-3 |
| **1,157** | **Eosinophils/100 leukocytes in Blood by Manual count** | 2 | 95.38 | 714-6 |
| **1,158** | **Basophils/100 leukocytes in Blood by Manual count** | 2 | 95.38 | 707-0 |
| **1,159** | **Carbon dioxide \| Blood arterial \| Chemistry - non-challenge** | 3 | 95.30 | LP383322-7 |

| | | | | |
|---|---|---|---|---|
| **1,160** | **Neutrophils [#/volume] in Blood by Manual count** | 2 | 95.23 | 753-4 |
| **1,161** | **Lymphocytes/100 leukocytes in Blood by Manual count** | 2 | 95.23 | 737-7 |
| **1,162** | **Basophils [#/volume] in Blood by Manual count** | 2 | 95.08 | 705-4 |
| **1,163** | **Erythrocytes [#/volume] in Urine** | 2 | 94.76 | 30391-7 |
| **1,164** | **Epstein Barr virus DNA \| XXX \| Microbiology** | 4 | 94.30 | LP377537-8 |
| **1,165** | **Neutrophils/100 leukocytes in Blood by Manual count** | 2 | 93.87 | 23761-0 |
| **1,166** | **Drugs of abuse panel \| Urine \| Drug and Toxicology Panels** | 4 | 93.81 | LP392070-1 |
| **1,167** | **Drugs of abuse panel - Urine by Screen method** | 3 | 93.81 | 69739-1 |
| **1,168** | **Epstein Barr virus Ab panel - Serum** | 3 | 93.78 | 87554-2 |
| **1,169** | **Epstein Barr virus Ab panel \| Serum \| Microbiology Panels** | 5 | 93.78 | LP379960-0 |
| **1,170** | **Leukocytes [#/volume] in Urine by Test strip** | 2 | 93.49 | 20408-1 |
| **1,171** | **Giardia lamblia** | 2 | 92.38 | LG41616-0 |
| **1,172** | **Viruses, non-human pathogens** | 4 | 92.37 | LP30610-7 |
| **1,173** | **Cryptosporidium** | 2 | 92.31 | LG41646-7 |
| **1,174** | **Complement C4\|Pt\|Ser/Plas** | 2 | 91.95 | LG43109-4 |
| **1,175** | **Complement C4 [Mass/volume] in Serum or Plasma** | 2 | 91.95 | 4498-2 |
| **1,176** | **Complement C4 \| Serum or Plasma \| Hematology and Cell counts** | 3 | 91.95 | LP393745-7 |
| **1,177** | **Microscopic observation \| Urine sediment \| Urinalysis** | 3 | 91.70 | LP402540-1 |
| **1,178** | **Microscopic observation [Identifier] in Urine sediment by Light microscopy** | 2 | 91.70 | 12235-8 |
| **1,179** | **Microscopic observation\|Urine/Urine sed** | 2 | 91.70 | LG40859-7 |
| **1,180** | **Retrograde ejaculation evaluation panel - Urine** | 3 | 90.88 | 98412-0 |
| **1,181** | **Fertility Panels** | 3 | 90.88 | LP36844-6 |
| **1,182** | **Retrograde ejaculation evaluation panel \| Urine \| Fertility Panels** | 4 | 90.88 | LP428682-1 |
| **1,183** | **Fertility testing order set** | 2 | 90.81 | PANEL.FERT |
| **1,184** | **Urate \| Serum or Plasma \| Chemistry - non-challenge** | 5 | 90.56 | LP383583-4 |
| **1,185** | **Urate\|Pt\|Ser/Plas\|168.112 g/mole** | 2 | 90.56 | LG49755-8 |
| **1,186** | **Urate\|MCnc\|Pt\|ANYBldSerPl** | 2 | 90.56 | LG51310-7 |
| **1,187** | **Urate [Mass/volume] in Serum or Plasma** | 2 | 90.56 | 3084-1 |
| **1,188** | **DNA double strand Ab \| Serum \| Serology - non-micro** | 3 | 90.18 | LP402693-8 |
| **1,189** | **Calcium.ionized [Moles/volume] in Blood by Ion-selective membrane electrode (ISE)** | 2 | 89.72 | 47596-2 |
| **1,190** | **Human parechovirus** | 4 | 89.39 | LP66495-0 |
| **1,191** | **Alanine aminotransferase [Enzymatic activity/volume] in Serum or Plasma by With P-5’-P** | 2 | 89.05 | 1743-4 |
| **1,192** | **Aspartate aminotransferase [Enzymatic activity/volume] in Serum or Plasma by With P-5’-P** | 2 | 88.20 | 30239-8 |
| **1,193** | **Home drug screening panel - Urine** | 3 | 88.06 | 55419-6 |
| **1,194** | **Home drug screening panel \| Urine \| Drug and Toxicology Panels** | 4 | 88.06 | LP392087-5 |
| **1,195** | **Chloride \| Blood \| Chemistry - non-challenge** | 4 | 87.94 | LP386574-0 |

| | | | | |
|---|---|---|---|---|
| **1,196** | **Chloride [Moles/volume] in Blood** | 2 | 87.94 | 2069-3 |
| **1,197** | **Gastrointestinal pathogens panel \| Stool \| Microbiology Panels** | 5 | 87.38 | LP379971-7 |
| **1,198** | **Cyclospora** | 2 | 87.09 | LG41642-6 |
| **1,199** | **Urobilinogen\|PrThr\|Urine** | 2 | 87.09 | LG34517-9 |
| **1,200** | **CD16+CD56+ cells/100 cells in Blood** | 2 | 86.92 | 18267-5 |
| **1,201** | **CD16+CD56+ cells [#/volume] in Blood** | 2 | 86.92 | 20402-4 |
| **1,202** | **CD16+CD56+ cells \| Blood \| Cell markers** | 4 | 86.92 | LP400507-2 |
| **1,203** | **Cells.CD16+CD56+/100 cells \| Blood \| Cell markers** | 4 | 86.92 | LP400509-8 |
| **1,204** | **Plesiomonas sp** | 4 | 86.91 | LP150159-4 |
| **1,205** | **Vancomycin [Mass/volume] in Serum or Plasma** | 2 | 86.77 | 20578-1 |
| **1,206** | **Vancomycin\|Pt\|Ser/Plas** | 2 | 86.77 | LG47183-5 |
| **1,207** | **FEF 25-75% Predicted** | 2 | 86.75 | 69971-0 |
| **1,208** | **PhenX - respiratory - bronchodilator responsiveness - BDR protocol 090301** | 3 | 86.34 | 62615-0 |
| **1,209** | **Forced vital capacity [Volume] Respiratory system – post bronchodilation** | 2 | 86.26 | 19875-4 |
| **1,210** | **Target cells [Presence] in Blood by Light microscopy** | 2 | 86.13 | 10381-2 |
| **1,211** | **Target cells \| Blood \| Hematology and Cell counts** | 3 | 86.13 | LP393431-4 |
| **1,212** | **Respiratory pathogens panel - Specimen by Organism specific culture** | 3 | 86.02 | 55101-0 |
| **1,213** | **Parainfluenza virus 2 \| XXX \| Microbiology** | 4 | 86.02 | LP378760-5 |
| **1,214** | **Respiratory pathogens panel \| XXX \| Microbiology Panels** | 5 | 86.02 | LP380089-5 |
| **1,215** | **Parainfluenza virus 2 [Presence] in Specimen by Organism specific culture** | 2 | 86.02 | 55098-8 |
| **1,216** | **Vibrio cholerae** | 2 | 84.55 | LG41625-1 |
| **1,217** | **Thyroxine and Thyroxine.free panel - Serum or Plasma** | 3 | 84.22 | 90224-7 |
| **1,218** | **Thyroxine and Thyroxine.free panel \| Serum or Plasma \| Chemistry Panels** | 5 | 84.22 | LP387074-0 |
| **1,219** | **Epstein Barr virus DNA [Log #/volume] (viral load) in Specimen by NAA with probe detection** | 2 | 83.91 | 53774-6 |
| **1,220** | **Platelet clump [Presence] in Blood by Light microscopy** | 2 | 83.80 | 7796-6 |
| **1,221** | **Volume of Urine** | 2 | 83.43 | 28009-9 |
| **1,222** | **Cells.CD3/100 cells \| Blood \| Cell markers** | 4 | 83.38 | LP399367-4 |
| **1,223** | **CD3 cells/100 cells in Blood** | 2 | 83.38 | 8124-0 |
| **1,224** | **Platelet clump \| Blood \| Hematology and Cell counts** | 3 | 83.13 | LP393380-3 |
| **1,225** | **Urinary cell count CNAMTS panel - 3 hour Urine** | 3 | 83.04 | 88846-1 |
| **1,226** | **Urinary cell count CNAMTS panel \| Urine \| Microbiology Panels** | 5 | 83.04 | LP380123-2 |
| **1,227** | **Protein and creatinine panel \| Urine \| Chemistry Panels** | 5 | 82.60 | LP387024-5 |
| **1,228** | **Protein and creatinine panel - Urine** | 3 | 82.60 | 87434-7 |
| **1,229** | **Metamyelocytes\|NCnc\|Pt\|Bld** | 2 | 81.85 | LG32786-2 |

| | | | | |
|---|---|---|---|---|
| **1,230** | **Metamyelocytes \| Blood \| Hematology and Cell counts** | 3 | 81.85 | LP392823-3 |
| **1,231** | **Carbon dioxide^^adjusted to patient's actual temperature\|PPres\|Pt\|ANYBldSerPl** | 2 | 81.74 | LG13382-3 |
| **1,232** | **Albumin [Mass/volume] in Serum or Plasma by Bromocresol purple (BCP) dye binding method** | 2 | 81.39 | 61152-5 |
| **1,233** | **Albumin\|Pt\|Ser/Plas\|BCP\|2754.105 g/mole** | 2 | 81.39 | LG33735-8 |
| **1,234** | **Macrocytes \| Blood \| Hematology and Cell counts** | 3 | 80.93 | LP393434-8 |
| **1,235** | **Macrocytes [Presence] in Blood by Light microscopy** | 2 | 80.93 | 738-5 |
| **1,236** | **Lipid panel with direct LDL - Serum or Plasma** | 3 | 80.78 | 57698-3 |
| **1,237** | **Lipid panel with direct LDL \| Serum or Plasma \| Chemistry Panels** | 5 | 80.78 | LP386974-2 |
| **1,238** | **Amino acids panel \| Serum or Plasma \| Chemistry Panels** | 5 | 80.60 | LP386808-2 |
| **1,239** | **Amino acids panel [Moles/volume] - Serum or Plasma** | 3 | 80.60 | 35083-5 |
| **1,240** | **Nucelotides/sides and derivatives** | 4 | 80.59 | LP31405-1 |
| **1,241** | **HLA antigens** | 2 | 79.41 | LP7806-5 |
| **1,242** | **Plesiomonas shigelloides DNA [Presence] in Stool by NAA with probe detection** | 2 | 79.18 | 70296-9 |
| **1,243** | **Abnormal Hemoglobin** | 3 | 79.06 | LP30925-9 |
| **1,244** | **CD3-CD16+CD56+ (Natural killer) cells/100 cells in Blood** | 2 | 78.96 | 8112-5 |
| **1,245** | **Cells.CD3-CD16+CD56+/100 cells \| Blood \| Cell markers** | 4 | 78.96 | LP400314-3 |
| **1,246** | **Stomatocytes \| Blood \| Hematology and Cell counts** | 3 | 78.85 | LP393430-6 |
| **1,247** | **Stomatocytes [Presence] in Blood by Light microscopy** | 2 | 78.85 | 10380-4 |
| **1,248** | **Gastrointestinal pathogens panel - Stool by NAA with probe detection** | 3 | 78.79 | 79381-0 |
| **1,249** | **Gastrointestinal bacterial pathogens panel \| Stool \| Microbiology Panels** | 5 | 78.79 | LP379968-3 |
| **1,250** | **Gastrointestinal bacterial pathogens panel - Stool by NAA with probe detection** | 3 | 78.79 | 92695-6 |
| **1,251** | **Polymorphonuclear cells [#/volume] in Blood** | 2 | 78.30 | 35003-3 |
| **1,252** | **Polymorphonuclear cells \| Blood \| Hematology and Cell counts** | 3 | 78.30 | LP393160-9 |
| **1,253** | **Polymorphonuclear cells/100 leukocytes in Blood** | 2 | 78.30 | 34999-3 |
| **1,254** | **Plesiomonas shigelloides DNA \| Stool \| Microbiology** | 5 | 78.26 | LP375551-1 |
| **1,255** | **Cells.CD3+CD4+/100 cells \| Blood \| Cell markers** | 4 | 77.42 | LP400179-0 |
| **1,256** | **CD3+CD4+ (T4 helper) cells/100 cells in Blood** | 2 | 77.42 | 8123-2 |
| **1,257** | **Lipid 1996 panel \| Serum or Plasma \| Chemistry Panels** | 5 | 77.29 | LP386973-4 |
| **1,258** | **Lipid 1996 panel - Serum or Plasma** | 3 | 77.29 | 24331-1 |
| **1,259** | **Globulin [Mass/volume] in Serum** | 2 | 77.14 | 2336-6 |
| **1,260** | **Reticulocytes/100 erythrocytes in Blood by Automated count** | 2 | 76.23 | 17849-1 |
| **1,261** | **Reticulocytes/100 erythrocytes \| Blood \| Hematology and Cell counts** | 3 | 76.13 | LP392556-9 |

| | | | | |
|---|---|---|---|---|
| **1,262** | **pH of Venous blood adjusted to patient's actual temperature** | 2 | 75.89 | 39486-6 |
| **1,263** | **Dacrocytes \| Blood \| Hematology and Cell counts** | 3 | 75.85 | LP393406-6 |
| **1,264** | **Dacrocytes [Presence] in Blood by Light microscopy** | 2 | 75.85 | 7791-7 |
| **1,265** | **CD3-CD16+CD56+ (Natural killer) cells \| Blood \| Cell markers** | 4 | 75.76 | LP400310-1 |
| **1,266** | **CD3-CD16+CD56+ (Natural killer) cells [#/volume] in Blood** | 2 | 75.76 | 9728-7 |
| **1,267** | **CD3 cells \| Blood \| Cell markers** | 4 | 75.64 | LP399355-9 |
| **1,268** | **CD3 cells [#/volume] in Blood** | 2 | 75.64 | 8122-4 |
| **1,269** | **CD3+CD4+ (T4 helper) cells [#/volume] in Blood** | 2 | 75.58 | 24467-3 |
| **1,270** | **CD3+CD4+ (T4 helper) cells \| Blood \| Cell markers** | 4 | 75.58 | LP400174-1 |
| **1,271** | **Cells.CD3+CD4+/Cells.CD3+CD8+ \| Blood \| Cell markers** | 4 | 74.50 | LP400186-5 |
| **1,272** | **CD3+CD4+ (T4 helper) cells/CD3+CD8+ (T8 suppressor cells) cells [# Ratio] in Blood** | 2 | 74.50 | 54218-3 |
| **1,273** | **Cells.CD3+CD8+/100 cells \| Blood \| Cell markers** | 4 | 74.48 | LP400217-8 |
| **1,274** | **CD3+CD8+ (T8 suppressor) cells/100 cells in Blood** | 2 | 74.48 | 8101-8 |
| **1,275** | **Lipoprofile panel \| Serum or Plasma \| Chemistry Panels** | 5 | 74.44 | LP386975-9 |
| **1,276** | **Lipoprofile panel - Serum or Plasma** | 3 | 74.44 | 59062-0 |
| **1,277** | **Service comment 14** | 2 | 74.43 | 8256-0 |
| **1,278** | **Thyrotropin \| Serum or Plasma \| Chemistry - non-challenge** | 3 | 74.16 | LP381405-2 |
| **1,279** | **Pituitary Hormones-** | 4 | 73.57 | LP31660-1 |
| **1,280** | **CD3+CD8+ (T8 suppressor) cells [#/volume] in Blood** | 2 | 73.36 | 14135-8 |
| **1,281** | **CD3+CD8+ (T8 suppressor) cells \| Blood \| Cell markers** | 4 | 73.36 | LP400212-9 |
| **1,282** | **Bicarbonate [Moles/volume] in Blood** | 2 | 73.33 | 1959-6 |
| **1,283** | **Bicarbonate \| Blood \| Chemistry - non-challenge** | 4 | 73.33 | LP386552-6 |
| **1,284** | **Urobilinogen\|Pt\|Urine\|Test strip\|592.737 g/mole** | 2 | 72.94 | LG47120-7 |
| **1,285** | **Urobilinogen [Mass/volume] in Urine by Test strip** | 2 | 72.94 | 20405-7 |
| **1,286** | **Schistocytes [Presence] in Blood by Light microscopy** | 2 | 72.03 | 800-3 |
| **1,287** | **Organic acids panel - Serum or Plasma** | 3 | 71.98 | 35866-3 |
| **1,288** | **Organic acids panel \| Serum or Plasma \| Chemistry Panels** | 5 | 71.98 | LP387008-8 |
| **1,289** | **Schistocytes \| Blood \| Hematology and Cell counts** | 3 | 71.92 | LP393421-5 |
| **1,290** | **Platelets.reticulated/100 platelets \| Blood \| Hematology and Cell counts** | 3 | 71.82 | LP393238-3 |
| **1,291** | **Platelets reticulated/100 platelets in Blood by Automated count** | 2 | 71.82 | 71693-6 |
| **1,292** | **Borrelia burgdorferi IgG and IgM bands panel - Serum by Immunoblot** | 3 | 71.37 | 98204-1 |
| **1,293** | **Borrelia burgdorferi IgG and IgM bands panel \| Serum \| Microbiology Panels** | 5 | 71.37 | LP428623-5 |
| **1,294** | **Bacteria [Presence] in Urine sediment by Light microscopy** | 2 | 71.04 | 25145-4 |

| | | | | |
|---|---|---|---|---|
| **1,295** | **Lipoprotein metabolism panel \| Serum or Plasma \| Chemistry Panels** | 5 | 70.93 | LP386976-7 |
| **1,296** | **Lipoprotein metabolism panel - Serum or Plasma** | 3 | 70.93 | 91136-2 |
| **1,297** | **SaO2 \| Blood arterial \| Chemistry - non-challenge** | 3 | 69.14 | LP383400-1 |
| **1,298** | **Urobilinogen\|ACnc\|Urine** | 2 | 68.94 | LG35109-4 |
| **1,299** | **Urobilinogen [Units/volume] in Urine by Test strip** | 2 | 68.94 | 19161-9 |
| **1,300** | **Leukocytes [#/volume] in Body fluid** | 2 | 68.81 | 26466-3 |
| **1,301** | **Bilirubin.total [Presence] in Urine by Automated test strip** | 2 | 68.70 | 50551-1 |
| **1,302** | **Protein and Glucose panel [Mass/volume] - Cerebral spinal fluid** | 3 | 68.49 | 34546-2 |
| **1,303** | **Protein and Glucose panel \| Cerebral spinal fluid \| Chemistry Panels** | 5 | 68.49 | LP387026-0 |
| **1,304** | **Nuclear Ab\|Titr\|Pt\|ANYBldSerPl** | 2 | 68.44 | LG7004-7 |
| **1,305** | **FEV1 –post bronchodilation** | 2 | 68.44 | 20155-8 |
| **1,306** | **Leukocytes\|NCnc\|Pt\|Body fld** | 2 | 68.42 | LG32859-7 |
| **1,307** | **Leukocytes \| Body fluid \| Hematology and Cell counts** | 3 | 68.42 | LP392602-1 |
| **1,308** | **HLA Panels** | 3 | 67.99 | LP36817-2 |
| **1,309** | **Shigella** | 2 | 67.85 | LG41635-0 |
| **1,310** | **Shigella sp DNA [Presence] in Specimen by NAA with probe detection** | 2 | 67.85 | 46455-2 |
| **1,311** | **Shigella sp DNA \| XXX \| Microbiology** | 4 | 67.85 | LP375772-3 |
| **1,312** | **Referral lab name [Identifier]** | 2 | 66.64 | 42216-2 |
| **1,313** | **Base deficit in Blood** | 2 | 66.13 | 30318-0 |
| **1,314** | **Base deficit \| Blood \| Chemistry - non-challenge** | 5 | 66.13 | LP383296-3 |
| **1,315** | **Bacteria identified in Body fluid by Culture** | 2 | 66.10 | 611-4 |
| **1,316** | **Bacteria identified \| Body fluid \| Microbiology** | 4 | 65.82 | LP373702-2 |
| **1,317** | **Body mass index (BMI) [Percentile] Per age and sex** | 3 | 65.67 | 59576-9 |
| **1,318** | **Body weight molecular** | 2 | 65.67 | BDYWGT.MOLEC |
| **1,319** | **Glucose in serum - glucose in synovial fluid [Molar concentration difference]** | 2 | 65.21 | 72648-9 |
| **1,320** | **Referral lab test reference range** | 2 | 65.07 | 19147-8 |
| **1,321** | **Referral lab test results** | 2 | 64.82 | 19146-0 |
| **1,322** | **Nitrite\|MCnc\|Urine** | 2 | 64.61 | LG35158-1 |
| **1,323** | **Nitrite [Mass/volume] in Urine by Test strip** | 2 | 64.61 | 20407-3 |
| **1,324** | **ERBB2 gene (HER2) duplication associated observations panel \| Tissue and Smears \| Molecular Pathology Panels** | 4 | 64.38 | LP401920-6 |
| **1,325** | **ERBB2 gene (HER2) duplication associated observations panel - Tissue by FISH** | 3 | 64.38 | 74885-5 |
| **1,326** | **Celiac disease IgA and HLA typing and serology panel - Blood or Tissue** | 3 | 64.27 | 94493-4 |
| **1,327** | **Celiac disease IgA and HLA typing and serology panel \| Blood or Tissue \| HLA Panels** | 4 | 64.27 | LP419323-3 |
| **1,328** | **Oxygen \| Blood arterial \| Chemistry - non-challenge** | 3 | 64.27 | LP383361-5 |

| | | | | |
|---|---|---|---|---|
| **1,329** | **Carbon dioxide [Partial pressure] adjusted to patient's actual temperature in Venous blood** | 2 | 64.16 | 40619-9 |
| **1,330** | **Eosinophils/100 leukocytes in Cerebral spinal fluid** | 2 | 64.06 | 26451-5 |
| **1,331** | **Eosinophils/100 leukocytes \| Cerebral spinal fluid \| Hematology and Cell counts** | 3 | 64.06 | LP392802-7 |
| **1,332** | **Promyelocytes/100 leukocytes \| Blood \| Hematology and Cell counts** | 3 | 63.88 | LP392861-3 |
| **1,333** | **Cytomegalovirus DNA \| XXX \| Microbiology** | 4 | 63.80 | LP377286-2 |
| **1,334** | **Mucus \| Urine \| Urinalysis** | 3 | 63.50 | LP402519-5 |
| **1,335** | **Mucus [#/volume] in Urine by Automated count** | 2 | 63.50 | 51478-6 |
| **1,336** | **Influenza virus** | 2 | 63.28 | LG32757-3 |
| **1,337** | **Antibiotic Susceptibility Panels** | 4 | 63.15 | LP29682-9 |
| **1,338** | **Susceptibility order sets** | 2 | 63.15 | PANEL.ABXBACT |
| **1,339** | **Thyrotropin [Units/volume] in Serum or Plasma** | 2 | 62.95 | 3016-3 |
| **1,340** | **Thyrotropin\|ACnc\|Pt\|ANYBldSerPl** | 2 | 62.95 | LG11757-8 |
| **1,341** | **ABO and Rh group \| Blood \| Blood bank** | 3 | 62.54 | LP399109-0 |
| **1,342** | **Specific gravity of Urine by Automated test strip** | 2 | 62.36 | 53326-5 |
| **1,343** | **Hepatitis B virus** | 2 | 62.12 | LG32752-4 |
| **1,344** | **Q-T interval corrected based on Bazett formula** | 3 | 62.04 | 76635-2 |
| **1,345** | **R-R interval by EKG** | 3 | 62.04 | 8637-1 |
| **1,346** | **R-R interval \| Heart \| EKG measurements** | 4 | 62.04 | LP407109-0 |
| **1,347** | **Drugs of abuse 7 and Alcohol and Tricyclics panel \| Urine \| Drug and Toxicology Panels** | 4 | 62.00 | LP392064-4 |
| **1,348** | **Drugs of abuse 7 and Alcohol and Tricyclics panel - Urine by Screen method** | 3 | 62.00 | 51782-1 |
| **1,349** | **Erythrocyte morphology [Interpretation] in Urine sediment by Light microscopy Narrative** | 2 | 61.93 | 53974-2 |
| **1,350** | **Erythrocyte morphology \| Urine sediment \| Urinalysis** | 3 | 61.93 | LP402480-0 |
| **1,351** | **Chronic urticaria index panel - Serum or Plasma** | 3 | 61.51 | 69040-4 |
| **1,352** | **pH \| Blood arterial \| Chemistry - non-challenge** | 4 | 61.44 | LP383432-4 |
| **1,353** | **Platelets Large [Presence] in Blood by Light microscopy** | 2 | 60.23 | 32146-3 |
| **1,354** | **Platelets Large \| Blood \| Hematology and Cell counts** | 3 | 60.01 | LP393231-8 |
| **1,355** | **Glucose\|Pt\|Urine\|Test strip.automated\|180.156 g/mole** | 2 | 59.56 | LG33611-1 |
| **1,356** | **pH of Urine by Automated test strip** | 2 | 59.56 | 50560-2 |
| **1,357** | **Salmonella sp DNA [Presence] in Specimen by NAA with probe detection** | 2 | 59.32 | 49612-5 |
| **1,358** | **Salmonella sp DNA \| XXX \| Microbiology** | 4 | 59.32 | LP375702-0 |
| **1,359** | **Respiratory syncytial virus RNA \| Nasopharynx \| Microbiology** | 4 | 59.01 | LP378956-9 |
| **1,360** | **Bilirubin.total [Mass/volume] in Urine by Test strip** | 2 | 58.61 | 20505-4 |
| **1,361** | **Bilirubin\|Pt\|Urine\|Test strip\|584.673 g/mole** | 2 | 58.61 | LG33179-9 |
| **1,362** | **Bilirubin\|MCnc\|Urine** | 2 | 58.61 | LG35154-0 |
| **1,363** | **Fatty acid comprehensive (C8-C26) panel - Serum or Plasma** | 3 | 57.92 | 43674-1 |

| | | | | |
|---|---|---|---|---|
| **1,364** | **Fatty acid comprehensive (C8-C26) panel \| Serum or Plasma \| Chemistry Panels** | 5 | 57.92 | LP386912-2 |
| **1,365** | **Gas panel \| Blood capillary \| Chemistry Panels** | 5 | 57.88 | LP386936-1 |
| **1,366** | **Gas panel - Capillary blood** | 3 | 57.88 | 24337-8 |
| **1,367** | **SARS-CoV-2 (COVID-19) IgG Ab [Units/volume] in Serum or Plasma by Immunoassay** | 2 | 56.60 | 94505-5 |
| **1,368** | **Molecular pathology** | 2 | 56.33 | LP7822-2 |
| **1,369** | **Leukocytes [#/volume] in Urine by Automated count** | 2 | 56.02 | 51487-7 |
| **1,370** | **Protein and Glucose panel - Urine by Test strip** | 3 | 56.00 | 45060-1 |
| **1,371** | **Protein and Glucose panel \| Urine \| Chemistry Panels** | 5 | 56.00 | LP387027-8 |
| **1,372** | **Sapovirus RNA [Presence] in Specimen by NAA with probe detection** | 2 | 54.95 | 72111-8 |
| **1,373** | **Astrovirus RNA [Presence] in Specimen by NAA with probe detection** | 2 | 54.95 | 69938-9 |
| **1,374** | **Sapovirus RNA \| XXX \| Microbiology** | 4 | 54.95 | LP379036-9 |
| **1,375** | **Astrovirus RNA \| XXX \| Microbiology** | 4 | 54.95 | LP377101-3 |
| **1,376** | **Acute hepatitis 2000 panel - Serum** | 3 | 54.95 | 24363-4 |
| **1,377** | **Acute hepatitis 2000 panel \| Serum \| Microbiology Panels** | 5 | 54.95 | LP379865-1 |
| **1,378** | **Molecular Pathology Panels** | 3 | 54.66 | LP62072-1 |
| **1,379** | **Carbon dioxide, total [Moles/volume] in Venous blood** | 2 | 54.55 | 2027-1 |
| **1,380** | **Molecular pathology order set** | 2 | 54.25 | PANEL.MOLPATH |
| **1,381** | **Gas and Carbon monoxide panel - Capillary blood** | 3 | 54.01 | 24342-8 |
| **1,382** | **Gas and Carbon monoxide panel \| Blood capillary \| Chemistry Panels** | 5 | 54.01 | LP386931-2 |
| **1,383** | **Oxygen [Partial pressure] in Arterial blood** | 2 | 53.56 | 2703-7 |
| **1,384** | **Drugs of abuse screen W Reflex confirm panel \| Urine \| Drug and Toxicology Panels** | 4 | 53.42 | LP392071-9 |
| **1,385** | **Drugs of abuse screen W Reflex confirm panel - Urine** | 3 | 53.42 | 87428-9 |
| **1,386** | **Vanillylmandelate, Homovanillate and Creatinine panel \| Urine \| Chemistry Panels** | 5 | 53.05 | LP387086-4 |
| **1,387** | **Vanillylmandelate, Homovanillate and Creatinine panel - 24 hour Urine** | 3 | 53.05 | 48644-9 |
| **1,388** | **Specimen Physical properties, artifacts** | 4 | 52.55 | LP31410-1 |
| **1,389** | **Vancomycin resistant enterococcus [Presence] in Specimen by Organism specific culture** | 2 | 52.20 | 13316-5 |
| **1,390** | **Vancomycin resistant enterococcus \| XXX \| Microbiology** | 4 | 52.20 | LP374761-7 |
| **1,391** | **ABO and Rh group [Type] in Blood** | 2 | 52.11 | 882-1 |
| **1,392** | **Bicarbonate \| Blood arterial \| Chemistry - non-challenge** | 4 | 51.69 | LP386553-4 |
| **1,393** | **Bicarbonate [Moles/volume] in Arterial blood** | 2 | 51.69 | 1960-4 |
| **1,394** | **Fatty acid essential (C12-C22) panel - Serum or Plasma** | 3 | 51.58 | 43676-6 |
| **1,395** | **Fatty acid essential (C12-C22) panel \| Serum or Plasma \| Chemistry Panels** | 5 | 51.58 | LP386913-0 |

| | | | | |
|---|---|---|---|---|
| **1,396** | **Glucose [Mass/volume] in Urine by Automated test strip** | 2 | 50.89 | 53328-1 |
| **1,397** | **Ketones [Mass/volume] in Urine by Automated test strip** | 2 | 50.89 | 50557-8 |
| **1,398** | **Ketones\|Pt\|Urine\|Test strip.automated\|270.413 g/mole** | 2 | 50.89 | LG48912-6 |
| **1,399** | **Microorganism identification and resistance pattern determination panel \| Isolate or Specimen \| Antibiotic Susceptibility Panels** | 5 | 50.74 | LP380959-9 |
| **1,400** | **Microorganism identification and resistance pattern determination panel by Molecular genetics method** | 3 | 50.74 | 92254-2 |
| **1,401** | **Escherichia coli DNA [Presence] in Specimen by NAA with probe detection** | 2 | 50.67 | 61398-4 |
| **1,402** | **Escherichia coli DNA \| XXX \| Microbiology** | 5 | 50.67 | LP374822-7 |
| **1,403** | **Blood pathogens panel \| Positive blood culture \| Microbiology Panels** | 5 | 50.56 | LP379915-4 |
| **1,404** | **Blood pathogens panel by NAA with non-probe detection in Positive blood culture** | 3 | 50.56 | 85762-3 |
| **1,405** | **Manganese panel \| Urine \| Drug and Toxicology Panels** | 4 | 50.40 | LP392093-3 |
| **1,406** | **Manganese panel - Urine** | 3 | 50.40 | 58736-0 |
| **1,407** | **Chromium panel - Urine** | 3 | 50.40 | 53780-3 |
| **1,408** | **Chromium panel \| Urine \| Drug and Toxicology Panels** | 4 | 50.40 | LP392045-3 |
| **1,409** | **Erythrocytes [#/volume] in Urine by Automated test strip** | 2 | 50.18 | 57747-8 |
| **1,410** | **Urobilinogen [Presence] in Urine by Automated test strip** | 2 | 50.18 | 62487-4 |
| **1,411** | **Nuclear Ab\|PrThr\|Pt\|ANYBldSerPl** | 2 | 49.68 | LG4652-6 |
| **1,412** | **Neutrophils.band form/100 leukocytes \| Body fluid \| Hematology and Cell counts** | 3 | 49.52 | LP392683-1 |
| **1,413** | **Band form neutrophils/100 leukocytes in Body fluid** | 2 | 49.52 | 26510-8 |
| **1,414** | **Cyclospora cayetanensis DNA [Presence] in Specimen by NAA with probe detection** | 2 | 49.10 | 41436-7 |
| **1,415** | **Cyclospora cayetanenis DNA \| XXX \| Microbiology** | 4 | 49.10 | LP376750-8 |
| **1,416** | **Yersinia sp DNA \| XXX \| Microbiology** | 4 | 49.10 | LP376123-8 |
| **1,417** | **Cryptosporidium sp DNA \| XXX \| Microbiology** | 4 | 49.10 | LP376746-6 |
| **1,418** | **Cryptosporidium sp DNA [Presence] in Specimen by NAA with probe detection** | 2 | 49.10 | 60545-1 |
| **1,419** | **Yersinia sp DNA [Identifier] in Specimen by NAA with probe detection** | 2 | 49.10 | 48646-4 |
| **1,420** | **Vibrio cholerae DNA \| XXX \| Microbiology** | 4 | 48.93 | LP376072-7 |
| **1,421** | **Vibrio cholerae DNA [Presence] in Specimen by NAA with probe detection** | 2 | 48.93 | 61371-1 |
| **1,422** | **Vibrio sp DNA \| XXX \| Microbiology** | 4 | 48.93 | LP376115-4 |
| **1,423** | **Escherichia coli Stx1 and Stx2 toxin stx1 and stx2 and H7 flagellar fliC genes [Identifier] in Specimen by NAA with probe detection** | 2 | 48.93 | 53947-8 |

| | | | | |
|---|---|---|---|---|
| **1,424** | **Campylobacter sp DNA.diarrheagenic \| Stool \| Microbiology** | 4 | 48.93 | LP374301-2 |
| **1,425** | **Campylobacter sp DNA.diarrheagenic [Presence] in Stool by NAA with probe detection** | 2 | 48.93 | 71429-5 |
| **1,426** | **Vibrio sp DNA [Identifier] in Specimen by NAA with probe detection** | 2 | 48.93 | 49609-1 |
| **1,427** | **Escherichia coli Stx1 and Stx2 toxin stx1 and stx2 and H7 flagellar fliC genes \| XXX \| Microbiology** | 5 | 48.93 | LP374782-3 |
| **1,428** | **Hemoglobin [Mass/volume] in Blood by calculation** | 2 | 48.89 | 20509-6 |
| **1,429** | **IgA \| Serum or Plasma \| Chemistry - non-challenge** | 4 | 48.86 | LP384793-8 |
| **1,430** | **IgA, IgA1 and IgA2 panel - Serum or Plasma** | 3 | 48.86 | 87552-6 |
| **1,431** | **IgA [Mass/volume] in Serum or Plasma** | 2 | 48.86 | 2458-8 |
| **1,432** | **IgA\|MCnc\|Pt\|ANYBldSerPl** | 2 | 48.86 | LG9788-3 |
| **1,433** | **IgA\|Pt\|Ser/Plas** | 2 | 48.86 | LG46917-7 |
| **1,434** | **IgA, IgA1 and IgA2 panel \| Serum or Plasma \| Chemistry Panels** | 5 | 48.86 | LP386954-4 |
| **1,435** | **Homovanillate panel \| Urine \| Chemistry Panels** | 5 | 48.82 | LP386950-2 |
| **1,436** | **Homovanillate panel - 24 hour Urine** | 3 | 48.82 | 44906-6 |
| **1,437** | **Giardia lamblia DNA \| XXX \| Microbiology** | 4 | 48.60 | LP376777-1 |
| **1,438** | **Giardia lamblia DNA [Presence] in Specimen by NAA with probe detection** | 2 | 48.60 | 60544-4 |
| **1,439** | **HIV 1 and 2 tests - Meaningful Use set** | 3 | 47.41 | 75622-1 |
| **1,440** | **HIV 1 and 2 tests - Meaningful Use set \| Patient \| Microbiology Panels** | 5 | 47.41 | LP380010-1 |
| **1,441** | **IgM [Mass/volume] in Serum or Plasma** | 2 | 47.07 | 2472-9 |
| **1,442** | **IgM\|Pt\|Ser/Plas** | 2 | 47.07 | LG48822-7 |
| **1,443** | **IgM \| Serum or Plasma \| Chemistry - non-challenge** | 4 | 47.07 | LP384862-1 |
| **1,444** | **IgM\|MCnc\|Pt\|ANYBldSerPl** | 2 | 47.07 | LG9802-2 |
| **1,445** | **Adrenal Cortex** | 4 | 46.93 | LP31653-6 |
| **1,446** | **25-Hydroxyvitamin D3+25-Hydroxyvitamin D2/24,25-dihydroxyvitamin D3+24,25-dihydroxyvitamin D2 ratio panel \| Serum or Plasma \| Chemistry Panels** | 5 | 46.77 | LP419292-0 |
| **1,447** | **Calcidiol and Calciferol panel \| Serum or Plasma \| Chemistry Panels** | 5 | 46.77 | LP386840-5 |
| **1,448** | **25-Hydroxyvitamin D3+25-Hydroxyvitamin D2/24,25-dihydroxyvitamin D3+24,25-dihydroxyvitamin D2 ratio panel in Serum or Plasma** | 3 | 46.77 | 94674-9 |
| **1,449** | **Calcidiol and Calciferol panel - Serum or Plasma** | 3 | 46.77 | 49590-3 |
| **1,450** | **Glucose \| Blood capillary \| Chemistry - non-challenge** | 3 | 45.91 | LP385528-7 |
| **1,451** | **Calcium.ionized\|MCnc\|Pt\|ANYBldSerPl** | 2 | 45.74 | LG7250-6 |
| **1,452** | **Blood bank order set** | 2 | 45.72 | PANEL.BLDBK |
| **1,453** | **Creatinine [Mass/volume] in Urine** | 2 | 45.36 | 2161-8 |
| **1,454** | **Creatinine\|MCnc\|Pt\|ANYUrine** | 2 | 45.36 | LG7133-4 |
| **1,455** | **Retinol binding protein panel - Urine** | 3 | 45.36 | 96401-5 |
| **1,456** | **Bacteria [#/area] in Urine sediment by Automated count** | 2 | 45.21 | 33218-9 |

| | | | | |
|---|---|---|---|---|
| **1,457** | **Creatinine\|Pt\|Urine\|113.12 g/mole** | 2 | 45.18 | LG50025-2 |
| **1,458** | **Creatinine\|MCnc\|Urine** | 2 | 45.18 | LG35227-4 |
| **1,459** | **Metanephrine, Normetanephrine, 3-Methoxytyramine & Creatinine panel\|-\|Urine** | 2 | 45.09 | LG37123-3 |
| **1,460** | **Metanephrine, Normetanephrine & Creatinine panel\|-\|Urine** | 2 | 45.09 | LG37118-3 |
| **1,461** | **Metanephrine, Normetanephrine, 3-Methoxytyramine and Creatinine panel \| Urine \| Drug and Toxicology Panels** | 4 | 45.09 | LP392101-4 |
| **1,462** | **Metanephrine, Normetanephrine, 3-Methoxytyramine and Creatinine panel - Urine** | 3 | 45.09 | 72670-3 |
| **1,463** | **Metanephrine, Normetanephrine and Creatinine panel - Urine** | 3 | 45.09 | 68317-7 |
| **1,464** | **Metanephrine, Normetanephrine and Creatinine panel \| Urine \| Drug and Toxicology Panels** | 4 | 45.09 | LP392100-6 |
| **1,465** | **Glucose [Presence] in Urine by Test strip** | 2 | 44.87 | 25428-4 |
| **1,466** | **5-Hydroxyindoleacetate panel - 24 hour Urine** | 3 | 44.78 | 44907-4 |
| **1,467** | **5-Hydroxyindoleacetate panel \| Urine \| Chemistry Panels** | 5 | 44.78 | LP386793-6 |
| **1,468** | **Base excess \| Blood arterial \| Chemistry - non-challenge** | 5 | 44.51 | LP383305-2 |
| **1,469** | **Creatinine \| Urine \| Chemistry - non-challenge** | 4 | 44.34 | LP385363-9 |
| **1,470** | **Thyroxine (T4) free [Mass/volume] in Serum or Plasma** | 2 | 44.26 | 3024-7 |
| **1,471** | **Thyroxine.free\|Pt\|Ser/Plas** | 2 | 44.26 | LG49672-5 |
| **1,472** | **Thyroxine.free\|MCnc\|Pt\|ANYBldSerPl** | 2 | 44.15 | LG12004-4 |
| **1,473** | **Thyroxine free \| Serum or Plasma \| Chemistry - non-challenge** | 3 | 44.15 | LP381519-0 |
| **1,474** | **Glucose\|Pt\|BldC\|Glucometer\|180.156 g/mole** | 2 | 44.08 | LG33190-6 |
| **1,475** | **Glucose [Mass/volume] in Capillary blood by Glucometer** | 2 | 44.08 | 41653-7 |
| **1,476** | **IgG and IgG subclass panel [Mass/volume] - Serum** | 3 | 43.07 | 47289-4 |
| **1,477** | **IgG and IgG subclass panel \| Serum \| Chemistry Panels** | 5 | 43.07 | LP386955-1 |
| **1,478** | **Vitamin B complex** | 3 | 42.64 | LP31703-9 |
| **1,479** | **HIV 1+2 Ab+HIV1 p24 Ag [Presence] in Serum or Plasma by Immunoassay** | 2 | 42.34 | 56888-1 |
| **1,480** | **HIV 1+2 Ab+HIV1 p24 Ag \| Serum or Plasma \| Microbiology** | 4 | 42.34 | LP378012-1 |
| **1,481** | **HIV 1+2 Ab and HIV1 p24 Ag panel \| Serum or Plasma \| Microbiology Panels** | 5 | 42.34 | LP380012-7 |
| **1,482** | **HIV 1+2 Ab and HIV1 p24 Ag panel - Serum or Plasma by Immunoassay** | 3 | 42.34 | 83101-6 |
| **1,483** | **HIV 1 and 2 Ab and HIV 1 p24 Ag panel \| Serum or Plasma \| Microbiology Panels** | 5 | 42.13 | LP380008-5 |
| **1,484** | **HIV 1 and 2 Ab and HIV 1 p24 Ag panel - Serum or Plasma by Immunoassay** | 3 | 42.13 | 85037-0 |

| | | | | |
|---|---|---|---|---|
| **1,485** | **Protein\|PrThr\|Urine** | 2 | 41.99 | LG35161-5 |
| **1,486** | **Nitrate [Presence] in Urine** | 2 | 41.97 | 32030-9 |
| **1,487** | **Nitrate \| Urine \| Drug toxicology** | 4 | 41.97 | LP387531-9 |
| **1,488** | **Protein [Mass/volume] in Urine by Automated test strip** | 2 | 41.97 | 50561-0 |
| **1,489** | **Leukocytes [#/volume] in Urine by Automated test strip** | 2 | 41.97 | 58805-3 |
| **1,490** | **Iron \| Serum or Plasma \| Chemistry - non-challenge** | 4 | 41.89 | LP385074-2 |
| **1,491** | **Iron [Mass/volume] in Serum or Plasma** | 2 | 41.89 | 2498-4 |
| **1,492** | **Iron\|Pt\|Ser/Plas\|55.845 g/mole** | 2 | 41.89 | LG48872-2 |
| **1,493** | **Blood group antibody screen \| Serum or Plasma \| Blood bank** | 3 | 41.72 | LP398980-5 |
| **1,494** | **Thyroxine binding globulin panel [Mass/volume] - Serum or Plasma by Electrophoresis** | 3 | 41.56 | 48073-1 |
| **1,495** | **Thyroxine (T4) [Mass/volume] in Serum or Plasma** | 2 | 41.56 | 3026-2 |
| **1,496** | **Thyroxine\|Pt\|Ser/Plas\|776.874 g/mole** | 2 | 41.56 | LG46307-1 |
| **1,497** | **Thyroxine binding globulin panel \| Serum or Plasma \| Chemistry Panels** | 5 | 41.56 | LP387075-7 |
| **1,498** | **Thyroxine \| Serum or Plasma \| Chemistry - non-challenge** | 3 | 41.56 | LP381510-9 |
| **1,499** | **Thyroxine\|MCnc\|Pt\|ANYBldSerPl** | 2 | 41.56 | LG12009-3 |
| **1,500** | **SaO2 \| Blood \| Chemistry - non-challenge** | 3 | 41.52 | LP383399-5 |
| **1,501** | **Miscellaneous toxic small organic and inorganic molecules** | 3 | 41.30 | LP31433-3 |
| **1,502** | **Electrolytes 3 panel - Urine** | 3 | 41.03 | 24329-5 |
| **1,503** | **Oxygen saturation Calculated from oxygen partial pressure in Blood** | 2 | 40.54 | 2713-6 |
| **1,504** | **Iron binding capacity \| Serum or Plasma \| Chemistry - non-challenge** | 4 | 40.13 | LP385085-8 |
| **1,505** | **Iron binding capacity\|Pt\|Ser/Plas** | 2 | 40.13 | LG48875-5 |
| **1,506** | **Epithelial cells.squamous [#/area] in Urine sediment by Microscopy high power field** | 2 | 39.38 | 11277-1 |
| **1,507** | **IgG [Mass/volume] in Serum or Plasma** | 2 | 39.10 | 2465-3 |
| **1,508** | **IgG \| Serum or Plasma \| Chemistry - non-challenge** | 4 | 39.10 | LP384832-4 |
| **1,509** | **IgG\|Pt\|Ser/Plas** | 2 | 39.10 | LG45120-9 |
| **1,510** | **IgG\|MCnc\|Pt\|ANYBldSerPl** | 2 | 39.10 | LG9795-8 |
| **1,511** | **Iron Satn \| Serum or Plasma \| Chemistry - non-challenge** | 4 | 38.74 | LP416972-0 |
| **1,512** | **Iron saturation [Mass Fraction] in Serum or Plasma** | 2 | 38.74 | 2502-3 |
| **1,513** | **Electrolytes 3 panel\|-\|Urine** | 2 | 38.01 | LG35255-5 |
| **1,514** | **Electrolytes 3 panel \| Urine \| Chemistry Panels** | 5 | 38.01 | LP386897-5 |
| **1,515** | **SARS-CoV-2 stimulated gamma interferon panel \| Blood \| Microbiology Panels** | 5 | 37.93 | LP427525-3 |
| **1,516** | **SARS-CoV-2 stimulated gamma interferon panel - Blood** | 3 | 37.93 | 95974-2 |
| **1,517** | **Blood Bank Panels** | 3 | 37.79 | LP31894-6 |

| | | | | |
|---|---|---|---|---|
| **1,518** | **Urobilinogen [Presence] in Urine by Test strip** | 2 | 37.33 | 5818-0 |
| **1,519** | **Urobilinogen\|PrThr\|Pt\|ANYUrine** | 2 | 37.33 | LG1334-4 |
| **1,520** | **Calcidiol and Calciferol and Calcitriol panel [Mass/volume] - Serum or Plasma** | 3 | 37.29 | 43134-6 |
| **1,521** | **Calcidiol and Calciferol and Calcitriol panel \| Serum or Plasma \| Chemistry Panels** | 5 | 37.29 | LP386839-7 |
| **1,522** | **Protein electrophoresis panel \| Cerebral spinal fluid \| Chemistry Panels** | 5 | 37.21 | LP387032-8 |
| **1,523** | **Protein electrophoresis panel - Cerebral spinal fluid** | 3 | 37.21 | 24352-7 |
| **1,524** | **Cobalamins and folate panel \| Serum \| Chemistry Panels** | 5 | 36.30 | LP427532-9 |
| **1,525** | **Cobalamin (Vitamin B12) and folate panel - Serum** | 3 | 36.30 | 96805-7 |
| **1,526** | **Benzodiazepines panel \| Urine \| Drug and Toxicology Panels** | 4 | 35.58 | LP392032-1 |
| **1,527** | **Benzodiazepines panel\|-\|Urine** | 2 | 35.58 | LG50130-0 |
| **1,528** | **Streptococcus pyogenes** | 2 | 35.01 | LG41610-3 |
| **1,529** | **Psychiatric drugs** | 3 | 34.91 | LP31449-9 |
| **1,530** | **Protein [Mass/volume] in Cerebral spinal fluid** | 2 | 34.25 | 2880-3 |
| **1,531** | **Glucose\|Pt\|CSF\|180.156 g/mole** | 2 | 34.25 | LG49888-7 |
| **1,532** | **Glucose [Mass/volume] in Cerebral spinal fluid** | 2 | 34.25 | 2342-4 |
| **1,533** | **Protein and Glucose panel - Cerebral spinal fluid** | 3 | 34.25 | 55394-1 |
| **1,534** | **Protein \| Cerebral spinal fluid \| Chemistry - non-challenge** | 4 | 34.25 | LP384457-0 |
| **1,535** | **Glucose \| Cerebral spinal fluid \| Chemistry - non-challenge** | 3 | 34.25 | LP385532-9 |
| **1,536** | **Respiratory syncytial virus RNA\|PrThr\|Sys:ANYResp** | 2 | 33.89 | LG34071-7 |
| **1,537** | **Reticulocytes \| Blood \| Hematology and Cell counts** | 3 | 33.24 | LP392535-3 |
| **1,538** | **Reticulocytes\|NCnc\|Pt\|Bld** | 2 | 33.24 | LG32882-9 |
| **1,540** | **25-hydroxyvitamin D3 [Mass/volume] in Serum or Plasma** | 2 | 31.08 | 1989-3 |
| **1,541** | **Calcidiol\|MCnc\|Pt\|ANYBldSerPl** | 2 | 31.08 | LG51030-1 |
| **1,542** | **25-hydroxyvitamin D3 \| Serum or Plasma \| Chemistry - non-challenge** | 3 | 31.08 | LP385191-4 |
| **1,543** | **Calcidiol\|Pt\|Ser/Plas\|400.647 g/mole** | 2 | 31.08 | LG43465-0 |
| **1,544** | **Influenza virus A RNA\|PrThr\|Sys:ANYResp** | 2 | 31.01 | LG34064-2 |
| **1,545** | **SARS-CoV-2 (COVID-19) IgM \| Serum or Plasma \| Microbiology** | 4 | 30.14 | LP418691-4 |
| **1,546** | **SARS-CoV-2 (COVID-19) IgM Ab [Presence] in Serum or Plasma by Immunoassay** | 2 | 30.14 | 94564-2 |
| **1,547** | **Influenza virus B RNA\|PrThr\|Sys:ANYResp** | 2 | 29.98 | LG34065-9 |
| **1,548** | **Epithelial cells.squamous \| Urine \| Urinalysis** | 3 | 29.66 | LP402471-9 |
| **1,549** | **Epithelial cells.squamous [#/volume] in Urine by Automated count** | 2 | 29.66 | 51486-9 |
| **1,550** | **Iron binding capacity [Mass/volume] in Serum or Plasma** | 2 | 29.16 | 2500-7 |
| **1,551** | **Protein [Presence] in Urine by Automated test strip** | 2 | 28.75 | 57735-3 |

| | | | | |
|---|---|---|---|---|
| **1,552** | **Leukocyte esterase [Presence] in Urine by Automated test strip** | 2 | 28.75 | 60026-2 |
| **1,553** | **Nitrite [Presence] in Urine by Automated test strip** | 2 | 28.75 | 50558-6 |
| **1,554** | **Blood Group Systems** | 3 | 28.17 | LP32652-7 |
| **1,555** | **Streptococcus pneumoniae 23 serotypes IgG panel [Mass/volume] - Serum** | 3 | 26.57 | 42366-5 |
| **1,556** | **Streptococcus pneumoniae 23 serotypes IgG panel \| Serum \| Microbiology Panels** | 5 | 26.57 | LP380109-1 |
| **1,557** | **Influenza virus A RNA \| Nasopharynx \| Microbiology** | 4 | 25.86 | LP378496-6 |
| **1,558** | **Respiratory syncytial virus RNA \| XXX \| Microbiology** | 4 | 25.50 | LP378960-1 |
| **1,559** | **Respiratory syncytial virus RNA [Presence] in Specimen by NAA with probe detection** | 2 | 25.50 | 40988-8 |
| **1,560** | **Influenza virus B RNA \| Nasopharynx \| Microbiology** | 4 | 24.70 | LP378553-4 |
| **1,561** | **Herpes virus 6 DNA panel \| XXX \| Microbiology Panels** | 5 | 23.58 | LP380002-8 |
| **1,562** | **Herpes virus 6 DNA panel - Specimen by NAA with probe detection** | 3 | 23.58 | 38351-3 |
| **1,563** | **Hematopoietic progenitor cell product characterization panel - Blood product unit by Flow cytometry (FC)** | 3 | 21.83 | 95830-6 |
| **1,564** | **HLA order set** | 2 | 21.15 | PANEL.HLA |
| **1,565** | **Drugs of abuse 5 panel - Urine by Screen method** | 3 | 20.11 | 65750-2 |
| **1,566** | **Tumor markers** | 4 | 20.11 | LP31412-7 |
| **1,567** | **Drugs of abuse 5 panel\|-\|Urine** | 2 | 20.05 | LG35257-1 |
| **1,568** | **Drugs of abuse 5 panel \| Urine \| Drug and Toxicology Panels** | 4 | 20.05 | LP392063-6 |
| **1,569** | **Coccidioides immitis IgG and IgM panel - Cerebral spinal fluid** | 3 | 19.89 | 88745-5 |
| **1,570** | **Coccidioides immitis IgG and IgM panel \| Cerebral spinal fluid \| Microbiology Panels** | 5 | 19.89 | LP379937-8 |
| **1,571** | **Mycoplasma sp and Ureaplasma sp panel - Specimen by Organism specific culture** | 3 | 19.89 | 43107-2 |
| **1,572** | **Specimen source identified** | 2 | 19.89 | 31208-2 |
| **1,573** | **Mycoplasma sp and Ureaplasma sp panel \| XXX \| Microbiology Panels** | 5 | 19.89 | LP380069-7 |
| **1,574** | **Stone analysis panel** | 3 | 19.79 | 74446-6 |
| **1,575** | **Stone analysis panel \| Calculus (stone) \| Chemistry Panels** | 5 | 19.79 | LP386842-1 |
| **1,576** | **Pancreatic exocrine function panel - Body fluid** | 3 | 19.70 | 95362-0 |
| **1,577** | **Pancreatic exocrine function panel \| Body fluid \| Challenge Bank Panels** | 5 | 19.70 | LP419315-9 |
| **1,578** | **Streptococcus pneumoniae 14 serotypes and Corynebacterium diphtheriae toxin and Clostridium tetani toxin Ab.IgG panel \| Serum \| Microbiology Panels** | 5 | 18.64 | LP380106-7 |
| **1,579** | **Streptococcus pneumoniae 14 serotypes and Corynebacterium diphtheriae toxin and Clostridium tetani toxin Ab.IgG panel - Serum** | 3 | 18.64 | 53932-0 |

| | | | | |
|---|---|---|---|---|
| **1,580** | **Medical devices order set** | 2 | 18.38 | PANEL.DEVICES |
| **1,581** | **Blood type and Crossmatch panel - Blood** | 3 | 17.97 | 34531-4 |
| **1,582** | **Blood type and Indirect antibody screen panel - Blood** | 3 | 17.97 | 34532-2 |
| **1,583** | **Blood type and Indirect antibody screen panel \| Blood \| Blood Bank Panels** | 4 | 17.97 | LP399317-9 |
| **1,584** | **Blood type and Crossmatch panel \| Blood \| Blood Bank Panels** | 4 | 17.97 | LP399316-1 |
| **1,585** | **Streptococcus pneumoniae 14 serotypes IgG panel B \| Serum \| Microbiology Panels** | 5 | 17.72 | LP380108-3 |
| **1,586** | **Streptococcus pneumoniae 14 serotypes IgG panel B [Mass/volume] - Serum** | 3 | 17.72 | 42359-0 |
| **1,587** | **Pregnancy Proteins-** | 4 | 17.54 | LP31662-7 |
| **1,588** | **Streptococcus pneumoniae 14 serotypes IgG panel A [Mass/volume] - Serum** | 3 | 16.73 | 42360-8 |
| **1,589** | **Streptococcus pneumoniae 14 serotypes IgG panel A \| Serum \| Microbiology Panels** | 5 | 16.73 | LP380107-5 |
| **1,590** | **Streptococcus pyogenes DNA [Presence] in Throat by NAA with probe detection** | 2 | 16.46 | 60489-2 |
| **1,591** | **Streptococcus pyogenes DNA\|PrThr\|Sys:ANYResp** | 2 | 16.46 | LG50970-9 |
| **1,592** | **Streptococcus pyogenes DNA \| Throat \| Microbiology** | 4 | 16.46 | LP375959-6 |
| **1,593** | **Bacteria identified \| XXX \| Microbiology** | 4 | 16.33 | LP373762-6 |
| **1,594** | **Celiac disease serology panel - Serum** | 3 | 15.91 | 94494-2 |
| **1,595** | **Leukocytes \| XXX \| Cell markers** | 3 | 14.45 | LP400781-3 |
| **1,596** | **Leukocytes [#/volume] in Specimen by Automated count** | 2 | 14.45 | 20584-9 |
| **1,597** | **Pregnancy test** | 2 | 13.77 | LG50141-7 |
| **1,598** | **Streptococcus pneumoniae 12 serotypes IgG panel \| Serum \| Microbiology Panels** | 5 | 13.68 | LP380105-9 |
| **1,599** | **Streptococcus pneumoniae 12 serotypes IgG panel [Mass/volume] - Serum** | 3 | 13.68 | 42361-6 |
| **1,600** | **25-Hydroxyvitamin D3+25-Hydroxyvitamin D2 \| Serum or Plasma \| Chemistry - non-challenge** | 3 | 13.65 | LP385185-6 |
| **1,601** | **Calcidiol+ercalcidiol\|MCnc\|Pt\|ANYBldSerPl** | 2 | 13.65 | LG25965-1 |
| **1,602** | **Calcidiol+ercalcidiol\|Pt\|Ser/Plas** | 2 | 12.91 | LG42015-4 |
| **1,603** | **25-Hydroxyvitamin D3+25-Hydroxyvitamin D2 [Mass/volume] in Serum or Plasma** | 2 | 12.91 | 62292-8 |
| **1,604** | **Tissue Transglutaminase IgA \| Serum \| Serology - non-micro** | 3 | 10.84 | LP404067-3 |
| **1,605** | **Assay related variables panel [PhenX]** | 3 | 9.85 | 95405-7 |
| **1,606** | **Influenza virus A RNA \| Respiratory specimen \| Microbiology** | 4 | 7.11 | LP378498-2 |
| **1,607** | **Influenza virus A RNA [Presence] in Respiratory specimen by NAA with probe detection** | 2 | 7.11 | 92142-9 |
| **1,608** | **Influenza virus B RNA \| Respiratory specimen \| Microbiology** | 4 | 7.09 | LP378555-9 |
| **1,609** | **Influenza virus B RNA [Presence] in Respiratory specimen by NAA with probe detection** | 2 | 7.09 | 92141-1 |

| | | | | |
|---|---|---|---|---|
| **1,610** | **Hemoglobin A1c/Hemoglobin.total\|MFr\|Pt\|ANYBldSerPl** | 2 | 6.99 | LG51070-7 |
| **1,611** | **Hemoglobin A1c measurement device panel** | 3 | 6.75 | 43150-2 |
| **1,612** | **Hemoglobin A1c/Hemoglobin.total in Blood** | 2 | 6.75 | 4548-4 |

Table 1c: TreeScan-selected procedure features

| Cut | Concept | Tree Level | Log Likelihood Ratio | Procedure code | Vocabulary |
|---|---|---|---|---|---|
| **1** | **Cardiovascular Procedures** | 1 | 8,169.44 | 1012974 | CPT4 |
| **2** | **Echocardiography Procedures** | 2 | 4,970.50 | 1013050 | CPT4 |
| **3** | **Cardiography Procedures** | 2 | 3,020.35 | 1013011 | CPT4 |
| **4** | **Electrocardiogram, routine ECG with at least 12 leads** | 3 | 2,941.79 | 1013012 | CPT4 |
| **5** | **Current Procedural Terminology Concept** | 1 | 2,763.16 | 1002795 | CPT4 |
| **6** | **Echocardiography, transthoracic, real-time with image documentation (2D), includes M-mode recording, when performed, complete, with spectral Doppler echocardiography, and with color flow Doppler echocardiography** | 3 | 2,030.62 | 93306 | CPT4 |
| **7** | **Radiology Procedures** | 2 | 1,901.97 | 1010251 | CPT4 |
| **8** | **Electrocardiogram, routine ECG with at least 12 leads; tracing only, without interpretation and report** | 4 | 1,566.51 | 93005 | CPT4 |
| **9** | **Diagnostic Radiology (Diagnostic Imaging) Procedures** | 3 | 1,413.65 | 1010252 | CPT4 |
| **10** | **Diagnostic Radiology (Diagnostic Imaging) Procedures of the Chest** | 4 | 1,027.31 | 1010334 | CPT4 |
| **11** | **Subsequent Hospital Care Services** | 1 | 928.56 | 1013668 | CPT4 |
| **12** | **Electrocardiogram, routine ECG with at least 12 leads; interpretation and report only** | 4 | 875.52 | 93010 | CPT4 |
| **13** | **Doppler echocardiography color flow velocity mapping (List separately in addition to codes for echocardiography)** | 3 | 862.06 | 93325 | CPT4 |
| **14** | **Echocardiography, transthoracic, real-time with image documentation (2D), includes M-mode recording, when performed, follow-up or limited study** | 3 | 805.65 | 93308 | CPT4 |
| **15** | **Radiologic examination, chest** | 5 | 780.53 | 1031050 | CPT4 |
| **16** | **Physical Medicine and Rehabilitation Evaluations** | 1 | 744.67 | 1013483 | CPT4 |
| **17** | **Doppler echocardiography, pulsed wave and/or continuous wave with spectral display (List separately in addition to codes for echocardiographic imaging)** | 3 | 733.35 | 1013066 | CPT4 |
| **18** | **Doppler echocardiography, pulsed wave and/or continuous wave with spectral display (List separately in addition to codes for echocardiographic imaging); follow-up or limited study (List separately in addition to codes for echocardiographic imaging)** | 4 | 725.40 | 93321 | CPT4 |
| **19** | **Administration @ Circulatory @ Transfusion @ Peripheral Vein @ Percutaneous @ Globulin** | 6 | 692.16 | 30233S | ICD10 PCS |

| | | | | | |
|---|---|---|---|---|---|
| **20** | **Transfusion of Nonautologous Globulin into Peripheral Vein, Percutaneous Approach** | 7 | 692.16 | 30233S1 | ICD10 PCS |
| **21** | **Myocardial strain imaging using speckle tracking-derived assessment of myocardial mechanics (List separately in addition to codes for echocardiography imaging)** | 3 | 685.11 | 93356 | CPT4 |
| **22** | **Pulmonary Procedures** | 1 | 659.96 | 1013214 | CPT4 |
| **23** | **Pathology and Laboratory Procedures** | 2 | 659.94 | 1011136 | CPT4 |
| **25** | **Critical Care Services** | 3 | 628.09 | 1013729 | CPT4 |
| **24** | **Critical care, evaluation and management of the critically ill or critically injured patient** | 4 | 628.09 | 1014309 | CPT4 |
| **26** | **Hydration, Therapeutic, Prophylactic, Diagnostic Injections and Infusions, and Chemotherapy and Other Highly Complex Drug or Highly Complex Biologic Agent Administration** | 1 | 617.37 | 1019103 | CPT4 |
| **27** | **Administration @ Circulatory @ Transfusion @ Peripheral Vein @ Percutaneous** | 5 | 573.51 | 30233 | ICD10 PCS |
| **28** | **Administration @ Circulatory @ Transfusion @ Peripheral Vein** | 4 | 573.51 | 3023 | ICD10 PCS |
| **30** | **Administration, Circulatory, Transfusion** | 3 | 560.74 | 302 | ICD10 PCS |
| **29** | **Administration @ Circulatory (Procedure)** | 2 | 560.74 | 30 | ICD10 PCS |
| **31** | **Pulmonary Diagnostic Testing and Therapies** | 2 | 545.94 | 1015099 | CPT4 |
| **32** | **Diagnostic Ultrasound Procedures** | 3 | 538.17 | 1010759 | CPT4 |
| **33** | **Electrocardiogram, routine ECG with at least 12 leads; with interpretation and report** | 4 | 499.92 | 93000 | CPT4 |
| **34** | **Administration (Procedure)** | 1 | 475.74 | 3 | ICD10 PCS |
| **35** | **Subsequent hospital care, per day, for the evaluation and management of a patient, which requires at least 2 of these 3 key components: A detailed interval history; A detailed examination; Medical decision making of high complexity. Counseling and/or coor** | 2 | 474.97 | 99233 | CPT4 |
| **36** | **Cardiovascular Monitoring Services** | 2 | 466.34 | 1020409 | CPT4 |
| **37** | **Initial Hospital Inpatient Care Services** | 1 | 438.99 | 1013660 | CPT4 |
| **38** | **New or Established Patient Initial Hospital Inpatient Care Services** | 2 | 438.99 | 1013661 | CPT4 |
| **39** | **Radiologic examination, chest; 2 views** | 6 | 422.90 | 71046 | CPT4 |
| **40** | **Subsequent hospital care, per day, for the evaluation and management of a patient, which requires at least 2 of these 3 key components: An expanded problem focused interval history; An expanded problem focused examination; Medical decision making of moder** | 2 | 404.71 | 99232 | CPT4 |
| **41** | **Hospital discharge day management** | 1 | 400.32 | 1013683 | CPT4 |
| **42** | **Diagnostic Ultrasound Procedures of the Abdomen and Retroperitoneum** | 4 | 394.34 | 1010774 | CPT4 |

| | | | | | |
|---|---|---|---|---|---|
| **43** | **Therapeutic, Prophylactic, and Diagnostic Injections and Infusions (Excludes Chemotherapy and Other Highly Complex Drug or Highly Complex Biologic Agent Administration)** | 2 | 385.86 | 1019108 | CPT4 |
| **44** | **Critical care, evaluation and management of the critically ill or critically injured patient; first 30-74 minutes** | 5 | 385.56 | 99291 | CPT4 |
| **45** | **Surgical Procedures on the Cardiovascular System** | 3 | 383.31 | 1006056 | CPT4 |
| **46** | **Surgical Procedures on Arteries and Veins** | 4 | 380.47 | 1006359 | CPT4 |
| **47** | **Transfusion Medicine Procedures** | 3 | 374.19 | 1012085 | CPT4 |
| **48** | **Radiologic examination, chest; single view** | 6 | 361.71 | 71045 | CPT4 |
| **49** | **Ultrasound, abdominal, real time with image documentation** | 5 | 348.37 | 1010775 | CPT4 |
| **50** | **Vascular Introduction and Injection Procedures** | 5 | 336.04 | 1013922 | CPT4 |
| **51** | **Evaluation and Management Services** | 2 | 333.29 | 1013625 | CPT4 |
| **52** | **Proprietary Laboratory Analyses** | 3 | 332.98 | 1029999 | CPT4 |
| **53** | **External electrocardiographic recording up to 48 hours by continuous rhythm recording and storage** | 3 | 326.97 | 1013030 | CPT4 |
| **55** | **Physical Medicine and Rehabilitation Therapeutic Procedures** | 2 | 319.68 | 1013510 | CPT4 |
| **56** | **Intravenous infusion, hydration** | 3 | 306.86 | 1019330 | CPT4 |
| **57** | **Hydration Infusion** | 2 | 306.86 | 1019105 | CPT4 |
| **58** | **Collection of venous blood by venipuncture** | 7 | 289.75 | 36415 | CPT4 |
| **59** | **Diagnostic Radiology (Diagnostic Imaging) Procedures of the Heart** | 4 | 283.67 | 1010594 | CPT4 |
| **60** | **Diagnostic Radiology (Diagnostic Imaging) Procedures of the Abdomen** | 4 | 280.84 | 1010520 | CPT4 |
| **61** | **Physical Therapy Evaluations** | 2 | 272.44 | 1029677 | CPT4 |
| **62** | **Hospital discharge day management; 30 minutes or less** | 2 | 264.69 | 99238 | CPT4 |
| **63** | **Initial hospital care, per day, for the evaluation and management of a patient, which requires these 3 key components: A comprehensive history; A comprehensive examination; and Medical decision making of high complexity. Counseling and/or coordination of** | 3 | 250.52 | 99223 | CPT4 |
| **64** | **Critical care, evaluation and management of the critically ill or critically injured patient; each additional 30 minutes (List separately in addition to code for primary service)** | 5 | 248.25 | 99292 | CPT4 |
| **65** | **Venous Procedures** | 6 | 243.23 | 1006654 | CPT4 |
| **66** | **Blood typing, serologic** | 4 | 225.13 | 1022269 | CPT4 |
| **67** | **Surgery** | 2 | 217.39 | 1003143 | CPT4 |
| **68** | **Diagnostic Radiology (Diagnostic Imaging) Procedures of the Head and Neck** | 4 | 204.62 | 1010253 | CPT4 |
| **69** | **Therapeutic, prophylactic, or diagnostic injection (specify substance or drug)** | 3 | 199.20 | 1019333 | CPT4 |
| **70** | **Intravenous infusion, for therapy, prophylaxis, or diagnosis (specify substance or drug)** | 3 | 195.63 | 1019331 | CPT4 |
| **72** | **Extracorporeal or Systemic Assistance and Performance @ Physiological Systems (Procedure)** | 2 | 188.61 | 5A | ICD10 PCS |

| | | | | | |
|---|---|---|---|---|---|
| **71** | **Extracorporeal or Systemic Assistance and Performance (Procedure)** | 1 | 188.61 | 5 | ICD10 PCS |
| **73** | **Bronchodilation responsiveness, spirometry as in 94010, pre- and post-bronchodilator administration** | 3 | 186.92 | 94060 | CPT4 |
| **74** | **Ultrasound, abdominal, real time with image documentation; complete** | 6 | 180.99 | 76700 | CPT4 |
| **75** | **Ultrasound, abdominal, real time with image documentation; limited (eg, single organ, quadrant, follow-up)** | 6 | 179.86 | 76705 | CPT4 |
| **76** | **Occupational Therapy Evaluations** | 2 | 178.49 | 1029678 | CPT4 |
| **77** | **Cardiac magnetic resonance imaging for morphology and function without contrast material(s), followed by contrast material(s) and further sequences** | 6 | 173.58 | 75561 | CPT4 |
| **78** | **Medical and Surgical (Procedure)** | 1 | 172.99 | 0 | ICD10 PCS |
| **79** | **Cardiac magnetic resonance imaging for morphology and function without contrast material(s), followed by contrast material(s) and further sequences** | 5 | 172.32 | 1018519 | CPT4 |
| **80** | **Imaging (Procedure)** | 1 | 165.26 | B | ICD10 PCS |
| **81** | **Upper Arteries, Insertion** | 3 | 161.86 | 03H | ICD10 PCS |
| **82** | **Intravenous infusion, hydration; each additional hour (List separately in addition to code for primary procedure)** | 4 | 160.30 | 96361 | CPT4 |
| **83** | **Intravenous infusion, for therapy, prophylaxis, or diagnosis (specify substance or drug); initial, up to 1 hour** | 4 | 158.78 | 96365 | CPT4 |
| **84** | **Medical and Surgical @ Upper Arteries (Procedure)** | 2 | 154.42 | 03 | ICD10 PCS |
| **85** | **Radiologic examination, abdomen** | 5 | 153.80 | 1031051 | CPT4 |
| **86** | **Physical therapy evaluation: moderate complexity, requiring these components: A history of present problem with 1-2 personal factors and/or comorbidities that impact the plan of care; An examination of body systems using standardized tests and measures in** | 3 | 153.11 | 97162 | CPT4 |
| **87** | **Imaging, Heart, Ultrasonography** | 3 | 152.65 | B24 | ICD10 PCS |
| **88** | **Non-Invasive Vascular Diagnostic Studies** | 1 | 149.27 | 1013175 | CPT4 |
| **89** | **External electrocardiographic recording up to 48 hours by continuous rhythm recording and storage; recording (includes connection, recording, and disconnection)** | 4 | 148.27 | 93225 | CPT4 |
| **90** | **Intravenous infusion, hydration; initial, 31 minutes to 1 hour** | 4 | 147.53 | 96360 | CPT4 |
| **91** | **Imaging @ Heart (Procedure)** | 2 | 143.39 | B2 | ICD10 PCS |
| **92** | **Ultrasonography of Pediatric Heart** | 7 | 142.88 | B24DZZZ | ICD10 PCS |

| | | | | | |
|---|---|---|---|---|---|
| **93** | **Imaging @ Heart @ Ultrasonography @ Pediatric Heart** | 4 | 139.64 | B24D | ICD10 PCS |
| **94** | **Therapeutic procedure, 1 or more areas, each 15 minutes** | 3 | 136.90 | 1013511 | CPT4 |
| **95** | **Hospital discharge day management; more than 30 minutes** | 2 | 136.17 | 99239 | CPT4 |
| **96** | **Diffusing capacity (eg, carbon monoxide, membrane) (List separately in addition to code for primary procedure)** | 3 | 135.07 | 94729 | CPT4 |
| **98** | **Imaging @ Heart @ Ultrasonography @ Pediatric Heart @ None @ None** | 6 | 132.01 | B24DZZ | ICD10 PCS |
| **97** | **Imaging @ Heart @ Ultrasonography @ Pediatric Heart @ None** | 5 | 132.01 | B24DZ | ICD10 PCS |
| **100** | **Insertion of Monitoring Device into Upper Artery, Percutaneous Approach** | 7 | 130.42 | 03HY32Z | ICD10 PCS |
| **99** | **Medical and Surgical @ Upper Arteries @ Insertion @ Upper Artery @ Percutaneous @ Monitoring Device** | 6 | 130.42 | 03HY32 | ICD10 PCS |
| **101** | **Medical and Surgical @ Upper Arteries @ Insertion @ Upper Artery @ Percutaneous** | 5 | 129.95 | 03HY3 | ICD10 PCS |
| **102** | **Medical and Surgical @ Upper Arteries @ Insertion @ Upper Artery** | 4 | 129.49 | 03HY | ICD10 PCS |
| **103** | **Cardiac magnetic resonance imaging for velocity flow mapping (List separately in addition to code for primary procedure)** | 5 | 127.09 | 75565 | CPT4 |
| **104** | **Therapeutic, prophylactic, or diagnostic injection (specify substance or drug); each additional sequential intravenous push of a new substance/drug (List separately in addition to code for primary procedure)** | 4 | 124.66 | 96375 | CPT4 |
| **105** | **Computed tomography, soft tissue neck; with contrast material(s)** | 6 | 124.45 | 70491 | CPT4 |
| **106** | **Ventilator Management** | 2 | 124.34 | 1015098 | CPT4 |
| **107** | **Ventilation assist and management, initiation of pressure or volume preset ventilators for assisted or controlled breathing** | 3 | 124.34 | 1014859 | CPT4 |
| **108** | **Computed tomography, soft tissue neck** | 5 | 124.07 | 1010308 | CPT4 |
| **109** | **Transthoracic echocardiography for congenital cardiac anomalies** | 3 | 123.02 | 1013051 | CPT4 |
| **110** | **Extracorporeal or Systemic Assistance and Performance, Physiological Systems, Performance** | 3 | 121.58 | 5A1 | ICD10 PCS |
| **111** | **Blood typing, serologic; ABO** | 5 | 117.37 | 86900 | CPT4 |
| **112** | **Antibody screen, RBC, each serum technique** | 4 | 115.17 | 86850 | CPT4 |
| **113** | **Pulmonary stress testing (eg, 6-minute walk test), including measurement of heart rate, oximetry, and oxygen titration, when performed** | 3 | 111.93 | 94618 | CPT4 |
| **114** | **Blood typing, serologic; Rh (D)** | 5 | 109.51 | 86901 | CPT4 |
| **115** | **Therapeutic, prophylactic, or diagnostic injection (specify substance or drug); intravenous push, single or initial substance/drug** | 4 | 107.50 | 96374 | CPT4 |

| | | | | | |
|---|---|---|---|---|---|
| **116** | **Plethysmography for determination of lung volumes and, when performed, airway resistance** | 3 | 106.12 | 94726 | CPT4 |
| **117** | **Therapeutic procedure, 1 or more areas, each 15 minutes; therapeutic exercises to develop strength and endurance, range of motion and flexibility** | 4 | 105.06 | 97110 | CPT4 |
| **118** | **Extracorporeal or Systemic Assistance and Performance @ Physiological Systems @ Performance @ Respiratory** | 4 | 103.74 | 5A19 | ICD10 PCS |
| **119** | **Therapeutic activities, direct (one-on-one) patient contact (use of dynamic activities to improve functional performance), each 15 minutes** | 3 | 103.63 | 97530 | CPT4 |
| **120** | **Initial hospital care, per day, for the evaluation and management of a patient, which requires these 3 key components: A detailed or comprehensive history; A detailed or comprehensive examination; and Medical decision making that is straightforward or of** | 3 | 102.72 | 99221 | CPT4 |
| **121** | **Continuous positive airway pressure ventilation (CPAP), initiation and management** | 3 | 101.32 | 94660 | CPT4 |
| **122** | **Insertion of Central Venous Access Device** | 7 | 101.10 | 1006697 | CPT4 |
| **123** | **3D rendering with interpretation and reporting of computed tomography, magnetic resonance imaging, ultrasound, or other tomographic modality with image postprocessing under concurrent supervision; not requiring image postprocessing on an independent works** | 6 | 96.91 | 76376 | CPT4 |
| **124** | **Occupational therapy evaluation, moderate complexity, requiring these components: An occupational profile and medical and therapy history, which includes an expanded review of medical and/or therapy records and additional review of physical, cognitive, or** | 3 | 96.64 | 97166 | CPT4 |
| **125** | **Radiologic examination, abdomen; 1 view** | 6 | 95.43 | 74018 | CPT4 |
| **126** | **Initial hospital care, per day, for the evaluation and management of a patient, which requires these 3 key components: A comprehensive history; A comprehensive examination; and Medical decision making of moderate complexity. Counseling and/or coordination** | 3 | 94.54 | 99222 | CPT4 |
| **127** | **Computed tomography, abdomen and pelvis; with contrast material(s)** | 6 | 94.30 | 74177 | CPT4 |
| **128** | **Central Venous Access Procedures** | 6 | 94.13 | 1006696 | CPT4 |
| **129** | **Computed tomography, abdomen and pelvis** | 5 | 94.04 | 1020544 | CPT4 |
| **130** | **Physical therapy evaluation: high complexity, requiring these components: A history of present problem with 3 or more personal factors and/or comorbidities that impact the plan of care; An examination of body systems using standardized tests and measures** | 3 | 90.24 | 97163 | CPT4 |
| **131** | **Subsequent hospital care, per day, for the evaluation and management of a patient, which requires at least 2 of these 3 key components: A problem focused interval** | 2 | 89.94 | 99231 | CPT4 |

| | | | | | |
|---|---|---|---|---|---|
| | **history; A problem focused examination; Medical decision making that is straightforward or o** | | | | |
| **132** | **Doppler echocardiography, pulsed wave and/or continuous wave with spectral display (List separately in addition to codes for echocardiographic imaging); complete** | 4 | 89.49 | 93320 | CPT4 |
| **133** | **Respiratory System, Insertion** | 3 | 89.31 | 0BH | ICD10 PCS |
| **134** | **Medical and Surgical @ Respiratory System @ Insertion @ Trachea** | 4 | 89.31 | 0BH1 | ICD10 PCS |
| **135** | **Self-care/home management training (eg, activities of daily living (ADL) and compensatory training, meal preparation, safety procedures, and instructions in use of assistive technology devices/adaptive equipment) direct one-on-one contact, each 15 minutes** | 3 | 88.50 | 97535 | CPT4 |
| **136** | **Inpatient Neonatal and Pediatric Critical Care Services** | 4 | 87.66 | 1019134 | CPT4 |
| **137** | **Heart and Great Vessels, Insertion** | 3 | 86.17 | 02H | ICD10 PCS |
| **138** | **Computed tomographic angiography, chest (noncoronary), with contrast material(s), including noncontrast images, if performed, and image postprocessing** | 5 | 84.52 | 71275 | CPT4 |
| **139** | **Inpatient Neonatal Intensive Care Services and Pediatric and Neonatal Critical Care Services** | 3 | 83.27 | 1019130 | CPT4 |
| **140** | **External electrocardiographic recording up to 48 hours by continuous rhythm recording and storage; scanning analysis with report** | 4 | 82.63 | 93226 | CPT4 |
| **141** | **3D rendering with interpretation and reporting of computed tomography, magnetic resonance imaging, ultrasound, or other tomographic modality with image postprocessing under concurrent supervision** | 5 | 81.14 | 1010748 | CPT4 |
| **142** | **Duplex scan of extremity veins including responses to compression and other maneuvers** | 3 | 79.30 | 1013199 | CPT4 |
| **143** | **Non-Invasive Extremity Venous Studies (Including Digits)** | 2 | 79.30 | 1013197 | CPT4 |
| **144** | **Medical and Surgical @ Respiratory System (Procedure)** | 2 | 79.29 | 0B | ICD10 PCS |
| **145** | **External electrocardiographic recording for more than 48 hours up to 7 days by continuous rhythm recording and storage** | 3 | 78.34 | 1036233 | CPT4 |
| **147** | **Medical and Surgical @ Heart and Great Vessels @ Insertion @ Superior Vena Cava @ Percutaneous @ Infusion Device** | 6 | 77.42 | 02HV33 | ICD10 PCS |
| **149** | **Medical and Surgical @ Heart and Great Vessels @ Insertion @ Superior Vena Cava** | 4 | 77.42 | 02HV | ICD10 PCS |
| **148** | **Insertion of Infusion Device into Superior Vena Cava, Percutaneous Approach** | 7 | 77.42 | 02HV33Z | ICD10 PCS |
| **146** | **Medical and Surgical @ Heart and Great Vessels @ Insertion @ Superior Vena Cava @ Percutaneous** | 5 | 77.42 | 02HV3 | ICD10 PCS |

| | | | | | |
|---|---|---|---|---|---|
| **150** | **Transthoracic echocardiography for congenital cardiac anomalies; complete** | 4 | 76.97 | 93303 | CPT4 |
| **151** | **Ultrasound guidance for vascular access requiring ultrasound evaluation of potential access sites, documentation of selected vessel patency, concurrent realtime ultrasound visualization of vascular needle entry, with permanent recording and reporting (Lis** | 5 | 75.05 | 76937 | CPT4 |
| **152** | **Radiologic examination, chest; single view, frontal** | 6 | 74.17 | 71010 | CPT4 |
| **153** | **Radiologic examination, chest** | 5 | 74.17 | 1010335 | CPT4 |
| **154** | **Insertion of non-tunneled centrally inserted central venous catheter** | 8 | 73.65 | 1006698 | CPT4 |
| **155** | **Arterial catheterization or cannulation for sampling, monitoring or transfusion (separate procedure)** | 6 | 72.92 | 1006739 | CPT4 |
| **157** | **Medical and Surgical @ Respiratory System @ Insertion @ Trachea @ Via Natural or Artificial Opening** | 5 | 72.12 | 0BH17 | ICD10 PCS |
| **158** | **Insertion of Endotracheal Airway into Trachea, Via Natural or Artificial Opening** | 7 | 72.12 | 0BH17EZ | ICD10 PCS |
| **156** | **Medical and Surgical @ Respiratory System @ Insertion @ Trachea @ Via Natural or Artificial Opening @ Intraluminal Device, Endotracheal Airway** | 6 | 72.12 | 0BH17E | ICD10 PCS |
| **159** | **Arterial catheterization or cannulation for sampling, monitoring or transfusion (separate procedure); percutaneous** | 7 | 71.86 | 36620 | CPT4 |
| **160** | **Ventilation assist and management, initiation of pressure or volume preset ventilators for assisted or controlled breathing; hospital inpatient/observation, each subsequent day** | 4 | 71.71 | 94003 | CPT4 |
| **161** | **Arterial Procedures** | 5 | 69.73 | 1006737 | CPT4 |
| **162** | **Ultrasonic Guidance Procedures** | 4 | 68.91 | 1010824 | CPT4 |
| **163** | **Other Diagnostic Radiology (Diagnostic Imaging) Related Procedures** | 4 | 67.80 | 1010693 | CPT4 |
| **164** | **Noninvasive ear or pulse oximetry for oxygen saturation; multiple determinations (eg, during exercise)** | 4 | 67.05 | 94761 | CPT4 |
| **165** | **Ultrasound, retroperitoneal (eg, renal, aorta, nodes), real time with image documentation** | 5 | 66.19 | 1010778 | CPT4 |
| **166** | **Medical and Surgical @ Heart and Great Vessels (Procedure)** | 2 | 65.87 | 02 | ICD10 PCS |
| **167** | **Spirometry, including graphic record, total and timed vital capacity, expiratory flow rate measurement(s), with or without maximal voluntary ventilation** | 3 | 63.33 | 94010 | CPT4 |
| **168** | **Ultrasound, soft tissues of head and neck (eg, thyroid, parathyroid, parotid), real time with image documentation** | 5 | 62.86 | 76536 | CPT4 |
| **169** | **Extracorporeal or Systemic Assistance and Performance, Physiological Systems, Assistance** | 3 | 61.54 | 5A0 | ICD10 PCS |
| **170** | **Extracorporeal or Systemic Assistance and Performance @ Physiological Systems @ Assistance @ Respiratory** | 4 | 58.70 | 5A09 | ICD10 PCS |
| **171** | **Unlisted pulmonary service or procedure** | 3 | 56.34 | 94799 | CPT4 |

| | | | | | |
|---|---|---|---|---|---|
| **172** | **Radiologic examination, chest, 2 views, frontal and lateral** | 5 | 56.13 | 1014245 | CPT4 |
| **173** | **Radiologic examination, chest, 2 views, frontal and lateral** | 6 | 56.13 | 71020 | CPT4 |
| **174** | **Radiologic examination, abdomen; 2 views** | 6 | 54.63 | 74019 | CPT4 |
| **175** | **Administration, Physiological Systems and Anatomical Regions, Introduction** | 3 | 53.32 | 3E0 | ICD10 PCS |
| **176** | **Ventilation assist and management, initiation of pressure or volume preset ventilators for assisted or controlled breathing; hospital inpatient/observation, initial day** | 4 | 52.99 | 94002 | CPT4 |
| **177** | **Magnetic resonance (eg, proton) imaging, brain (including brain stem)** | 5 | 52.80 | 1010326 | CPT4 |
| **178** | **Administration @ Physiological Systems and Anatomical Regions (Procedure)** | 2 | 52.62 | 3E | ICD10 PCS |
| **179** | **Ultrasound, retroperitoneal (eg, renal, aorta, nodes), real time with image documentation; complete** | 6 | 51.37 | 76770 | CPT4 |
| **180** | **Duplex scan of arterial inflow and venous outflow of abdominal, pelvic, scrotal contents and/or retroperitoneal organs** | 3 | 47.79 | 1013203 | CPT4 |
| **181** | **Transthoracic echocardiography for congenital cardiac anomalies; follow-up or limited study** | 4 | 47.48 | 93304 | CPT4 |
| **182** | **Non-Invasive Visceral and Penile Vascular Studies** | 2 | 46.51 | 1013202 | CPT4 |
| **183** | **Diagnostic Ultrasound Procedures of the Head and Neck** | 4 | 45.74 | 1010760 | CPT4 |
| **184** | **Surgical Procedures on the Respiratory System** | 3 | 45.68 | 1005690 | CPT4 |
| **185** | **Magnetic resonance (eg, proton) imaging, brain (including brain stem); without contrast material, followed by contrast material(s) and further sequences** | 6 | 45.40 | 70553 | CPT4 |
| **186** | **Duplex scan of extremity veins including responses to compression and other maneuvers; unilateral or limited study** | 4 | 45.37 | 93971 | CPT4 |
| **187** | **Computed tomography, thorax, diagnostic** | 5 | 42.06 | 1036223 | CPT4 |
| **188** | **Occupational therapy evaluation, low complexity, requiring these components: An occupational profile and medical and therapy history, which includes a brief history including review of medical and/or therapy records relating to the presenting problem; An** | 3 | 40.08 | 97165 | CPT4 |
| **189** | **Computed tomography, head or brain** | 5 | 39.87 | 1010296 | CPT4 |
| **190** | **Duplex scan of arterial inflow and venous outflow of abdominal, pelvic, scrotal contents and/or retroperitoneal organs; complete study** | 4 | 38.64 | 93975 | CPT4 |
| **191** | **Measurement and Monitoring @ Physiological Systems (Procedure)** | 2 | 38.11 | 4A | ICD10 PCS |
| **192** | **Measurement and Monitoring (Procedure)** | 1 | 37.79 | 4 | ICD10 PCS |
| **193** | **Measurement and Monitoring, Physiological Systems, Monitoring** | 3 | 37.12 | 4A1 | ICD10 PCS |
| **194** | **Physical therapy evaluation: low complexity, requiring these components: A history with no personal factors and/or comorbidities that impact the plan of care; An** | 3 | 36.49 | 97161 | CPT4 |

| | | | | | |
|---|---|---|---|---|---|
| | **examination of body system(s) using standardized tests and measures addressing 1-2 elements f** | | | | |
| **195** | **Noninvasive ear or pulse oximetry for oxygen saturation; by continuous overnight monitoring (separate procedure)** | 4 | 36.30 | 94762 | CPT4 |
| **196** | **Surgical Procedures on the Larynx** | 4 | 35.15 | 1005814 | CPT4 |
| **197** | **Special EEG Testing Procedures** | 2 | 33.75 | 1013381 | CPT4 |
| **198** | **Neurology and Neuromuscular Procedures** | 1 | 32.71 | 1013309 | CPT4 |
| **199** | **Long-term EEG Monitoring** | 3 | 30.75 | 1035629 | CPT4 |
| **200** | **Medical and Surgical @ Gastrointestinal System (Procedure)** | 2 | 30.68 | 0D | ICD10 PCS |
| **201** | **Health behavior assessment, or re-assessment (ie, health-focused clinical interview, behavioral observations, clinical decision making)** | 2 | 30.52 | 96156 | CPT4 |
| **202** | **Noninvasive ear or pulse oximetry for oxygen saturation** | 3 | 29.92 | 1013256 | CPT4 |
| **203** | **Manipulation chest wall, such as cupping, percussing, and vibration to facilitate lung function** | 3 | 29.77 | 1013246 | CPT4 |
| **204** | **Computed tomography, head or brain; without contrast material** | 6 | 28.71 | 70450 | CPT4 |
| **205** | **Gastrointestinal System, Excision** | 3 | 28.58 | 0DB | ICD10 PCS |
| **206** | **Radiologic examination, abdomen** | 5 | 25.43 | 1010521 | CPT4 |
| **208** | **Prolonged office or other outpatient evaluation and management service(s) beyond the minimum required time of the primary procedure which has been selected using total time, requiring total time with or without direct patient contact beyond the usual s…** | 2 | 22.86 | 99417 | CPT4 |
| **207** | **Prolonged Service With or Without Direct Patient Contact on the Date of an Office or Other Outpatient Service** | 1 | 22.86 | 1036204 | CPT4 |
| **209** | **Medical and Surgical @ Central Nervous System and Cranial Nerves (Procedure)** | 2 | 22.42 | 00 | ICD10 PCS |
| **210** | **Health Behavior Assessment and Intervention Procedures** | 1 | 21.83 | 1013429 | CPT4 |
| **211** | **Pressurized or nonpressurized inhalation treatment for acute airway obstruction for therapeutic purposes and/or for diagnostic purposes such as sputum induction with an aerosol generator, nebulizer, metered dose inhaler or intermittent positive pressure b** | 3 | 17.81 | 94640 | CPT4 |
| **212** | **Drug Assay Procedures** | 3 | 15.68 | 1021848 | CPT4 |
| **213** | **Magnetic resonance (eg, proton) imaging, brain (including brain stem); without contrast material** | 6 | 13.15 | 70551 | CPT4 |
| **214** | **Initial Hospital Observation Care Services** | 1 | 12.08 | 1013651 | CPT4 |
| **215** | **New or Established Patient Initial Hospital Observation Care Services** | 2 | 12.08 | 1013652 | CPT4 |
| **216** | **Diagnostic Radiology (Diagnostic Imaging) Procedures of the Spine and Pelvis** | 4 | 10.01 | 1010367 | CPT4 |
| **217** | **Definitive Drug Testing Procedures** | 4 | 6.75 | 1021853 | CPT4 |

**Table 1d: TreeScan-selected medication features**

| Cut | Concept | Tree Level | Log Likelihood Ratio | Medication code | Vocabulary |
|---|---|---|---|---|---|
| 1 | Electrolyte solutions | 2 | 6,365.92 | B05XA | ATC |
| 2 | electrolytes in combination with other drugs; parenteral (electrolyte solutions) | 3 | 6,318.12 | B05XA31 | ATC |
| 3 | Other antidiarrheals | 2 | 5,875.08 | A07XA | ATC |
| 4 | Calcium | 2 | 5,875.08 | A12AA | ATC |
| 5 | calcium (different salts in combination); oral | 3 | 5,875.08 | A12AA20 | ATC |
| 6 | calcium compounds; oral | 3 | 5,804.92 | A07XA03 | ATC |
| 7 | Osmotically acting laxatives | 2 | 5,698.74 | A06AD | ATC |
| 8 | mineral salts in combination; oral | 3 | 5,698.74 | A06AD10 | ATC |
| 9 | Other sclerosing agents | 2 | 5,665.37 | C05BX | ATC |
| 10 | calcium dobesilate, combinations; oral, topical | 3 | 5,664.64 | C05BX51 | ATC |
| 11 | Imidazole derivatives | 2 | 5,656.12 | G01AF | ATC |
| 12 | combinations of imidazole derivatives; systemic, vaginal | 3 | 5,632.92 | G01AF20 | ATC |
| 13 | electrolytes with carbohydrates; parenteral | 3 | 5,509.14 | B05BB02 | ATC |
| 14 | Solutions affecting the electrolyte balance | 2 | 5,506.52 | B05BB | ATC |
| 15 | electrolytes; parenteral | 3 | 5,506.44 | B05BB01 | ATC |
| 16 | Amides | 2 | 4,212.22 | N01BB | ATC |
| 17 | amides - combinations; parenteral | 3 | 4,209.95 | N01BB20 | ATC |
| 18 | combinations of electrolytes; parenteral | 3 | 4,147.61 | B05XA30 | ATC |
| 19 | Centrally acting antiobesity products | 2 | 4,138.24 | A08AA | ATC |
| 20 | ephedrine, combinations; systemic | 3 | 4,136.79 | A08AA56 | ATC |
| 21 | lidocaine, combinations; parenteral | 3 | 4,102.08 | N01BB52 | ATC |
| 22 | electrolytes in combination with other drugs; parenteral (solutions affecting the electrolyte balance) | 3 | 4,099.21 | B05BB04 | ATC |
| 23 | Solutions for parenteral nutrition | 2 | 4,073.73 | B05BA | ATC |
| 24 | Salicylic acid and derivatives | 2 | 4,024.41 | N02BA | ATC |
| 25 | carbohydrates; parenteral | 3 | 3,970.89 | B05BA03 | ATC |
| 26 | glucose, combinations; parenteral | 3 | 3,966.08 | C05BB56 | ATC |
| 27 | Sclerosing agents for local injection | 2 | 3,961.57 | C05BB | ATC |
| 28 | Belladonna alkaloids, tertiary amines | 2 | 3,693.82 | A03BA | ATC |
| 29 | Antiinfectives | 2 | 3,668.13 | S03AA | ATC |
| 30 | belladonna total alkaloids; oral | 3 | 3,644.04 | A03BA04 | ATC |
| 31 | Antipropulsives | 2 | 3,642.70 | A07DA | ATC |
| 32 | loperamide, combinations; oral | 3 | 3,544.89 | A07DA53 | ATC |
| 33 | Esters of aminobenzoic acid | 2 | 3,541.29 | N01BA | ATC |
| 34 | Heparin group | 2 | 3,515.66 | B01AB | ATC |
| 35 | procaine, combinations; parenteral | 3 | 3,475.84 | N01BA52 | ATC |
| 36 | Potassium | 2 | 3,475.30 | A12BA | ATC |
| 37 | Vitamin B12 (cyanocobalamin and analogues) | 2 | 3,472.57 | B03BA | ATC |
| 38 | antiinfectives, combinations; ophthalmic, otic | 3 | 3,467.30 | S03AA30 | ATC |
| 39 | cyanocobalamin, combinations; systemic | 3 | 3,446.95 | B03BA51 | ATC |
| 40 | medicated shampoos - others; topical | 3 | 3,371.34 | D11AC30 | ATC |
| 41 | Medicated shampoos | 2 | 3,366.33 | D11AC | ATC |

| | | | | | |
|---|---|---|---|---|---|
| **42** | **Natural opium alkaloids** | 2 | 3,296.76 | N02AA | ATC |
| **43** | **Xanthines** | 2 | 3,221.25 | R03DA | ATC |
| **44** | **potassium chloride, combinations; systemic** | 3 | 3,177.74 | A12BA51 | ATC |
| **45** | **Contact laxatives** | 2 | 3,163.12 | A06AB | ATC |
| **46** | **Organic nitrates** | 2 | 3,135.55 | C01DA | ATC |
| **47** | **codeine, combinations excl. psycholeptics; systemic** | 3 | 3,089.50 | N02AA59 | ATC |
| **48** | **Hypnotics and sedatives in combination, excl. barbiturates** | 2 | 3,071.49 | N05CX | ATC |
| **49** | **meprobamate, comb.; systemic (hypnotics and sedatives in comb., excl. barbiturates)** | 3 | 3,062.22 | N05CX01 | ATC |
| **50** | **meprobamate, comb.; systemic (carbamates)** | 3 | 3,062.22 | N05BC51 | ATC |
| **51** | **Carbamates** | 2 | 3,062.22 | N05BC | ATC |
| **52** | **Pyrazolones** | 2 | 3,018.06 | N02BB | ATC |
| **53** | **combinations of xanthines; systemic** | 3 | 3,014.03 | R03DA20 | ATC |
| **54** | **ethenzamide, combinations excl. psycholeptics; oral, rectal, topical** | 3 | 2,996.57 | N02BA57 | ATC |
| **55** | **heparin, combinations; systemic** | 3 | 2,980.39 | B01AB51 | ATC |
| **56** | **amino acids; parenteral** | 3 | 2,967.57 | B05BA01 | ATC |
| **57** | **Anilides** | 2 | 2,952.01 | N02BE | ATC |
| **58** | **Iron in other combinations** | 2 | 2,919.14 | B03AE | ATC |
| **59** | **various combinations; systemic** | 3 | 2,918.87 | B03AE10 | ATC |
| **60** | **acetylsalicylic acid, combinations excl. psycholeptics; systemic** | 3 | 2,877.58 | N02BA51 | ATC |
| **61** | **combinations; parenteral (solutions for parenteral nutrition)** | 3 | 2,859.75 | B05BA10 | ATC |
| **62** | **arginine and lysine; systemic** | 3 | 2,858.93 | V03AF11 | ATC |
| **63** | **Detoxifying agents for antineoplastic treatment** | 2 | 2,839.22 | V03AF | ATC |
| **64** | **Corticosteroids, weak, other combinations** | 2 | 2,819.22 | D07XA | ATC |
| **65** | **potassium phosphate, incl. combinations with other potassium salts; parenteral** | 3 | 2,812.88 | B05XA06 | ATC |
| **66** | **Salt solutions** | 2 | 2,779.46 | B05CB | ATC |
| **67** | **salt solutions - combinations; irrigating solution** | 3 | 2,779.46 | B05CB10 | ATC |
| **68** | **colecalciferol, combinations; systemic** | 3 | 2,773.36 | A11CC55 | ATC |
| **69** | **combinations of corticosteroids; topical (corticosteroids, moderately potent, other comb.)** | 3 | 2,769.51 | D07XB30 | ATC |
| **70** | **Corticosteroids, moderately potent, other combinations** | 2 | 2,769.51 | D07XB | ATC |
| **71** | **combinations of corticosteroids; topical (corticosteroids, moderately potent (group ii))** | 3 | 2,765.35 | D07AB30 | ATC |
| **72** | **Corticosteroids, moderately potent (group II)** | 2 | 2,765.35 | D07AB | ATC |
| **73** | **Vitamin D and analogues** | 2 | 2,745.04 | A11CC | ATC |
| **74** | **paracetamol, combinations excl. psycholeptics; systemic** | 3 | 2,721.47 | N02BE51 | ATC |
| **75** | **Ergot alkaloids** | 2 | 2,703.74 | N02CA | ATC |
| **76** | **sulfur compounds; topical** | 3 | 2,695.44 | D11AC08 | ATC |
| **77** | **chlorphenamine, combinations; systemic** | 3 | 2,660.41 | R06AB54 | ATC |
| **78** | **Substituted alkylamines** | 2 | 2,660.34 | R06AB | ATC |
| **79** | **fat emulsions; parenteral** | 3 | 2,649.06 | B05BA02 | ATC |

| | | | | | |
|---|---|---|---|---|---|
| **80** | **Local hemostatics** | 2 | 2,617.59 | B02BC | ATC |
| **81** | **combinations; topical (local hemostatics)** | 3 | 2,617.59 | B02BC30 | ATC |
| **82** | **Other irrigating solutions** | 2 | 2,580.11 | B05CX | ATC |
| **83** | **other irrigating solutions - combinations** | 3 | 2,580.11 | B05CX10 | ATC |
| **84** | **iodine/octylphenoxypolyglycolether; topical** | 3 | 2,551.23 | D08AG01 | ATC |
| **85** | **Iodine products** | 2 | 2,550.43 | D08AG | ATC |
| **86** | **Antidiarrheal microorganisms** | 2 | 2,523.42 | A07FA | ATC |
| **87** | **propyphenazone, combinations with psycholeptics; oral, otic, rectal, topical** | 3 | 2,512.79 | N02BB74 | ATC |
| **88** | **Other agents for treatment of hemorrhoids and anal fissures for topical use** | 2 | 2,499.51 | C05AX | ATC |
| **89** | **propyphenazone, combinations excl. psycholeptics; oral** | 3 | 2,497.52 | N02BB54 | ATC |
| **90** | **Other ophthalmologicals** | 2 | 2,484.39 | S01XA | ATC |
| **91** | **artificial tears and other indifferent preparations; ophthalmic** | 3 | 2,470.95 | S01XA20 | ATC |
| **92** | **bisacodyl, combinations; oral, rectal** | 3 | 2,429.55 | A06AB52 | ATC |
| **93** | **carbamide, combinations; topical** | 3 | 2,425.55 | D02AE51 | ATC |
| **94** | **Carbamide products** | 2 | 2,423.06 | D02AE | ATC |
| **95** | **glyceryl trinitrate, combinations; systemic** | 3 | 2,408.26 | C01DA52 | ATC |
| **96** | **Nicotinic acid and derivatives** | 2 | 2,405.55 | C10AD | ATC |
| **97** | **nicotinic acid, combinations; systemic** | 3 | 2,405.55 | C10AD52 | ATC |
| **98** | **lactic acid producing organisms, combinations; oral** | 3 | 2,398.19 | A07FA51 | ATC |
| **99** | **Aminoalkyl ethers** | 2 | 2,385.57 | R06AA | ATC |
| **100** | **folic acid, combinations; systemic** | 3 | 2,298.67 | B03BB51 | ATC |
| **101** | **Folic acid and derivatives** | 2 | 2,297.93 | B03BB | ATC |
| **102** | **Third-generation cephalosporins** | 2 | 2,272.98 | J01DD | ATC |
| **103** | **Other antiemetics** | 2 | 2,268.77 | A04AD | ATC |
| **104** | **macrogol, combinations; oral** | 3 | 2,267.95 | A06AD65 | ATC |
| **105** | **Corticosteroids for systemic use, combinations** | 2 | 2,258.86 | H02BX | ATC |
| **106** | **hydrocortisone, combinations; topical** | 3 | 2,255.00 | D07XA01 | ATC |
| **107** | **Caries prophylactic agents** | 2 | 2,191.77 | A01AA | ATC |
| **108** | **combinations; oral, local oral (caries prophylactic agents)** | 3 | 2,191.77 | A01AA30 | ATC |
| **109** | **medicinal charcoal, combinations; oral** | 3 | 2,188.76 | A07BA51 | ATC |
| **110** | **Charcoal preparations** | 2 | 2,188.19 | A07BA | ATC |
| **111** | **Drugs used in erectile dysfunction** | 2 | 2,187.74 | G04BE | ATC |
| **112** | **sodium fluoride, combinations; oral, local oral** | 3 | 2,165.23 | A01AA51 | ATC |
| **113** | **paracetamol, combinations with psycholeptics; systemic** | 3 | 2,147.00 | N02BE71 | ATC |
| **114** | **papaverine, combinations; systemic** | 3 | 2,137.94 | G04BE52 | ATC |
| **115** | **Diphenylpropylamine derivatives** | 2 | 2,100.51 | N02AC | ATC |
| **116** | **salicylamide, combinations with psycholeptics; systemic** | 3 | 2,099.90 | N02BA75 | ATC |
| **117** | **diphenhydramine, combinations; systemic** | 3 | 2,096.48 | R06AA52 | ATC |
| **118** | **chlorobutanol, combinations; systemic** | 3 | 2,063.30 | A04AD54 | ATC |
| **119** | **fluoride, combinations; systemic** | 3 | 2,058.69 | A12CD51 | ATC |
| **120** | **Fluoride** | 2 | 2,050.00 | A12CD | ATC |

| | | | | | |
|---|---|---|---|---|---|
| **121** | **phenacetin, combinations excl. psycholeptics; inhalant, oral, rectal** | 3 | 2,048.98 | N02BE53 | ATC |
| **122** | **iron, vitamin B12 and folic acid; systemic** | 3 | 2,045.95 | B03AE01 | ATC |
| **123** | **Propionic acid derivatives** | 2 | 2,036.34 | M01AE | ATC |
| **124** | **phenylpropanolamine, combinations; oral** | 3 | 2,012.97 | R01BA51 | ATC |
| **125** | **ergotamine, combinations excl. psycholeptics; oral** | 3 | 1,997.09 | N02CA52 | ATC |
| **126** | **dihydrocodeine and other non-opioid analgesics; systemic** | 3 | 1,989.69 | N02AJ03 | ATC |
| **127** | **codeine and other non-opioid analgesics; systemic** | 3 | 1,988.92 | N02AJ09 | ATC |
| **128** | **phenylephrine, combinations; oral** | 3 | 1,985.66 | R01BA53 | ATC |
| **129** | **dihydrocodeine, combinations; systemic** | 3 | 1,970.30 | N02AA58 | ATC |
| **130** | **brompheniramine, combinations; systemic** | 3 | 1,958.53 | R06AB51 | ATC |
| **131** | **Opioids in combination with non-opioid analgesics** | 2 | 1,939.17 | N02AJ | ATC |
| **132** | **codeine, combinations with psycholeptics; systemic** | 3 | 1,937.36 | N02AA79 | ATC |
| **133** | **Other nervous system drugs** | 2 | 1,935.71 | N07XX | ATC |
| **134** | **dextromethorphan, combinations; oral** | 3 | 1,935.38 | N07XX59 | ATC |
| **135** | **Belladonna and derivatives in combination with psycholeptics** | 2 | 1,921.09 | A03CB | ATC |
| **136** | **cascara, combinations; systemic** | 3 | 1,918.13 | A06AB57 | ATC |
| **137** | **Oxazol, thiazine, and triazine derivatives** | 2 | 1,915.52 | M03BB | ATC |
| **138** | **dextropropoxyphene, combinations excl. psycholeptics; systemic** | 3 | 1,908.34 | N02AC54 | ATC |
| **139** | **pseudoephedrine, combinations; oral** | 3 | 1,878.15 | R01BA52 | ATC |
| **140** | **Sympathomimetics** | 2 | 1,874.99 | R01BA | ATC |
| **141** | **metamizole sodium, combinations excl. psycholeptics; systemic** | 3 | 1,861.98 | N02BB52 | ATC |
| **142** | **Rauwolfia alkaloids** | 2 | 1,857.41 | C02AA | ATC |
| **143** | **dihydroergotamine, combinations; systemic** | 3 | 1,848.33 | N02CA51 | ATC |
| **144** | **theophylline, combinations excl. psycholeptics; systemic** | 3 | 1,837.54 | R03DA54 | ATC |
| **145** | **aluminium preparations; rectal, topical** | 3 | 1,837.21 | C05AX01 | ATC |
| **146** | **ibuprofen, combinations; systemic** | 3 | 1,827.74 | M01AE51 | ATC |
| **147** | **combinations of penicillins; systemic** | 3 | 1,826.42 | J01CR50 | ATC |
| **148** | **Combinations of penicillins, incl. beta-lactamase inhibitors** | 2 | 1,826.42 | J01CR | ATC |
| **149** | **aspirin** | 3 | 1,812.38 | 1191 | RxNorm |
| **150** | **aspirin Oral Product** | 2 | 1,800.83 | 1154069 | RxNorm |
| **151** | **acetylsalicylic acid; systemic, rectal** | 3 | 1,798.25 | N02BA01 | ATC |
| **152** | **aspirin Pill** | 2 | 1,797.82 | 1154070 | RxNorm |
| **153** | **analgesics and anesthetics - combinations; otic** | 3 | 1,797.10 | S02DA30 | ATC |
| **154** | **Analgesics and anesthetics** | 2 | 1,797.10 | S02DA | ATC |
| **155** | **platelet aggregation inhibitors excl. heparin - combinations** | 3 | 1,794.81 | B01AC30 | ATC |
| **156** | **other preparations, combinations; rectal** | 3 | 1,783.25 | C05AX03 | ATC |

| | | | | | |
|---|---|---|---|---|---|
| **157** | **pravastatin and acetylsalicylic acid; systemic** | 3 | 1,781.23 | C10BX02 | ATC |
| **158** | **Carbamic acid esters** | 2 | 1,780.52 | M03BA | ATC |
| **159** | **dipyrocetyl, combinations excl. psycholeptics** | 3 | 1,766.49 | N02BA59 | ATC |
| **160** | **Carbapenems** | 2 | 1,760.98 | J01DH | ATC |
| **161** | **gonadotropins - combinations; systemic** | 3 | 1,759.83 | G03GA30 | ATC |
| **162** | **Gonadotropins** | 2 | 1,759.83 | G03GA | ATC |
| **163** | **cefoperazone and beta-lactamase inhibitor; parenteral** | 3 | 1,759.68 | J01DD62 | ATC |
| **164** | **sodium chloride; oral** | 3 | 1,755.85 | A12CA01 | ATC |
| **165** | **aspirin 81 MG** | 4 | 1,751.41 | 315431 | RxNorm |
| **166** | **Sodium** | 2 | 1,744.21 | A12CA | ATC |
| **167** | **sodium chloride** | 3 | 1,744.21 | 9863 | RxNorm |
| **168** | **imipenem and cilastatin; parenteral** | 3 | 1,744.21 | J01DH51 | ATC |
| **169** | **ceftriaxone, combinations; systemic** | 3 | 1,744.21 | J01DD54 | ATC |
| **170** | **Corticosteroids, weak, combinations with antibiotics** | 2 | 1,723.67 | D07CA | ATC |
| **171** | **nifedipine, combinations; systemic** | 3 | 1,719.64 | C08CA55 | ATC |
| **172** | **sodium chloride 9 MG/ML** | 4 | 1,708.98 | 1661411 | RxNorm |
| **173** | **tetracaine, combinations; parenteral** | 3 | 1,706.44 | N01BA53 | ATC |
| **174** | **atropine and psycholeptics; systemic** | 3 | 1,688.28 | A03CB03 | ATC |
| **175** | **sodium chloride; irrigating solution** | 3 | 1,683.22 | B05CB01 | ATC |
| **176** | **Opioid anesthetics** | 2 | 1,678.06 | N01AH | ATC |
| **177** | **phenacetin, combinations with psycholeptics; inhalant, oral, rectal** | 3 | 1,671.55 | N02BE73 | ATC |
| **178** | **sodium chloride; parenteral** | 3 | 1,667.55 | B05XA03 | ATC |
| **179** | **Immunoglobulins, normal human** | 2 | 1,666.67 | J06BA | ATC |
| **180** | **immunoglobulins, normal human, for extravascular adm.; systemic** | 3 | 1,666.67 | J06BA01 | ATC |
| **181** | **immunoglobulins, normal human, for intravascular adm.; systemic** | 3 | 1,666.67 | J06BA02 | ATC |
| **182** | **immunoglobulin G** | 4 | 1,666.67 | 5666 | RxNorm |
| **183** | **Antiinfectives** | 2 | 1,666.15 | S02AA | ATC |
| **184** | **sodium picosulfate, combinations; oral, topical** | 3 | 1,658.16 | A06AB58 | ATC |
| **185** | **bupivacaine, combinations; parenteral, transdermal** | 3 | 1,655.36 | N01BB51 | ATC |
| **188** | **Platelet aggregation inhibitors excl. heparin** | 2 | 1,630.34 | B01AC | ATC |
| **189** | **fentanyl, combinations; systemic** | 3 | 1,617.87 | N01AH51 | ATC |
| **190** | **codeine and paracetamol; systemic** | 3 | 1,616.30 | N02AJ06 | ATC |
| **191** | **acetylsalicylic acid, combinations with proton pump inhibitors; systemic** | 3 | 1,608.95 | B01AC56 | ATC |
| **192** | **ergotamine, combinations with psycholeptics; oral** | 3 | 1,604.84 | N02CA72 | ATC |
| **193** | **theophylline, combinations with psycholeptics; systemic** | 3 | 1,587.29 | R03DA74 | ATC |
| **194** | **antibiotics in combination with other drugs; ophthalmic** | 3 | 1,583.71 | S01AA20 | ATC |
| **195** | **Antibiotics** | 2 | 1,580.68 | S01AA | ATC |
| **196** | **Beta blocking agents, other combinations** | 2 | 1,558.59 | C07FX | ATC |

| | | | | | |
|---|---|---|---|---|---|
| **197** | **Dihydropyridine derivatives** | 2 | 1,556.36 | C08CA | ATC |
| **198** | **dexamethasone and antiinfectives; otic** | 3 | 1,551.41 | S02CA06 | ATC |
| **199** | **Zinc bandages** | 2 | 1,529.45 | D09AB | ATC |
| **200** | **zinc bandage without supplements; topical** | 3 | 1,527.98 | D09AB01 | ATC |
| **201** | **zinc bandage with supplements; topical** | 3 | 1,527.98 | D09AB02 | ATC |
| **202** | **dexamethasone and antiinfectives; ophthalmic, otic (ophth. and otological prep., corticosteroids and antiinfectives in comb.)** | 3 | 1,526.16 | S03CA01 | ATC |
| **203** | **zinc preparations; rectal, topical** | 3 | 1,524.22 | C05AX04 | ATC |
| **204** | **Enemas** | 2 | 1,517.60 | A06AG | ATC |
| **205** | **HMG CoA reductase inhibitors, other combinations** | 2 | 1,514.59 | C10BX | ATC |
| **206** | **antiinfectives, combinations; otic** | 3 | 1,513.44 | S02AA30 | ATC |
| **207** | **carisoprodol, combinations excl. psycholeptics; oral** | 3 | 1,445.27 | M03BA52 | ATC |
| **208** | **lactulose, combinations; oral** | 3 | 1,435.94 | A06AD61 | ATC |
| **209** | **aminophenazone, combinations excl. psycholeptics; systemic** | 3 | 1,408.04 | N02BB53 | ATC |
| **210** | **phenazone, combinations with psycholeptics; systemic** | 3 | 1,407.20 | N02BB71 | ATC |
| **211** | **butylscopolamine and analgesics; systemic** | 3 | 1,402.41 | A03DB04 | ATC |
| **212** | **Belladonna and derivatives in combination with analgesics** | 2 | 1,402.41 | A03DB | ATC |
| **213** | **tiemonium iodide and analgesics; oral** | 3 | 1,391.55 | A03DA07 | ATC |
| **214** | **Synthetic anticholinergic agents in combination with analgesics** | 2 | 1,391.30 | A03DA | ATC |
| **215** | **trospium and analgesics; systemic** | 3 | 1,389.05 | A03DA06 | ATC |
| **216** | **bevonium and analgesics** | 3 | 1,389.05 | A03DA03 | ATC |
| **217** | **tropenzilone and analgesics** | 3 | 1,389.05 | A03DA01 | ATC |
| **218** | **ciclonium and analgesics; systemic** | 3 | 1,388.79 | A03DA04 | ATC |
| **219** | **pitofenone and analgesics; systemic** | 3 | 1,388.79 | A03DA02 | ATC |
| **220** | **camylofin and analgesics; systemic** | 3 | 1,387.81 | A03DA05 | ATC |
| **221** | **chlormezanone, combinations excl. psycholeptics; oral, rectal** | 3 | 1,383.26 | M03BB52 | ATC |
| **222** | **tramadol and other non-opioid analgesics; systemic** | 3 | 1,381.48 | N02AJ15 | ATC |
| **223** | **acetylsalicylic acid, combinations with psycholeptics; systemic** | 3 | 1,366.26 | N02BA71 | ATC |
| **224** | **methylprednisolone** | 3 | 1,356.98 | 6902 | RxNorm |
| **225** | **rauwolfia alkaloids, whole root; systemic** | 3 | 1,355.68 | C02AA04 | ATC |
| **228** | **methylprednisolone Injectable Product** | 2 | 1,344.39 | 1163486 | RxNorm |
| **230** | **Corticosteroids and antiinfectives in combination** | 2 | 1,336.44 | S02CA | ATC |
| **231** | **Corticosteroids and antiinfectives in combination** | 2 | 1,332.64 | S03CA | ATC |
| **232** | **Other agents for local oral treatment** | 2 | 1,332.29 | A01AD | ATC |
| **233** | **diphenylpyraline, combinations; systemic** | 3 | 1,327.14 | R06AA57 | ATC |
| **234** | **doxylamine, combinations; systemic** | 3 | 1,320.18 | R06AA59 | ATC |
| **235** | **Belladonna alkaloids, semisynthetic, quaternary ammonium compounds** | 2 | 1,297.32 | A03BB | ATC |

| | | | | | |
|---|---|---|---|---|---|
| **236** | **methylprednisolone; systemic** | 3 | 1,289.98 | H02AB04 | ATC |
| **237** | **nikethamide, combinations; systemic** | 3 | 1,286.55 | R07AB52 | ATC |
| **238** | **salicylamide, combinations excl. psycholeptics; systemic** | 3 | 1,280.04 | N02BA55 | ATC |
| **239** | **Ethers, chemically close to antihistamines** | 2 | 1,278.84 | M03BC | ATC |
| **240** | **orphenadrine, combinations; systemic** | 3 | 1,278.84 | M03BC51 | ATC |
| **242** | **homatropine methylbromide; systemic** | 3 | 1,273.75 | A03BB06 | ATC |
| **243** | **glucose; systemic (carbohydrates)** | 3 | 1,267.96 | V06DC01 | ATC |
| **244** | **Tests for diabetes** | 2 | 1,259.89 | V04CA | ATC |
| **245** | **Carbohydrates** | 2 | 1,259.89 | V06DC | ATC |
| **246** | **glucose** | 3 | 1,259.89 | 4850 | RxNorm |
| **247** | **glucose; irrigating solution** | 3 | 1,253.22 | B05CX01 | ATC |
| **248** | **glucose 50 MG/ML** | 4 | 1,253.22 | 315789 | RxNorm |
| **249** | **Sulfonamides and potassium in combination** | 2 | 1,246.88 | C03CB | ATC |
| **250** | **Piperazine derivatives** | 2 | 1,231.11 | R06AE | ATC |
| **251** | **methocarbamol, combinations excl. psycholeptics; systemic** | 3 | 1,230.33 | M03BA53 | ATC |
| **252** | **dihydroergocristine, combinations; systemic** | 3 | 1,199.63 | C04AE54 | ATC |
| **253** | **methylprednisolone, combinations; systemic** | 3 | 1,192.31 | H02BX01 | ATC |
| **254** | **phenazone, combinations excl. psycholeptics; oral** | 3 | 1,169.06 | N02BB51 | ATC |
| **255** | **codeine and acetylsalicylic acid; systemic** | 3 | 1,163.89 | N02AJ07 | ATC |
| **256** | **diprophylline, combinations; systemic** | 3 | 1,162.55 | R03DA51 | ATC |
| **257** | **Ergot alkaloids** | 2 | 1,126.98 | C04AE | ATC |
| **258** | **ethenzamide, combinations with psycholeptics; oral, rectal, topical** | 3 | 1,109.24 | N02BA77 | ATC |
| **259** | **Opium derivatives and expectorants** | 2 | 1,098.91 | R05FA | ATC |
| **260** | **opium derivatives and expectorants; systemic** | 3 | 1,091.98 | R05FA02 | ATC |
| **261** | **quinine, combinations with psycholeptics; systemic** | 3 | 1,091.73 | M09AA72 | ATC |
| **262** | **Quinine and derivatives** | 2 | 1,091.73 | M09AA | ATC |
| **263** | **Glucocorticoids** | 2 | 1,089.01 | H02AB | ATC |
| **264** | **fludrocortisone and antiinfectives; otic** | 3 | 1,087.97 | S02CA07 | ATC |
| **265** | **fludrocortisone and antiinfectives; ophthalmic, otic (ophth. and otological prep., corticosteroids and antiinfectives in comb.)** | 3 | 1,087.97 | S03CA05 | ATC |
| **266** | **Corticosteroids, weak (group I)** | 2 | 1,084.81 | D07AA | ATC |
| **267** | **hydrocortisone and antiinfectives; otic** | 3 | 1,077.92 | S02CA03 | ATC |
| **268** | **Iron in combination with folic acid** | 2 | 1,070.17 | B03AD | ATC |
| **269** | **Other cough suppressants and expectorants** | 2 | 1,067.11 | R05FB | ATC |
| **270** | **Phenothiazine derivatives** | 2 | 1,066.11 | R06AD | ATC |
| **271** | **promethazine, combinations; systemic** | 3 | 1,063.03 | R06AD52 | ATC |
| **272** | **cough suppressants and expectorants** | 3 | 1,061.89 | R05FB02 | ATC |
| **273** | **hydrocortisone and antiinfectives; ophthalmic, otic (ophth. and otological prep., corticosteroids and antiinfectives in comb.)** | 3 | 1,057.62 | S03CA04 | ATC |
| **274** | **hydroxyethylpromethazine, combinations; systemic** | 3 | 1,054.08 | R06AD55 | ATC |

| | | | | | |
|---|---|---|---|---|---|
| **275** | **oil; rectal** | 3 | 1,053.48 | A06AG06 | ATC |
| **276** | **ferrous sulfate, combinations; systemic** | 3 | 1,014.19 | B03AD03 | ATC |
| **277** | **scopolamine, combinations; systemic** | 3 | 1,007.07 | A04AD51 | ATC |
| **278** | **famotidine** | 3 | 1,002.79 | 4278 | RxNorm |
| **279** | **cyclizine, combinations; systemic** | 3 | 1,002.29 | R06AE53 | ATC |
| **280** | **bismuth preparations, combinations; rectal, topical** | 3 | 992.36 | C05AX02 | ATC |
| **281** | **Ascorbic acid (vitamin C), combinations** | 2 | 984.62 | A11GB | ATC |
| **282** | **chlorzoxazone, combinations excl. psycholeptics; oral** | 3 | 983.11 | M03BB53 | ATC |
| **283** | **famotidine; systemic** | 3 | 978.40 | A02BA03 | ATC |
| **284** | **sodium chloride Injectable Product** | 2 | 975.10 | 1159317 | RxNorm |
| **285** | **Proton pump inhibitors** | 2 | 972.88 | A02BC | ATC |
| **286** | **Phenylpiperidine derivatives** | 2 | 965.93 | N02AB | ATC |
| **287** | **combinations of different antibiotics; ophthalmic** | 3 | 953.61 | S01AA30 | ATC |
| **288** | **ferrous fumarate, combinations; systemic (iron in comb. with folic acid)** | 3 | 938.95 | B03AD02 | ATC |
| **289** | **fumaric acid derivatives, combinations; systemic** | 3 | 934.45 | D05BX51 | ATC |
| **290** | **Other antipsoriatics for systemic use** | 2 | 934.45 | D05BX | ATC |
| **291** | **prednisone** | 3 | 915.05 | 8640 | RxNorm |
| **292** | **prednisone; oral** | 3 | 915.05 | H02AB07 | ATC |
| **293** | **prednisone Oral Product** | 2 | 915.05 | 1161705 | RxNorm |
| **294** | **hydroxocobalamin, combinations; systemic** | 3 | 903.93 | B03BA53 | ATC |
| **295** | **prednisone Oral Tablet** | 3 | 902.50 | 373585 | RxNorm |
| **296** | **prednisone Pill** | 2 | 902.16 | 1161706 | RxNorm |
| **297** | **antiinfectives - combinations; irrigating solution** | 3 | 896.55 | B05CA10 | ATC |
| **298** | **Antiinfectives** | 2 | 896.55 | B05CA | ATC |
| **299** | **Corticosteroids acting locally** | 2 | 886.08 | A07EA | ATC |
| **300** | **triamcinolone and antiinfectives; otic** | 3 | 880.65 | S02CA04 | ATC |
| **301** | **hydrochlorothiazide, combinations; oral** | 3 | 874.59 | C03AX01 | ATC |
| **302** | **Thiazides, combinations with other drugs** | 2 | 874.11 | C03AX | ATC |
| **303** | **fluocinolone acetonide and antiinfectives; otic** | 3 | 855.42 | S02CA05 | ATC |
| **304** | **flumetasone and antiinfectives; otic** | 3 | 854.85 | S02CA02 | ATC |
| **305** | **hydrocortisone and antibiotics; topical** | 3 | 854.82 | D07CA01 | ATC |
| **306** | **betamethasone and antiinfectives; ophthalmic, otic (ophth. and otological prep., corticosteroids and antiinfectives in comb.)** | 3 | 853.54 | S03CA06 | ATC |
| **307** | **triamcinolone and antibiotics; topical** | 3 | 848.79 | D07CB01 | ATC |
| **308** | **Antiinflammatory/antirheumatic agents in combination with corticosteroids** | 2 | 846.47 | M01BA | ATC |
| **309** | **Adrenergics in combination with corticosteroids or other drugs, excl. anticholinergics** | 2 | 844.19 | R03AK | ATC |

| | | | | | |
|---|---|---|---|---|---|
| **310** | **Respiratory stimulants** | 2 | 839.32 | R07AB | ATC |
| **311** | **H2-receptor antagonists** | 2 | 829.21 | A02BA | ATC |
| **312** | **dexamethasone, combinations; topical** | 3 | 828.85 | D07XB05 | ATC |
| **313** | **Thiazides and potassium in combination** | 2 | 796.27 | C03AB | ATC |
| **314** | **potassium chloride** | 3 | 790.16 | 8591 | RxNorm |
| **315** | **Sulfonamides and potassium in combination** | 2 | 789.21 | C03BB | ATC |
| **316** | **potassium (different salts in combination)** | 3 | 787.92 | A12BA30 | ATC |
| **317** | **ascorbic acid (vit C) and calcium; systemic** | 3 | 786.49 | A11GB01 | ATC |
| **318** | **theobromine, combinations; systemic** | 3 | 774.00 | R03DA57 | ATC |
| **319** | **ketoprofen, combinations; systemic** | 3 | 745.40 | M01AE53 | ATC |
| **320** | **benzethonium chloride, combinations; topical** | 3 | 736.35 | D08AJ58 | ATC |
| **321** | **Quaternary ammonium compounds** | 2 | 736.35 | D08AJ | ATC |
| **322** | **Corticosteroids, moderately potent, combinations with antibiotics** | 2 | 728.29 | D07CB | ATC |
| **323** | **pentaerithrityl tetranitrate, combinations; oral** | 3 | 726.42 | C01DA55 | ATC |
| **324** | **Sympathomimetics used as decongestants** | 2 | 721.33 | S01GA | ATC |
| **326** | **methaqualone, combinations; oral, rectal, topical** | 3 | 701.00 | N05CX02 | ATC |
| **327** | **diclofenac, combinations; oral** | 3 | 695.28 | M01AB55 | ATC |
| **328** | **Corticosteroids, weak, combinations with antiseptics** | 2 | 693.32 | D07BA | ATC |
| **329** | **combinations of tetracyclines; systemic** | 3 | 685.96 | J01AA20 | ATC |
| **330** | **Antihistamines for topical use** | 2 | 685.93 | D04AA | ATC |
| **331** | **Tetracyclines** | 2 | 685.76 | J01AA | ATC |
| **332** | **lansoprazole, combinations; systemic** | 3 | 683.96 | A02BC53 | ATC |
| **333** | **Rauwolfia alkaloids and diuretics in combination** | 2 | 678.83 | C02LA | ATC |
| **334** | **diphenhydramine** | 3 | 674.23 | 3498 | RxNorm |
| **335** | **Ethers chemically close to antihistamines** | 2 | 674.23 | N04AB | ATC |
| **336** | **Other agents against amoebiasis and other protozoal diseases** | 2 | 671.96 | P01AX | ATC |
| **337** | **Sulfonamides** | 2 | 671.73 | G01AE | ATC |
| **338** | **emetine, combinations; systemic** | 3 | 671.65 | P01AX52 | ATC |
| **339** | **ceftriaxone** | 3 | 664.50 | 2193 | RxNorm |
| **340** | **bifonazole, combinations; topical** | 3 | 662.95 | D01AC60 | ATC |
| **341** | **famotidine Oral Product** | 2 | 658.56 | 1159021 | RxNorm |
| **342** | **prednisolone, combinations; topical** | 3 | 656.48 | D07XA02 | ATC |
| **343** | **methylpentynol, combinations; oral, rectal** | 3 | 656.33 | N05CX03 | ATC |
| **344** | **ceftriaxone; parenteral** | 3 | 656.28 | J01DD04 | ATC |
| **345** | **ceftriaxone Injectable Product** | 2 | 656.28 | 1152108 | RxNorm |
| **346** | **Corticosteroids, very potent, combinations with antibiotics** | 2 | 653.73 | D07CD | ATC |
| **347** | **clobetasol and antibiotics; topical** | 3 | 653.73 | D07CD01 | ATC |
| **348** | **Hydrazinophthalazine derivatives and diuretics** | 2 | 652.97 | C02LG | ATC |

| | | | | | |
|---|---|---|---|---|---|
| **349** | **prednisolone and antibiotics; topical** | 3 | 651.15 | D07CA03 | ATC |
| **350** | **fluocinolone acetonide and antibiotics; topical** | 3 | 648.66 | D07CC02 | ATC |
| **351** | **methylprednisolone and antibiotics; topical** | 3 | 648.59 | D07CA02 | ATC |
| **352** | **beclometasone and antibiotics; topical** | 3 | 647.72 | D07CC04 | ATC |
| **353** | **fluprednidene and antibiotics; topical** | 3 | 647.72 | D07CB02 | ATC |
| **354** | **fluocortolone and antibiotics; topical** | 3 | 647.72 | D07CC06 | ATC |
| **355** | **flumetasone and antibiotics; topical** | 3 | 647.72 | D07CB05 | ATC |
| **356** | **fluorometholone and antibiotics; topical** | 3 | 647.68 | D07CB03 | ATC |
| **357** | **fludroxycortide and antibiotics; topical** | 3 | 647.68 | D07CC03 | ATC |
| **358** | **dihydrocodeine and paracetamol; systemic** | 3 | 647.06 | N02AJ01 | ATC |
| **359** | **fluocinonide and antibiotics; topical** | 3 | 646.86 | D07CC05 | ATC |
| **360** | **Biguanides and amidines** | 2 | 645.96 | D08AC | ATC |
| **361** | **Acetic acid derivatives and related substances** | 2 | 644.25 | M01AB | ATC |
| **362** | **pethidine, combinations excl. psycholeptics; systemic** | 3 | 639.65 | N02AB52 | ATC |
| **363** | **chlorhexidine, combinations; topical** | 3 | 638.34 | D08AC52 | ATC |
| **364** | **dexamethasone and antibiotics; topical** | 3 | 634.82 | D07CB04 | ATC |
| **365** | **prednisolone and antiinfectives; otic** | 3 | 631.44 | S02CA01 | ATC |
| **366** | **betamethasone and antibiotics; topical** | 3 | 630.84 | D07CC01 | ATC |
| **367** | **reserpine and diuretics, combinations with psycholeptics; systemic** | 3 | 630.81 | C02LA71 | ATC |
| **368** | **potassium chloride; parenteral** | 3 | 630.70 | B05XA01 | ATC |
| **369** | **Corticosteroids, potent, combinations with antibiotics** | 2 | 630.59 | D07CC | ATC |
| **370** | **prednisolone and antiinfectives; ophthalmic, otic (ophth. and otological prep., corticosteroids and antiinfectives in comb.)** | 3 | 630.17 | S03CA02 | ATC |
| **371** | **picodralazine and diuretics, combinations with psycholeptics** | 3 | 618.71 | C02LG73 | ATC |
| **372** | **Tests for gastric secretion** | 2 | 618.15 | V04CG | ATC |
| **373** | **caffeine and sodium benzoate; systemic** | 3 | 616.58 | V04CG30 | ATC |
| **374** | **combinations; topical (antifungals for topical use)** | 3 | 614.21 | D01AE20 | ATC |
| **375** | **Other antifungals for topical use** | 2 | 614.21 | D01AE | ATC |
| **376** | **combinations of sulfonamides; oral, topical** | 3 | 606.97 | G01AE10 | ATC |
| **377** | **buclizine, combinations; systemic** | 3 | 605.17 | R06AE51 | ATC |
| **378** | **ACE inhibitors and diuretics** | 2 | 605.02 | C09BA | ATC |
| **379** | **acetaminophen** | 3 | 602.85 | 161 | RxNorm |
| **380** | **thiocolchicoside, combinations; systemic** | 3 | 602.85 | M03BX55 | ATC |
| **381** | **codeine and ibuprofen; systemic** | 3 | 602.85 | N02AJ08 | ATC |
| **382** | **paracetamol; systemic, rectal** | 3 | 600.33 | N02BE01 | ATC |
| **383** | **Other centrally acting agents** | 2 | 598.35 | M03BX | ATC |
| **385** | **dexbrompheniramine, combinations; systemic** | 3 | 594.75 | R06AB56 | ATC |
| **386** | **dexchlorpheniramine, combinations; systemic** | 3 | 594.75 | R06AB52 | ATC |
| **387** | **atenolol and other diuretics, combinations; systemic** | 3 | 588.11 | C07CB53 | ATC |
| **388** | **lisinopril and diuretics; oral** | 3 | 584.80 | C09BA03 | ATC |
| **389** | **Corticosteroids, potent (group III)** | 2 | 584.76 | D07AC | ATC |
| **390** | **Beta blocking agents, selective, and other diuretics** | 2 | 583.58 | C07CB | ATC |

| | | | | | |
|---|---|---|---|---|---|
| **391** | **Beta blocking agents, non-selective, and other diuretics** | 2 | 582.37 | C07CA | ATC |
| **392** | **isoprenaline and other drugs for obstructive airway diseases; inhalant** | 3 | 579.13 | R03AK02 | ATC |
| **393** | **enalapril and diuretics; systemic** | 3 | 575.95 | C09BA02 | ATC |
| **394** | **Alkaloids, excl. rauwolfia, in combination with diuretics** | 2 | 575.31 | C02LK | ATC |
| **395** | **veratrum and diuretics; systemic** | 3 | 575.31 | C02LK01 | ATC |
| **396** | **irbesartan and diuretics; systemic** | 3 | 572.94 | C09DA04 | ATC |
| **397** | **azilsartan medoxomil and diuretics; oral** | 3 | 572.94 | C09DA09 | ATC |
| **398** | **pinacidil and diuretics; oral** | 3 | 572.94 | C02LX01 | ATC |
| **399** | **Other antihypertensives and diuretics** | 2 | 572.94 | C02LX | ATC |
| **400** | **benazepril and diuretics; oral** | 3 | 572.94 | C09BA07 | ATC |
| **401** | **fosinopril and diuretics; oral** | 3 | 572.94 | C09BA09 | ATC |
| **402** | **delapril and diuretics; oral** | 3 | 572.94 | C09BA12 | ATC |
| **403** | **moexipril and diuretics; oral** | 3 | 572.94 | C09BA13 | ATC |
| **404** | **methyldopa (levorotatory) and diuretics; systemic** | 3 | 572.94 | C02LB01 | ATC |
| **405** | **fimasartan and diuretics; oral** | 3 | 572.94 | C09DA10 | ATC |
| **406** | **Methyldopa and diuretics in combination** | 2 | 572.94 | C02LB | ATC |
| **407** | **quinapril and diuretics; systemic** | 3 | 572.94 | C09BA06 | ATC |
| **408** | **syrosingopine and diuretics** | 3 | 572.94 | C02LA09 | ATC |
| **409** | **moxonidine and diuretics; oral** | 3 | 572.94 | C02LC05 | ATC |
| **410** | **eprosartan and diuretics; oral** | 3 | 572.94 | C09DA02 | ATC |
| **411** | **zofenopril and diuretics; oral** | 3 | 572.94 | C09BA15 | ATC |
| **412** | **bietaserpine and diuretics; oral** | 3 | 572.94 | C02LA07 | ATC |
| **413** | **picodralazine and diuretics** | 3 | 572.94 | C02LG03 | ATC |
| **414** | **pargyline and diuretics; oral** | 3 | 572.94 | C02LL01 | ATC |
| **415** | **methoserpidine and diuretics; oral** | 3 | 572.94 | C02LA04 | ATC |
| **416** | **Guanidine derivatives and diuretics** | 2 | 572.94 | C02LF | ATC |
| **417** | **MAO inhibitors and diuretics** | 2 | 572.94 | C02LL | ATC |
| **418** | **deserpidine and diuretics; oral** | 3 | 572.94 | C02LA03 | ATC |
| **419** | **cilazapril and diuretics; oral** | 3 | 572.94 | C09BA08 | ATC |
| **420** | **guanethidine and diuretics; systemic** | 3 | 572.94 | C02LF01 | ATC |
| **421** | **candesartan and diuretics; oral** | 3 | 572.72 | C09DA06 | ATC |
| **422** | **nifedipine and diuretics; systemic** | 3 | 572.72 | C08GA01 | ATC |
| **423** | **captopril and diuretics; systemic** | 3 | 571.20 | C09BA01 | ATC |
| **425** | **dihydralazine and diuretics; systemic** | 3 | 570.55 | C02LG01 | ATC |
| **428** | **Sulfonamides, plain** | 2 | 567.77 | C03CA | ATC |
| **429** | **Calcium channel blockers and diuretics** | 2 | 564.01 | C08GA | ATC |
| **430** | **prazosin and diuretics; oral, topical** | 3 | 563.57 | C02LE01 | ATC |
| **431** | **Alpha-adrenoreceptor antagonists and diuretics** | 2 | 563.57 | C02LE | ATC |
| **432** | **perindopril and diuretics; systemic** | 3 | 560.99 | C09BA04 | ATC |
| **433** | **ramipril and diuretics; oral** | 3 | 560.99 | C09BA05 | ATC |
| **434** | **olmesartan medoxomil and diuretics; oral** | 3 | 560.99 | C09DA08 | ATC |
| **435** | **telmisartan and diuretics; oral** | 3 | 560.99 | C09DA07 | ATC |
| **436** | **valsartan and diuretics; oral** | 3 | 560.80 | C09DA03 | ATC |
| **437** | **sodium chloride Injectable Solution** | 3 | 559.51 | 373901 | RxNorm |

| | | | | | |
|---|---|---|---|---|---|
| **438** | **hydralazine and diuretics; systemic** | 3 | 559.20 | C02LG02 | ATC |
| **439** | **penbutolol and other diuretics; oral** | 3 | 557.52 | C07CA23 | ATC |
| **440** | **furosemide; systemic** | 3 | 557.52 | C03CA01 | ATC |
| **441** | **furosemide** | 3 | 557.52 | 4603 | RxNorm |
| **442** | **losartan and diuretics; oral** | 3 | 555.31 | C09DA01 | ATC |
| **443** | **rauwolfia alkaloids, whole root and diuretics; systemic** | 3 | 555.17 | C02LA08 | ATC |
| **444** | **combination of rauwolfia alkaloids and diuretics incl. other combinations; systemic** | 3 | 555.17 | C02LA50 | ATC |
| **445** | **Angiotensin II receptor blockers (ARBs) and diuretics** | 2 | 554.62 | C09DA | ATC |
| **446** | **amlodipine and diuretics; oral** | 3 | 551.46 | C08GA02 | ATC |
| **447** | **High-ceiling diuretics and potassium-sparing agents** | 2 | 546.82 | C03EB | ATC |
| **448** | **sodium chloride 9 MG/ML Injectable Solution** | 3 | 540.26 | 313002 | RxNorm |
| **449** | **Imidazole and triazole derivatives** | 2 | 531.69 | D01AC | ATC |
| **450** | **reserpine and diuretics; systemic** | 3 | 529.22 | C02LA01 | ATC |
| **451** | **naphazoline, combinations; ophthalmic** | 3 | 523.73 | S01GA51 | ATC |
| **452** | **Corticosteroids and antiinfectives in combination** | 2 | 521.95 | S01CA | ATC |
| **453** | **reserpine and diuretics, combinations with other drugs; systemic** | 3 | 517.01 | C02LA51 | ATC |
| **454** | **Imidazoline receptor agonists in combination with diuretics** | 2 | 510.76 | C02LC | ATC |
| **455** | **homatropine methylbromide and psycholeptics; systemic** | 3 | 505.51 | A03CB04 | ATC |
| **456** | **various combinations; topical (anti-acne prep.)** | 3 | 503.62 | D10AX30 | ATC |
| **457** | **clonidine and diuretics, combinations with other drugs; systemic** | 3 | 503.20 | C02LC51 | ATC |
| **458** | **clonidine and diuretics; systemic** | 3 | 503.20 | C02LC01 | ATC |
| **459** | **Other anti-acne preparations for topical use** | 2 | 502.40 | D10AX | ATC |
| **460** | **Corticosteroids** | 2 | 497.67 | R01AD | ATC |
| **461** | **rescinnamine and diuretics; systemic** | 3 | 496.40 | C02LA02 | ATC |
| **462** | **combinations; rectal** | 3 | 485.79 | A06AG20 | ATC |
| **463** | **prilocaine, combinations; parenteral** | 3 | 475.58 | N01BB54 | ATC |
| **464** | **vancomycin** | 3 | 473.69 | 11124 | RxNorm |
| **465** | **Glycopeptide antibacterials** | 2 | 472.76 | J01XA | ATC |
| **470** | **diphenhydramine hydrochloride 50 MG/ML** | 4 | 468.25 | 1049288 | RxNorm |
| **471** | **diphenhydramine Injectable Product** | 2 | 468.01 | 1158446 | RxNorm |
| **472** | **etanautine; systemic** | 3 | 465.69 | N04AB01 | ATC |
| **473** | **vancomycin; parenteral** | 3 | 454.74 | J01XA01 | ATC |
| **474** | **furosemide Injectable Product** | 2 | 446.27 | 1162712 | RxNorm |
| **477** | **furosemide 10 MG/ML** | 4 | 428.08 | 317376 | RxNorm |

| | | | | | |
|---|---|---|---|---|---|
| **478** | **Local anesthetics** | 2 | 419.09 | S01HA | ATC |
| **479** | **combinations; ophthalmic (local anesthetics)** | 3 | 419.09 | S01HA30 | ATC |
| **480** | **Corticosteroids, potent, other combinations** | 2 | 418.12 | D07XC | ATC |
| **482** | **clemastine, combinations; systemic** | 3 | 412.35 | R06AA54 | ATC |
| **483** | **epinephrine and other drugs for obstructive airway diseases; inhalant** | 3 | 405.69 | R03AK01 | ATC |
| **484** | **Softeners, emollients** | 2 | 404.74 | A06AA | ATC |
| **486** | **vancomycin Injectable Product** | 2 | 392.52 | 1160499 | RxNorm |
| **488** | **glucose / sodium chloride Injection** | 3 | 383.65 | 1794561 | RxNorm |
| **491** | **glucose / sodium chloride Injectable Product** | 2 | 382.87 | 1165817 | RxNorm |
| **492** | **liquid paraffin, combinations; topical** | 3 | 377.47 | A06AA51 | ATC |
| **493** | **choline theophyllinate and adrenergics; systemic** | 3 | 375.23 | R03DB02 | ATC |
| **494** | **diprophylline and adrenergics; systemic** | 3 | 375.23 | R03DB01 | ATC |
| **495** | **etamiphylline and adrenergics; systemic** | 3 | 375.23 | R03DB06 | ATC |
| **496** | **aminophylline and adrenergics; systemic** | 3 | 375.23 | R03DB05 | ATC |
| **497** | **theophylline and adrenergics; systemic** | 3 | 375.23 | R03DB04 | ATC |
| **498** | **proxyphylline and adrenergics; systemic** | 3 | 375.23 | R03DB03 | ATC |
| **499** | **Xanthines and adrenergics** | 2 | 375.01 | R03DB | ATC |
| **500** | **reserpine, combinations; systemic** | 3 | 373.20 | C02AA52 | ATC |
| **501** | **bietaserpine, combinations; systemic** | 3 | 373.20 | C02AA57 | ATC |
| **502** | **1000 ML glucose 50 MG/ML / potassium chloride 0.02 MEQ/ML / sodium chloride 9 MG/ML Injection** | 3 | 373.05 | 615107 | RxNorm |
| **504** | **Other drugs for peptic ulcer and gastro-oesophageal reflux disease (GORD)** | 2 | 368.38 | A02BX | ATC |
| **505** | **500 ML glucose 50 MG/ML / sodium chloride 9 MG/ML Injection** | 3 | 367.36 | 1795344 | RxNorm |
| **506** | **Bioflavonoids** | 2 | 364.00 | C05CA | ATC |
| **507** | **ceftriaxone Injection** | 3 | 363.54 | 1664992 | RxNorm |
| **510** | **mepivacaine, combinations; parenteral** | 3 | 362.47 | N01BB53 | ATC |
| **511** | **Multivitamins with minerals** | 2 | 361.95 | A11AA | ATC |
| **512** | **prednisolone; systemic** | 3 | 360.81 | H02AB06 | ATC |
| **513** | **hyoscyamine and psycholeptics; systemic** | 3 | 360.30 | A03CB31 | ATC |
| **515** | **prednisolone Oral Product** | 2 | 359.03 | 1165759 | RxNorm |
| **516** | **prednisolone** | 3 | 358.75 | 8638 | RxNorm |
| **517** | **acetaminophen Oral Product** | 2 | 358.08 | 1152842 | RxNorm |
| **518** | **Corticosteroids, combinations for treatment of acne** | 2 | 357.02 | D10AA | ATC |
| **519** | **adrenergic and dopaminergic agents - combinations** | 3 | 356.63 | C01CA30 | ATC |
| **520** | **glucose Injectable Product** | 2 | 356.49 | 1165819 | RxNorm |

| | | | | | |
|---|---|---|---|---|---|
| **521** | **Adrenergic and dopaminergic agents** | 2 | 356.18 | C01CA | ATC |
| **522** | **prednisolone Oral Solution** | 3 | 354.24 | 373575 | RxNorm |
| **523** | **prednisolone Oral Liquid Product** | 2 | 354.24 | 1165758 | RxNorm |
| **524** | **prednisolone; ophthalmic (corticosteroids, plain)** | 3 | 354.14 | S01BA04 | ATC |
| **525** | **prednisolone; ophthalmic, otic (corticosteroids)** | 3 | 354.14 | S03BA02 | ATC |
| **526** | **prednisolone; ophthalmic (corticosteroids/antiinfectives/mydriatics in comb.)** | 3 | 354.14 | S01CB02 | ATC |
| **527** | **prednisolone 3 MG/ML** | 4 | 353.33 | 317474 | RxNorm |
| **528** | **prednisolone; otic** | 3 | 352.51 | S02BA03 | ATC |
| **533** | **famotidine Oral Tablet** | 3 | 351.00 | 372140 | RxNorm |
| **534** | **famotidine Pill** | 2 | 350.77 | 1159022 | RxNorm |
| **535** | **calcium compounds - combinations** | 3 | 350.38 | A02AC10 | ATC |
| **536** | **Calcium compounds** | 2 | 350.38 | A02AC | ATC |
| **537** | **rutoside, combinations; systemic** | 3 | 345.58 | C05CA51 | ATC |
| **544** | **potassium chloride 0.02 MEQ/ML** | 4 | 342.51 | 316538 | RxNorm |
| **545** | **prednisolone and antiseptics; topical** | 3 | 330.37 | D07BA01 | ATC |
| **547** | **glucose / potassium chloride / sodium chloride Injectable Product** | 2 | 327.03 | 1165813 | RxNorm |
| **548** | **glucose / potassium chloride / sodium chloride Injection** | 3 | 327.03 | 1863604 | RxNorm |
| **549** | **Non-selective beta-adrenoreceptor agonists** | 2 | 323.97 | R03CB | ATC |
| **550** | **isoprenaline, combinations; systemic** | 3 | 322.56 | R03CB51 | ATC |
| **551** | **famotidine Oral Suspension** | 3 | 315.95 | 372139 | RxNorm |
| **552** | **famotidine Oral Liquid Product** | 2 | 315.95 | 1159020 | RxNorm |
| **553** | **famotidine 8 MG/ML** | 4 | 315.95 | 315922 | RxNorm |
| **556** | **acetaminophen Pill** | 2 | 307.06 | 1152843 | RxNorm |
| **559** | **Bisphosphonates, combinations** | 2 | 305.52 | M05BB | ATC |
| **563** | **prednisolone 3 MG/ML Oral Solution** | 3 | 298.99 | 283077 | RxNorm |
| **565** | **Antiseptics** | 2 | 295.00 | R02AA | ATC |
| **567** | **Blood coagulation factors** | 2 | 293.27 | B02BD | ATC |
| **568** | **ceftriaxone Injectable Solution** | 3 | 293.03 | 376593 | RxNorm |
| **569** | **Other intestinal adsorbents** | 2 | 292.29 | A07BC | ATC |
| **570** | **combinations; oral (intestinal adsorbents)** | 3 | 292.29 | A07BC30 | ATC |
| **571** | **heparin** | 3 | 289.88 | 5224 | RxNorm |

| | | | | | |
|---|---|---|---|---|---|
| **573** | **quinidine, combinations with psycholeptics; systemic** | 3 | 287.36 | C01BA71 | ATC |
| **574** | **coagulation factor IX, II, VII and X in combination; parenteral** | 3 | 287.00 | B02BD01 | ATC |
| **575** | **Synthetic anticholinergic agents in combination with psycholeptics** | 2 | 286.55 | A03CA | ATC |
| **576** | **epinephrine 1 MG/ML** | 4 | 285.29 | 328316 | RxNorm |
| **577** | **epinephrine; topical** | 3 | 285.29 | B02BC09 | ATC |
| **578** | **Heparins or heparinoids for topical use** | 2 | 284.98 | C05BA | ATC |
| **581** | **amitriptyline and psycholeptics; systemic** | 3 | 280.61 | N06CA01 | ATC |
| **582** | **combinations; inhalant, nasal** | 3 | 279.58 | R01AX30 | ATC |
| **583** | **Other nasal preparations** | 2 | 279.58 | R01AX | ATC |
| **584** | **epinephrine, combinations; ophthalmic** | 3 | 277.57 | S01EA51 | ATC |
| **585** | **phenylbutazone and corticosteroids; systemic** | 3 | 277.33 | M01BA01 | ATC |
| **586** | **Antiarrhythmics, class Ia** | 2 | 276.43 | C01BA | ATC |
| **587** | **propacetamol; parenteral** | 3 | 275.18 | N02BE05 | ATC |
| **588** | **Antidotes** | 2 | 274.21 | V03AB | ATC |
| **589** | **epinephrine Injectable Product** | 2 | 273.62 | 1163887 | RxNorm |
| **590** | **Antidepressants in combination with psycholeptics** | 2 | 272.96 | N06CA | ATC |
| **591** | **aminophenazone, combinations with psycholeptics; systemic** | 3 | 271.29 | N02BB73 | ATC |
| **592** | **methylscopolamine and psycholeptics; oral, rectal** | 3 | 269.60 | A03CB01 | ATC |
| **593** | **dextropropoxyphene, combinations with psycholeptics; systemic** | 3 | 268.32 | N02AC74 | ATC |
| **594** | **Enzymes** | 2 | 265.53 | M09AB | ATC |
| **595** | **trypsin, combinations; systemic** | 3 | 265.53 | M09AB52 | ATC |
| **596** | **Sulfonamides, plain** | 2 | 265.53 | C03BA | ATC |
| **597** | **glycopyrronium bromide and psycholeptics; systemic** | 3 | 265.24 | A03CA05 | ATC |
| **598** | **clorexolone, combinations with psycholeptics; oral** | 3 | 265.22 | C03BA82 | ATC |
| **599** | **fluoxetine and psycholeptics; oral** | 3 | 265.22 | N06CA03 | ATC |
| **600** | **melitracen and psycholeptics; systemic** | 3 | 265.22 | N06CA02 | ATC |
| **601** | **gefarnate, combinations with psycholeptics; oral** | 3 | 265.22 | A02BX77 | ATC |
| **602** | **bucetin, combinations with psycholeptics** | 3 | 265.22 | N02BE74 | ATC |
| **603** | **clidinium and psycholeptics; oral** | 3 | 265.22 | A03CA02 | ATC |
| **604** | **carbenoxolone, combinations with psycholeptics; oral, oral local, topical, vaginal** | 3 | 265.22 | A02BX71 | ATC |
| **605** | **chlorzoxazone, combinations with psycholeptics; oral** | 3 | 265.22 | M03BB73 | ATC |
| **606** | **chlormezanone, combinations with psycholeptics; oral, rectal** | 3 | 265.22 | M03BB72 | ATC |
| **607** | **emepronium and psycholeptics; oral** | 3 | 265.22 | A03CA30 | ATC |
| **608** | **diphemanil and psycholeptics; oral, topical** | 3 | 265.22 | A03CA08 | ATC |
| **609** | **isopropamide and psycholeptics; oral** | 3 | 265.22 | A03CA01 | ATC |
| **610** | **methocarbamol, combinations with psycholeptics; systemic** | 3 | 265.22 | M03BA73 | ATC |
| **611** | **carisoprodol, combinations with psycholeptics; oral** | 3 | 265.22 | M03BA72 | ATC |

| | | | | | |
|---|---|---|---|---|---|
| **612** | **otilonium bromide and psycholeptics; oral** | 3 | 265.22 | A03CA04 | ATC |
| **613** | **ambutonium and psycholeptics; oral** | 3 | 265.22 | A03CA07 | ATC |
| **614** | **bevonium and psycholeptics** | 3 | 265.22 | A03CA06 | ATC |
| **615** | **dipyrocetyl, combinations with psycholeptics** | 3 | 265.22 | N02BA79 | ATC |
| **616** | **organic nitrates in combination with psycholeptics** | 3 | 265.07 | C01DA70 | ATC |
| **617** | **sodium chloride Injection** | 3 | 264.33 | 1807545 | RxNorm |
| **618** | **belladonna total alkaloids and psycholeptics; systemic** | 3 | 264.24 | A03CB02 | ATC |
| **619** | **phenprobamate, combinations with psycholeptics; oral, rectal** | 3 | 264.19 | M03BA71 | ATC |
| **620** | **oxyphencyclimine and psycholeptics; oral** | 3 | 264.19 | A03CA03 | ATC |
| **621** | **propantheline and psycholeptics; oral, rectal, urethral** | 3 | 264.19 | A03CA34 | ATC |
| **622** | **pipenzolate and psycholeptics; oral** | 3 | 264.19 | A03CA09 | ATC |
| **623** | **pethidine, combinations with psycholeptics; systemic** | 3 | 264.14 | N02AB72 | ATC |
| **628** | **metamizole sodium, combinations with psycholeptics; systemic** | 3 | 262.55 | N02BB72 | ATC |
| **629** | **epinephrine; ophthalmic** | 3 | 262.40 | S01EA01 | ATC |
| **630** | **acetaminophen Injection** | 3 | 261.78 | 1803956 | RxNorm |
| **631** | **acetaminophen Injectable Product** | 2 | 261.78 | 1152840 | RxNorm |
| **634** | **combinations; topical (antibiotics)** | 3 | 259.75 | D01AA20 | ATC |
| **635** | **Antibiotics** | 2 | 259.75 | D01AA | ATC |
| **636** | **acetaminophen 10 MG/ML** | 4 | 259.66 | 483015 | RxNorm |
| **638** | **Anesthetics, local** | 2 | 258.68 | R02AD | ATC |
| **639** | **various; local oral** | 3 | 254.95 | R02AA20 | ATC |
| **640** | **heparin; parenteral** | 3 | 252.78 | B01AB01 | ATC |
| **642** | **acetaminophen Oral Tablet** | 3 | 252.07 | 369097 | RxNorm |
| **643** | **Anesthetics for topical use** | 2 | 250.67 | D04AB | ATC |
| **644** | **Local anesthetics** | 2 | 250.67 | C05AD | ATC |
| **648** | **Corticosteroids and mydriatics in combination** | 2 | 242.96 | S01BB | ATC |
| **649** | **fluorescein, combinations; ophthalmic** | 3 | 240.33 | S01JA51 | ATC |
| **650** | **dodeclonium bromide, combinations; topical** | 3 | 239.79 | D08AJ59 | ATC |
| **651** | **lidocaine** | 3 | 239.79 | 6387 | RxNorm |
| **652** | **diphenhydramine; oral, rectal** | 3 | 239.40 | R06AA02 | ATC |
| **653** | **Antiarrhythmics, class Ib** | 2 | 239.13 | C01BB | ATC |
| **654** | **diflucortolone, combinations; topical** | 3 | 238.65 | D07XC04 | ATC |
| **655** | **fluocortolone, combinations; topical** | 3 | 238.65 | D07XC05 | ATC |
| **656** | **diphenhydramine Oral Product** | 2 | 238.39 | 1158449 | RxNorm |
| **657** | **Colouring agents** | 2 | 238.26 | S01JA | ATC |
| **658** | **hydrocortisone, combinations; inhalant, nasal** | 3 | 238.14 | R01AD60 | ATC |

| | | | | | |
|---|---|---|---|---|---|
| **659** | **glucose Injectable Solution** | 3 | 237.96 | 376937 | RxNorm |
| **660** | **combinations; systemic (beta-lactamase sensitive penicillins)** | 3 | 237.52 | J01CE30 | ATC |
| **661** | **Beta-lactamase sensitive penicillins** | 2 | 237.52 | J01CE | ATC |
| **669** | **hydrocortisone and antiseptics; topical** | 3 | 236.09 | D07BA04 | ATC |
| **674** | **betamethasone, combinations; topical** | 3 | 234.20 | D07XC01 | ATC |
| **675** | **hyaluronic acid, combinations; ophthalmic** | 3 | 233.48 | S01KA51 | ATC |
| **676** | **Viscoelastic substances** | 2 | 232.33 | S01KA | ATC |
| **677** | **Sympathomimetics in glaucoma therapy** | 2 | 227.84 | S01EA | ATC |
| **678** | **lidocaine; rectal, topical** | 3 | 227.41 | C05AD01 | ATC |
| **679** | **lidocaine; topical** | 3 | 227.41 | D04AB01 | ATC |
| **680** | **Antivirals** | 2 | 226.58 | D06BB | ATC |
| **683** | **famotidine 8 MG/ML Oral Suspension** | 3 | 224.54 | 310274 | RxNorm |
| **685** | **oxytetracycline, combinations; systemic** | 3 | 223.35 | J01AA56 | ATC |
| **686** | **Serotonin (5HT3) antagonists** | 2 | 222.89 | A04AA | ATC |
| **687** | **Other antiinfectives** | 2 | 222.60 | S01AX | ATC |
| **688** | **morphine, combinations; systemic** | 3 | 221.21 | N02AA51 | ATC |
| **689** | **ondansetron; systemic, rectal** | 3 | 220.93 | A04AA01 | ATC |
| **690** | **ondansetron** | 3 | 220.93 | 26225 | RxNorm |
| **691** | **epinephrine; parenteral** | 3 | 220.02 | C01CA24 | ATC |
| **692** | **100 ML acetaminophen 10 MG/ML Injection** | 3 | 218.91 | 483017 | RxNorm |
| **693** | **Antibiotics** | 2 | 218.80 | A07AA | ATC |
| **694** | **heparin Injectable Product** | 2 | 218.69 | 1856274 | RxNorm |
| **697** | **aciclovir, combinations; topical** | 3 | 216.75 | D06BB53 | ATC |
| **698** | **Acidifiers** | 2 | 216.17 | G04BA | ATC |
| **699** | **calcium chloride** | 3 | 216.17 | 1901 | RxNorm |
| **700** | **propanol, combinations; topical** | 3 | 214.06 | D08AX53 | ATC |
| **701** | **potassium chloride; oral** | 3 | 213.82 | A12BA01 | ATC |
| **702** | **Other antiseptics and disinfectants** | 2 | 212.65 | D08AX | ATC |
| **703** | **senna glycosides; systemic** | 3 | 212.35 | A06AB06 | ATC |
| **704** | **oxycodone and paracetamol; systemic** | 3 | 211.93 | N02AJ17 | ATC |
| **705** | **phenylephrine, combinations; ophthalmic** | 3 | 210.09 | S01GA55 | ATC |
| **707** | **combinations; systemic (penicillins with extended spectrum)** | 3 | 208.73 | J01CA20 | ATC |
| **708** | **Penicillins with extended spectrum** | 2 | 208.73 | J01CA | ATC |
| **709** | **zinc compounds; ophthalmic** | 3 | 206.45 | S01AX03 | ATC |
| **710** | **Antiinflammatory preparations, non-steroids for topical use** | 2 | 205.78 | M02AA | ATC |
| **711** | **Pyrimidine analogues** | 2 | 204.43 | L01BC | ATC |
| **712** | **epinephrine; inhalant** | 3 | 203.46 | R03AA01 | ATC |

| | | | | | |
|---|---|---|---|---|---|
| **713** | **epinephrine; inhalant, nasal** | 3 | 203.46 | R01AA14 | ATC |
| **714** | **articaine, combinations; inhalant, parenteral** | 3 | 203.24 | N01BB58 | ATC |
| **715** | **etidocaine, combinations; parenteral** | 3 | 201.12 | N01BB57 | ATC |
| **716** | **epinephrine** | 3 | 201.12 | 3992 | RxNorm |
| **717** | **Alpha- and beta-adrenoreceptor agonists** | 2 | 201.12 | R03AA | ATC |
| **718** | **tramadol and paracetamol; systemic** | 3 | 199.90 | N02AJ13 | ATC |
| **719** | **fluorouracil, combinations; systemic** | 3 | 199.44 | L01BC52 | ATC |
| **720** | **Antiinflammatory products for vaginal administration** | 2 | 198.91 | G02CC | ATC |
| **721** | **calcium chloride / glucose / lactate / potassium chloride / sodium chloride Injectable Product** | 2 | 196.16 | 1154150 | RxNorm |
| **722** | **calcium chloride / glucose / lactate / potassium chloride / sodium chloride Injectable Solution** | 3 | 196.16 | 876076 | RxNorm |
| **723** | **famotidine, combinations; systemic** | 3 | 195.80 | A02BA53 | ATC |
| **724** | **calcium chloride 0.001 MEQ/ML / glucose 50 MG/ML / potassium chloride 0.004 MEQ/ML / sodium chloride 0.103 MEQ/ML / sodium lactate 0.028 MEQ/ML Injectable Solution** | 3 | 195.56 | 847627 | RxNorm |
| **725** | **calcium chloride 0.001 MEQ/ML** | 4 | 194.28 | 847619 | RxNorm |
| **726** | **Ergot alkaloids** | 2 | 194.16 | G02AB | ATC |
| **727** | **ergot alkaloids; systemic** | 3 | 194.16 | G02AB02 | ATC |
| **728** | **acetaminophen 325 MG** | 4 | 193.16 | 315263 | RxNorm |
| **729** | **Other drugs for constipation** | 2 | 191.16 | A06AX | ATC |
| **731** | **Combinations for eradication of Helicobacter pylori** | 2 | 189.49 | A02BD | ATC |
| **734** | **acetylsalicylic acid and corticosteroids; systemic** | 3 | 183.06 | M01BA03 | ATC |
| **735** | **dipyrocetyl and corticosteroids** | 3 | 183.06 | M01BA02 | ATC |
| **737** | **Other dermatologicals** | 2 | 181.98 | D11AX | ATC |
| **738** | **carbon dioxide producing drugs** | 3 | 181.24 | A06AX02 | ATC |
| **740** | **lidocaine Topical Product** | 2 | 177.98 | 1164666 | RxNorm |
| **741** | **lidocaine Topical Cream** | 3 | 177.25 | 377740 | RxNorm |
| **742** | **lactate** | 3 | 176.19 | 114202 | RxNorm |
| **743** | **sodium lactate 0.028 MEQ/ML** | 4 | 175.18 | 797836 | RxNorm |
| **744** | **sodium chloride 0.103 MEQ/ML** | 4 | 175.18 | 797835 | RxNorm |
| **745** | **potassium chloride 0.004 MEQ/ML** | 4 | 174.82 | 665002 | RxNorm |
| **746** | **pilocarpine, combinations; ophthalmic** | 3 | 174.46 | S01EB51 | ATC |
| **747** | **Parasympathomimetics** | 2 | 174.00 | S01EB | ATC |
| **748** | **Other cardiac preparations** | 2 | 173.97 | C01EB | ATC |
| **749** | **mometasone, combinations; topical (corticosteroids, potent, other comb.)** | 3 | 173.76 | D07XC03 | ATC |

| | | | | | |
|---|---|---|---|---|---|
| **750** | **mometasone, combinations; topical (corticosteroids)** | 3 | 173.76 | R01AD59 | ATC |
| **751** | **heparin; topical** | 3 | 173.19 | C05BA03 | ATC |
| **752** | **Organic acids** | 2 | 173.00 | G01AD | ATC |
| **753** | **omeprazole** | 3 | 170.74 | 7646 | RxNorm |
| **754** | **lidocaine; ophthalmic** | 3 | 169.20 | S01HA07 | ATC |
| **757** | **lidocaine; local oral** | 3 | 168.12 | R02AD02 | ATC |
| **763** | **ibuprofen** | 3 | 163.96 | 5640 | RxNorm |
| **764** | **Other throat preparations** | 2 | 163.96 | R02AX | ATC |
| **768** | **ibuprofen; systemic, rectal** | 3 | 163.40 | M01AE01 | ATC |
| **769** | **ibuprofen Oral Product** | 2 | 163.40 | 1156277 | RxNorm |
| **770** | **ibuprofen; oral** | 3 | 163.40 | R02AX02 | ATC |
| **771** | **Sympathomimetics, plain** | 2 | 163.30 | R01AA | ATC |
| **772** | **diastase; inhalant, oral** | 3 | 162.65 | A09AA01 | ATC |
| **773** | **Enzyme preparations** | 2 | 162.65 | A09AA | ATC |
| **775** | **acetaminophen 325 MG Oral Tablet** | 3 | 161.78 | 313782 | RxNorm |
| **778** | **senna glycosides, combinations; systemic** | 3 | 157.33 | A06AB56 | ATC |
| **779** | **diphenhydramine Oral Liquid Product** | 2 | 156.48 | 1158448 | RxNorm |
| **780** | **diphenhydramine Oral Solution** | 3 | 156.48 | 371889 | RxNorm |
| **781** | **diphenhydramine hydrochloride 2.5 MG/ML** | 4 | 156.11 | 997155 | RxNorm |
| **782** | **triamcinolone, combinations; topical** | 3 | 152.51 | D07XB02 | ATC |
| **783** | **polyethylene glycol 3350 17000 MG** | 4 | 147.56 | 1870356 | RxNorm |
| **784** | **macrogol; oral** | 3 | 147.56 | A06AD15 | ATC |
| **785** | **sodium chloride Prefilled Syringe** | 3 | 147.10 | 727633 | RxNorm |
| **786** | **sodium chloride 9 MG/ML Prefilled Syringe** | 3 | 146.78 | 1359867 | RxNorm |
| **788** | **Watersoluble, nephrotropic, low osmolar X-ray contrast media** | 2 | 145.28 | V08AB | ATC |
| **789** | **magnesium (different salts in combination)** | 3 | 144.56 | A12CC30 | ATC |
| **790** | **Magnesium** | 2 | 144.56 | A12CC | ATC |
| **791** | **magnesium compounds - combinations** | 3 | 144.56 | A02AA10 | ATC |
| **792** | **Magnesium compounds** | 2 | 144.56 | A02AA | ATC |
| **793** | **polyethylene glycol 3350** | 3 | 143.84 | 221147 | RxNorm |
| **795** | **ondansetron Pill** | 2 | 143.71 | 1161034 | RxNorm |
| **797** | **ondansetron Oral Product** | 2 | 142.89 | 1161033 | RxNorm |

| | | | | | |
|---|---|---|---|---|---|
| **799** | **diphenhydramine hydrochloride 2.5 MG/ML Oral Solution** | 3 | 141.03 | 1049906 | RxNorm |
| **800** | **imidazoles/triazoles in combination with corticosteroids; topical** | 3 | 140.94 | D01AC20 | ATC |
| **801** | **lidocaine; parenteral** | 3 | 140.19 | C01BB01 | ATC |
| **808** | **heparin Injectable Solution** | 3 | 133.65 | 1856275 | RxNorm |
| **809** | **sodium chloride 9 MG/ML Injection** | 3 | 132.63 | 1807628 | RxNorm |
| **810** | **polyethylene glycol 3350 Oral Product** | 2 | 131.68 | 1162922 | RxNorm |
| **811** | **polyethylene glycol 3350 Powder for Oral Solution** | 3 | 131.68 | 1870358 | RxNorm |
| **812** | **ondansetron Disintegrating Oral Product** | 2 | 131.46 | 1295332 | RxNorm |
| **813** | **ondansetron Disintegrating Oral Tablet** | 3 | 131.46 | 373149 | RxNorm |
| **814** | **polyethylene glycol 3350 Oral Powder Product** | 2 | 130.89 | 1870357 | RxNorm |
| **815** | **morphine, combinations; oral** | 3 | 129.58 | A07DA52 | ATC |
| **818** | **ondansetron 4 MG** | 4 | 126.35 | 328450 | RxNorm |
| **819** | **glucose Injection** | 3 | 125.93 | 1794567 | RxNorm |
| **821** | **acetaminophen 32 MG/ML** | 4 | 122.87 | 315262 | RxNorm |
| **822** | **acetaminophen Oral Liquid Product** | 2 | 122.21 | 1152841 | RxNorm |
| **831** | **carbenoxolone, combinations excl. psycholeptics; oral, oral local, topical, vaginal** | 3 | 116.98 | A02BX51 | ATC |
| **837** | **contact laxatives in combination with belladonna alkaloids** | 3 | 116.70 | A06AB30 | ATC |
| **838** | **Adrenergics in combination with anticholinergics incl. triple combinations with corticosteroids** | 2 | 115.72 | R03AL | ATC |
| **842** | **polyethylene glycol 3350 17000 MG Powder for Oral Solution** | 3 | 112.10 | 876193 | RxNorm |
| **843** | **heparin, combinations; topical** | 3 | 112.03 | C05BA53 | ATC |
| **847** | **water** | 3 | 109.98 | 11295 | RxNorm |
| **848** | **midazolam Injectable Product** | 2 | 109.63 | 1164715 | RxNorm |
| **855** | **lidocaine hydrochloride 40 MG/ML** | 4 | 106.64 | 1010843 | RxNorm |
| **856** | **sodium bicarbonate** | 3 | 105.85 | 36676 | RxNorm |
| **858** | **omeprazole Pill** | 2 | 104.73 | 1161029 | RxNorm |

| | | | | | |
|---|---|---|---|---|---|
| **859** | **omeprazole Oral Product** | 2 | 104.45 | 1161028 | RxNorm |
| **861** | **omeprazole; systemic** | 3 | 104.26 | A02BC01 | ATC |
| **864** | **benzoyl peroxide, combinations; topical** | 3 | 100.82 | D10AE51 | ATC |
| **865** | **Peroxides** | 2 | 100.69 | D10AE | ATC |
| **866** | **omeprazole Delayed Release Oral Capsule** | 3 | 100.45 | 378236 | RxNorm |
| **872** | **ibuprofen Pill** | 2 | 98.68 | 1156278 | RxNorm |
| **873** | **Corticosteroids for local oral treatment** | 2 | 98.15 | A01AC | ATC |
| **883** | **Opioids in combination with antispasmodics** | 2 | 97.51 | N02AG | ATC |
| **890** | **acetaminophen Oral Suspension** | 3 | 93.44 | 370509 | RxNorm |
| **895** | **Antiinfectives and antiseptics for local oral treatment** | 2 | 89.88 | A01AB | ATC |
| **898** | **attapulgite, combinations; oral** | 3 | 89.67 | A07BC54 | ATC |
| **904** | **Natural and semisynthetic estrogens, plain** | 2 | 88.30 | G03CA | ATC |
| **910** | **estradiol, combinations; systemic** | 3 | 87.13 | G03CA53 | ATC |
| **911** | **ibuprofen Oral Tablet** | 3 | 86.61 | 370674 | RxNorm |
| **912** | **penicillins, combinations with other antibacterials; systemic** | 3 | 86.13 | J01RA01 | ATC |
| **913** | **ampicillin, combinations; systemic** | 3 | 85.68 | J01CA51 | ATC |
| **915** | **Selective beta-2-adrenoreceptor agonists** | 2 | 85.59 | R03AC | ATC |
| **916** | **Combinations of antibacterials** | 2 | 85.43 | J01RA | ATC |
| **919** | **Corticosteroids** | 2 | 84.60 | S02BA | ATC |
| **921** | **ondansetron 4 MG Disintegrating Oral Tablet** | 3 | 83.41 | 104894 | RxNorm |
| **922** | **Corticosteroids/antiinfectives/mydriatics in combination** | 2 | 83.38 | S01CB | ATC |
| **923** | **diphenhydramine Pill** | 2 | 82.89 | 1158450 | RxNorm |
| **926** | **ondansetron 2 MG/ML** | 4 | 81.44 | 328448 | RxNorm |
| **927** | **ondansetron Injectable Product** | 2 | 81.25 | 1161031 | RxNorm |
| **928** | **clindamycin** | 3 | 80.98 | 2582 | RxNorm |
| **929** | **Lincosamides** | 2 | 80.98 | J01FF | ATC |
| **930** | **Other quaternary ammonium compounds** | 2 | 80.63 | M03AC | ATC |
| **932** | **morphine and antispasmodics; systemic** | 3 | 79.83 | N02AG01 | ATC |
| **933** | **Corticosteroids** | 2 | 79.69 | C05AA | ATC |
| **935** | **omeprazole, amoxicillin and clarithromycin; systemic** | 3 | 79.46 | A02BD05 | ATC |
| **936** | **midazolam Injectable Solution** | 3 | 78.82 | 379133 | RxNorm |
| **937** | **Corticosteroids, plain** | 2 | 78.66 | S01BA | ATC |

| | | | | | |
|---|---|---|---|---|---|
| **939** | **omega-3-triglycerides incl. other esters and acids; systemic** | 3 | 78.11 | C10AX06 | ATC |
| **940** | **miconazole, combinations; topical** | 3 | 78.03 | D01AC52 | ATC |
| **941** | **Other lipid modifying agents** | 2 | 77.71 | C10AX | ATC |
| **946** | **Antiinfectives for treatment of acne** | 2 | 75.44 | D10AF | ATC |
| **947** | **ibuprofen 20 MG/ML** | 4 | 75.40 | 316073 | RxNorm |
| **948** | **lorazepam 2 MG/ML** | 4 | 75.25 | 316171 | RxNorm |
| **949** | **ibuprofen Oral Liquid Product** | 2 | 75.19 | 1156276 | RxNorm |
| **950** | **ibuprofen Oral Suspension** | 3 | 75.19 | 370672 | RxNorm |
| **951** | **hydromorphone and antispasmodics; systemic** | 3 | 75.11 | N02AG04 | ATC |
| **955** | **Corticosteroids** | 2 | 73.07 | S03BA | ATC |
| **956** | **lorazepam; systemic, sublingual** | 3 | 72.62 | N05BA06 | ATC |
| **957** | **lorazepam** | 3 | 72.56 | 6470 | RxNorm |
| **958** | **melatonin** | 3 | 72.38 | 6711 | RxNorm |
| **959** | **Melatonin receptor agonists** | 2 | 72.38 | N05CH | ATC |
| **960** | **clindamycin; systemic** | 3 | 72.36 | J01FF01 | ATC |
| **961** | **fentanyl** | 3 | 71.91 | 4337 | RxNorm |
| **965** | **melatonin Oral Product** | 2 | 71.24 | 1159785 | RxNorm |
| **966** | **melatonin; oral** | 3 | 71.17 | N05CH01 | ATC |
| **967** | **Antiinflammatory agents, non-steroids** | 2 | 70.71 | S01BC | ATC |
| **968** | **tretinoin, combinations; topical** | 3 | 70.15 | D10AD51 | ATC |
| **970** | **Benzodiazepine derivatives** | 2 | 70.01 | N05CD | ATC |
| **972** | **contact laxatives in combination** | 3 | 69.53 | A06AB20 | ATC |
| **976** | **ondansetron Injectable Solution** | 3 | 68.26 | 376327 | RxNorm |
| **977** | **hydrocortisone; systemic** | 3 | 68.16 | H02AB09 | ATC |
| **978** | **midazolam** | 3 | 67.95 | 6960 | RxNorm |
| **979** | **midazolam 1 MG/ML** | 4 | 67.95 | 328485 | RxNorm |
| **981** | **lidocaine Injectable Product** | 2 | 66.06 | 1164658 | RxNorm |
| **982** | **midazolam; nasal, systemic** | 3 | 66.02 | N05CD08 | ATC |
| **984** | **sulfonamides, combinations with other antibacterials (excl. trimethoprim); systemic** | 3 | 65.81 | J01RA02 | ATC |
| **986** | **Retinoids for topical use in acne** | 2 | 65.05 | D10AD | ATC |
| **996** | **ketobemidone and antispasmodics; systemic** | 3 | 64.07 | N02AG02 | ATC |
| **998** | **ketorolac; systemic** | 3 | 63.82 | M01AB15 | ATC |

| | | | | | |
|---|---|---|---|---|---|
| **999** | **pethidine and antispasmodics; systemic** | 3 | 63.79 | N02AG03 | ATC |
| **1,004** | **ketorolac** | 3 | 63.66 | 35827 | RxNorm |
| **1,005** | **Androgens and estrogens** | 2 | 63.56 | G03EA | ATC |
| **1,012** | **acetaminophen 32 MG/ML Oral Suspension** | 3 | 62.35 | 307668 | RxNorm |
| **1,013** | **ketorolac Injectable Product** | 2 | 61.98 | 1160966 | RxNorm |
| **1,014** | **diflucortolone and antiseptics; topical** | 3 | 61.70 | D07BC04 | ATC |
| **1,015** | **fluocortolone and antiseptics; topical** | 3 | 61.70 | D07BC03 | ATC |
| **1,016** | **fluocinolone acetonide and antiseptics; topical** | 3 | 61.70 | D07BC02 | ATC |
| **1,017** | **betamethasone and antiseptics; topical** | 3 | 61.70 | D07BC01 | ATC |
| **1,018** | **hydrocortisone butyrate and antiseptics; topical** | 3 | 61.70 | D07BB04 | ATC |
| **1,019** | **triamcinolone and antiseptics; topical** | 3 | 61.70 | D07BB03 | ATC |
| **1,020** | **desonide and antiseptics; topical** | 3 | 61.70 | D07BB02 | ATC |
| **1,021** | **flumetasone and antiseptics; topical** | 3 | 61.70 | D07BB01 | ATC |
| **1,024** | **Allergen extracts** | 2 | 61.27 | V01AA | ATC |
| **1,025** | **lorazepam Injectable Product** | 2 | 61.13 | 1165338 | RxNorm |
| **1,030** | **formoterol and budesonide; inhalant** | 3 | 60.48 | R03AK07 | ATC |
| **1,033** | **ondansetron 2 MG/ML Injectable Solution** | 3 | 58.27 | 283504 | RxNorm |
| **1,041** | **rocuronium bromide 10 MG/ML Injectable Solution** | 3 | 57.24 | 1234995 | RxNorm |
| **1,045** | **aminophylline, combinations; systemic** | 3 | 56.60 | R03DA55 | ATC |
| **1,046** | **rocuronium bromide 10 MG/ML** | 4 | 55.72 | 998229 | RxNorm |
| **1,047** | **rocuronium Injectable Product** | 2 | 55.72 | 1156444 | RxNorm |
| **1,048** | **rocuronium** | 3 | 55.72 | 68139 | RxNorm |
| **1,049** | **rocuronium bromide; parenteral** | 3 | 55.72 | M03AC09 | ATC |
| **1,050** | **rocuronium Injectable Solution** | 3 | 55.72 | 375623 | RxNorm |
| **1,051** | **influenza B virus B/Phuket/3073/2013 antigen 0.03 MG/ML** | 4 | 55.63 | 1657229 | RxNorm |
| **1,052** | **influenza A virus (H1N1) antigen / influenza A virus (H3N2) antigen / influenza B virus antigen Prefilled Syringe** | 3 | 55.59 | 1657137 | RxNorm |
| **1,053** | **ispaghula, combinations; systemic** | 3 | 55.57 | A06AC51 | ATC |
| **1,054** | **sennosides, USP** | 3 | 55.57 | 36387 | RxNorm |
| **1,055** | **influenza, inactivated, split virus or surface antigen; systemic** | 3 | 55.45 | J07BB02 | ATC |
| **1,056** | **influenza A virus (H1N1) antigen / influenza A virus (H3N2) antigen / influenza B virus antigen Injectable Product** | 2 | 55.45 | 1657136 | RxNorm |

| | | | | | |
|---|---|---|---|---|---|
| **1,057** | **docusate sodium, incl. combinations; rectal** | 3 | 55.34 | A06AG10 | ATC |
| **1,059** | **fentanyl Injectable Product** | 2 | 55.20 | 1159056 | RxNorm |
| **1,060** | **dexmedetomidine** | 3 | 55.15 | 48937 | RxNorm |
| **1,061** | **influenza A virus (H3N2) antigen** | 3 | 55.14 | 1657131 | RxNorm |
| **1,062** | **influenza B virus antigen** | 3 | 55.14 | 1657134 | RxNorm |
| **1,063** | **influenza A virus (H1N1) antigen** | 3 | 55.14 | 1657128 | RxNorm |
| **1,064** | **Influenza vaccines** | 2 | 55.14 | J07BB | ATC |
| **1,066** | **sennosides, USP Oral Product** | 2 | 55.03 | 1159068 | RxNorm |
| **1,071** | **Other hypnotics and sedatives** | 2 | 54.55 | N05CM | ATC |
| **1,072** | **fentanyl; systemic** | 3 | 54.54 | N01AH01 | ATC |
| **1,074** | **feather; systemic** | 3 | 53.89 | V01AA01 | ATC |
| **1,079** | **Corticosteroids, potent, combinations with antiseptics** | 2 | 53.28 | D07BC | ATC |
| **1,080** | **albuterol Inhalant Product** | 2 | 52.94 | 1154602 | RxNorm |
| **1,081** | **salbutamol; inhalant** | 3 | 52.88 | R03AC02 | ATC |
| **1,082** | **neomycin, combinations; oral** | 3 | 52.15 | A07AA51 | ATC |
| **1,084** | **petrolatum** | 4 | 51.51 | 8091 | RxNorm |
| **1,085** | **neostigmine, combinations; systemic** | 3 | 51.49 | N07AA51 | ATC |
| **1,086** | **ACE inhibitors and calcium channel blockers** | 2 | 51.46 | C09BB | ATC |
| **1,087** | **Antiallergic agents, excl. corticosteroids** | 2 | 51.41 | R01AC | ATC |
| **1,088** | **calcium chloride 0.0014 MEQ/ML** | 4 | 51.04 | 847624 | RxNorm |
| **1,089** | **cromoglicic acid, combinations; inhalant, nasal** | 3 | 50.95 | R01AC51 | ATC |
| **1,090** | **opium alkaloids and derivatives - combinations** | 3 | 50.68 | R05DA20 | ATC |
| **1,091** | **Opium alkaloids and derivatives** | 2 | 50.68 | R05DA | ATC |
| **1,092** | **calcium chloride 0.0014 MEQ/ML / potassium chloride 0.004 MEQ/ML / sodium chloride 0.103 MEQ/ML / sodium lactate 0.028 MEQ/ML Injectable Solution** | 3 | 50.62 | 847630 | RxNorm |
| **1,093** | **calcium chloride / lactate / potassium chloride / sodium chloride Injectable Solution** | 3 | 50.62 | 876077 | RxNorm |
| **1,094** | **calcium chloride / lactate / potassium chloride / sodium chloride Injectable Product** | 2 | 50.62 | 1154438 | RxNorm |
| **1,096** | **Selective beta-2-adrenoreceptor agonists** | 2 | 49.39 | R03CC | ATC |
| **1,097** | **albuterol** | 3 | 49.24 | 435 | RxNorm |
| **1,098** | **Bulk-forming laxatives** | 2 | 48.88 | A06AC | ATC |
| **1,099** | **ibuprofen 20 MG/ML Oral Suspension** | 3 | 48.80 | 197803 | RxNorm |
| **1,100** | **ampicillin and beta-lactamase inhibitor; parenteral** | 3 | 48.73 | J01CR01 | ATC |

| | | | | | |
|---|---|---|---|---|---|
| **1,101** | **ketorolac Injection** | 3 | 48.62 | 1665000 | RxNorm |
| **1,102** | **phenytoin, combinations; systemic** | 3 | 48.46 | N03AB52 | ATC |
| **1,107** | **Other general anesthetics** | 2 | 48.08 | N01AX | ATC |
| **1,110** | **Hydantoin derivatives** | 2 | 47.04 | N03AB | ATC |
| **1,114** | **ibuprofen 200 MG** | 4 | 46.68 | 316074 | RxNorm |
| **1,115** | **ibuprofen; topical** | 3 | 46.68 | M02AA13 | ATC |
| **1,116** | **tetryzoline, combinations; ophthalmic** | 3 | 46.66 | S01GA52 | ATC |
| **1,118** | **Other cephalosporins and penems** | 2 | 45.85 | J01DI | ATC |
| **1,119** | **fluticasone, combinations; inhalant, nasal** | 3 | 45.75 | R01AD58 | ATC |
| **1,120** | **ProAir Inhalant Product** | 2 | 45.71 | 1649960 | RxNorm |
| **1,121** | **albuterol 0.09 MG/ACTUAT [ProAir]** | 4 | 45.71 | 1649958 | RxNorm |
| **1,122** | **methyltestosterone and estrogen; systemic** | 3 | 45.63 | G03EA01 | ATC |
| **1,124** | **formoterol and glycopyrronium bromide; inhalant** | 3 | 45.36 | R03AL07 | ATC |
| **1,126** | **ceftriaxone and beta-lactamase inhibitor; systemic** | 3 | 44.93 | J01DD63 | ATC |
| **1,127** | **piperacillin and beta-lactamase inhibitor; parenteral** | 3 | 44.93 | J01CR05 | ATC |
| **1,128** | **ticarcillin and beta-lactamase inhibitor; parenteral** | 3 | 44.93 | J01CR03 | ATC |
| **1,129** | **cefotaxime and beta-lactamase inhibitor; systemic** | 3 | 44.93 | J01DD51 | ATC |
| **1,130** | **ceftolozane and beta-lactamase inhibitor; parenteral** | 3 | 44.93 | J01DI54 | ATC |
| **1,131** | **cefpodoxime and beta-lactamase inhibitor; systemic** | 3 | 44.93 | J01DD64 | ATC |
| **1,135** | **lidocaine hydrochloride 10 MG/ML** | 4 | 44.27 | 1010032 | RxNorm |
| **1,136** | **albuterol Metered Dose Inhaler [ProAir]** | 3 | 44.16 | 1649967 | RxNorm |
| **1,137** | **Anticholinesterases** | 2 | 44.12 | N07AA | ATC |
| **1,138** | **ceftazidime and beta-lactamase inhibitor; parenteral** | 3 | 43.89 | J01DD52 | ATC |
| **1,141** | **albuterol 0.09 MG/ACTUAT Metered Dose Inhaler [ProAir]** | 3 | 43.07 | 1359561 | RxNorm |
| **1,142** | **lidocaine; otic** | 3 | 42.86 | S02DA01 | ATC |
| **1,143** | **Benzodiazepine derivatives** | 2 | 42.61 | N05BA | ATC |
| **1,145** | **Glucocorticoids** | 2 | 42.43 | R03BA | ATC |
| **1,146** | **melatonin Oral Tablet** | 3 | 42.37 | 372751 | RxNorm |
| **1,149** | **ketorolac tromethamine 15 MG/ML** | 4 | 41.90 | 860091 | RxNorm |
| **1,150** | **melatonin Pill** | 2 | 41.53 | 1159786 | RxNorm |
| **1,151** | **dexmedetomidine Injectable Product** | 2 | 41.26 | 1154546 | RxNorm |
| **1,152** | **dexmedetomidine; parenteral** | 3 | 41.16 | N05CM18 | ATC |
| **1,154** | **lidocaine Injectable Solution** | 3 | 40.71 | 372599 | RxNorm |

| | | | | | |
|---|---|---|---|---|---|
| **1,155** | **Corticosteroids, moderately potent, combinations with antiseptics** | 2 | 40.59 | D07BB | ATC |
| **1,161** | **acetaminophen Rectal Suppository** | 3 | 39.31 | 370508 | RxNorm |
| **1,162** | **acetaminophen Rectal Product** | 2 | 39.31 | 1152844 | RxNorm |
| **1,163** | **fluticasone furoate; nasal** | 3 | 39.06 | R01AD12 | ATC |
| **1,164** | **fluticasone furoate; inhalant** | 3 | 39.00 | R03BA09 | ATC |
| **1,171** | **Antibiotics** | 2 | 37.26 | G01AA | ATC |
| **1,175** | **Macrolides** | 2 | 37.09 | J01FA | ATC |
| **1,176** | **Zofran Oral Product** | 2 | 37.01 | 1188160 | RxNorm |
| **1,177** | **azithromycin** | 3 | 36.93 | 18631 | RxNorm |
| **1,178** | **azithromycin; systemic** | 3 | 36.93 | J01FA10 | ATC |
| **1,179** | **midazolam 5 MG/ML** | 4 | 36.38 | 330809 | RxNorm |
| **1,182** | **albuterol 0.09 MG/ACTUAT** | 4 | 35.20 | 329498 | RxNorm |
| **1,183** | **albuterol Metered Dose Inhaler** | 3 | 34.85 | 745678 | RxNorm |
| **1,184** | **Zofran Pill** | 2 | 34.29 | 1188161 | RxNorm |
| **1,187** | **dexmedetomidine Injection** | 3 | 34.02 | 1718899 | RxNorm |
| **1,190** | **fentanyl Injection** | 3 | 33.76 | 1735002 | RxNorm |
| **1,192** | **lidocaine hydrochloride 10 MG/ML Injectable Solution** | 3 | 33.65 | 1010033 | RxNorm |
| **1,199** | **hydrocortisone; rectal** | 3 | 33.14 | A07EA02 | ATC |
| **1,200** | **Tylenol Oral Product** | 2 | 33.10 | 1187311 | RxNorm |
| **1,202** | **acetaminophen Oral Suspension [Tylenol]** | 3 | 32.94 | 828554 | RxNorm |
| **1,203** | **acetaminophen 32 MG/ML Oral Suspension [Tylenol]** | 3 | 32.94 | 828555 | RxNorm |
| **1,204** | **Tylenol Oral Liquid Product** | 2 | 32.94 | 1187310 | RxNorm |
| **1,205** | **acetaminophen 32 MG/ML [Tylenol]** | 4 | 32.94 | 828553 | RxNorm |
| **1,206** | **fentanyl 0.05 MG/ML** | 4 | 32.60 | 328264 | RxNorm |
| **1,207** | **cough suppressants and mucolytics** | 3 | 32.52 | R05FB01 | ATC |
| **1,208** | **fentanyl; nasal, sublingual, transdermal** | 3 | 32.42 | N02AB03 | ATC |
| **1,211** | **ondansetron 4 MG [Zofran]** | 4 | 31.95 | 563812 | RxNorm |
| **1,213** | **salbutamol and ipratropium bromide; inhalant** | 3 | 31.64 | R03AL02 | ATC |

| | | | | | |
|---|---|---|---|---|---|
| **1,214** | **opium derivatives and mucolytics; systemic** | 3 | 31.58 | R05FA01 | ATC |
| **1,215** | **formoterol and beclometasone; inhalant** | 3 | 31.12 | R03AK08 | ATC |
| **1,217** | **Nitrofuran derivatives** | 2 | 30.60 | J01XE | ATC |
| **1,223** | **albuterol 5 MG/ML** | 4 | 30.26 | 2108264 | RxNorm |
| **1,224** | **Zofran Disintegrating Oral Product** | 2 | 29.95 | 1297043 | RxNorm |
| **1,225** | **ondansetron Disintegrating Oral Tablet [Zofran]** | 3 | 29.95 | 876689 | RxNorm |
| **1,226** | **midazolam Injection** | 3 | 29.85 | 1666797 | RxNorm |
| **1,227** | **ibuprofen 200 MG Oral Tablet** | 3 | 29.68 | 310965 | RxNorm |
| **1,228** | **lactic acid producing organisms; oral** | 3 | 29.66 | A07FA01 | ATC |
| **1,229** | **nitrofurantoin, combinations; systemic** | 3 | 29.65 | J01XE51 | ATC |
| **1,232** | **Other cough suppressants** | 2 | 29.55 | R05DB | ATC |
| **1,233** | **other cough suppressants - combinations** | 3 | 29.55 | R05DB20 | ATC |
| **1,234** | **albuterol 5 MG/ML Inhalation Solution** | 3 | 29.55 | 245314 | RxNorm |
| **1,236** | **acetaminophen 32 MG/ML Oral Solution** | 3 | 28.97 | 307675 | RxNorm |
| **1,237** | **acetaminophen Oral Solution** | 3 | 28.91 | 370506 | RxNorm |
| **1,241** | **ibuprofen 20 MG/ML [Junifen]** | 4 | 28.51 | 564197 | RxNorm |
| **1,242** | **ibuprofen Oral Suspension [Junifen]** | 3 | 28.51 | 365908 | RxNorm |
| **1,243** | **Junifen Oral Liquid Product** | 2 | 28.51 | 1167873 | RxNorm |
| **1,244** | **Junifen Oral Product** | 2 | 28.51 | 1167874 | RxNorm |
| **1,245** | **ibuprofen 20 MG/ML Oral Suspension [Junifen]** | 3 | 28.51 | 105850 | RxNorm |
| **1,246** | **2 ML fentanyl 0.05 MG/ML Injection** | 3 | 28.40 | 1735003 | RxNorm |
| **1,250** | **Short-acting sulfonamides** | 2 | 27.80 | J01EB | ATC |
| **1,251** | **combinations; systemic (short-acting sulfonamides)** | 3 | 27.80 | J01EB20 | ATC |
| **1,252** | **Intermediate-acting sulfonamides** | 2 | 27.80 | J01EC | ATC |
| **1,253** | **combinations; systemic (intermediate-acting sulfonamides)** | 3 | 27.80 | J01EC20 | ATC |
| **1,254** | **Long-acting sulfonamides** | 2 | 27.80 | J01ED | ATC |
| **1,255** | **combinations; systemic (long-acting sulfonamides)** | 3 | 27.80 | J01ED20 | ATC |
| **1,267** | **ondansetron 4 MG Disintegrating Oral Tablet [Zofran]** | 3 | 26.51 | 876690 | RxNorm |
| **1,270** | **ketorolac tromethamine 30 MG/ML** | 4 | 26.29 | 860095 | RxNorm |

| | | | | | |
|---|---|---|---|---|---|
| **1,273** | **hydromorphone** | 3 | 25.51 | 3423 | RxNorm |
| **1,276** | **lansoprazole, amoxicillin and clarithromycin; systemic** | 3 | 25.20 | A02BD07 | ATC |
| **1,282** | **Other antiepileptics** | 2 | 24.06 | N03AX | ATC |
| **1,285** | **Low-ceiling diuretics and potassium-sparing agents** | 2 | 23.77 | C03EA | ATC |
| **1,286** | **prednisolone, combinations; local oral** | 3 | 23.56 | A01AC54 | ATC |
| **1,287** | **hydrochlorothiazide and potassium-sparing agents; oral** | 3 | 23.54 | C03EA01 | ATC |
| **1,290** | **clindamycin, combinations; topical** | 3 | 22.93 | D10AF51 | ATC |
| **1,306** | **Diphenylmethane derivatives** | 2 | 21.40 | N05BB | ATC |
| **1,307** | **Anticholinergics** | 2 | 21.34 | S01FA | ATC |
| **1,308** | **azithromycin Oral Product** | 2 | 21.27 | 1155011 | RxNorm |
| **1,309** | **edetates** | 3 | 21.26 | V03AB03 | ATC |
| **1,316** | **oxycodone and acetylsalicylic acid; systemic** | 3 | 19.32 | N02AJ18 | ATC |
| **1,317** | **oxycodone and ibuprofen; systemic** | 3 | 19.22 | N02AJ19 | ATC |
| **1,318** | **Thyroid hormones** | 2 | 18.76 | H03AA | ATC |
| **1,319** | **cholecalciferol Pill** | 2 | 18.67 | 1156134 | RxNorm |
| **1,322** | **amoxicillin and beta-lactamase inhibitor; systemic** | 3 | 18.54 | J01CR02 | ATC |
| **1,326** | **albuterol Inhalation Solution** | 3 | 18.03 | 2108226 | RxNorm |
| **1,335** | **fluticasone** | 3 | 17.75 | 41126 | RxNorm |
| **1,336** | **vilanterol and umeclidinium bromide; inhalant** | 3 | 17.75 | R03AL03 | ATC |
| **1,338** | **amoxicillin Pill** | 2 | 17.25 | 1152901 | RxNorm |
| **1,339** | **clioquinol, combinations; oral, otic, rectal, topical, transdermal, vaginal** | 3 | 16.62 | P01AA52 | ATC |
| **1,340** | **Hydroxyquinoline derivatives** | 2 | 16.62 | P01AA | ATC |
| **1,342** | **multivitamins and trace elements; systemic** | 3 | 16.47 | A11AA04 | ATC |
| **1,343** | **Insulins and analogues for injection, fast-acting** | 2 | 16.43 | A10AB | ATC |
| **1,344** | **combinations; parenteral (insulins and analog. for injection, fast-acting)** | 3 | 16.43 | A10AB30 | ATC |
| **1,345** | **erythromycin, combinations; topical** | 3 | 16.31 | D10AF52 | ATC |
| **1,346** | **thyroid gland preparations; systemic** | 3 | 16.18 | H03AA05 | ATC |
| **1,347** | **combinations; parenteral (insulins and analog. for injection, intermediate- or long-acting comb. with fast-acting)** | 3 | 16.11 | A10AD30 | ATC |
| **1,348** | **Insulins and analogues for injection, intermediate- or long-acting combined with fast-acting** | 2 | 16.11 | A10AD | ATC |
| **1,349** | **Antivertigo preparations** | 2 | 15.82 | N07CA | ATC |
| **1,350** | **Progestogens and estrogens, fixed combinations** | 2 | 15.81 | G03AA | ATC |
| **1,352** | **rabeprazole, combinations; oral** | 3 | 15.67 | A02BC54 | ATC |
| **1,353** | **fluticasone; inhalant** | 3 | 15.59 | R03BA05 | ATC |
| **1,354** | **sodium chloride 4.5 MG/ML** | 4 | 15.29 | 1794564 | RxNorm |

| | | | | | |
|---|---|---|---|---|---|
| **1,356** | **Nitroimidazole derivatives** | 2 | 15.12 | P01AB | ATC |
| **1,357** | **Progestogens and estrogens, sequential preparations** | 2 | 15.05 | G03AB | ATC |
| **1,358** | **dexamethasone, combinations; inhalant, nasal** | 3 | 14.96 | R01AD53 | ATC |
| **1,359** | **Digitalis glycosides** | 2 | 14.74 | C01AA | ATC |
| **1,360** | **norethisterone and ethinylestradiol; systemic (progestogens and estrogens, fixed comb.)** | 3 | 14.69 | G03AA05 | ATC |
| **1,361** | **acetyldigoxin, combinations; systemic** | 3 | 14.65 | C01AA52 | ATC |
| **1,362** | **grass pollen; systemic** | 3 | 14.56 | V01AA02 | ATC |
| **1,363** | **Progestogens and estrogens, fixed combinations** | 2 | 14.36 | G03FA | ATC |
| **1,365** | **influenza B virus B/Washington/02/2019 antigen 0.03 MG/ML** | 4 | 14.14 | 2280740 | RxNorm |
| **1,367** | **Anticholinergics** | 2 | 13.96 | R03BB | ATC |
| **1,368** | **Sulfur containing products** | 2 | 13.95 | P03AA | ATC |
| **1,369** | **disulfiram, combinations; topical** | 3 | 13.95 | P03AA54 | ATC |
| **1,371** | **2 ML ondansetron 2 MG/ML Injection** | 3 | 13.69 | 1740467 | RxNorm |
| **1,374** | **meclozine, combinations; systemic** | 3 | 13.02 | R06AE55 | ATC |
| **1,375** | **ondansetron Injection** | 3 | 13.00 | 1740463 | RxNorm |
| **1,376** | **epinephrine Auto-Injector** | 3 | 12.97 | 1661387 | RxNorm |
| **1,377** | **morphine** | 3 | 12.65 | 7052 | RxNorm |
| **1,378** | **Progestogens and estrogens, sequential preparations** | 2 | 12.57 | G03FB | ATC |
| **1,379** | **vitamin d and analogues - combinations** | 3 | 12.49 | A11CC20 | ATC |
| **1,380** | **collagen, combinations; topical** | 3 | 12.47 | D11AX57 | ATC |
| **1,381** | **Intravaginal contraceptives** | 2 | 12.30 | G02BB | ATC |
| **1,382** | **hydrocortisone** | 3 | 12.04 | 5492 | RxNorm |
| **1,383** | **morphine; systemic, rectal** | 3 | 11.88 | N02AA01 | ATC |
| **1,384** | **dexmedetomidine 0.004 MG/ML** | 4 | 11.71 | 1249680 | RxNorm |
| **1,385** | **ondansetron Injectable Solution [Zofran]** | 3 | 11.57 | 94648 | RxNorm |
| **1,386** | **ondansetron 2 MG/ML [Zofran]** | 4 | 11.57 | 563814 | RxNorm |
| **1,387** | **ondansetron 2 MG/ML Injectable Solution [Zofran]** | 3 | 11.57 | 104897 | RxNorm |
| **1,388** | **Zofran Injectable Product** | 2 | 11.57 | 1188158 | RxNorm |
| **1,389** | **norethisterone and estrogen; systemic (progestogens and estrogens, fixed comb.)** | 3 | 11.53 | G03FA01 | ATC |
| **1,390** | **clonidine; systemic** | 3 | 11.42 | C02AC01 | ATC |
| **1,391** | **clonidine** | 3 | 11.36 | 2599 | RxNorm |
| **1,392** | **Other antimigraine preparations** | 2 | 11.36 | N02CX | ATC |

| | | | | | |
|---|---|---|---|---|---|
| **1,394** | **risedronic acid and calcium, sequential; oral** | 3 | 10.90 | M05BB02 | ATC |
| **1,395** | **Enzyme and acid preparations, combinations** | 2 | 10.82 | A09AC | ATC |
| **1,396** | **pepsin and acid preparations; systemic** | 3 | 10.82 | A09AC01 | ATC |
| **1,397** | **clavulanate** | 4 | 10.54 | 48203 | RxNorm |
| **1,398** | **amoxicillin / clavulanate Oral Product** | 2 | 10.54 | 1152874 | RxNorm |
| **1,400** | **morphine Injectable Product** | 2 | 10.12 | 1156360 | RxNorm |
| **1,401** | **Intrauterine contraceptives** | 2 | 10.09 | G02BA | ATC |
| **1,402** | **Diphtheria vaccines** | 2 | 10.05 | J07AF | ATC |
| **1,403** | **tetanus toxoid, combinations with diphtheria toxoid; systemic** | 3 | 10.05 | J07AM51 | ATC |
| **1,404** | **Tetanus vaccines** | 2 | 10.05 | J07AM | ATC |
| **1,405** | **colecalciferol; oral** | 3 | 9.86 | A11CC05 | ATC |
| **1,406** | **cetirizine Pill** | 2 | 9.79 | 1152447 | RxNorm |
| **1,407** | **pertussis, inactivated, whole cell, combinations with toxoids; systemic** | 3 | 9.67 | J07AJ51 | ATC |
| **1,408** | **cetirizine Oral Tablet** | 3 | 9.58 | 371364 | RxNorm |
| **1,409** | **lidocaine hydrochloride 20 MG/ML** | 4 | 9.38 | 1010670 | RxNorm |
| **1,410** | **cinnarizine, combinations; oral** | 3 | 9.33 | N07CA52 | ATC |
| **1,411** | **hydrocortisone; rectal, topical** | 3 | 9.19 | C05AA01 | ATC |
| **1,413** | **cholecalciferol** | 3 | 9.03 | 2418 | RxNorm |
| **1,415** | **influenza A virus A/Tasmania/503/2020 (H3N2) antigen 0.03 MG/ML** | 4 | 8.90 | 2561272 | RxNorm |
| **1,416** | **influenza A virus A/Victoria/2570/2019 (H1N1) antigen 0.03 MG/ML** | 4 | 8.90 | 2479042 | RxNorm |
| **1,417** | **Other drugs affecting bone structure and mineralization** | 2 | 8.87 | M05BX | ATC |
| **1,418** | **cholecalciferol Oral Product** | 2 | 8.83 | 1156133 | RxNorm |
| **1,420** | **hydroxyzine, combinations; systemic** | 3 | 8.66 | N05BB51 | ATC |
| **1,421** | **Imidazoline receptor agonists** | 2 | 8.63 | C02AC | ATC |
| **1,423** | **hydrocortisone aceponate; topical** | 3 | 8.11 | D07AC16 | ATC |
| **1,424** | **hydrocortisone buteprate; topical** | 3 | 8.11 | D07AB11 | ATC |
| **1,425** | **hydrocortisone; topical** | 3 | 8.11 | D07AA02 | ATC |
| **1,426** | **undecylenic acid, combinations; topical** | 3 | 8.08 | D01AE54 | ATC |
| **1,428** | **fluticasone; nasal** | 3 | 8.03 | R01AD08 | ATC |
| **1,429** | **Other antihistamines for systemic use** | 2 | 8.01 | R06AX | ATC |
| **1,430** | **fluticasone Inhalant Product** | 2 | 7.87 | 1165655 | RxNorm |
| **1,431** | **morphine sulfate 2 MG/ML** | 4 | 7.79 | 892588 | RxNorm |

| | | | | | |
|---|---|---|---|---|---|
| **1,432** | **Other antibacterials** | 2 | 7.70 | J01XX | ATC |
| **1,433** | **troxerutin, combinations; systemic** | 3 | 7.42 | C05CA54 | ATC |
| **1,434** | **200 ACTUAT albuterol 0.09 MG/ACTUAT Metered Dose Inhaler** | 3 | 7.09 | 745679 | RxNorm |
| **1,435** | **Iron bivalent, oral preparations** | 2 | 7.03 | B03AA | ATC |
| **1,436** | **cetirizine hydrochloride 10 MG** | 4 | 6.92 | 1011480 | RxNorm |
| **1,438** | **Progestogens** | 2 | 6.76 | G03AC | ATC |
